# Exon variants associated with asthma and allergy^1^

**DOI:** 10.1101/2022.02.09.22270730

**Authors:** Matthias Wjst

**Affiliations:** Institute of Lung Health and Immunity (LHI), Helmholtz Zentrum München German Research Center for Environmental Health, (GmbH), Ingolstädter Landstr. 1, 85764 München-Neuherberg, Germany; Institut für Medizinische Informatik, Statistik und Epidemiologie, Lehrstuhl für Medizinische Informatik, Klinikum rechts der Isar, Grillparzerstr. 18, 81675 München, Germany

## Abstract

Recent biobank based exon sequencing studies included thousands of traits while the mutational spectrum of asthma and allergy associated genes is still unknown.

**Methods:** Meta-analysis of exome data from 281,104 UK Biobank samples that were analyzed for association of mostly rare variants with asthma, allergic rhinitis and atopic dermatitis. Variants of interest (VOI) were tabulated, shared genes annotated and compared to earlier GWAS, WGBS, WES and selected candidate gene studies.

**Results:** 354 VOI were significantly associated with the traits examined. They cluster mainly in two large regions on chromosome 6 and 17 while there is basically no overlap of atopic dermatitis with both other diseases. After exclusion of the two atopic dermatitis variants, 321 unique VOI remain in 122 unique genes. 30 genes are shared by the group of 87 genes with increased and the group of 65 genes with decreased risk for allergic disease. 85% of genes identified earlier by common SNPs in GWAS can not be replicated.

**Discussion:** Most identified genes are involved in interferon γ and IL33 signaling pathway. They highlight already known but also new pharmacological targets, including the IL33 receptor ST2/IL1RL1, TLR1, ALOX15, GSDMA, BTNL2, IL13 and IKZF3. Future pharmacological studies will need to included these VOI for stratification of the study population.

---

It has been a long way from the first molecular study of David Marsh in the early seventies (3) until the asthma genetics meeting at the Transatlantic Airway Conference 1997 in Key Biscaine where the first positional candidate gene from Tristan da Cunha was announced (4). A genome-wide linkage scan (5) confirmed the chromosome 6 linkage (3), while only the following two studies tagged chromosome 17 (6) or chromosome 2 and 9 (7) where later the IKZF3-ORMDL3-GSDMA cluster, IL33 and its receptor (8) could be identified as asthma genes (9). The genomic resolution was however poor during that time. Only until thousands of SNPs had been discovered (10), the technology opened the door to hundreds of association studies. It was reaching a new height by a meta-analysis of 180,000 cases (11) in 2017 but unfortunately leaving the genetics of asthma and allergy basically stuck with long lists of SNPs (12). The situation changed only by the last year when two ultra-large exome scans releasing the variants of 281,000 (13) respective 455,000 participants of the UK biobank (14).

Rare exon variants (15) are not uniformly distributed across the genome but depend on many factors from sequence context to selective pressure of the population. Highly deleterious mutations will not be transmitted to offspring so mainly missense and synonymous variants are found with milder but nevertheless important effects. As most of these variants are in the coding region of genes, they are expected to identify directly functional active genes just like knock-out or knock-in experiments in animals. As humans are outbred, a significantly increased and decreased risk by a mutation may not be associated with disease due to the variable genetic background or possible rescue mutations (16). Nevertheless a significant association of a mutation with reasonable effects size will highlight the chain of events protecting from or leading to complex disease.

As the recent biobank based exon studies included thousands of different traits, no particular attention could be given to allergy and asthma phenotypes. An meta-analysis in this large database is therefore timely.

## Methods

Whole genome exome sequencing (WES) data for UK Biobank participants were generated at the Regeneron Genetics Center as described before (13) in a collaboration between AbbVie, Alnylam Pharmaceuticals, AstraZeneca, Biogen, Bristol-Myers Squibb, Pfizer, Regeneron and Takeda with the UK Biobank. Genomic DNA underwent paired-end 75bp whole exome sequencing using the IDT xGen v1 capture kit on the NovaSeq 6000 platform (13). The kit consists of 5′ biotin–modified oligonucleotide probes that are individually synthesized and analyzed by electrospray ionization-mass spectrometry (ESI-MS) covering (v2) 415,115 probes that spans a 34 Mb target region (19,433 genes) of the human genome.

For the present meta-analysis reference sequence BSgenome.Hsapiens.UCSC.hg38 was selected along with genomic coordinates hg38. Earlier data were converted to this format using the Golden Path Liftover tools from https://hgdownload.cse.ucsc.edu/goldenpath/hg19/liftOver. For gene names HUGO nomenclature (17) (https://www.genenames.org) was used while in a few cases SNPs or genes were matched using surrounding base sequence. Exome data were downloaded on Jan, 18, 2022 from https://azphewas.com. Allergic rhinitis was defined as “#20002#1387#hayfever|allergic_rhinitis (UK Biobank)”, atopic dermatitis as “Union#L20#L20 Atopic dermatitis” and asthma as “#6152#8#Asthma (UK Biobank) “ using the option variant-level results. As several known gene hits for example in IL33 were not found using the above asthma definition “Union#J45#J45 Asthma” was instead analyzed. Taken together, these are the results of 998 terabytes of raw exome sequence data from 302,355 UKB participants (18) resulting in 2,108,983 variants observed in at least six European ancestry participants corresponding to a MAF > 0.001%.

Using R software (19), the tidyverse framework and the biomaRt library, gene transcript data were obtained on Jan, 25, 2022 from https://www.ensembl.org/biomart/martview. GWAS data (11) were already received by Nov, 3, 2017 from
https://genepi.qimr.edu.au/staff/manuelf/gwas_results/intro.html This dataset is not complete as 23andme did not respond to repeated requests for data sharing. Linkage disequilibrium data were downloaded on Nov, 11, 2017 from https://data.broadinstitute.org/alkesgroup/LDSCORE. Gene set enrichment and pathway analysis were were done using the Enrichr API https://maayanlab.cloud/Enrichr (20) including Reactome and Human Gene Atlas pathways. Data were plotted using ggplot and gviz libraries (21). The data analysis pipeline is available under https://github.com/under-score/allergy_exome

## Results

For the trait allergic rhinitis allelic results of 6,917 variants exome variants were downloaded, leading to 50 significantly associated variants of interest (VOI). For asthma 9,695 VOI were identified, where 302 allelic VOI remain after applying a stringent significance threshold of P≤2×10^−9^ as used before (13). For atopic dermatitis 5,965 VOI were found but only 2 significant VOI remain after filtering. Both VOI, 1_152312600 and 1_152313385 are located in filaggrin. There are a few more f missense variants with odds ratios >3 for allergic rhinitis and >5 for asthma that are not significant by the applied genome-wide threshold. As these VOI do not include any known asthma or allergy gene, it is unlikely that important genes have been missed by the current threshold.

Genome positions for all VOI are given in Figure 1 as well as in the supplemental table. Clearly, there two major gene cluster on chromosome 6 and 17 that are known from earlier association studies.

**Figure 1:**
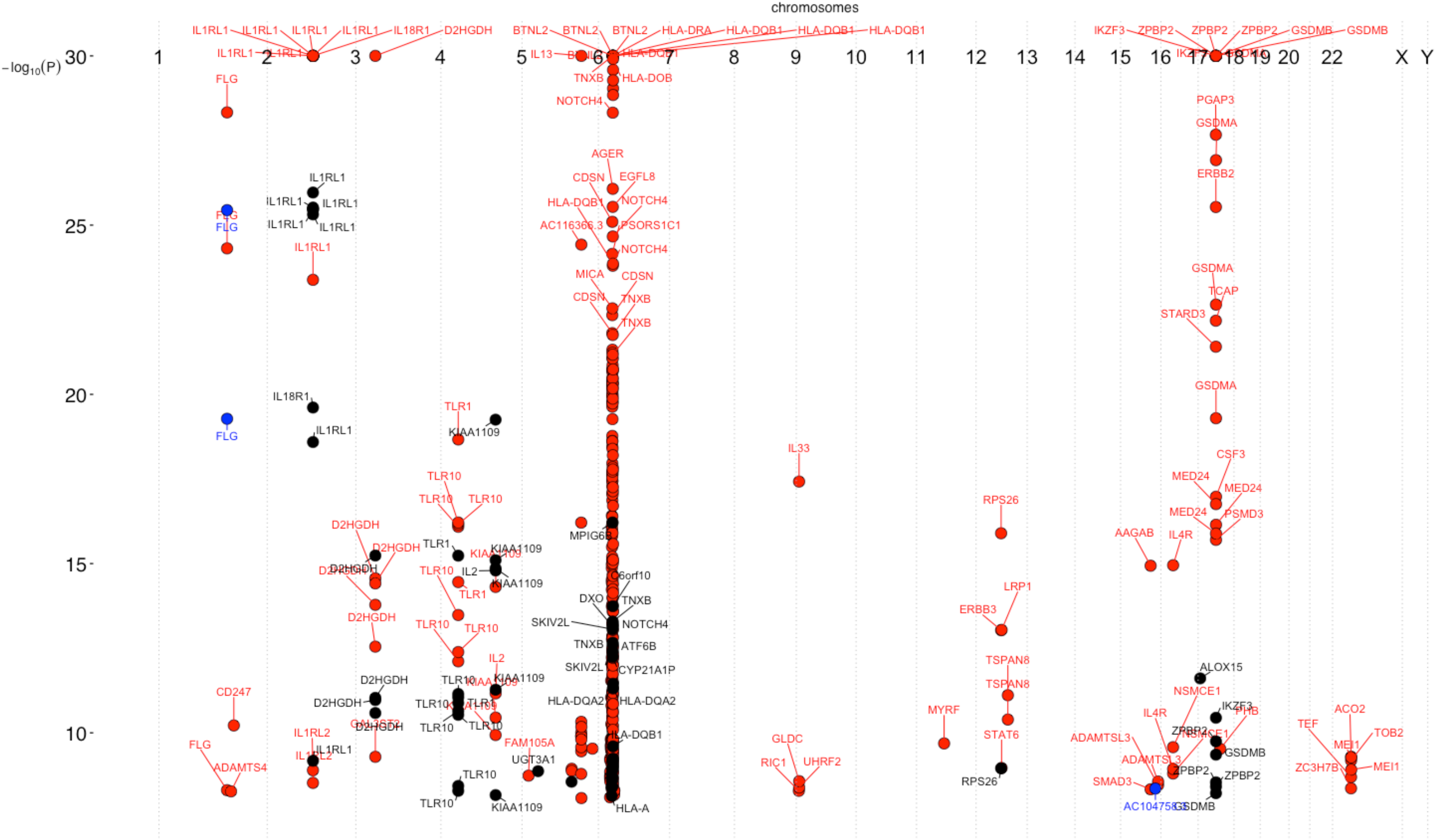
Exon variants of interest (N=354) significantly associated with asthma, allergic rhinitis oder atopic dermatitis in 281,104 UK Biobank samples. The plot shows significance using a threshold of P≤2×10^−9^ versus chromosomal position as Manhattan plot. Atopic dermatitis is indicated by blue dots, asthma by red dots and allergic rhinitis by black dots. Variant clustering is observed at chromosome 6 and 17.

After exclusion of the two atopic dermatitis variants, 352 VOI remain of which 321 are unique. They are situated in 122 unique genes while there is an overlap of 30 genes in the 87 genes with increased and the 65 genes associated with decreased odds ratios (Figure 2). Most VOI are leading to missense base exchanges (41%), followed by synonymous (30%), non coding (12%) and splice (5%) variants among others (Fig S1). VOI usually have rather low allele frequencies (Fig S2) and modest odds ratios (Fig S3). Most interesting are probably missense variants of high allele frequency but decreased risk for asthma and allergic rhinitis (Fig S4 until Fig S10) where primarily VOI in IL1RL1 (IL33 receptor), PGAP3 (a phospholipase), IKZF3 (B cell transcription factor), ZPBP2 (zona pellucida binding protein) and GSDMB (gasdermin) are interesting as they have been identified already in earlier association studies. Fig S13 to S127 contain genomic plots for all these genes.

**Figure 2:**
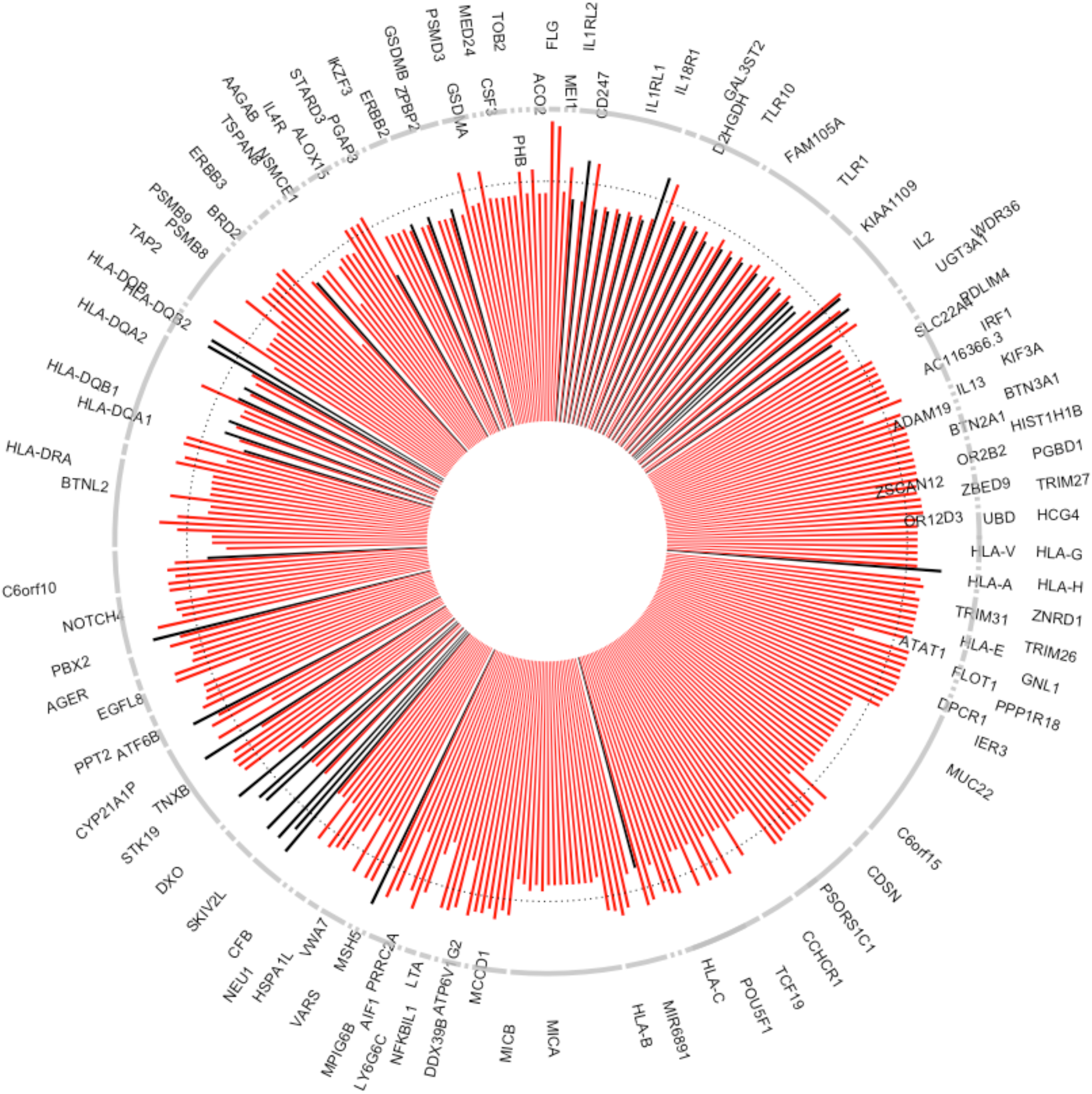
Exon variants of interest (N=352) significantly associated with asthma or allergic rhinitis in 281,104 UK Biobank samples. The plot shows odds ratios of variants filter by a threshold of P≤2×10^−9^ as circular barplot where OR of 1 is indicated by a dotted line. Results for asthma are shown as red bars, allergic rhinitis as black bars. There were more variants associated with asthma than with alllergic rhinitis while also number of variants per gene is highly variable with most VOI seen in MUC22 (N=18), MICA (N=15) and IL1RL1 (N=14).

**Figure 3:**
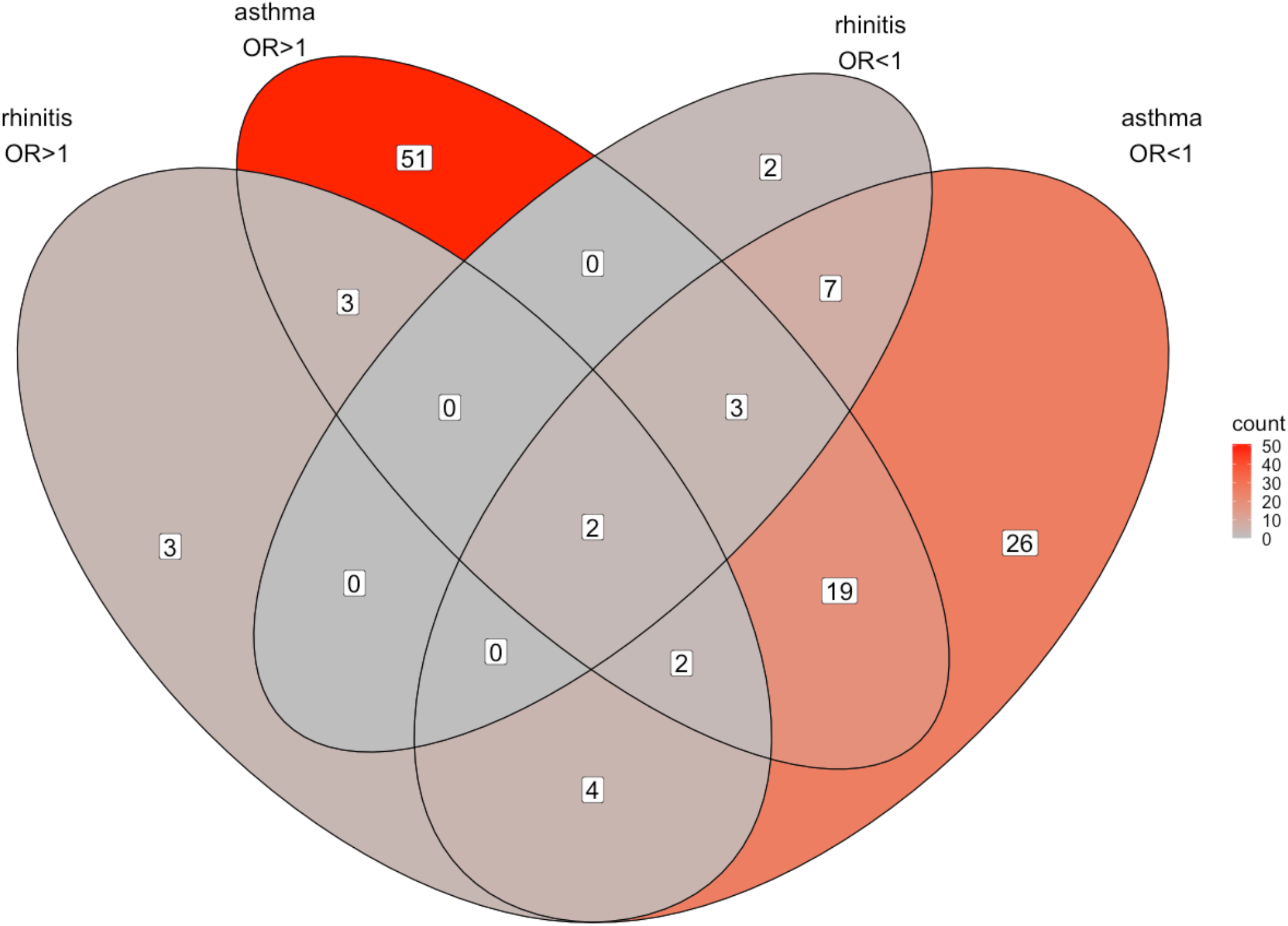
Shared genes (N=122) significantly associated with asthma or allergic rhinitis in 281,104 UK Biobank sample by either increased or decreased odds ratios. Most genes are observed in the group with increased asthma risk, followed by the group with decreased asthma risk.

An annotation of the VOI showed rather identical pathways for both protective and risk VOI. While increased asthma/allergy risk is more associated with defunct genes of the interferon γ pathway, an increased risk was found more with general cytokine signaling (Fig S11 and Fig S12).

As a last question we ask if there are any shared genes with earlier GWAS studies. When comparing the currently defined 122 genes with the 193 genes in a recent meta-analysis (11) there is only a small group of 21 shared genes, including IL1RL1, IL18R1, D2HGDH, TLR1, KIAA1109, RPS26, ALOX15, ZPBP2, GSDMB, FLG, CD247, FAM105A, WDR36, SLC22A4, IL13, HLA-C, PRRC2A, ERBB3, STAT6, AAGAB and MEI1.

## Discussion

Exome sequencing has been around now for a decade (22) and is even increasingly used also in clinical diagnostics. While the wet technology is advanced, data analysis remains a bottleneck even by using preprocessed data. At present false positive or negative findings cannot be excluded as there is neither a second sample of that size. Although we can assume from the large UK Biobank Consortium that there are no genotyping errors, that phenotypes have been correctly ascribed and data correctly handled there is still some risk of the known “Weisburd’s paradox”, the difficulty in maintaining quality control as studies get larger (23). Non-exon and some ultra-rare exonic variants will also not been included here. Nevertheless some hypotheses can already be generated.

### Atopic dermatitis

It seems from this analysis, that the genetics of atopic dermatitis is clearly separated from allergic rhinitis and asthma. Atopic dermatitis is already sufficiently described by filaggrin mutations (24) and in line with our early observation that atopic dermatitis is a genetically a largely different disease (25). Filaggrin mutations may act on top of an allergic predisposition while there have been no further shared variants in this analysis.

### Asthma

The asthma risk is increased by a large number of variants. Most of them are being private and basically limited to two genomic regions in know inflammatory gene cluster that are thought to be jointly regulated (26). The risk can be also diminished by a number of missense variants in the same gene cluster, requiring more detailed molecular studies. It should be noted that also VOI in pseudogenes may have functional effects by partial recombination with functional genes or by providing inactive transcription factor bindings sites. (27). Also synonymous base exchanges (28) may affect translational efficiency by indirect effects on mRNA stability.

### Allergic rhinitis

risk is increased by a much lower number of variants that are more heterogeneously distributed. A major peak is found at chromosome 2 that is mainly caused by IL1RL1/ST2, the IL33 receptor (29).

### Earlier GWAS

While there are numerous explanations for the “missing heritability” (30-32) there are less explanations of the “vague associations” in recent GWAS studies. The focus on the common disease/common variants may have lead to inadequate conclusions where inflated statistical methods have led to synthetic association (33). There might be local linkage disequilibrium effects along with undetectable selective sweeps. Recent GWAS hits (eg 83%) are either irrelevant but more likely, many are caused by functional, not structural gene variants. To resolve the apparent discrepancy of the numbers of significant GWAS SNPs and significant exon variants a combined analysis of common and rare variants in the same individuals would be helpful.

### Earlier WES (whole exome sequencing)

Backman (14) reported already two associations in the UK Biobank sample, the missense variant G>C 1_153775113 in SLC27A3 (OR_asthma_=0.65, p=8.2×10^−8^) as well as another G>C variant 9_6255967 in IL33 (OR_allergy_=0.60, P=9,52×10^−27^). The first variant is missing from the current dataset probably as there were not enough carriers; the variant seems to be also upstream of the ATG start site. The second VOI is also found in the current dataset with similar effect size (see p28 of the supplement) and identical to rs146597587 (34) whcih has a strong effect by disrupting a canonical splice acceptor site before the last coding exon of IL33.

### Earlier WGBS (whole genome bisulfite sequencing)

Comparing the current 122 genes with the 65 genes of a genomewide methylation scan of IgE in peripheral lymphocytes (35) we found only one shared gene (IL1RL1). Of the 24 genes found in whole blood of children with asthma (36) no shared gene could be identified.

### Other genes

Although ORMDL3 has been repeatedly favored by the Oxford group (10) along with their earlier claim on FcεR1ß/MS4A2 (37), a structural contribution of both genes to asthma and allergy is unlikely. Although evidence of absence is not absence of evidence, there is no further support that blocking MS4A2 or ORMDL3 expression (38) would be beneficial for asthma patients. CYSLTR2, the Tristan da Cunha asthma gene (39) was also not tagged here neither ADAM33 (40). FCER1A is also missing although a formal analysis of total IgE in UK Biobank is still pending. Genetic association for serum IL4 and IL13 were also surprisingly weak as well as for serum IFN***γ*** and IL10 (https://azphewas.com).

### Clinical relevance

In future studies, replication of this results with disease related QTL would be useful as well as interaction testing of VOI and the construction of improved risk scores (22). It is an open question if the previously described interaction of variants in IL1RL1 and IL5 or IL13 (41) will be also seen with the VOI described here.

The location of some variants can now even elucidate biology. All IL1RL1 VOI show a negative association with allergy except 2_102341256 in an exon that is exclusively used by the ST2V transcript (42) leading to an shortened version of the soluble decoy receptor sST2. If we assume a weaker IL33 blocking capacity, it seems reasonable to develop recombinant fusion proteins as alternative IL33 traps (43).

The VOI results provide also a basis for planning and re-analyzing clinical trials. While therapies to restore or substitute broken gene functions (eg VOI with increased disease risk) are are difficult to target at the moment, dampening or turning off genes (eg VOI that show decreased disease risk) are promising. IL33 is already one of the most relevant therapy targets where monoclonal antibodies including etokimab, itepekimab, tozorakimab have been tested in clinical trials. From the current analysis they are expected to have even stronger clinical effects than anti-IL4R (dupilumab), IL5 (mepolizumab, reslizumab), IL5R (benralizumab) or IL-13 (tralokinumab) antibodies while anti ST2 antibodies like astegolimab may be even more effective. A major condition for future clinical trial will be an adequate stratification of patients as for example individuals with a deleterious ALOX5 mutation will probably not respond to zileuton, neither will patients with an already defunct IL33 receptor respond to astegolimab. There are also refined or repurposed treatment options as for example lenalidomide is targeting IKZF3 (44) or disulfiram which was used earlier to treat atopic dermatitis (45) and may be able to block gasdermins (46). There are also new treatment possibilities of small-molecule antagonists like MMG-11 on TLR1 (47) or ZINC59514725 on IL1RL1 (48).

Whatever future studies will provide, there seems now a rational guideposts on the long and winding road (49). UK Biobank released here the largest structural genomic study of asthma and allergy genetics right at the 25th anniversary of the Transatlantic Airway Conference on genetics. There are still many efforts necessary to bring these results finally from bench to bedside.

## Data Availability

All data produced in the present study are available upon reasonable request to the authors

https://azphewas.com

https://www.ensembl.org/biomart/martview

https://genepi.qimr.edu.au/staff/manuelf/gwas_results/intro.html

https://data.broadinstitute.org/alkesgroup/LDSCORE

## Funding

none declared

## Conflicts of Interest

none declared

## Author contribution

study design, analysis, writing and review by M.W.

## Acknowledgments

I acknowledge the work by authors of the original study in particular K. Carss for providing help with the data. I thank also numerous other colleagues including S. Dold, H. Wickham, J.J. Allaire, Y. Xie and many anonymous colleagues at Github and Stackoverflow.

## Note

The paper summarizes various entries on the author’s blog on asthma genetics (keyword “Key Biscaine”).

## Notes

### Competing Interest Statement

The authors have declared no competing interest.

### Funding Statement

This study did not receive any funding

### Author Declarations

This study involves openly available human data, which can be obtained from https://azphewas.com

## References

1. Werenskiold AK, Hoffmann S, Klemenz R. Induction of a mitogen-responsive gene after expression of the Ha-ras oncogene in NIH 3T3 fibroblasts. Molecular and Cellular Biology. 1989;9:5207–5214.

2. Kakkar R, Lee RT. The IL-33/ST2 pathway: Therapeutic target and novel biomarker. Nature Reviews Drug Discovery. 2008;7:827–840.

3. Marsh DG, Bias WB, Hsu SH, Goodfriend L. Association of the HL-A7 cross-reacting group with a specific reaginic antibody response in allergic man. Science. 1973;179:691–693.

4. Vogel G. A scientific result without the science. Science. 1997;276:1327–1327.

5. Daniels SE, Bhattacharrya S, James A et al. A genome-wide search for quantitative trait loci underlying asthma. Nature. 1996;383:247–250.

6. CSGA. A genome-wide search for asthma susceptibility loci in ethnically diverse populations. Nat Genet. 1997;15:389–392.

7. Wjst M, Fischer G, Immervoll T et al. A genome-wide search for linkage to asthma. Genomics. 1999;58:1–8.

8. Yanagisawa K, Takagi T, Tsukamoto T, Tetsuka T, Tominaga S-i. Presence of a novel primary response gene ST2L, encoding a product highly similar to the interleukin 1 receptor type 1. FEBS letters. 1993;318:83–87.

9. Moffatt MF, Gut IG, Demenais F et al. A large-scale, consortium-based genomewide association study of asthma. New England Journal of Medicine. 2010;363:1211–1221.

10. Moffatt MF, Kabesch M, Liang L et al. Genetic variants regulating ORMDL3 expression contribute to the risk of childhood asthma. Nature. 2007;448:470–473.

11. Ferreira MA, Vonk JM, Baurecht H et al. Shared genetic origin of asthma, hay fever and eczema elucidates allergic disease biology. Nat Genet. 2017;49,1752–1757.

12. Wjst M, Sargurupremraj M, Arnold M. Genome-wide association studies in asthma: What they really told us about pathogenesis. Curr Opin Allergy Clin Immunol. 2012;13:112–118.

13. Wang Q, Dhindsa RS, Carss K et al. Rare variant contribution to human disease in 281,104 UK Biobank exomes. Nature. 2021;597:527–532.

14. Backman JD, Li AH, Marcketta A et al. Exome sequencing and analysis of 454,787 UK Biobank participants. Nature. 2021;599:628–634.

15. Lek M, Karczewski KJ, Minikel EV et al. Analysis of protein-coding genetic variation in 60,706 humans. Nature. 2016;536:285–291.

16. Lopes-Pacheco M, Boinot C, Sabirzhanova I, Rapino D, Cebotaru L. Combination of correctors rescues CFTR transmembrane-domain mutants by mitigating their interactions with proteostasis. Cell Physiol Biochem. 2017;41:2194–2210.

17. Standardized nomenclature and open science in Human Genomics. [editorial]. Hum Genomics 2021;15:13.

18. Bycroft C, Freeman C, Petkova D et al. The UK Biobank resource with deep phenotyping and genomic data. Nature. 2018;562:203–209.

19. Team RC. R: A language and environment for statistical computing. R Foundation for Statistical Computing, Vienna, Austria. URL https://www.R-project.org, last visited 2/2/22

20. Chen EY, Tan CM, Kou Y et al. Enrichr: interactive and collaborative HTML5 gene list enrichment analysis tool. BMC Bioinformatics. 2013;14:128.

21. Buja A, Cook D, Hofmann H et al. Statistical inference for exploratory data analysis and model diagnostics. Philos Trans A Math Phys Eng Sci. 2009;367:4361–4383.

22. Ripatti S, Tikkanen E, Orho-Melander M et al. A multilocus genetic risk score for coronary heart disease: case-control and prospective cohort analyses. Lancet. 2010;376:1393–1400.

23. Gelman A, Skardhamar T, Aaltonen M. Type M error might explain Weisburd’s paradox. Journal of Quantitative Criminology. 2020;36:295–304.

24. Hoober JK, Eggink LL. The discovery and function of filaggrin. International Journal of Molecular Sciences. 2022;23:1455.

25. Dold S, Wjst M, von Mutius E, Reitmeir P, Stiepel E. Genetic risk for asthma, allergic rhinitis, and atopic dermatitis. Arch Dis Child. 1992;67:1018–1022.

26. Makino T, McLysaght A. Interacting gene clusters and the evolution of the vertebrate immune system. Mol Biol Evol. 2008;25:1855–1862.

27. Balakirev ES, Ayala FJ. Pseudogenes: Are they “junk” or functional DNA? Annu Rev Genet. 2003;37:123–151.

28. Kristofich J, Morgenthaler AB, Kinney WR et al. Synonymous mutations make dramatic contributions to fitness when growth is limited by a weak-link enzyme. PLoS Genet. 2018;14:e1007615.

29. Portelli MA, Dijk FN, Ketelaar ME et al. Phenotypic and functional translation of IL1RL1 locus polymorphisms in lung tissue and asthmatic airway epithelium. JCI Insight. 2020;5

30. Manolio TA, Collins FS, Cox NJ et al. Finding the missing heritability of complex diseases. Nature. 2009;461:747–753.

31. Zuk O, Schaffner SF, Samocha K et al. Searching for missing heritability: designing rare variant association studies. Proc Natl Acad Sci U S A. 2014;111:E455–64.

32. Génin E. Missing heritability of complex diseases: case solved? Hum Genet. 2019; 139:103–113

33. Dickson SP, Wang K, Krantz I, Hakonarson H, Goldstein DB. Rare variants create synthetic genome-wide associations. PLoS Biology. 2010;8:e1000294.

34. Smith D, Helgason H, Sulem P et al. A rare IL33 loss-of-function mutation reduces blood eosinophil counts and protects from asthma. PLoS genetics. 2017;13:e1006659.

35. Liang L, Willis-Owen SAG, Laprise C et al. An epigenome-wide association study of total serum immunoglobulin E concentration. Nature. 2015; 520:670–674.

36. Xu C-J, Söderhäll C, Bustamante M et al. DNA methylation in childhood asthma: An epigenome-wide meta-analysis. The Lancet Respiratory Medicine. 2018;6:379–388.

37. Cookson WO, Sharp PA, Faux JA, Hopkin JM. Linkage between immunoglobulin E responses underlying asthma and rhinitis and chromosome 11q. The Lancet. 1989;1:1292–1295.

38. Luthers CR, Dunn TM, Snow AL. ORMDL3 and asthma: Linking sphingolipid regulation to altered T cell function. Front Immunol. 2020;11:597945.

39. Thompson MD, Capra V, Takasaki J et al. A functional G300S variant of the cysteinyl leukotriene 1 receptor is associated with atopy in a Tristan da Cunha isolate. Pharmacogenet Genomics. 2007;17:539–549.

40. Van Eerdewegh P, Little RD, Dupuis J et al. Association of the ADAM33 gene with asthma and bronchial hyperresponsiveness. Nature. 2002;418:426–430.

41. Demenais F, Margaritte-Jeannin P, Barnes KC et al. Multiancestry association study identifies new asthma risk loci that colocalize with immune-cell enhancer marks. Nat Genet. 2018;50:42–53.

42. Tominaga S-i, Kuroiwa K, Tago K, Iwahana H, Yanagisawa K, Komatsu N. Presence and expression of a novel variant form of ST2 gene product in human leukemic cell line UT-7/GM. Biochemical and biophysical research communications. 1999;264:14–18.

43. Holgado A, Braun H, Verstraete K et al. Single-chain soluble receptor fusion proteins as versatile cytokine inhibitors. Front Immunol. 2020;11:1422.

44. Krönke J, Udeshi ND, Narla A et al. Lenalidomide causes selective degradation of IKZF1 and IKZF3 in multiple myeloma cells. Science. 2014;343:301–305.

45. Kaaber K, Menne T, Tjell JC, Veien N. Antabuse treatment of nickel dermatitis. Contact Dermatitis. 1979;5:221–228.

46. Hu JJ, Liu X, Xia S et al. FDA-approved disulfiram inhibits pyroptosis by blocking gasdermin D pore formation. Nat Immunol. 2020;21:736–745.

47. Grabowski M, Murgueitio MS, Bermudez M, Wolber G, Weindl G. The novel small-molecule antagonist MMG-11 preferentially inhibits TLR2/1 signaling. Biochem Pharmacol. 2020;171:113687.

48. Mai TT, Nguyen PG, L. MT et al. Discovery of small molecular inhibitors for interleukin-33/ST2 protein-protein interaction: a virtual screening, molecular dynamics simulations and binding free energy calculations. Mol Divers. 2022

49. Ober C, Hoffjan S. Asthma genetics 2006: the long and winding road to gene discovery. Genes and Immunity. 2006;7:95–100.

