## Supplemental Table and Figures for "Exon variants associated with asthma and allergy^1^"

### Supplement

#### Figures general

##### Fig S1 distribution of exome variants by functional consequence

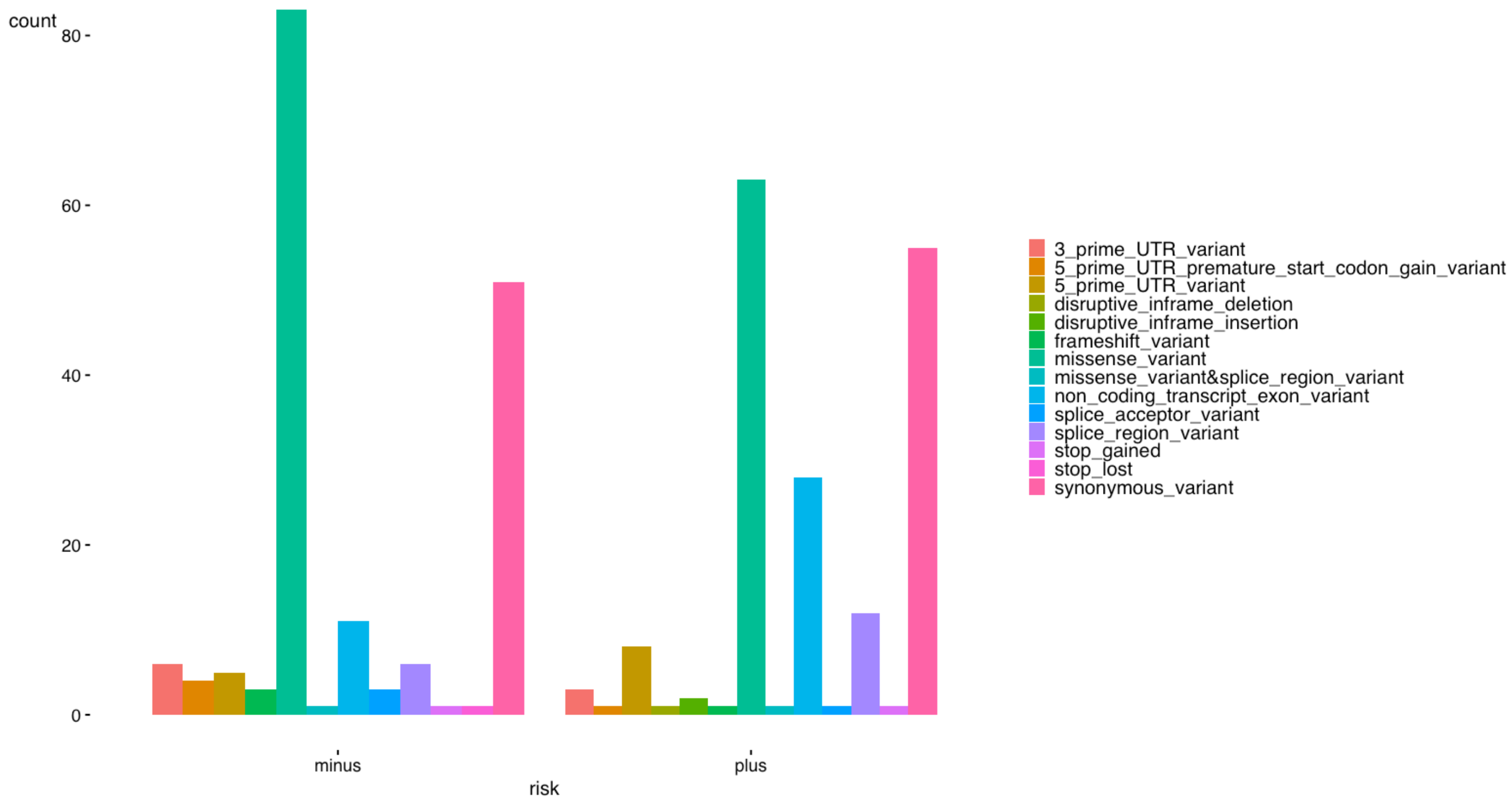

**Fig S2 distribution of exome variants by minor allele frequency in cases**

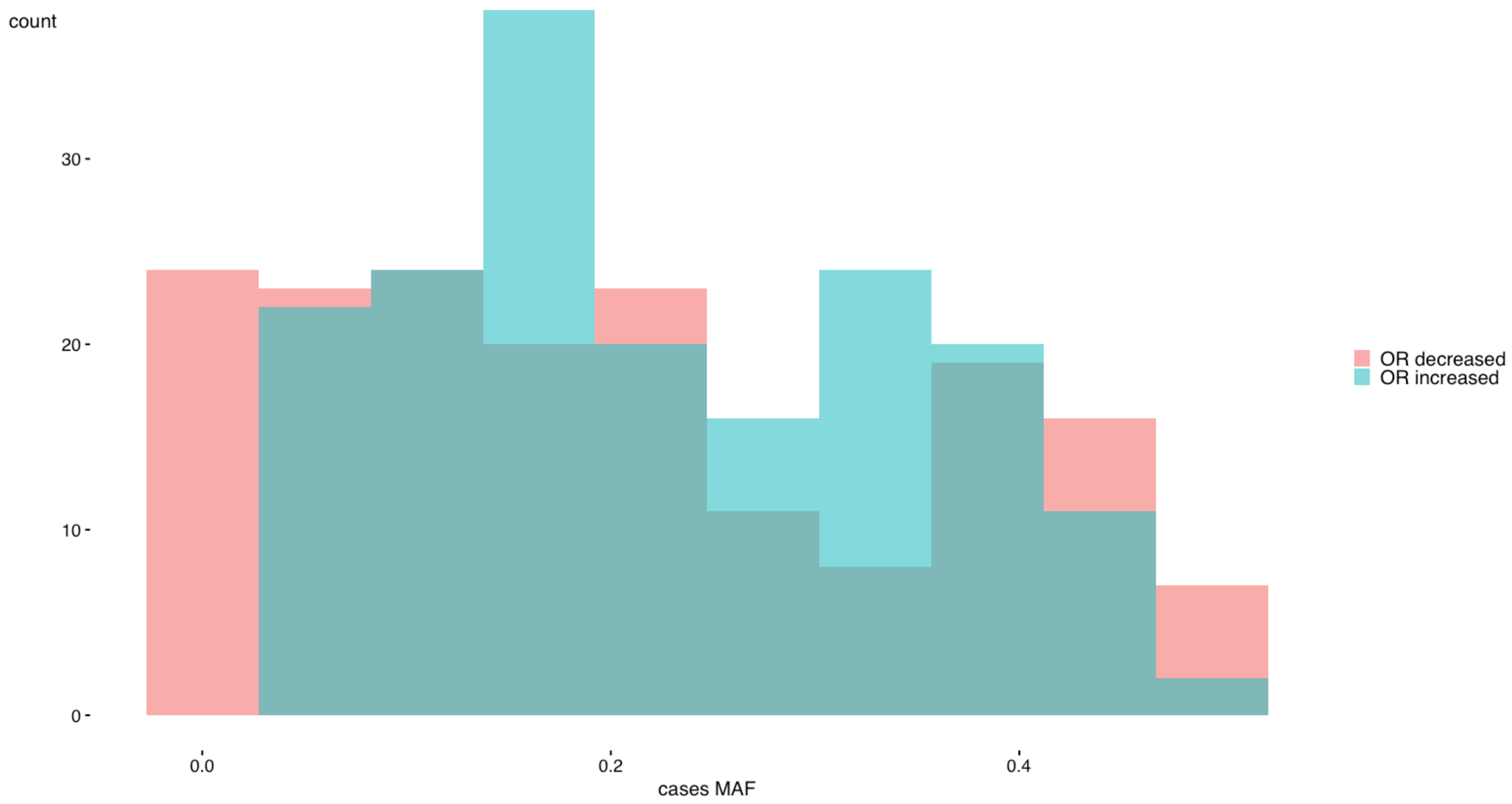

**Fig S3 distribution of odds ratios**

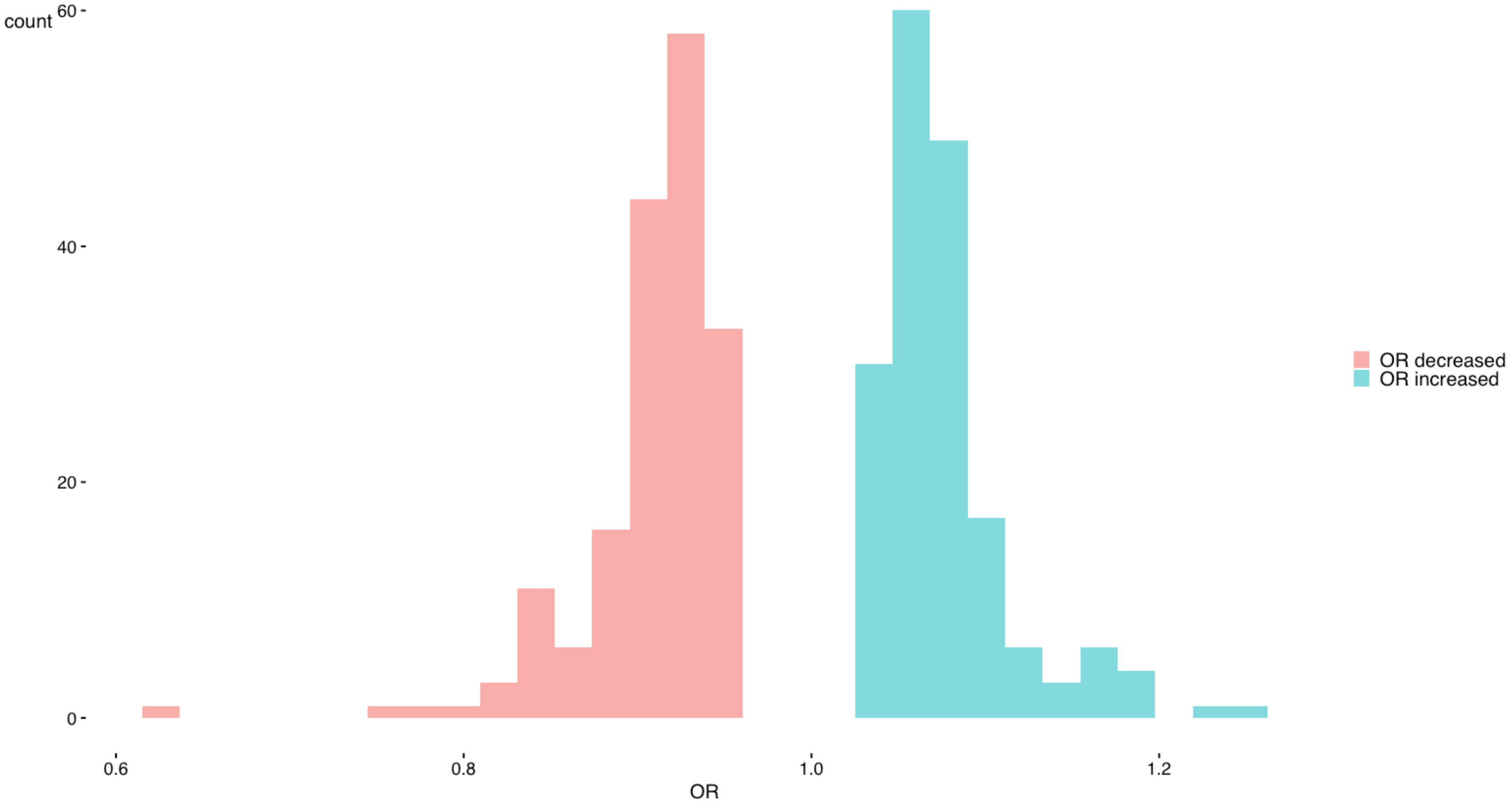

**Fig S4 exome variants by functional consequence: minor allele frequency in cases versus odds ratio**

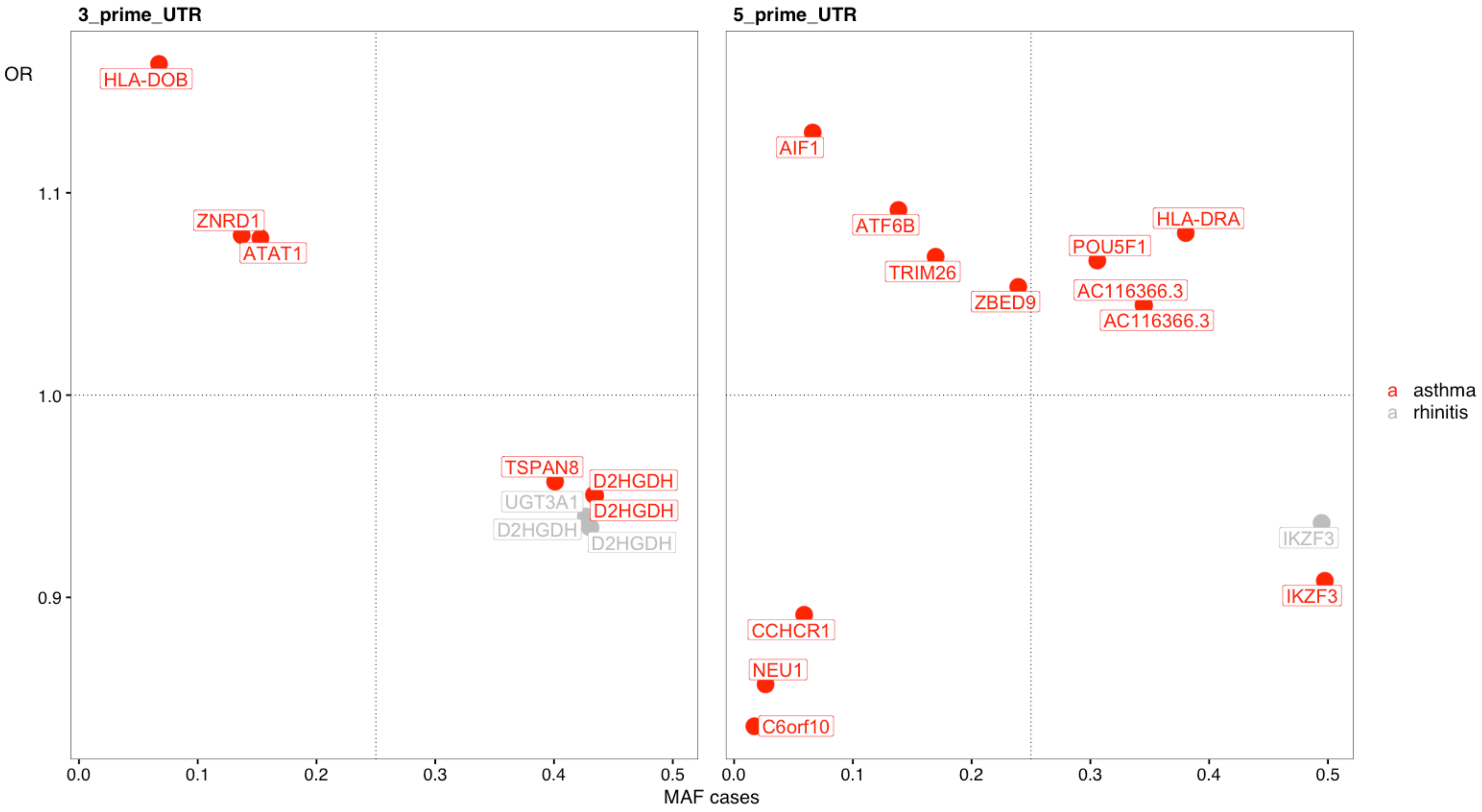

**Fig S5 exome variants by functional consequence: minor allele frequency in cases versus odds ratio**

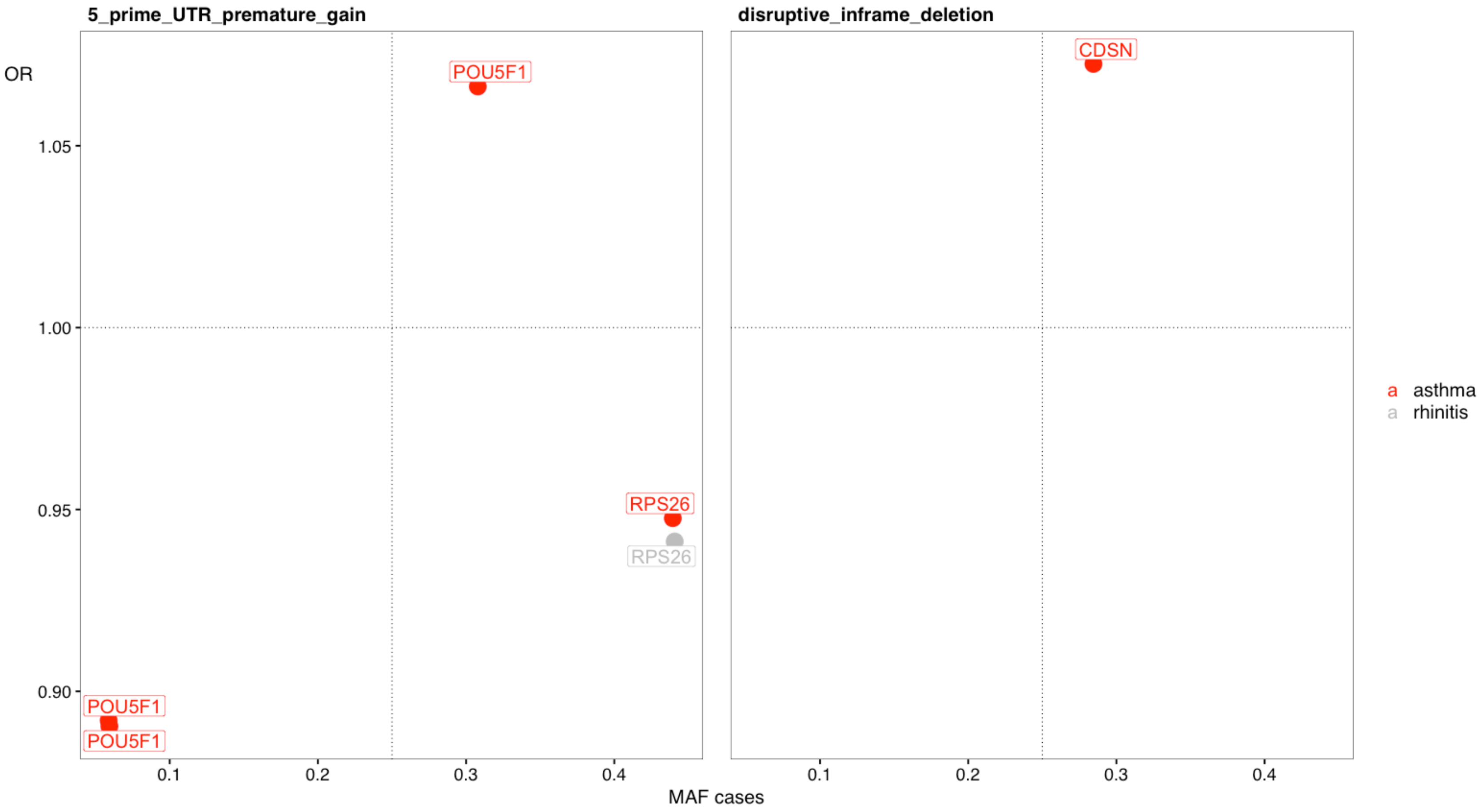

**Fig S6 exome variants by functional consequence: minor allele frequency in cases versus odds ratio**

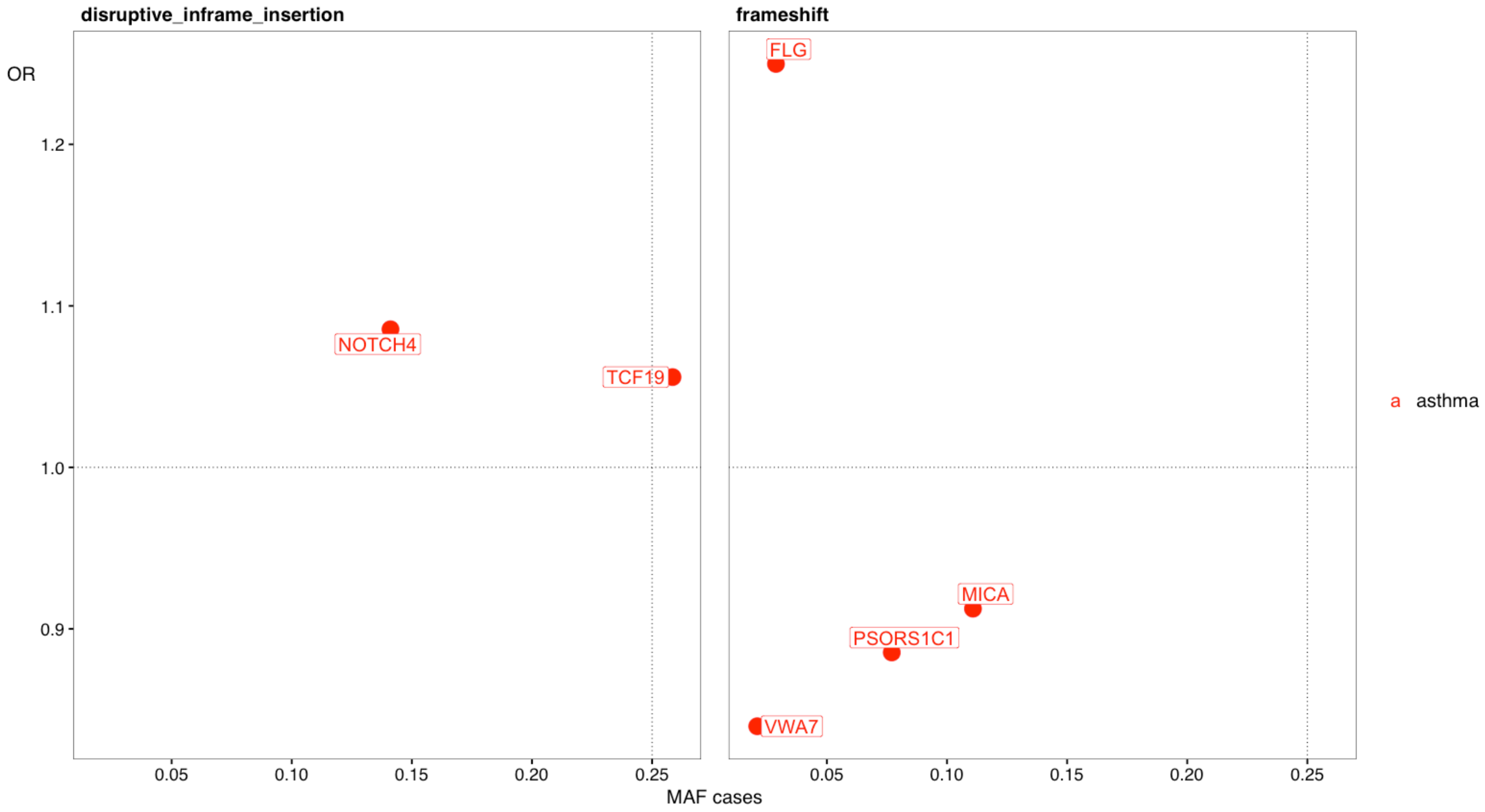

#### Fig S7 exome variants by functional consequence: minor allele frequency in cases versus odds ratio

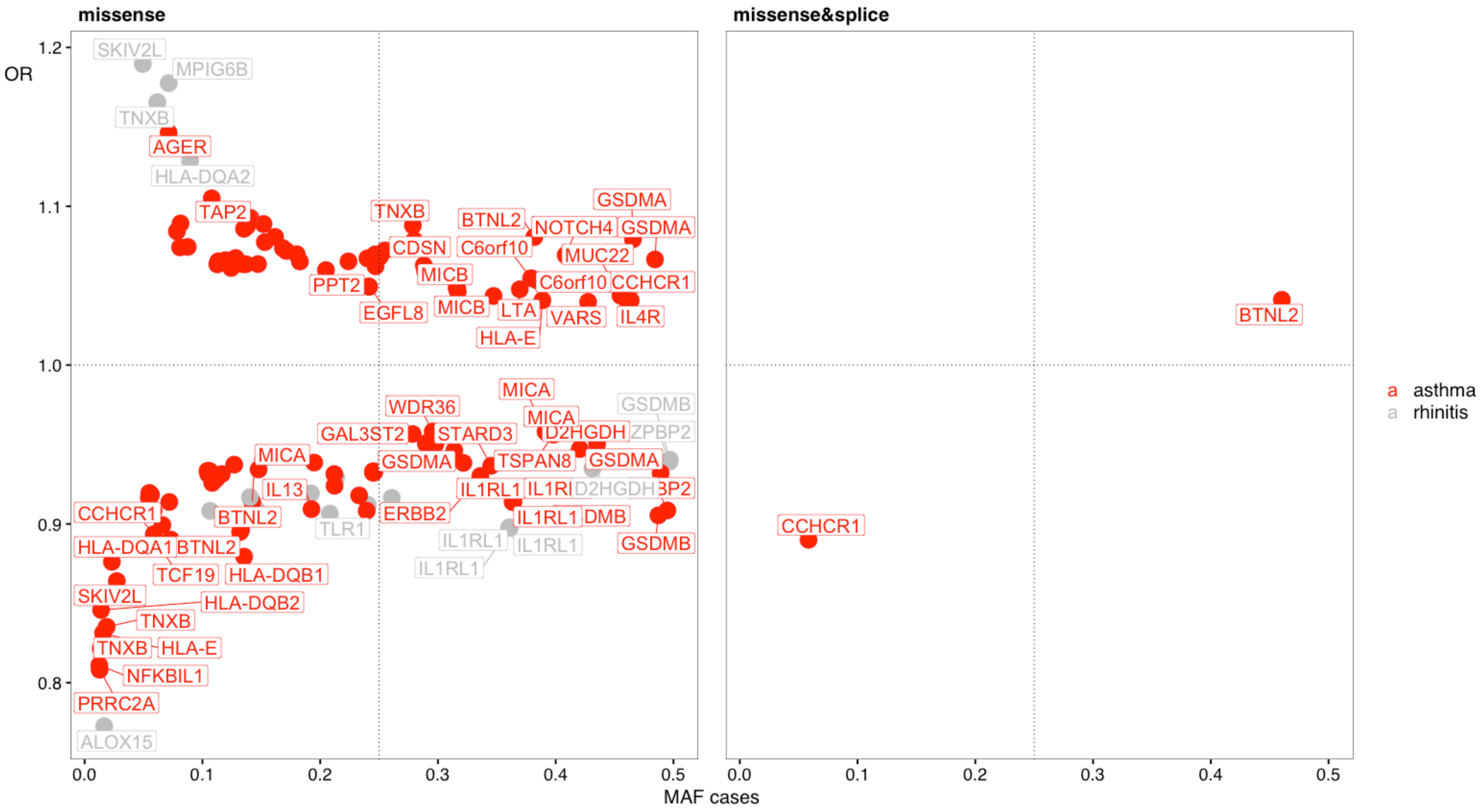

**Fig S8 exome variants by functional consequence: minor allele frequency in cases versus odds ratio**

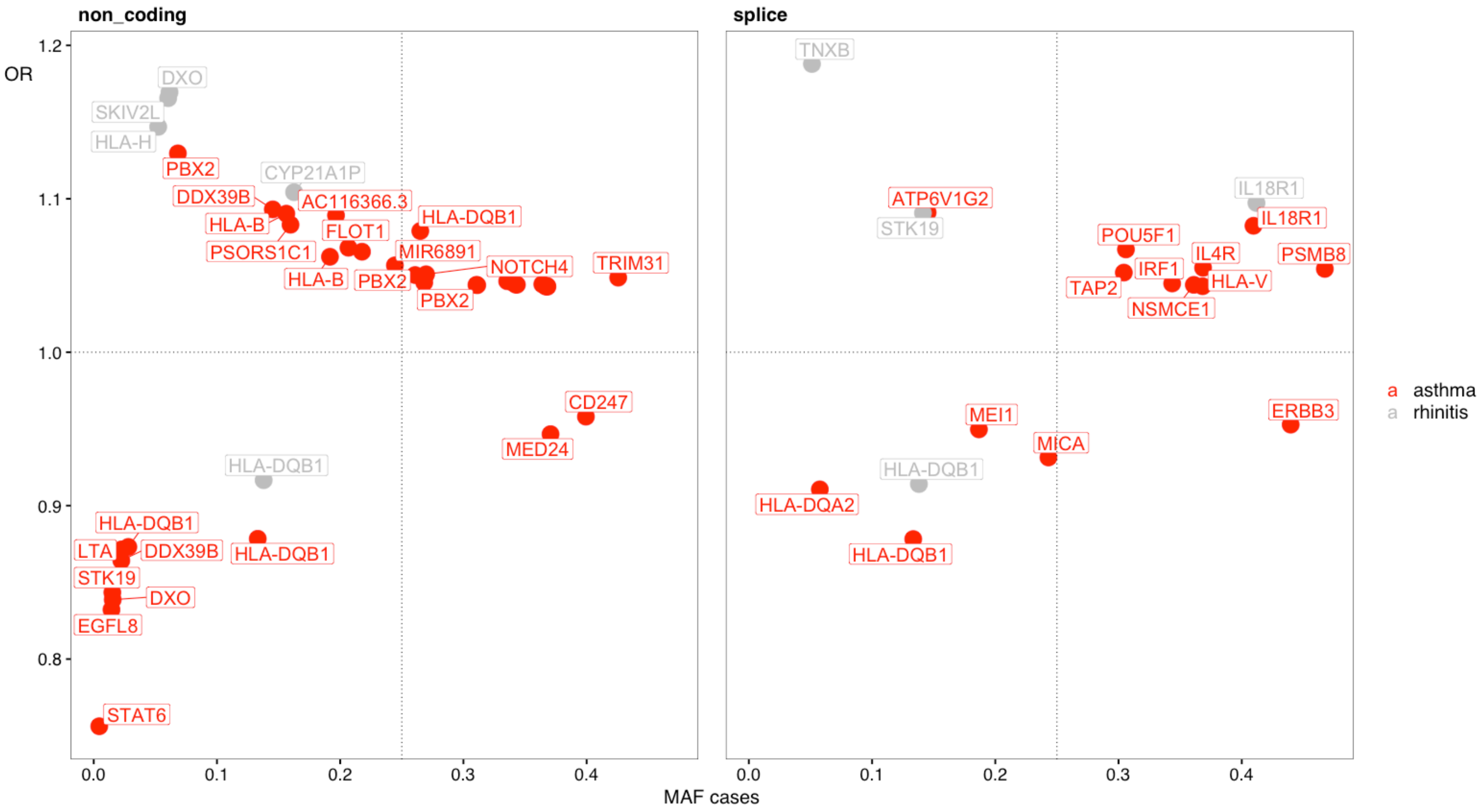

**Fig S9 exome variants by functional consequence: minor allele frequency in cases versus odds ratio**

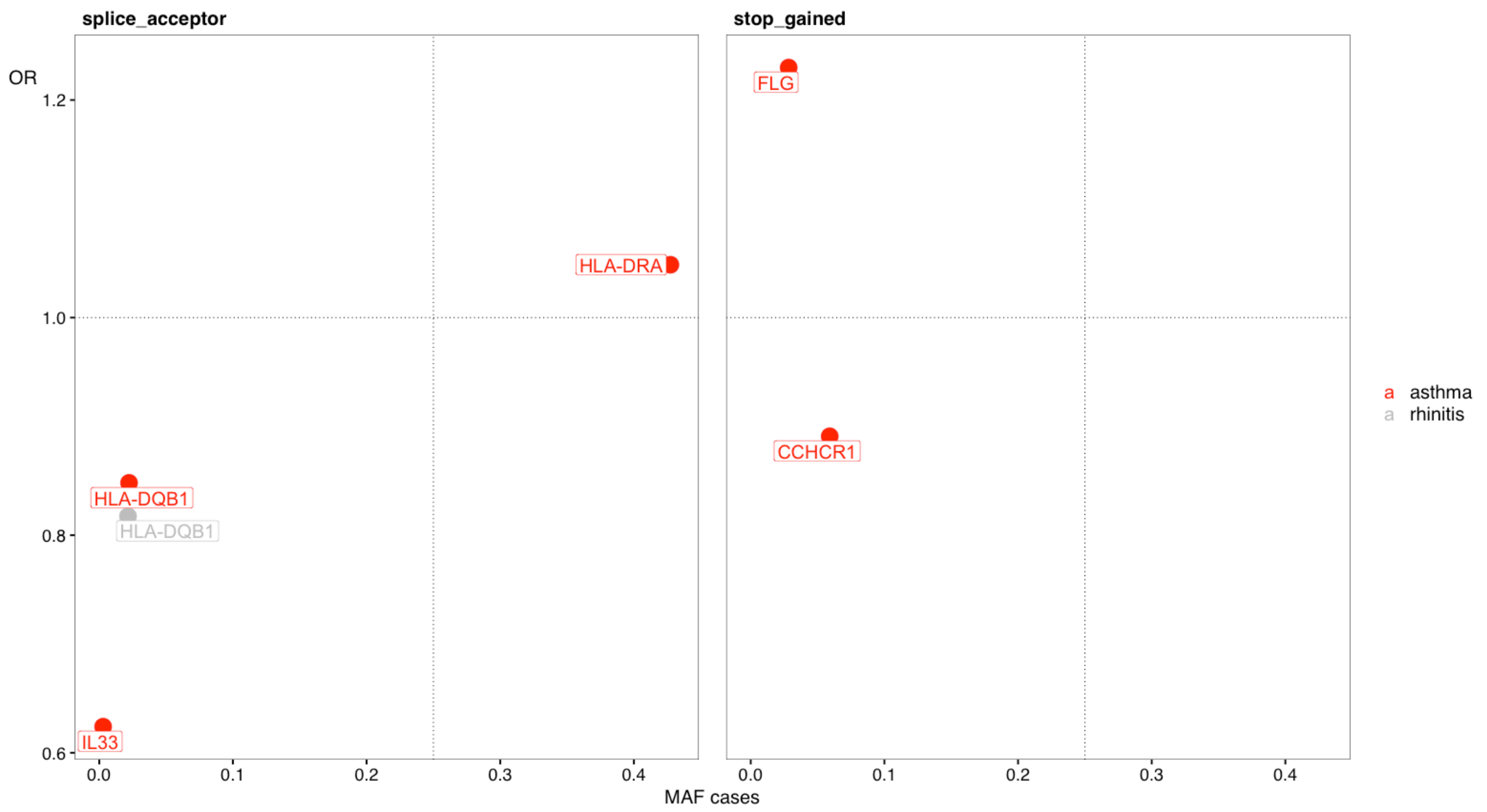

#### Fig S10 exome variants by functional consequence: minor allele frequency in cases versus odds ratio

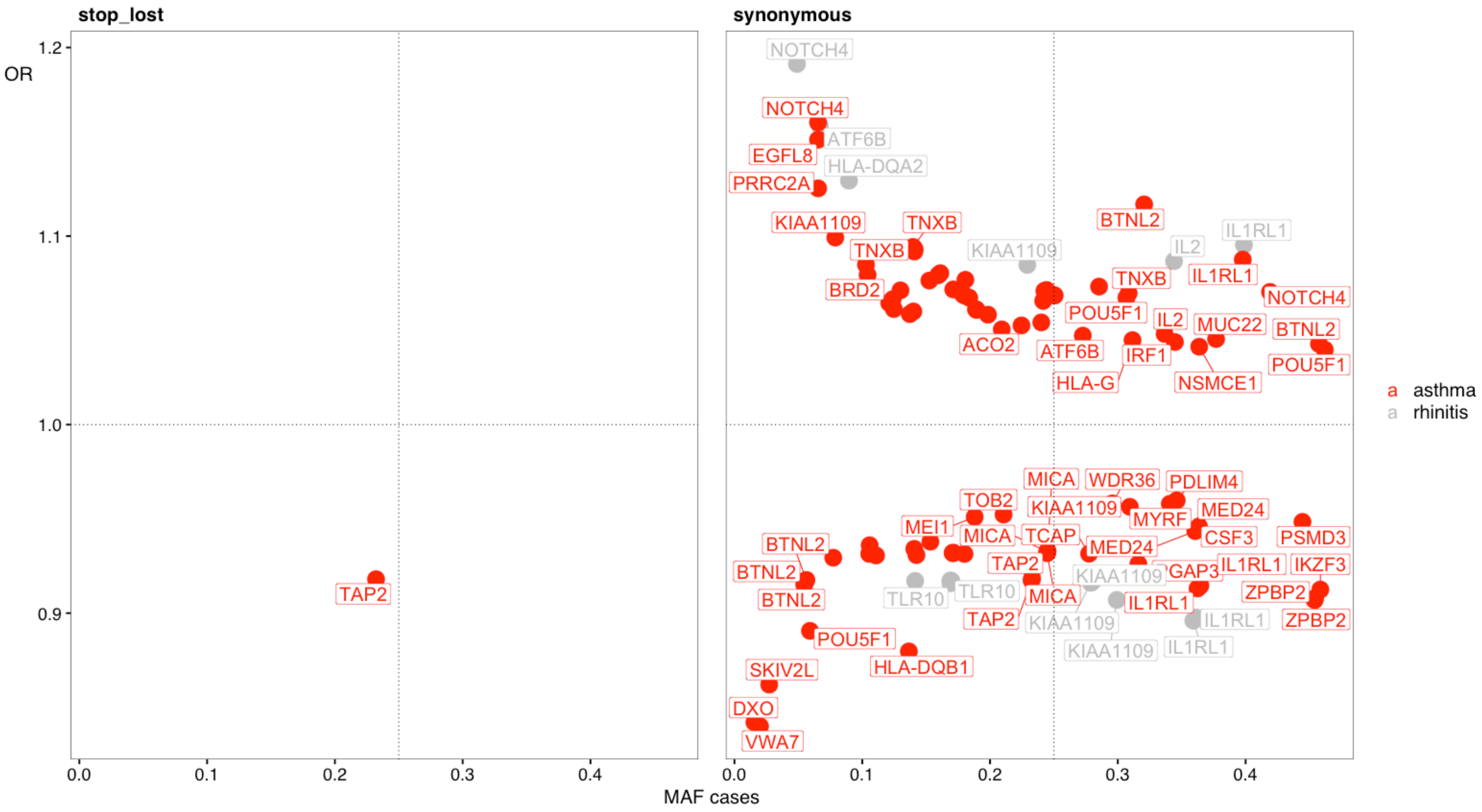

#### Fig S11 Reactome annotation of exome variants

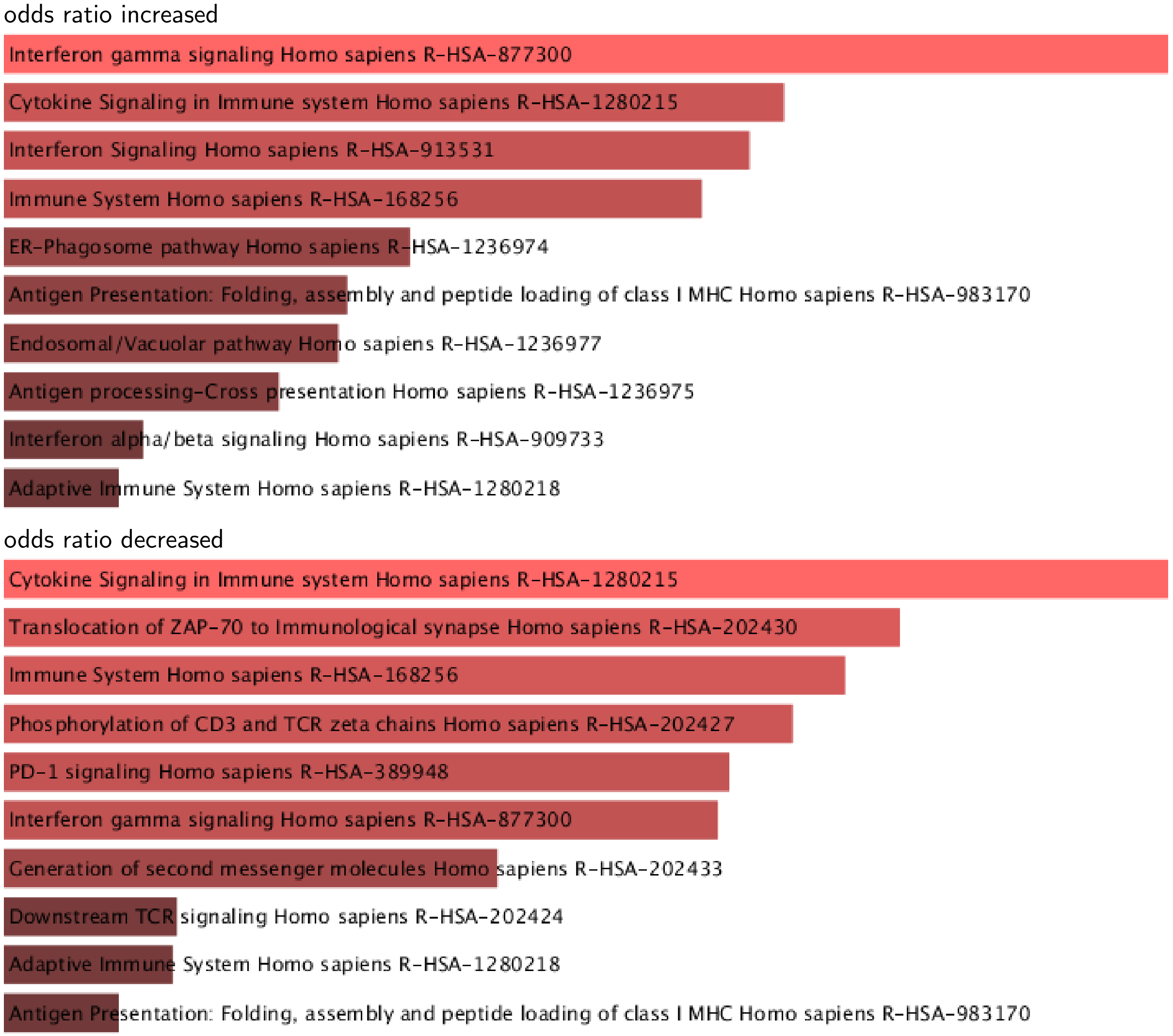

#### Fig S12 Human Gene Atlas annotation of exome variants

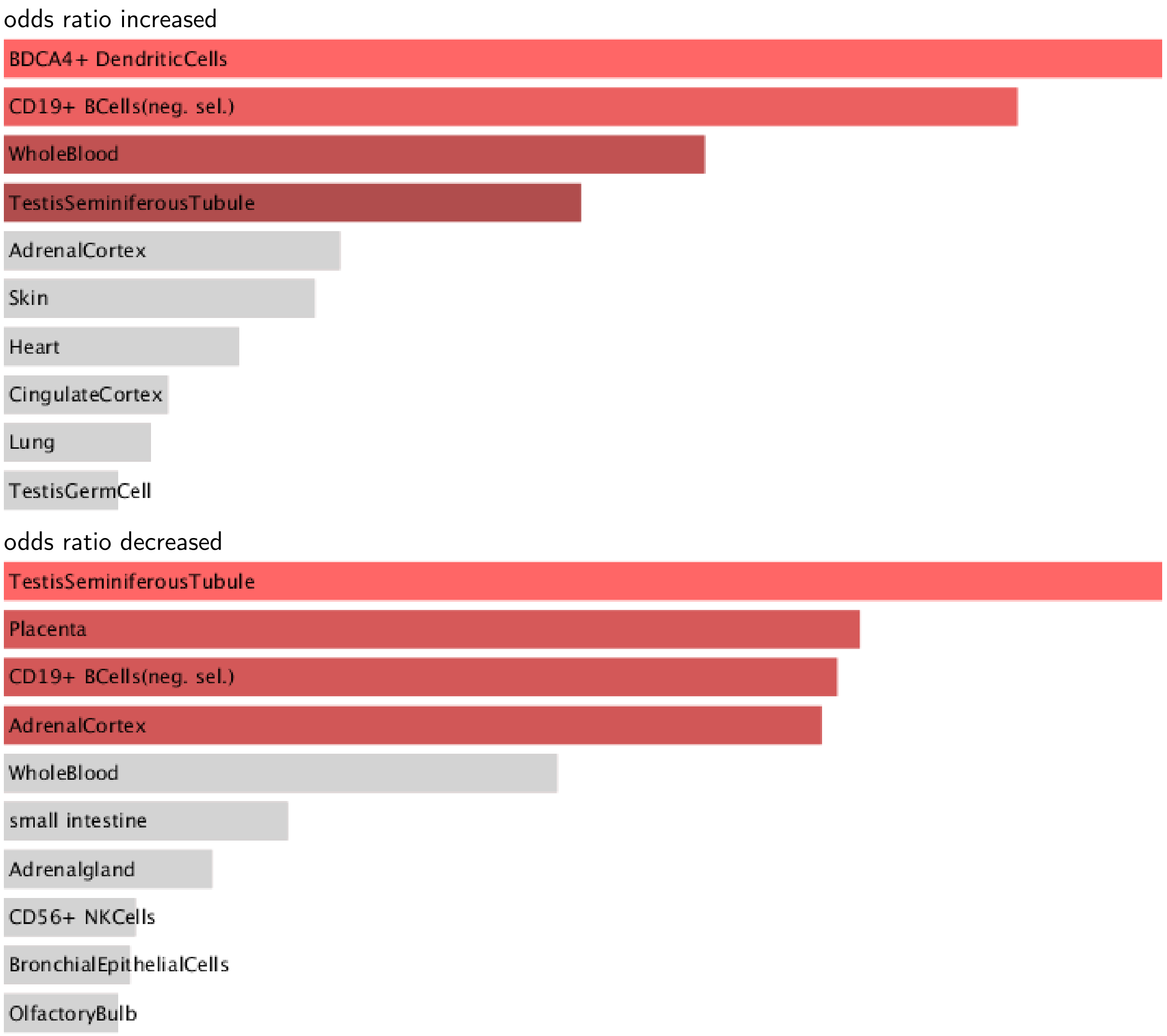

#### Table exome variants

- Sequence coordinates are using hg38 position
- Only results with P > -log10(8^10-7) are given for the exome analysis while all P > -log10(1^10-5´30) are truncated at this value
- Variants may appear twice with different traits

| | chr | pos | Gene | cDNA | Consequence | Case.MAF | Control.MAF | Odds.ratio | p | trait |
|---|---|---|---|---|---|---|---|---|---|---|
| 1 | 1 | 152312600 | FLG | c.2282_22... | frameshift | 0.0288 | 0.0232 | 1.2498 | 28.329940 | asthma |
| 2 | 1 | 152313385 | FLG | c.1501C>T | stop_gained | 0.0284 | 0.0232 | 1.2298 | 24.313006 | asthma |
| 3 | 1 | 167439433 | CD247 | n.208G>A | non_coding | 0.3994 | 0.4097 | 0.9580 | 10.217958 | asthma |
| 4 | 2 | 102226007 | IL1RL2 | c.1101T>C | synonymous | 0.1247 | 0.1183 | 1.0613 | 8.894149 | asthma |
| 5 | 2 | 102339008 | IL1RL1 | c.233C>A | missense | 0.2135 | 0.2261 | 0.9291 | 9.175289 | rhinitis |
| 6 | 2 | 102339008 | IL1RL1 | c.233C>A | missense | 0.2122 | 0.2257 | 0.9240 | 23.383419 | asthma |
| 7 | 2 | 102341256 | IL1RL1 | c.679C>T | synonymous | 0.3987 | 0.3771 | 1.0953 | 18.591251 | rhinitis |
| 8 | 2 | 102341256 | IL1RL1 | c.679C>T | synonymous | 0.3978 | 0.3779 | 1.0875 | 36.071450 | asthma |
| 9 | 2 | 102351547 | IL1RL1 | c.1297G>A | missense | 0.3604 | 0.3856 | 0.8977 | 25.315963 | rhinitis |
| 10 | 2 | 102351547 | IL1RL1 | c.1297G>A | missense | 0.3634 | 0.3845 | 0.9138 | 40.882397 | asthma |
| 11 | 2 | 102351615 | IL1RL1 | c.1365T>C | synonymous | 0.3613 | 0.3866 | 0.8977 | 25.518844 | rhinitis |
| 12 | 2 | 102351615 | IL1RL1 | c.1365T>C | synonymous | 0.3646 | 0.3855 | 0.9146 | 40.414652 | asthma |
| 13 | 2 | 102351751 | IL1RL1 | c.1501C>A | missense | 0.3608 | 0.3861 | 0.8977 | 25.472112 | rhinitis |
| 14 | 2 | 102351751 | IL1RL1 | c.1501C>A | missense | 0.3639 | 0.3850 | 0.9139 | 40.922996 | asthma |
| 15 | 2 | 102351752 | IL1RL1 | c.1502A>G | missense | 0.3608 | 0.3860 | 0.8976 | 25.460422 | rhinitis |
| 16 | 2 | 102351752 | IL1RL1 | c.1502A>G | missense | 0.3639 | 0.3849 | 0.9139 | 40.884057 | asthma |
| 17 | 2 | 102351825 | IL1RL1 | c.1575T>C | synonymous | 0.3591 | 0.3848 | 0.8960 | 25.962574 | rhinitis |
| 18 | 2 | 102351825 | IL1RL1 | c.1575T>C | synonymous | 0.3624 | 0.3836 | 0.9131 | 41.242680 | asthma |
| 19 | 2 | 102367819 | IL18R1 | c.59-6T>C | splice | 0.4119 | 0.3896 | 1.0973 | 19.610834 | rhinitis |
| 20 | 2 | 102367819 | IL18R1 | c.59-6T>C | splice | 0.4095 | 0.3905 | 1.0824 | 32.717831 | asthma |
| 21 | 2 | 241735388 | D2HGDH | c.164G>A | missense | 0.2978 | 0.3086 | 0.9503 | 12.546529 | asthma |
| 22 | 2 | 241748877 | D2HGDH | c.427C>T | missense | 0.4312 | 0.4478 | 0.9348 | 10.967784 | rhinitis |
| 23 | 2 | 241748877 | D2HGDH | c.427C>T | missense | 0.4351 | 0.4477 | 0.9501 | 14.562408 | asthma |
| 24 | 2 | 241748951 | D2HGDH | c.*191C>T | 3_prime_UTR | 0.4307 | 0.4473 | 0.9345 | 11.034046 | rhinitis |
| 25 | 2 | 241748951 | D2HGDH | c.*191C>T | 3_prime_UTR | 0.4347 | 0.4473 | 0.9503 | 14.415217 | asthma |
| 26 | 2 | 241749257 | D2HGDH | c.*497C>T | 3_prime_UTR | 0.4296 | 0.4461 | 0.9350 | 10.585863 | rhinitis |
| 27 | 2 | 241749257 | D2HGDH | c.*497C>T | 3_prime_UTR | 0.4336 | 0.4460 | 0.9507 | 13.782253 | asthma |
| 28 | 2 | 241751260 | D2HGDH | c.1012G>A | missense | 0.2404 | 0.2576 | 0.9120 | 15.237922 | rhinitis |

*(continued)*

| | chr | pos | Gene | cDNA | Consequence | Case.MAF | Control.MAF | Odds.ratio | p | trait |
|---|---|---|---|---|---|---|---|---|---|---|
| 29 | 2 | 241751260 | D2HGDH | c.1012G>A | missense | 0.2395 | 0.2575 | 0.9082 | 37.580209 | asthma |
| 30 | 2 | 241776965 | GAL3ST2 | c.10A>T | missense | 0.2785 | 0.2875 | 0.9566 | 9.291070 | asthma |
| 31 | 4 | 38773164 | TLR10 | c.2427T>C | synonymous | 0.1690 | 0.1815 | 0.9173 | 10.531800 | rhinitis |
| 32 | 4 | 38773164 | TLR10 | c.2427T>C | synonymous | 0.1707 | 0.1809 | 0.9320 | 16.088310 | asthma |
| 33 | 4 | 38773268 | TLR10 | c.2323A>G | missense | 0.1450 | 0.1571 | 0.9099 | 11.148680 | rhinitis |
| 34 | 4 | 38773268 | TLR10 | c.2323A>G | missense | 0.1477 | 0.1565 | 0.9343 | 13.480960 | asthma |
| 35 | 4 | 38773419 | TLR10 | c.2172T>C | synonymous | 0.1697 | 0.1822 | 0.9171 | 10.650334 | rhinitis |
| 36 | 4 | 38773419 | TLR10 | c.2172T>C | synonymous | 0.1715 | 0.1817 | 0.9320 | 16.158766 | asthma |
| 37 | 4 | 38774486 | TLR10 | c.1105A>C | missense | 0.2900 | 0.3005 | 0.9508 | 12.111596 | asthma |
| 38 | 4 | 38774559 | TLR10 | c.1032G>T | synonymous | 0.1691 | 0.1818 | 0.9159 | 10.881405 | rhinitis |
| 39 | 4 | 38774559 | TLR10 | c.1032G>T | synonymous | 0.1710 | 0.1812 | 0.9318 | 16.213036 | asthma |
| 40 | 4 | 38774870 | TLR10 | c.721A>C | missense | 0.2899 | 0.3005 | 0.9502 | 12.386475 | asthma |
| 41 | 4 | 38797027 | TLR1 | c.1805G>T | missense | 0.2082 | 0.2248 | 0.9065 | 15.234182 | rhinitis |
| 42 | 4 | 38797027 | TLR1 | c.1805G>T | missense | 0.2120 | 0.2241 | 0.9314 | 18.666956 | asthma |
| 43 | 4 | 38798089 | TLR1 | c.743A>G | missense | 0.1922 | 0.2057 | 0.9192 | 11.046289 | rhinitis |
| 44 | 4 | 38798089 | TLR1 | c.743A>G | missense | 0.1949 | 0.2051 | 0.9386 | 14.453828 | asthma |
| 45 | 4 | 122224596 | KIAA1109 | c.2712T>A | synonymous | 0.2789 | 0.2969 | 0.9161 | 15.100344 | rhinitis |
| 46 | 4 | 122271228 | KIAA1109 | c.7704T>C | synonymous | 0.2803 | 0.2981 | 0.9169 | 14.858550 | rhinitis |
| 47 | 4 | 122307977 | KIAA1109 | c.9870C>T | synonymous | 0.2992 | 0.3200 | 0.9070 | 19.252122 | rhinitis |
| 48 | 4 | 122307977 | KIAA1109 | c.9870C>T | synonymous | 0.3094 | 0.3191 | 0.9564 | 9.935168 | asthma |
| 49 | 4 | 122355806 | KIAA1109 | c.14316G>A | synonymous | 0.0789 | 0.0723 | 1.0992 | 14.310691 | asthma |
| 50 | 4 | 122359705 | KIAA1109 | c.14784T>C | synonymous | 0.2292 | 0.2151 | 1.0847 | 11.274660 | rhinitis |
| 51 | 4 | 122359705 | KIAA1109 | c.14784T>C | synonymous | 0.2246 | 0.2158 | 1.0526 | 10.449405 | asthma |
| 52 | 4 | 122456327 | IL2 | c.114G>T | synonymous | 0.3440 | 0.3255 | 1.0867 | 14.795609 | rhinitis |
| 53 | 4 | 122456327 | IL2 | c.114G>T | synonymous | 0.3366 | 0.3262 | 1.0481 | 11.165643 | asthma |
| 54 | 5 | 14610200 | FAM105A | c.957C>G | missense | 0.0810 | 0.0758 | 1.0741 | 8.735418 | asthma |
| 55 | 5 | 35955580 | UGT3A1 | c.*49G>C | 3_prime_UTR | 0.4268 | 0.4420 | 0.9400 | 8.867420 | rhinitis |
| 56 | 5 | 111103810 | WDR36 | c.790A>G | missense | 0.2956 | 0.3045 | 0.9582 | 8.937794 | asthma |
| 57 | 5 | 111121006 | WDR36 | c.2181A>T | synonymous | 0.2957 | 0.3047 | 0.9582 | 8.876475 | asthma |
| 58 | 5 | 132266473 | PDLIM4 | c.255T>C | synonymous | 0.3461 | 0.3369 | 0.9598 | 8.787280 | asthma |
| 59 | 5 | 132340627 | SLC22A4 | c.1507C>T | missense | 0.4204 | 0.4337 | 0.9471 | 16.210278 | asthma |
| 60 | 5 | 132484106 | IRF1 | n.4132A>G | non_coding | 0.3355 | 0.3255 | 1.0463 | 10.325139 | asthma |
| 61 | 5 | 132484108 | IRF1 | n.4130C>T | non_coding | 0.3402 | 0.3303 | 1.0453 | 10.005903 | asthma |

*(continued)*

| | chr | pos | Gene | cDNA | Consequence | Case.MAF | Control.MAF | Odds.ratio | p | trait |
|---|---|---|---|---|---|---|---|---|---|---|
| 62 | 5 | 132486174 | IRF1 | n.1180A>C | non_coding | 0.3434 | 0.3337 | 1.0440 | 9.453951 | asthma |
| 63 | 5 | 132486363 | IRF1 | c.555A>G | synonymous | 0.3447 | 0.3351 | 1.0438 | 9.574792 | asthma |
| 64 | 5 | 132486380 | IRF1 | c.545-7T>C | splice | 0.3435 | 0.3337 | 1.0448 | 9.905529 | asthma |
| 65 | 5 | 132486441 | AC116366.3 | c.-388T>C | 5_prime_UTR | 0.3447 | 0.3349 | 1.0446 | 9.903438 | asthma |
| 66 | 5 | 132486532 | AC116366.3 | c.-297T>C | 5_prime_UTR | 0.3452 | 0.3354 | 1.0443 | 9.803548 | asthma |
| 67 | 5 | 132579521 | AC116366.3 | n.730G>A | non_coding | 0.1966 | 0.1835 | 1.0892 | 24.424466 | asthma |
| 68 | 5 | 132660272 | IL13 | c.431A>G | missense | 0.1926 | 0.1783 | 0.9093 | 30.164690 | asthma |
| 69 | 5 | 132706454 | KIF3A | c.1234G>A | missense | 0.1369 | 0.1298 | 1.0635 | 10.176070 | asthma |
| 70 | 5 | 157509356 | ADAM19 | c.850A>G | missense | 0.3472 | 0.3376 | 1.0435 | 9.533429 | asthma |
| 71 | 6 | 26409662 | BTN3A1 | c.845G>C | missense | 0.1126 | 0.1066 | 1.0634 | 8.729321 | asthma |
| 72 | 6 | 26463346 | BTN2A1 | c.533G>T | missense | 0.1135 | 0.1073 | 1.0653 | 9.241845 | asthma |
| 73 | 6 | 26463347 | BTN2A1 | c.534G>T | missense | 0.1135 | 0.1073 | 1.0651 | 9.187154 | asthma |
| 74 | 6 | 26463432 | BTN2A1 | c.619G>A | missense | 0.1244 | 0.1181 | 1.0611 | 8.851397 | asthma |
| 75 | 6 | 27867440 | HIST1H1B | c.90C>T | synonymous | 0.1211 | 0.1146 | 1.0645 | 9.536406 | asthma |
| 76 | 6 | 27911422 | OR2B2 | c.898G>T | missense | 0.1189 | 0.1124 | 1.0655 | 9.641494 | asthma |
| 77 | 6 | 27912204 | OR2B2 | c.116T>C | missense | 0.1196 | 0.1130 | 1.0660 | 9.824488 | asthma |
| 78 | 6 | 28301047 | PGBD1 | c.1193A>G | missense | 0.1206 | 0.1141 | 1.0649 | 9.610302 | asthma |
| 79 | 6 | 28398374 | ZSCAN12 | c.32T>C | missense | 0.1199 | 0.1133 | 1.0657 | 9.775726 | asthma |
| 80 | 6 | 28575487 | ZBED9 | c.1218G>A | synonymous | 0.2401 | 0.2306 | 1.0542 | 11.540155 | asthma |
| 81 | 6 | 28586766 | ZBED9 | c.-49C>A | 5_prime_UTR | 0.2393 | 0.2300 | 1.0535 | 11.044842 | asthma |
| 82 | 6 | 28923399 | TRIM27 | c.234A>G | synonymous | 0.1238 | 0.1170 | 1.0665 | 10.261854 | asthma |
| 83 | 6 | 29374998 | OR12D3 | c.290C>T | missense | 0.1282 | 0.1211 | 1.0674 | 10.803548 | asthma |
| 84 | 6 | 29556180 | UBD | c.198T>C | synonymous | 0.1299 | 0.1223 | 1.0712 | 12.052664 | asthma |
| 85 | 6 | 29792099 | HCG4 | n.652C>T | non_coding | 0.3667 | 0.3570 | 1.0429 | 9.423428 | asthma |
| 86 | 6 | 29792108 | HCG4 | n.643C>T | non_coding | 0.3106 | 0.3015 | 1.0439 | 9.119529 | asthma |
| 87 | 6 | 29792146 | HCG4 | n.605A>G | non_coding | 0.3112 | 0.3021 | 1.0437 | 9.057298 | asthma |
| 88 | 6 | 29792219 | HLA-V | n.102-6T>C | splice | 0.3682 | 0.3585 | 1.0430 | 9.575282 | asthma |
| 89 | 6 | 29792334 | HCG4 | n.417C>G | non_coding | 0.3681 | 0.3584 | 1.0427 | 9.450016 | asthma |
| 90 | 6 | 29792411 | HCG4 | n.340C>T | non_coding | 0.3679 | 0.3581 | 1.0432 | 9.623423 | asthma |
| 91 | 6 | 29792575 | HCG4 | n.155_175... | non_coding | 0.3640 | 0.3540 | 1.0443 | 9.799971 | asthma |
| 92 | 6 | 29829862 | HLA-G | c.942C>T | synonymous | 0.3115 | 0.3022 | 1.0448 | 9.488518 | asthma |
| 93 | 6 | 29888649 | HLA-H | n.532C>G | non_coding | 0.0523 | 0.0459 | 1.1469 | 8.792635 | rhinitis |
| 94 | 6 | 29942982 | HLA-A | c.299T>C | missense | 0.2880 | 0.2756 | 1.0628 | 16.091140 | asthma |
| 95 | 6 | 30064745 | ZNRD1 | c.*48A>C | 3_prime_UTR | 0.1370 | 0.1282 | 1.0790 | 14.865504 | asthma |

*(continued)*

| | chr | pos | Gene | cDNA | Consequence | Case.MAF | Control.MAF | Odds.ratio | p | trait |
|---|---|---|---|---|---|---|---|---|---|---|
| 96 | 6 | 30108978 | TRIM31 | n.361C>T | non_coding | 0.4256 | 0.4140 | 1.0486 | 12.227165 | asthma |
| 97 | 6 | 30110498 | TRIM31 | c.694G>A | missense | 0.1801 | 0.1704 | 1.0698 | 14.977572 | asthma |
| 98 | 6 | 30110553 | TRIM31 | c.639G>A | synonymous | 0.1789 | 0.1693 | 1.0689 | 14.520713 | asthma |
| 99 | 6 | 30112497 | TRIM31 | c.309C>T | synonymous | 0.1837 | 0.1742 | 1.0673 | 14.232918 | asthma |
| 100 | 6 | 30112719 | TRIM31 | c.87C>T | synonymous | 0.1803 | 0.1707 | 1.0684 | 14.441291 | asthma |
| 101 | 6 | 30198489 | TRIM26 | c.474G>A | synonymous | 0.2416 | 0.2301 | 1.0656 | 16.417482 | asthma |
| 102 | 6 | 30199109 | TRIM26 | c.-6G>A | 5_prime_UTR | 0.1697 | 0.1606 | 1.0684 | 13.735418 | asthma |
| 103 | 6 | 30490287 | HLA-E | c.382G>A | missense | 0.3887 | 0.3983 | 1.0408 | 8.931072 | asthma |
| 104 | 6 | 30491388 | HLA-E | c.862C>T | missense | 0.0156 | 0.0187 | 0.8313 | 12.765736 | asthma |
| 105 | 6 | 30547266 | GNL1 | c.1287A>G | synonymous | 0.1525 | 0.1433 | 1.0764 | 15.488651 | asthma |
| 106 | 6 | 30643573 | ATAT1 | c.*558C>A | 3_prime_UTR | 0.1528 | 0.1434 | 1.0776 | 15.999132 | asthma |
| 107 | 6 | 30685004 | PPP1R18 | c.1015G>A | missense | 0.1530 | 0.1436 | 1.0774 | 15.924453 | asthma |
| 108 | 6 | 30730764 | FLOT1 | n.761T>C | non_coding | 0.2067 | 0.1961 | 1.0681 | 15.815025 | asthma |
| 109 | 6 | 30744028 | IER3 | c.379G>C | missense | 0.0875 | 0.0934 | 1.0744 | 9.778847 | asthma |
| 110 | 6 | 30951614 | DPCR1 | c.3150G>A | synonymous | 0.1712 | 0.1616 | 1.0717 | 15.119587 | asthma |
| 111 | 6 | 30952347 | DPCR1 | c.1255G>A | missense | 0.1712 | 0.1616 | 1.0719 | 15.201626 | asthma |
| 112 | 6 | 31025536 | MUC22 | c.105C>T | synonymous | 0.1895 | 0.1806 | 1.0608 | 12.105130 | asthma |
| 113 | 6 | 31025722 | MUC22 | c.291C>T | synonymous | 0.3769 | 0.3874 | 1.0454 | 10.715344 | asthma |
| 114 | 6 | 31025756 | MUC22 | c.325A>G | missense | 0.4550 | 0.4656 | 1.0437 | 10.460548 | asthma |
| 115 | 6 | 31026089 | MUC22 | c.658A>T | missense | 0.1164 | 0.1239 | 0.9314 | 12.037157 | asthma |
| 116 | 6 | 31026103 | MUC22 | c.672G>C | missense | 0.1270 | 0.1343 | 0.9373 | 10.905529 | asthma |
| 117 | 6 | 31027401 | MUC22 | c.1970C>T | missense | 0.1051 | 0.1120 | 0.9313 | 11.200935 | asthma |
| 118 | 6 | 31027410 | MUC22 | c.1979T>C | missense | 0.1052 | 0.1120 | 0.9322 | 10.926282 | asthma |
| 119 | 6 | 31027520 | MUC22 | c.2089A>G | missense | 0.1052 | 0.1121 | 0.9316 | 11.100234 | asthma |
| 120 | 6 | 31027755 | MUC22 | c.2324T>C | missense | 0.1061 | 0.1128 | 0.9333 | 10.650917 | asthma |
| 121 | 6 | 31028355 | MUC22 | c.2924C>T | missense | 0.1061 | 0.1130 | 0.9322 | 11.015653 | asthma |
| 122 | 6 | 31028713 | MUC22 | c.3282C>T | synonymous | 0.1055 | 0.1124 | 0.9314 | 11.205303 | asthma |
| 123 | 6 | 31028732 | MUC22 | c.3301G>A | missense | 0.1043 | 0.1109 | 0.9334 | 10.453087 | asthma |
| 124 | 6 | 31029301 | MUC22 | c.3870T>C | synonymous | 0.1108 | 0.1181 | 0.9306 | 11.970616 | asthma |
| 125 | 6 | 31029557 | MUC22 | c.4126A>G | missense | 0.1051 | 0.1119 | 0.9321 | 10.946537 | asthma |
| 126 | 6 | 31029575 | MUC22 | c.4144A>G | missense | 0.1101 | 0.1175 | 0.9288 | 12.462433 | asthma |
| 127 | 6 | 31030039 | MUC22 | c.4608G>T | synonymous | 0.1533 | 0.1618 | 0.9379 | 12.389021 | asthma |
| 128 | 6 | 31030047 | MUC22 | c.4616T>C | missense | 0.1116 | 0.1192 | 0.9283 | 12.778325 | asthma |

*(continued)*

| | chr | pos | Gene | cDNA | Consequence | Case.MAF | Control.MAF | Odds.ratio | p | trait |
|---|---|---|---|---|---|---|---|---|---|---|
| 129 | 6 | 31032362 | MUC22 | c.4836C>T | synonymous | 0.1060 | 0.1128 | 0.9322 | 11.004233 | asthma |
| 130 | 6 | 31111867 | C6orf15 | c.492C>T | synonymous | 0.1611 | 0.1510 | 1.0803 | 17.706858 | asthma |
| 131 | 6 | 31112217 | C6orf15 | c.142G>A | missense | 0.0660 | 0.0728 | 0.8993 | 16.203842 | asthma |
| 132 | 6 | 31112239 | C6orf15 | c.120G>C | missense | 0.2049 | 0.1956 | 1.0598 | 12.408601 | asthma |
| 133 | 6 | 31116036 | CDSN | c.1579A>G | missense | 0.2467 | 0.2594 | 1.0697 | 18.771600 | asthma |
| 134 | 6 | 31116271 | CDSN | c.1344T>C | synonymous | 0.2853 | 0.2711 | 1.0732 | 22.340369 | asthma |
| 135 | 6 | 31117165 | CDSN | c.447_449... | disruptiv... | 0.2845 | 0.2704 | 1.0725 | 21.808270 | asthma |
| 136 | 6 | 31117187 | CDSN | c.428A>G | missense | 0.2506 | 0.2631 | 1.0681 | 18.456801 | asthma |
| 137 | 6 | 31117423 | CDSN | c.192T>C | synonymous | 0.2506 | 0.2632 | 1.0685 | 18.621602 | asthma |
| 138 | 6 | 31117492 | CDSN | c.123T>C | synonymous | 0.2501 | 0.2628 | 1.0686 | 18.615109 | asthma |
| 139 | 6 | 31117579 | PSORS1C1 | n.2586C>T | non_coding | 0.1597 | 0.1492 | 1.0831 | 17.635637 | asthma |
| 140 | 6 | 31120368 | CDSN | c.52A>T | missense | 0.2798 | 0.2648 | 1.0786 | 25.097780 | asthma |
| 141 | 6 | 31138682 | PSORS1C1 | c.70C>A | missense | 0.0730 | 0.0813 | 0.8902 | 21.071963 | asthma |
| 142 | 6 | 31138723 | PSORS1C1 | c.118dupC | frameshift | 0.0770 | 0.0862 | 0.8854 | 24.149721 | asthma |
| 143 | 6 | 31144707 | CCHCR1 | c.1880G>A | missense | 0.0587 | 0.0653 | 0.8935 | 16.391902 | asthma |
| 144 | 6 | 31151121 | CCHCR1 | c.536T>A | missense&... | 0.0584 | 0.0652 | 0.8900 | 17.439735 | asthma |
| 145 | 6 | 31154538 | CCHCR1 | c.492C>G | missense | 0.4637 | 0.4538 | 1.0407 | 9.235973 | asthma |
| 146 | 6 | 31157480 | CCHCR1 | c.121G>T | stop_gained | 0.0591 | 0.0658 | 0.8909 | 17.289883 | asthma |
| 147 | 6 | 31157625 | CCHCR1 | c.-25G>C | 5_prime_UTR | 0.0590 | 0.0657 | 0.8914 | 17.075359 | asthma |
| 148 | 6 | 31161533 | TCF19 | c.325C>T | missense | 0.1617 | 0.1515 | 1.0807 | 17.964971 | asthma |
| 149 | 6 | 31161930 | TCF19 | c.722C>T | missense | 0.0591 | 0.0659 | 0.8899 | 17.628563 | asthma |
| 150 | 6 | 31164637 | POU5F1 | c.1047C>T | synonymous | 0.3067 | 0.3208 | 1.0673 | 20.194771 | asthma |
| 151 | 6 | 31164872 | POU5F1 | c.817-5T>G | splice | 0.3060 | 0.3199 | 1.0670 | 19.939302 | asthma |
| 152 | 6 | 31165179 | POU5F1 | c.765C>G | synonymous | 0.0591 | 0.0659 | 0.8906 | 17.445753 | asthma |
| 153 | 6 | 31165732 | POU5F1 | c.-93C>T | 5_prime_UTR | 0.3058 | 0.3196 | 1.0665 | 19.635261 | asthma |
| 154 | 6 | 31165886 | POU5F1 | c.-247A>G | 5_prime_U... | 0.0588 | 0.0655 | 0.8920 | 16.867740 | asthma |
| 155 | 6 | 31166093 | POU5F1 | c.-229C>T | 5_prime_U... | 0.0593 | 0.0661 | 0.8904 | 17.539403 | asthma |
| 156 | 6 | 31166166 | POU5F1 | c.-302G>T | 5_prime_U... | 0.3079 | 0.3217 | 1.0663 | 19.779892 | asthma |
| 157 | 6 | 31166341 | TCF19 | c.620_622... | disruptiv... | 0.2585 | 0.2691 | 1.0559 | 12.846795 | asthma |
| 158 | 6 | 31170600 | POU5F1 | c.21G>A | synonymous | 0.4620 | 0.4523 | 1.0397 | 8.832092 | asthma |
| 159 | 6 | 31271830 | HLA-C | c.112C>T | missense | 0.0719 | 0.0781 | 0.9138 | 11.616723 | asthma |
| 160 | 6 | 31355235 | MIR6891 | n.82C>T | non_coding | 0.2444 | 0.2344 | 1.0566 | 11.714443 | asthma |
| 161 | 6 | 31355544 | HLA-B | c.668C>T | missense | 0.1683 | 0.1586 | 1.0736 | 15.059932 | asthma |

*(continued)*

| | chr | pos | Gene | cDNA | Consequence | Case.MAF | Control.MAF | Odds.ratio | p | trait |
|---|---|---|---|---|---|---|---|---|---|---|
| 162 | 6 | 31355560 | HLA-B | c.652A>G | missense | 0.1065 | 0.1160 | 0.9082 | 8.977984 | rhinitis |
| 163 | 6 | 31355560 | HLA-B | c.652A>G | missense | 0.1083 | 0.1160 | 0.9258 | 13.255551 | asthma |
| 164 | 6 | 31355632 | HLA-B | n.171A>G | non_coding | 0.1917 | 0.2012 | 1.0622 | 12.863279 | asthma |
| 165 | 6 | 31355639 | HLA-B | n.164C>G | non_coding | 0.1563 | 0.1452 | 1.0903 | 21.318397 | asthma |
| 166 | 6 | 31356638 | HLA-B | n.414T>G | non_coding | 0.2177 | 0.2071 | 1.0655 | 13.804100 | asthma |
| 167 | 6 | 31403653 | MICA | c.21T>C | synonymous | 0.1984 | 0.2076 | 1.0583 | 11.872571 | asthma |
| 168 | 6 | 31410581 | MICA | c.70T>G | missense | 0.1423 | 0.1537 | 0.9138 | 22.540909 | asthma |
| 169 | 6 | 31410610 | MICA | c.99T>C | synonymous | 0.2450 | 0.2581 | 0.9330 | 20.027334 | asthma |
| 170 | 6 | 31410648 | MICA | c.137A>G | missense | 0.2449 | 0.2579 | 0.9331 | 19.970616 | asthma |
| 171 | 6 | 31411200 | MICA | c.454G>A | missense | 0.2458 | 0.2590 | 0.9325 | 20.425621 | asthma |
| 172 | 6 | 31411332 | MICA | c.586G>A | missense | 0.2457 | 0.2588 | 0.9330 | 20.119587 | asthma |
| 173 | 6 | 31411996 | MICA | c.336C>T | synonymous | 0.2447 | 0.2580 | 0.9318 | 20.713993 | asthma |
| 174 | 6 | 31412017 | MICA | c.357C>T | synonymous | 0.2449 | 0.2581 | 0.9325 | 20.322941 | asthma |
| 175 | 6 | 31412018 | MICA | c.358A>G | missense | 0.2449 | 0.2580 | 0.9326 | 20.234481 | asthma |
| 176 | 6 | 31412030 | MICA | c.370C>T | missense | 0.2450 | 0.2582 | 0.9322 | 20.493766 | asthma |
| 177 | 6 | 31412040 | MICA | c.380T>C | missense | 0.3914 | 0.4017 | 0.9581 | 10.138764 | asthma |
| 178 | 6 | 31412046 | MICA | c.386C>G | missense | 0.2455 | 0.2587 | 0.9326 | 20.338187 | asthma |
| 179 | 6 | 31412154 | MICA | c.494G>A | missense | 0.3917 | 0.4020 | 0.9580 | 10.205721 | asthma |
| 180 | 6 | 31412314 | MICA | c.566-4dupT | splice | 0.2431 | 0.2565 | 0.9314 | 20.759201 | asthma |
| 181 | 6 | 31412384 | MICA | c.625_626... | frameshift | 0.1108 | 0.1201 | 0.9125 | 17.718058 | asthma |
| 182 | 6 | 31505769 | MICB | c.223A>G | missense | 0.2240 | 0.2352 | 1.0653 | 15.904831 | asthma |
| 183 | 6 | 31505784 | MICB | c.238A>G | missense | 0.3160 | 0.3058 | 1.0486 | 11.163802 | asthma |
| 184 | 6 | 31506180 | MICB | c.363C>G | missense | 0.1519 | 0.1413 | 1.0888 | 20.307153 | asthma |
| 185 | 6 | 31506223 | MICB | c.406G>A | missense | 0.3161 | 0.3060 | 1.0485 | 11.122859 | asthma |
| 186 | 6 | 31509904 | MICB | c.1147A>G | missense | 0.2550 | 0.2685 | 1.0722 | 20.655215 | asthma |
| 187 | 6 | 31529929 | MCCD1 | c.354C>T | synonymous | 0.2442 | 0.2572 | 1.0714 | 19.767512 | asthma |
| 188 | 6 | 31530467 | DDX39B | n.393dupG | non_coding | 0.1453 | 0.1346 | 1.0931 | 21.230401 | asthma |
| 189 | 6 | 31531452 | DDX39B | n.293G>A | non_coding | 0.0224 | 0.0258 | 0.8642 | 10.656001 | asthma |
| 190 | 6 | 31538871 | DDX39B | c.408C>A | synonymous | 0.2425 | 0.2554 | 1.0709 | 19.268170 | asthma |
| 191 | 6 | 31546470 | ATP6V1G2 | c.82+8T>C | splice | 0.1451 | 0.1347 | 1.0911 | 20.346498 | asthma |
| 192 | 6 | 31558135 | NFKBIL1 | c.625C>T | missense | 0.0814 | 0.0880 | 1.0892 | 12.823330 | asthma |
| 193 | 6 | 31558303 | NFKBIL1 | c.793G>A | missense | 0.0124 | 0.0152 | 0.8110 | 13.204398 | asthma |
| 194 | 6 | 31572916 | LTA | n.336G>C | non_coding | 0.0227 | 0.0260 | 0.8715 | 10.221415 | asthma |
| 195 | 6 | 31573007 | LTA | c.179C>A | missense | 0.3695 | 0.3587 | 1.0477 | 11.518414 | asthma |

*(continued)*

| | chr | pos | Gene | cDNA | Consequence | Case.MAF | Control.MAF | Odds.ratio | p | trait |
|---|---|---|---|---|---|---|---|---|---|---|
| 196 | 6 | 31616064 | AIF1 | c.-48C>G | 5_prime_UTR | 0.0662 | 0.0591 | 1.1298 | 19.730954 | asthma |
| 197 | 6 | 31626851 | PRRC2A | c.1062T>A | synonymous | 0.1400 | 0.1472 | 1.0600 | 9.673255 | asthma |
| 198 | 6 | 31632329 | PRRC2A | c.3656C>A | missense | 0.0127 | 0.0157 | 0.8082 | 13.989700 | asthma |
| 199 | 6 | 31636267 | PRRC2A | c.5683T>G | missense | 0.0783 | 0.0844 | 1.0842 | 11.143997 | asthma |
| 200 | 6 | 31636578 | PRRC2A | c.5904C>A | synonymous | 0.0655 | 0.0587 | 1.1253 | 18.390192 | asthma |
| 201 | 6 | 31719231 | LY6G6C | c.243C>T | synonymous | 0.1800 | 0.1907 | 0.9314 | 17.031144 | asthma |
| 202 | 6 | 31724609 | MPIG6B | c.432C>G | missense | 0.0714 | 0.0613 | 1.1775 | 16.215240 | rhinitis |
| 203 | 6 | 31760120 | MSH5 | c.1716C>T | synonymous | 0.1408 | 0.1305 | 1.0918 | 20.318849 | asthma |
| 204 | 6 | 31765689 | VWA7 | c.2581A>G | missense | 0.1409 | 0.1305 | 1.0927 | 20.718285 | asthma |
| 205 | 6 | 31765982 | VWA7 | c.2400C>T | synonymous | 0.0200 | 0.0237 | 0.8400 | 14.388170 | asthma |
| 206 | 6 | 31766568 | VWA7 | c.2079C>A | synonymous | 0.1410 | 0.1306 | 1.0931 | 20.868061 | asthma |
| 207 | 6 | 31772986 | VWA7 | c.1041_10... | frameshift | 0.0210 | 0.0249 | 0.8397 | 15.107516 | asthma |
| 208 | 6 | 31781043 | VARS | c.2625C>T | synonymous | 0.1424 | 0.1513 | 0.9308 | 14.505845 | asthma |
| 209 | 6 | 31795066 | VARS | c.152C>G | missense | 0.4275 | 0.4179 | 1.0398 | 8.703774 | asthma |
| 210 | 6 | 31810169 | HSPA1L | c.1804G>A | missense | 0.3168 | 0.3071 | 1.0463 | 10.223371 | asthma |
| 211 | 6 | 31810495 | HSPA1L | c.1478C>T | missense | 0.1828 | 0.1924 | 1.0652 | 13.761452 | asthma |
| 212 | 6 | 31862816 | NEU1 | c.-40T>G | 5_prime_UTR | 0.0265 | 0.0307 | 0.8570 | 14.671213 | asthma |
| 213 | 6 | 31947158 | CFB | c.450A>G | synonymous | 0.1890 | 0.1983 | 1.0612 | 12.534023 | asthma |
| 214 | 6 | 31962574 | SKIV2L | c.1200A>G | synonymous | 0.0272 | 0.0315 | 0.8621 | 13.994391 | asthma |
| 215 | 6 | 31967790 | SKIV2L | c.2659G>A | missense | 0.0273 | 0.0314 | 0.8640 | 13.586030 | asthma |
| 216 | 6 | 31967973 | SKIV2L | c.2749G>A | missense | 0.0493 | 0.0418 | 1.1896 | 13.060281 | rhinitis |
| 217 | 6 | 31968902 | SKIV2L | c.3212C>T | missense | 0.1394 | 0.1291 | 1.0927 | 9.190912 | rhinitis |
| 218 | 6 | 31969260 | SKIV2L | n.586C>T | non_coding | 0.0604 | 0.0523 | 1.1656 | 12.406050 | rhinitis |
| 219 | 6 | 31970240 | DXO | n.1683delC | non_coding | 0.0154 | 0.0183 | 0.8387 | 11.487983 | asthma |
| 220 | 6 | 31970858 | DXO | n.1066C>G | non_coding | 0.0615 | 0.0530 | 1.1695 | 13.138227 | rhinitis |
| 221 | 6 | 31971568 | DXO | c.108C>T | synonymous | 0.0156 | 0.0184 | 0.8421 | 11.094798 | asthma |
| 222 | 6 | 31980623 | STK19 | n.233T>G | non_coding | 0.0151 | 0.0178 | 0.8434 | 10.571217 | asthma |
| 223 | 6 | 31980644 | STK19 | c.902-6_9... | splice | 0.1413 | 0.1311 | 1.0907 | 8.914709 | rhinitis |
| 224 | 6 | 32007072 | CYP21A1P | n.1141C>G | non_coding | 0.1626 | 0.1495 | 1.1043 | 12.240106 | rhinitis |
| 225 | 6 | 32053637 | TNXB | c.8542G>A | missense | 0.0187 | 0.0223 | 0.8352 | 14.397831 | asthma |
| 226 | 6 | 32056126 | TNXB | c.8192C>G | missense | 0.0617 | 0.0534 | 1.1655 | 12.660747 | rhinitis |
| 227 | 6 | 32056618 | TNXB | c.8111G>A | missense | 0.0549 | 0.0594 | 0.9195 | 8.818442 | asthma |
| 228 | 6 | 32058086 | TNXB | c.7797G>A | synonymous | 0.1807 | 0.1700 | 1.0768 | 17.846185 | asthma |

*(continued)*

| | chr | pos | Gene | cDNA | Consequence | Case.MAF | Control.MAF | Odds.ratio | p | trait |
|---|---|---|---|---|---|---|---|---|---|---|
| 229 | 6 | 32058330 | TNXB | c.7553G>A | missense | 0.2787 | 0.2959 | 1.0879 | 31.731422 | asthma |
| 230 | 6 | 32061428 | TNXB | c.7461C>T | synonymous | 0.1394 | 0.1290 | 1.0940 | 21.052370 | asthma |
| 231 | 6 | 32061449 | TNXB | c.7440T>C | synonymous | 0.3085 | 0.3230 | 1.0697 | 21.753994 | asthma |
| 232 | 6 | 32061654 | TNXB | c.7235C>T | missense | 0.0545 | 0.0591 | 0.9167 | 9.365825 | asthma |
| 233 | 6 | 32062152 | TNXB | c.7168+5G>A | splice | 0.0512 | 0.0435 | 1.1879 | 13.288868 | rhinitis |
| 234 | 6 | 32067917 | TNXB | c.6288G>A | synonymous | 0.1398 | 0.1293 | 1.0942 | 21.173472 | asthma |
| 235 | 6 | 32084667 | TNXB | c.3191G>A | missense | 0.0137 | 0.0166 | 0.8215 | 12.810790 | asthma |
| 236 | 6 | 32096949 | TNXB | c.904A>G | missense | 0.1392 | 0.1289 | 1.0926 | 20.472241 | asthma |
| 237 | 6 | 32121077 | ATF6B | c.603C>G | synonymous | 0.2726 | 0.2635 | 1.0472 | 9.734946 | asthma |
| 238 | 6 | 32126145 | ATF6B | c.441C>T | synonymous | 0.0668 | 0.0582 | 1.1584 | 12.605198 | rhinitis |
| 239 | 6 | 32128224 | ATF6B | c.-17G>T | 5_prime_UTR | 0.1384 | 0.1283 | 1.0915 | 19.875496 | asthma |
| 240 | 6 | 32154609 | PPT2 | c.15C>G | missense | 0.1344 | 0.1418 | 1.0635 | 10.386158 | asthma |
| 241 | 6 | 32154695 | PPT2 | c.101C>A | missense | 0.2411 | 0.2324 | 1.0495 | 9.823041 | asthma |
| 242 | 6 | 32166733 | EGFL8 | c.257G>A | missense | 0.2416 | 0.2329 | 1.0493 | 9.787812 | asthma |
| 243 | 6 | 32167051 | EGFL8 | n.415C>T | non_coding | 0.0144 | 0.0172 | 0.8322 | 11.739452 | asthma |
| 244 | 6 | 32167360 | EGFL8 | c.612G>A | synonymous | 0.0654 | 0.0573 | 1.1511 | 25.538952 | asthma |
| 245 | 6 | 32183666 | AGER | c.244G>A | missense | 0.0713 | 0.0628 | 1.1463 | 26.071963 | asthma |
| 246 | 6 | 32184217 | AGER | c.6T>A | synonymous | 0.1372 | 0.1440 | 1.0586 | 9.108351 | asthma |
| 247 | 6 | 32186508 | PBX2 | n.668T>C | non_coding | 0.2682 | 0.2596 | 1.0454 | 8.851397 | asthma |
| 248 | 6 | 32187221 | PBX2 | n.1072G>A | non_coding | 0.2608 | 0.2514 | 1.0503 | 10.616005 | asthma |
| 249 | 6 | 32188712 | PBX2 | n.105C>G | non_coding | 0.0683 | 0.0609 | 1.1296 | 20.176983 | asthma |
| 250 | 6 | 32200994 | NOTCH4 | c.4152C>A | synonymous | 0.1594 | 0.1494 | 1.0792 | 17.138227 | asthma |
| 251 | 6 | 32201155 | NOTCH4 | c.4101G>A | synonymous | 0.0489 | 0.0414 | 1.1913 | 13.177832 | rhinitis |
| 252 | 6 | 32201368 | NOTCH4 | c.3888C>T | synonymous | 0.0654 | 0.0568 | 1.1603 | 28.322119 | asthma |
| 253 | 6 | 32216928 | NOTCH4 | n.2007C>T | non_coding | 0.2697 | 0.2796 | 1.0508 | 11.177244 | asthma |
| 254 | 6 | 32222613 | NOTCH4 | c.349A>C | missense | 0.4084 | 0.3924 | 1.0692 | 23.800519 | asthma |
| 255 | 6 | 32222629 | NOTCH4 | c.333T>C | synonymous | 0.4191 | 0.4027 | 1.0704 | 24.664542 | asthma |
| 256 | 6 | 32222707 | NOTCH4 | c.255C>T | synonymous | 0.0773 | 0.0827 | 0.9294 | 9.150765 | asthma |
| 257 | 6 | 32223881 | NOTCH4 | c.42_47du... | disruptiv... | 0.1411 | 0.1315 | 1.0855 | 17.780154 | asthma |
| 258 | 6 | 32293475 | C6orf10 | c.1192A>C | missense | 0.1377 | 0.1281 | 1.0873 | 18.200797 | asthma |
| 259 | 6 | 32293994 | C6orf10 | c.673T>C | missense | 0.3791 | 0.3667 | 1.0547 | 15.006740 | asthma |
| 260 | 6 | 32335915 | C6orf10 | c.442A>T | missense | 0.1353 | 0.1259 | 1.0858 | 17.257589 | asthma |
| 261 | 6 | 32366178 | C6orf10 | c.206A>G | missense | 0.3842 | 0.3720 | 1.0533 | 14.240861 | asthma |

*(continued)*

| | chr | pos | Gene | cDNA | Consequence | Case.MAF | Control.MAF | Odds.ratio | p | trait |
|---|---|---|---|---|---|---|---|---|---|---|
| 262 | 6 | 32369909 | C6orf10 | c.88T>C | missense | 0.2608 | 0.2443 | 0.9164 | 13.741123 | rhinitis |
| 263 | 6 | 32371721 | C6orf10 | c.-15G>C | 5_prime_UTR | 0.0171 | 0.0203 | 0.8363 | 13.040577 | asthma |
| 264 | 6 | 32394964 | BTNL2 | c.1140G>A | missense | 0.1320 | 0.1452 | 0.8949 | 31.664943 | asthma |
| 265 | 6 | 32394968 | BTNL2 | c.1136C>T | missense | 0.1328 | 0.1459 | 0.8962 | 31.070785 | asthma |
| 266 | 6 | 32396039 | BTNL2 | c.1078A>G | missense&... | 0.4603 | 0.4503 | 1.0411 | 9.167810 | asthma |
| 267 | 6 | 32396067 | BTNL2 | c.1050G>A | synonymous | 0.3206 | 0.2971 | 1.1168 | 50.000000 | asthma |
| 268 | 6 | 32396178 | BTNL2 | c.939A>G | synonymous | 0.4574 | 0.4470 | 1.0428 | 10.087406 | asthma |
| 269 | 6 | 32403017 | BTNL2 | c.627G>A | synonymous | 0.0556 | 0.0603 | 0.9170 | 9.455932 | asthma |
| 270 | 6 | 32403039 | BTNL2 | c.605C>T | missense | 0.0585 | 0.0640 | 0.9083 | 12.025396 | asthma |
| 271 | 6 | 32403058 | BTNL2 | c.586A>G | missense | 0.3818 | 0.3636 | 1.0806 | 30.815025 | asthma |
| 272 | 6 | 32403102 | BTNL2 | c.542G>A | missense | 0.0549 | 0.0596 | 0.9165 | 9.439137 | asthma |
| 273 | 6 | 32403131 | BTNL2 | c.513A>T | synonymous | 0.0546 | 0.0594 | 0.9151 | 9.702896 | asthma |
| 274 | 6 | 32405086 | BTNL2 | c.280T>A | missense | 0.0564 | 0.0611 | 0.9183 | 9.300596 | asthma |
| 275 | 6 | 32405186 | BTNL2 | c.180C>T | synonymous | 0.0563 | 0.0611 | 0.9176 | 9.433327 | asthma |
| 276 | 6 | 32439932 | HLA-DRA | c.-19C>A | 5_prime_UTR | 0.3803 | 0.3623 | 1.0801 | 30.184157 | asthma |
| 277 | 6 | 32443258 | HLA-DRA | c.329-2A>C | splice_ac... | 0.4273 | 0.4389 | 1.0486 | 12.536704 | asthma |
| 278 | 6 | 32637532 | HLA-DQA1 | c.74A>G | missense | 0.0231 | 0.0263 | 0.8761 | 8.833570 | asthma |
| 279 | 6 | 32642029 | HLA-DQA1 | c.389C>T | missense | 0.2401 | 0.2285 | 1.0672 | 16.707966 | asthma |
| 280 | 6 | 32642175 | HLA-DQA1 | c.535T>C | missense | 0.2470 | 0.2359 | 1.0622 | 14.633390 | asthma |
| 281 | 6 | 32660883 | HLA-DQB1 | c.773-1A>G | splice_ac... | 0.0215 | 0.0262 | 0.8174 | 9.050074 | rhinitis |
| 282 | 6 | 32660883 | HLA-DQB1 | c.773-1A>G | splice_ac... | 0.0223 | 0.0262 | 0.8481 | 14.133594 | asthma |
| 283 | 6 | 32661352 | HLA-DQB1 | c.767A>G | missense | 0.1403 | 0.1511 | 0.9168 | 9.208730 | rhinitis |
| 284 | 6 | 32661352 | HLA-DQB1 | c.767A>G | missense | 0.1354 | 0.1512 | 0.8794 | 43.232177 | asthma |
| 285 | 6 | 32661378 | HLA-DQB1 | c.741G>T | synonymous | 0.1414 | 0.1522 | 0.9171 | 9.235450 | rhinitis |
| 286 | 6 | 32661378 | HLA-DQB1 | c.741G>T | synonymous | 0.1365 | 0.1523 | 0.8797 | 43.459420 | asthma |
| 287 | 6 | 32661480 | HLA-DQB1 | n.3728A>C | non_coding | 0.0281 | 0.0321 | 0.8731 | 11.988853 | asthma |
| 288 | 6 | 32661939 | HLA-DQB1 | n.3269A>G | non_coding | 0.2650 | 0.2505 | 1.0790 | 23.858864 | asthma |
| 289 | 6 | 32661941 | HLA-DQB1 | n.3267C>T | non_coding | 0.1379 | 0.1486 | 0.9166 | 8.982549 | rhinitis |
| 290 | 6 | 32661941 | HLA-DQB1 | n.3267C>T | non_coding | 0.1331 | 0.1487 | 0.8784 | 42.583859 | asthma |
| 291 | 6 | 32661960 | HLA-DQB1 | c.661+7G>A | splice | 0.1380 | 0.1490 | 0.9141 | 9.600153 | rhinitis |
| 292 | 6 | 32661960 | HLA-DQB1 | c.661+7G>A | splice | 0.1334 | 0.1491 | 0.8782 | 43.232251 | asthma |
| 293 | 6 | 32746077 | HLA-DQA2 | c.613+5G>A | splice | 0.0576 | 0.0629 | 0.9106 | 11.313185 | asthma |
| 294 | 6 | 32746306 | HLA-DQA2 | c.680T>C | missense | 0.0895 | 0.0801 | 1.1286 | 11.319665 | rhinitis |
| 295 | 6 | 32746391 | HLA-DQA2 | c.765A>G | synonymous | 0.0896 | 0.0801 | 1.1294 | 11.451365 | rhinitis |

*(continued)*

| | chr | pos | Gene | cDNA | Consequence | Case.MAF | Control.MAF | Odds.ratio | p | trait |
|---|---|---|---|---|---|---|---|---|---|---|
| 296 | 6 | 32758976 | HLA-DQB2 | c.520C>T | missense | 0.0139 | 0.0164 | 0.8458 | 9.500725 | asthma |
| 297 | 6 | 32813180 | HLA-DOB | c.*36G>A | 3_prime_UTR | 0.0674 | 0.0585 | 1.1637 | 29.909037 | asthma |
| 298 | 6 | 32828876 | TAP2 | c.2088G>T | synonymous | 0.2325 | 0.2482 | 0.9174 | 29.584526 | asthma |
| 299 | 6 | 32828908 | TAP2 | c.2059T>C | stop_lost | 0.2322 | 0.2478 | 0.9180 | 29.033952 | asthma |
| 300 | 6 | 32828974 | TAP2 | c.1993A>G | missense | 0.2331 | 0.2487 | 0.9179 | 29.278272 | asthma |
| 301 | 6 | 32829520 | TAP2 | c.1812A>G | synonymous | 0.2329 | 0.2484 | 0.9186 | 28.844057 | asthma |
| 302 | 6 | 32830032 | TAP2 | c.1693G>A | missense | 0.1079 | 0.0986 | 1.1050 | 20.737312 | asthma |
| 303 | 6 | 32830771 | TAP2 | c.1308C>T | synonymous | 0.1058 | 0.1122 | 0.9360 | 9.812197 | asthma |
| 304 | 6 | 32832635 | TAP2 | c.1135G>A | missense | 0.1472 | 0.1397 | 1.0636 | 10.850165 | asthma |
| 305 | 6 | 32837530 | TAP2 | c.608+7G>A | splice | 0.3043 | 0.2937 | 1.0519 | 12.312382 | asthma |
| 306 | 6 | 32842666 | PSMB8 | c.395+6C>T | splice | 0.4674 | 0.4542 | 1.0543 | 15.576263 | asthma |
| 307 | 6 | 32857313 | PSMB9 | c.179G>A | missense | 0.2891 | 0.2775 | 1.0590 | 15.109915 | asthma |
| 308 | 6 | 32976317 | BRD2 | c.678G>C | synonymous | 0.1029 | 0.0956 | 1.0848 | 13.588380 | asthma |
| 309 | 6 | 32980649 | BRD2 | c.2337C>T | synonymous | 0.1042 | 0.0973 | 1.0794 | 12.211973 | asthma |
| 310 | 9 | 6255967 | IL33 | c.613-1G>C | splice_ac... | 0.0029 | 0.0047 | 0.6240 | 17.423198 | asthma |
| 311 | 11 | 61783884 | MYRF | c.3033T>C | synonymous | 0.3407 | 0.3504 | 0.9580 | 9.686344 | asthma |
| 312 | 12 | 56042145 | RPS26 | c.-22C>G | 5_prime_U... | 0.4408 | 0.4259 | 0.9412 | 8.951558 | rhinitis |
| 313 | 12 | 56042145 | RPS26 | c.-22C>G | 5_prime_U... | 0.4395 | 0.4262 | 0.9476 | 15.896196 | asthma |
| 314 | 12 | 56083910 | ERBB3 | c.234+8A>T | splice | 0.4397 | 0.4279 | 0.9528 | 13.032358 | asthma |
| 315 | 12 | 57104437 | STAT6 | n.405A>G | non_coding | 0.0045 | 0.0059 | 0.7562 | 8.960983 | asthma |
| 316 | 12 | 57141483 | LRP1 | c.300C>T | synonymous | 0.1407 | 0.1491 | 0.9341 | 13.041102 | asthma |
| 317 | 12 | 71125316 | TSPAN8 | c.*18C>G | 3_prime_UTR | 0.4009 | 0.4115 | 0.9572 | 10.392223 | asthma |
| 318 | 12 | 71139754 | TSPAN8 | c.218G>C | missense | 0.3982 | 0.4089 | 0.9562 | 11.111091 | asthma |
| 319 | 15 | 67236036 | AAGAB | c.394A>C | missense | 0.3139 | 0.3259 | 0.9462 | 14.936667 | asthma |
| 320 | 16 | 27226713 | NSMCE1 | c.600+7G>A | splice | 0.3611 | 0.3512 | 1.0439 | 9.576099 | asthma |
| 321 | 16 | 27226789 | NSMCE1 | c.531G>A | synonymous | 0.3637 | 0.3544 | 1.0413 | 8.798603 | asthma |
| 322 | 16 | 27344882 | IL4R | c.178A>G | missense | 0.4612 | 0.4515 | 1.0400 | 8.932557 | asthma |
| 323 | 16 | 27345038 | IL4R | n.700C>T | splice | 0.3686 | 0.3562 | 1.0550 | 14.951558 | asthma |
| 324 | 17 | 4632019 | ALOX15 | c.1679C>T | missense | 0.0166 | 0.0214 | 0.7726 | 11.609065 | rhinitis |
| 325 | 17 | 39657827 | STARD3 | c.350G>A | missense | 0.3452 | 0.3305 | 0.9366 | 21.407601 | asthma |
| 326 | 17 | 39666058 | TCAP | c.453A>C | synonymous | 0.2775 | 0.2635 | 0.9315 | 22.176526 | asthma |
| 327 | 17 | 39674647 | PGAP3 | c.465T>C | synonymous | 0.3161 | 0.2997 | 0.9261 | 27.670602 | asthma |
| 328 | 17 | 39727784 | ERBB2 | c.3508C>G | missense | 0.3360 | 0.3200 | 0.9302 | 25.534320 | asthma |

*(continued)*

| | chr | pos | Gene | cDNA | Consequence | Case.MAF | Control.MAF | Odds.ratio | p | trait |
|---|---|---|---|---|---|---|---|---|---|---|
| 329 | 17 | 39766006 | IKZF3 | c.1080C>T | synonymous | 0.4585 | 0.4813 | 0.9125 | 45.298259 | asthma |
| 330 | 17 | 39864166 | IKZF3 | c.-40A>G | 5_prime_UTR | 0.4947 | 0.4784 | 0.9368 | 10.445269 | rhinitis |
| 331 | 17 | 39864166 | IKZF3 | c.-40A>G | 5_prime_UTR | 0.4974 | 0.4786 | 0.9082 | 49.755723 | asthma |
| 332 | 17 | 39868373 | ZPBP2 | c.19C>T | synonymous | 0.4544 | 0.4784 | 0.9080 | 50.000000 | asthma |
| 333 | 17 | 39872381 | ZPBP2 | c.518G>T | missense | 0.4967 | 0.4810 | 0.9393 | 9.748362 | rhinitis |
| 334 | 17 | 39872381 | ZPBP2 | c.518G>T | missense | 0.4948 | 0.4812 | 0.9085 | 49.846490 | asthma |
| 335 | 17 | 39875421 | ZPBP2 | c.876C>T | synonymous | 0.4540 | 0.4783 | 0.9069 | 50.000000 | asthma |
| 336 | 17 | 39905943 | GSDMB | c.892C>T | missense | 0.4469 | 0.4717 | 0.9052 | 50.000000 | asthma |
| 337 | 17 | 39905964 | GSDMB | c.871G>A | missense | 0.4968 | 0.4878 | 0.9404 | 9.355561 | rhinitis |
| 338 | 17 | 39905964 | GSDMB | c.871G>A | missense | 0.4872 | 0.4880 | 0.9054 | 50.000000 | asthma |
| 339 | 17 | 39965740 | GSDMA | c.53G>A | missense | 0.4656 | 0.4467 | 1.0792 | 31.557834 | asthma |
| 340 | 17 | 39966427 | GSDMA | c.382G>T | missense | 0.4891 | 0.4933 | 0.9320 | 26.919013 | asthma |
| 341 | 17 | 39966433 | GSDMA | c.388G>A | missense | 0.3216 | 0.3356 | 0.9384 | 19.294393 | asthma |
| 342 | 17 | 39974934 | GSDMA | c.941C>A | missense | 0.4843 | 0.4682 | 1.0665 | 22.652085 | asthma |
| 343 | 17 | 39981111 | PSMD3 | c.141G>A | synonymous | 0.4445 | 0.4576 | 0.9484 | 15.701365 | asthma |
| 344 | 17 | 40016890 | CSF3 | c.555G>A | synonymous | 0.3624 | 0.3757 | 0.9446 | 16.975514 | asthma |
| 345 | 17 | 40020003 | MED24 | c.237T>A | synonymous | 0.3606 | 0.3741 | 0.9435 | 16.757459 | asthma |
| 346 | 17 | 40023239 | MED24 | c.2103T>C | synonymous | 0.3635 | 0.3764 | 0.9460 | 16.148375 | asthma |
| 347 | 17 | 40026837 | MED24 | n.313C>A | non_coding | 0.3707 | 0.3836 | 0.9466 | 15.887060 | asthma |
| 348 | 17 | 49406866 | PHB | n.608C>G | non_coding | 0.3424 | 0.3328 | 1.0441 | 9.526075 | asthma |
| 349 | 22 | 41437112 | TOB2 | c.234G>A | synonymous | 0.2106 | 0.2026 | 0.9523 | 9.182369 | asthma |
| 350 | 22 | 41507809 | ACO2 | c.192A>C | synonymous | 0.2092 | 0.2011 | 1.0505 | 9.293965 | asthma |
| 351 | 22 | 41743074 | MEI1 | c.1332-6A>G | splice | 0.1867 | 0.1790 | 0.9497 | 9.274742 | asthma |
| 352 | 22 | 41763225 | MEI1 | c.2172G>T | synonymous | 0.1880 | 0.1805 | 0.9510 | 8.912574 | asthma |

#### Figures genome context

- From top: Cytogenetic bands were obtained from http://genome.ucsc.edu
- Gene transcripts were downloaded from http://www.ensembl.org/biomart/martview
- GWAS ß values obtained from http://genepi.qimr.edu.au/staff/manuelf/gwas_results/SHARE-without23andMe.LDSCORE-GC.SE-META.v0(A. Sample does not include 23andme samples
- Only results with P > -log10(8^10-7) are given for the exome analysis while all P > -log10(1^10-30) are truncated at this value

#### Fig S13 AAGAB

alpha and gamma adaptin binding protein [Source:HGNC Symbol;Acc:HGNC:25662]

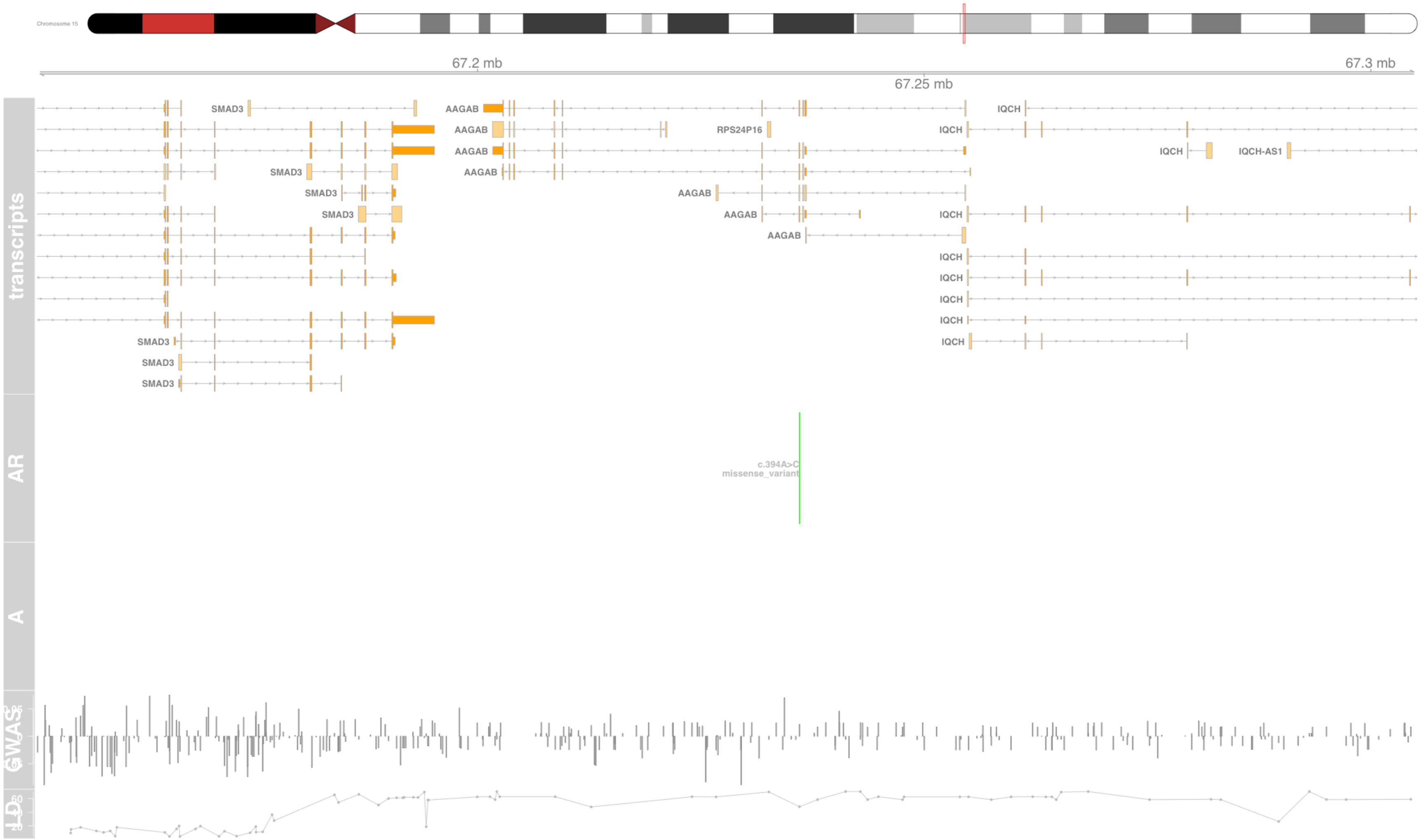

#### Fig S14 ACO2

aconitase 2 [Source:HGNC Symbol;Acc:HGNC:118]

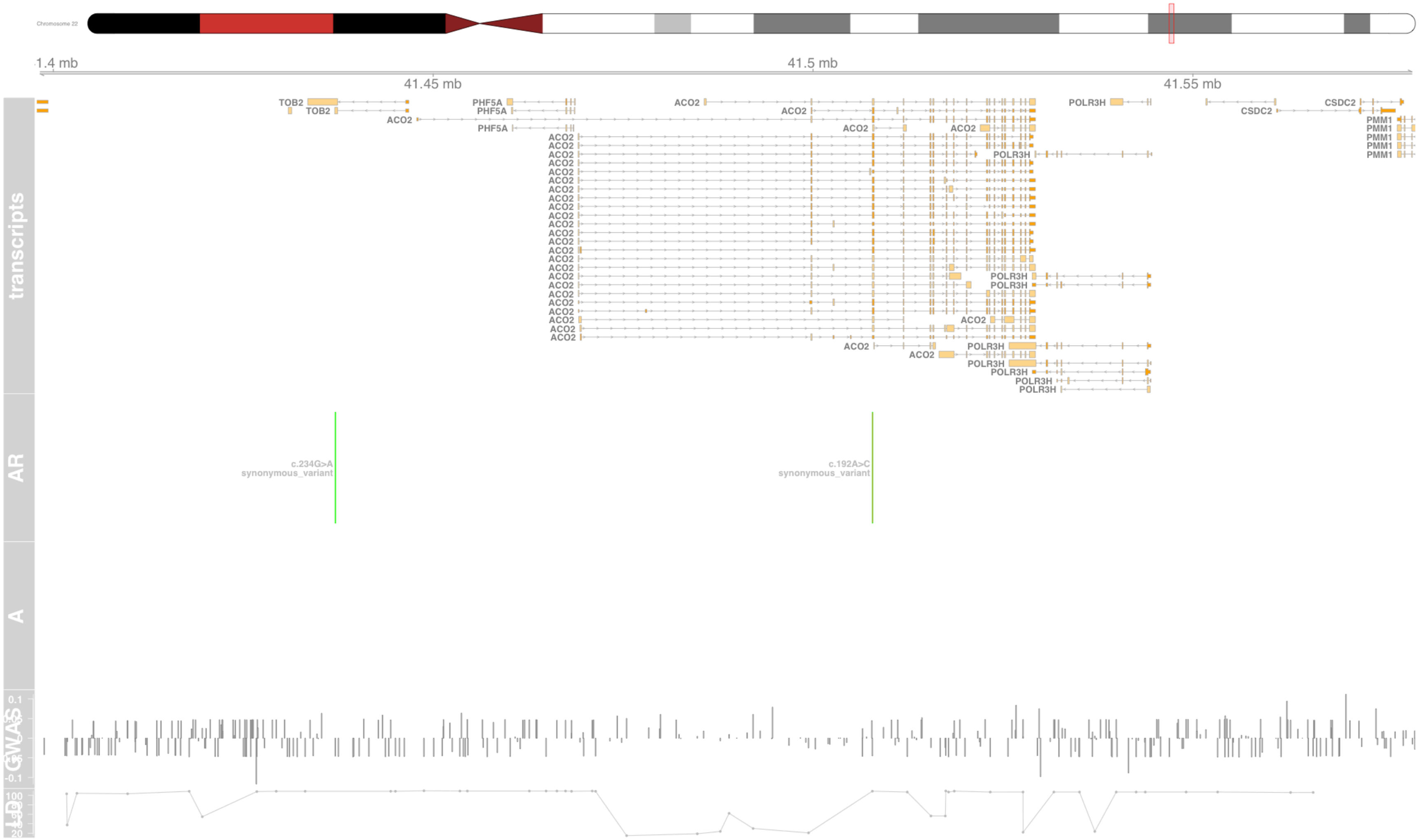

#### Fig S15 ADAM19

ADAM metallopeptidase domain 19 [Source:HGNC Symbol;Acc:HGNC:197]

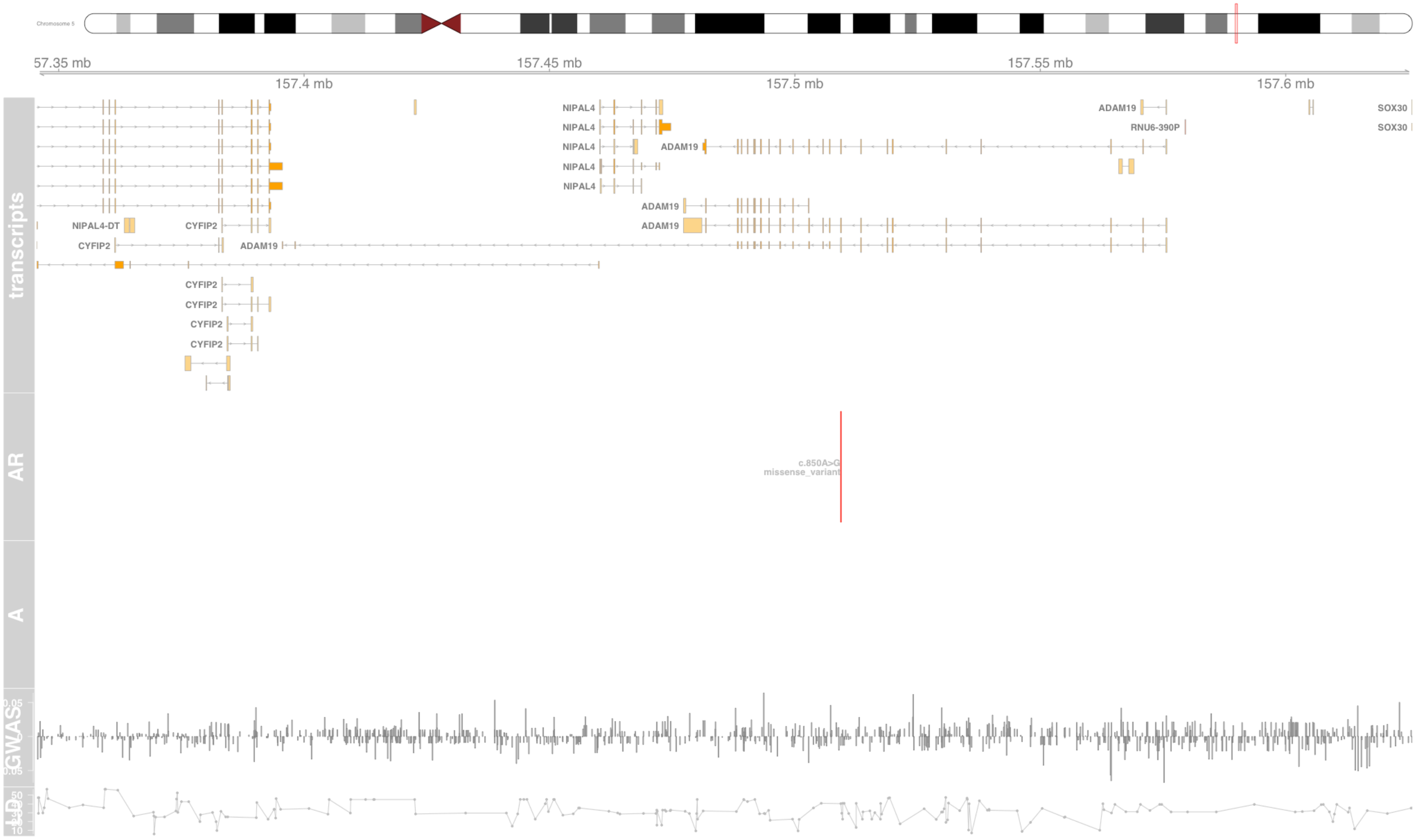

#### Fig S16 AGER

advanced glycosylation end-product specific receptor [Source:HGNC Symbol;Acc:HGNC:320]

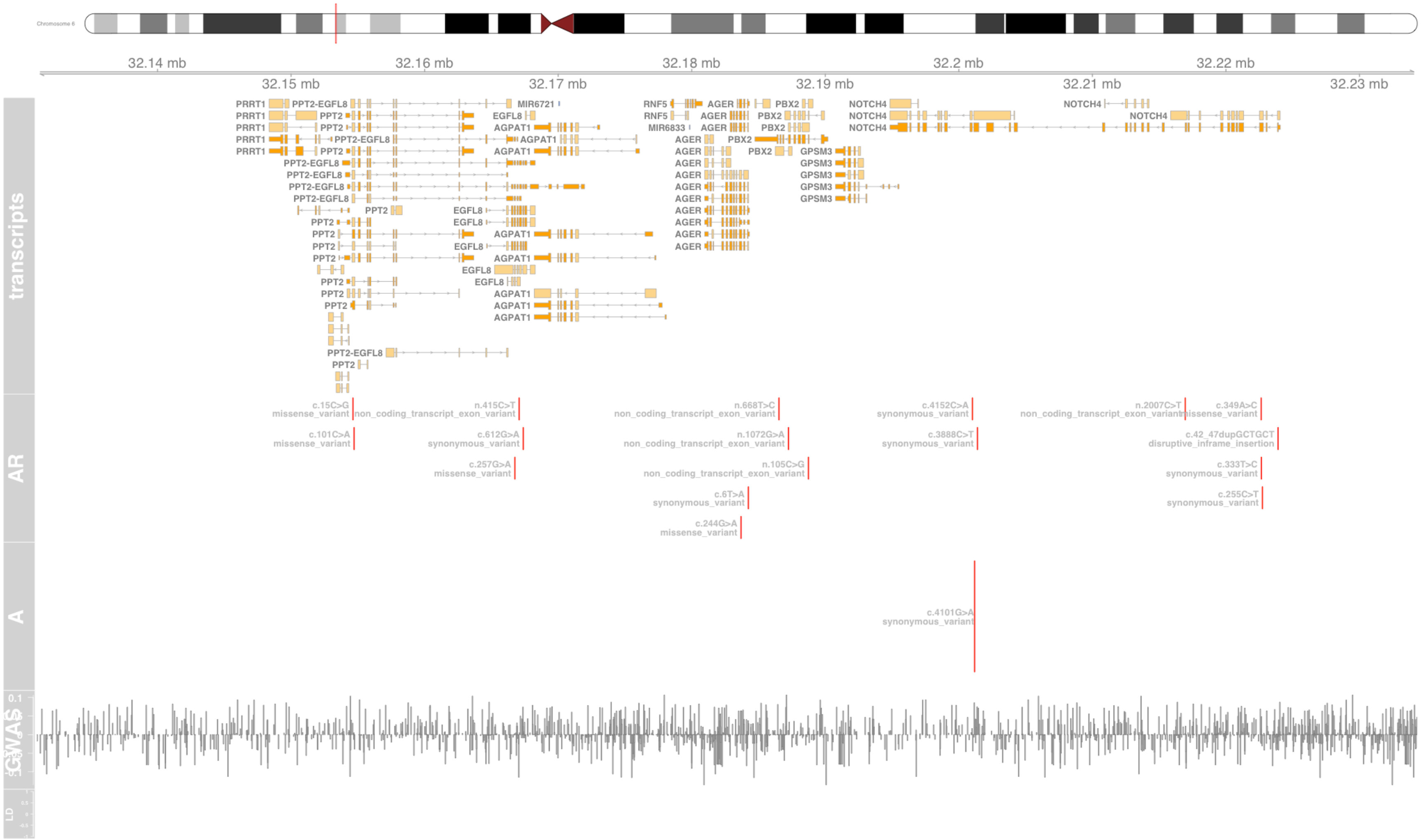

#### Fig S17 AIF1

allograft inflammatory factor 1 [Source:HGNC Symbol;Acc:HGNC:352]

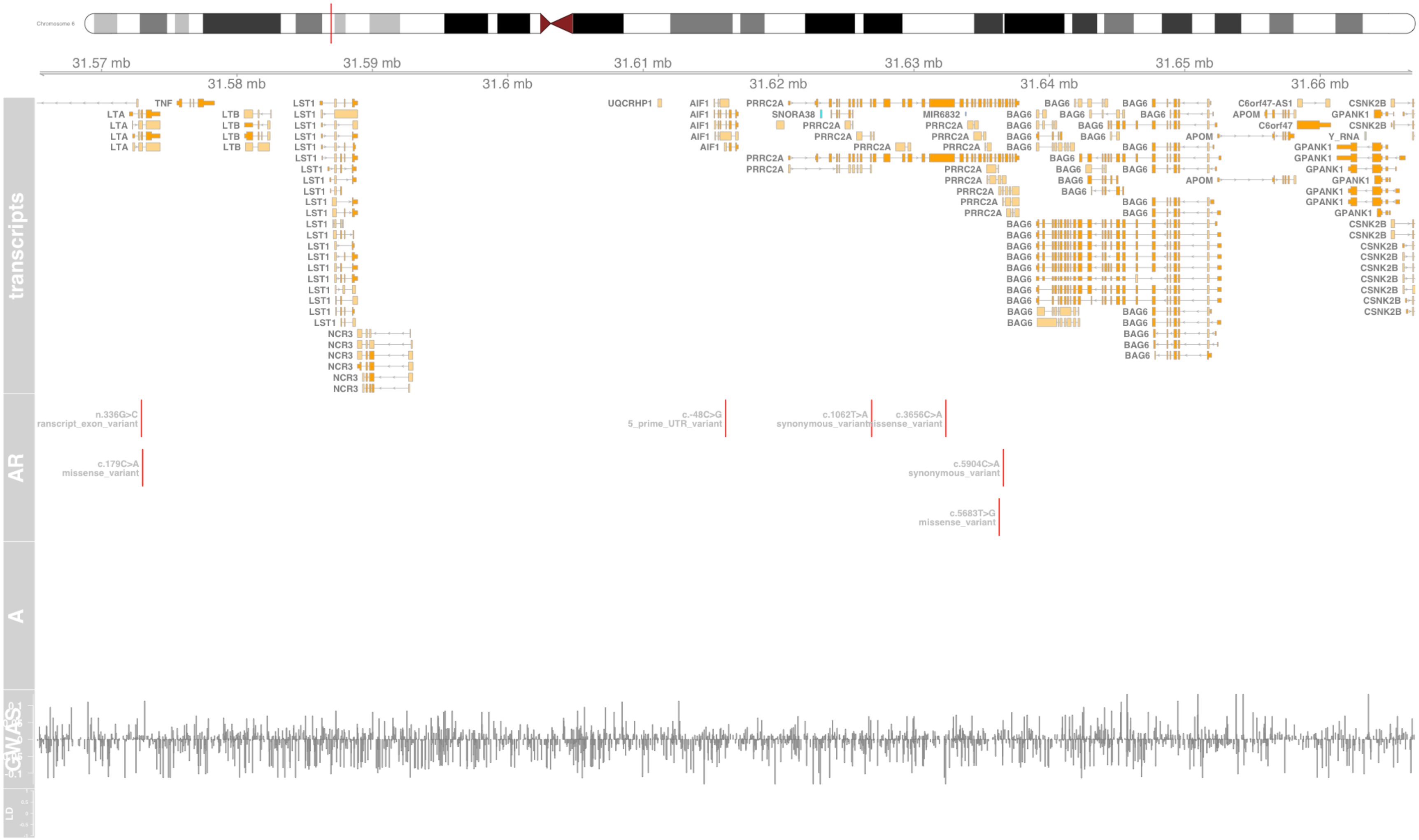

#### Fig S18 ALOX15

arachidonate 15-lipoxygenase [Source:HGNC Symbol;Acc:HGNC:433]

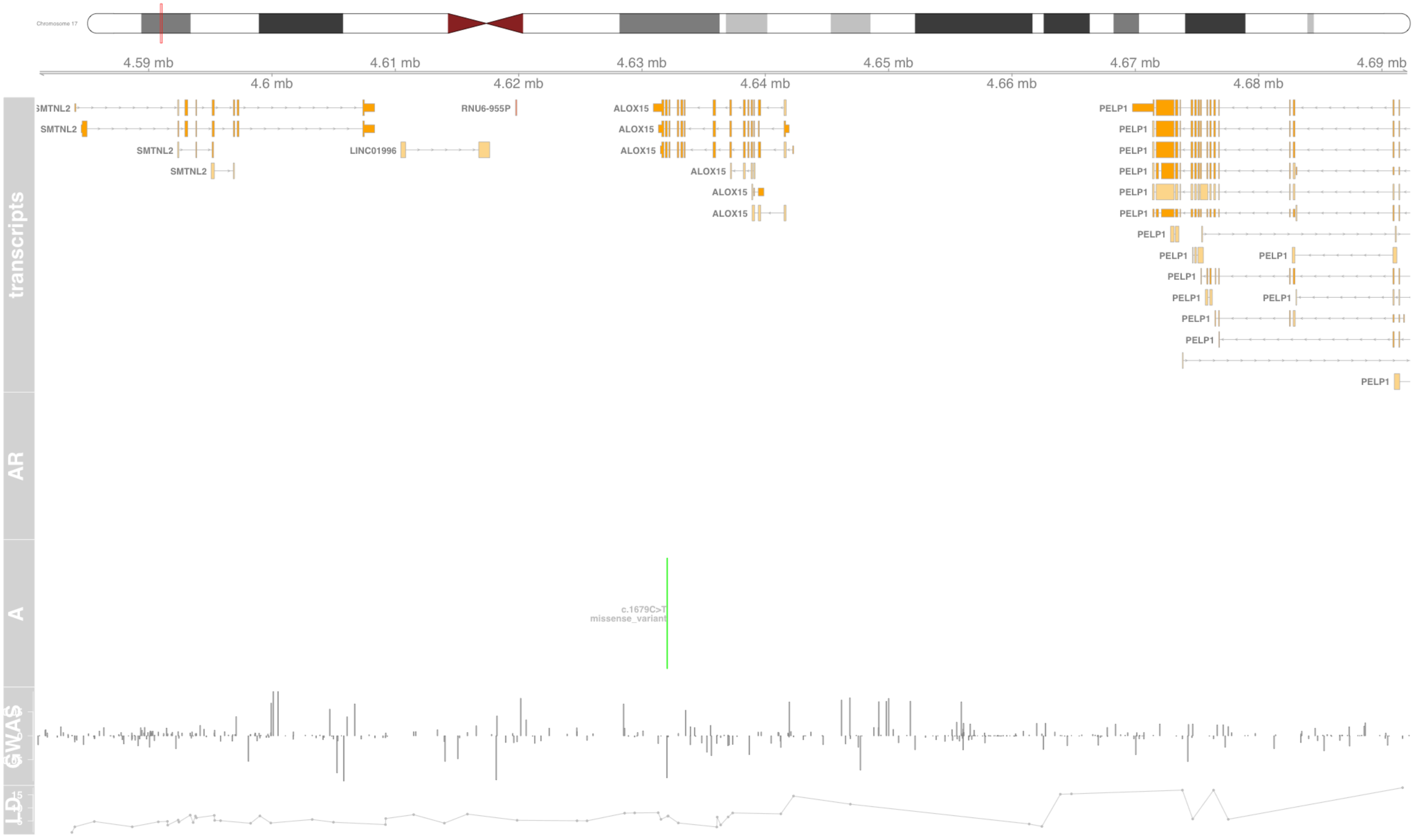

#### Fig S19 ATAT1

alpha tubulin acetyltransferase 1 [Source:HGNC Symbol;Acc:HGNC:21186]

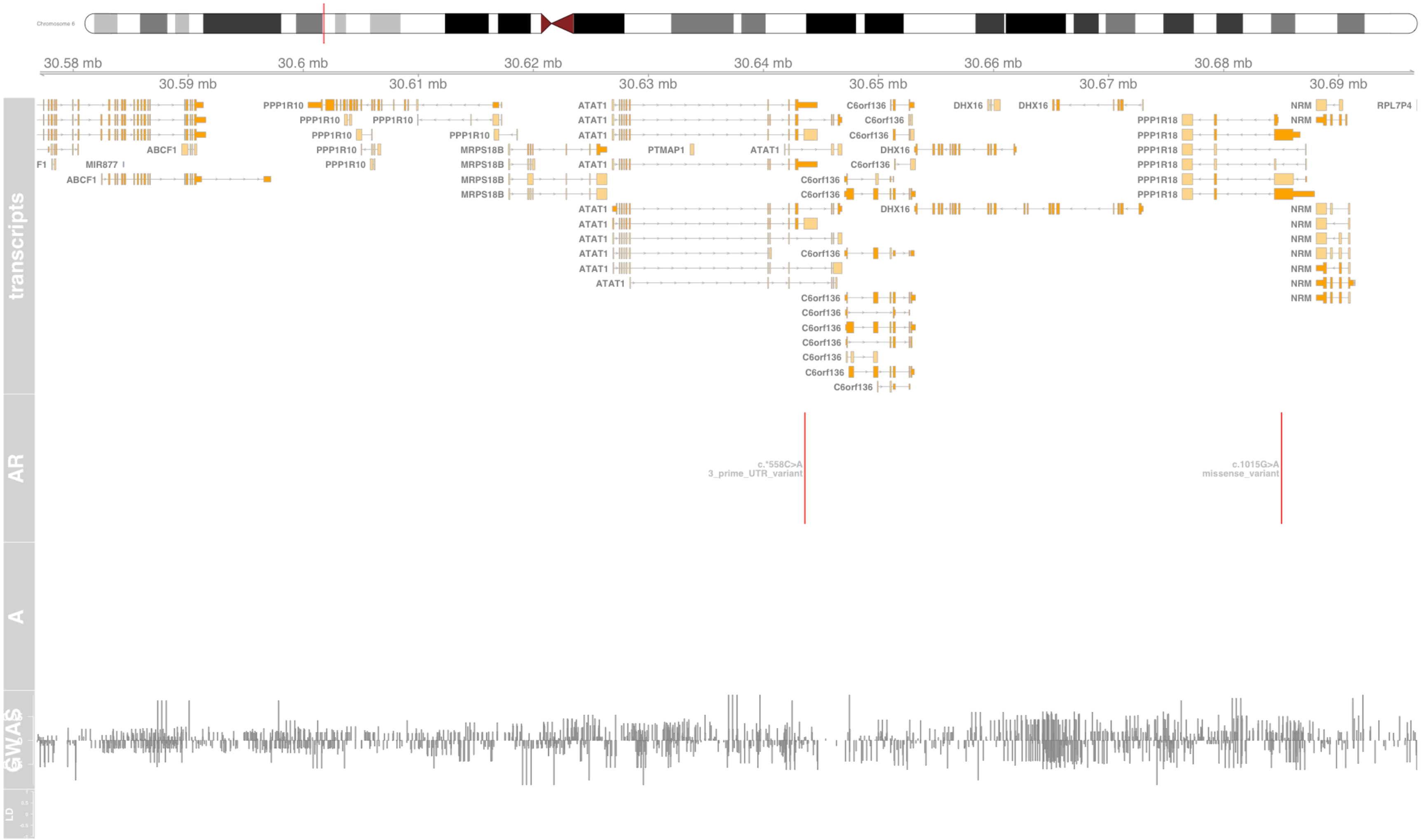

#### Fig S20 ATF6B

activating transcription factor 6 beta [Source:HGNC Symbol;Acc:HGNC:2349]

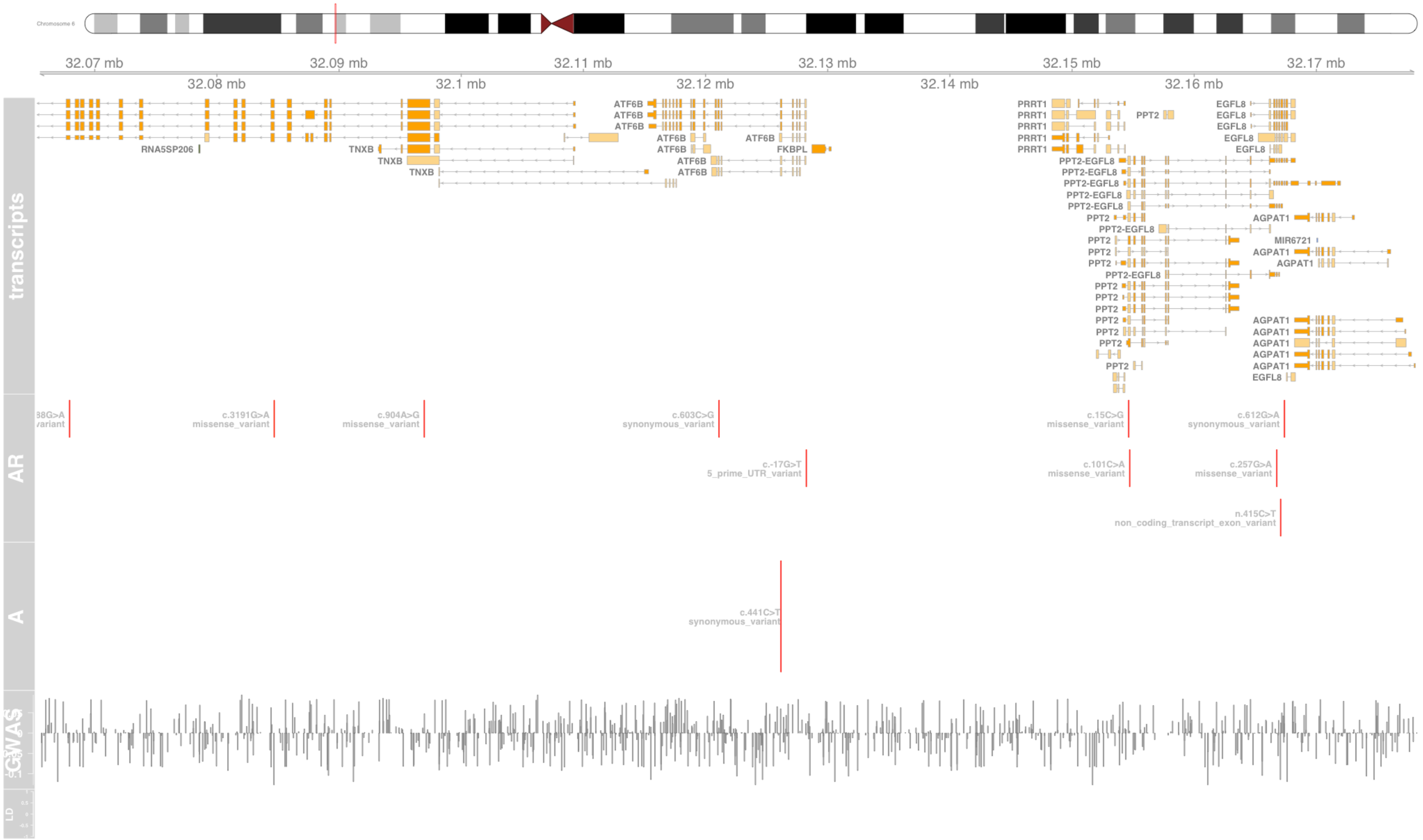

#### Fig S21 ATP6V1G2

ATPase H+ transporting V1 subunit G2 [Source:HGNC Symbol;Acc:HGNC:862]

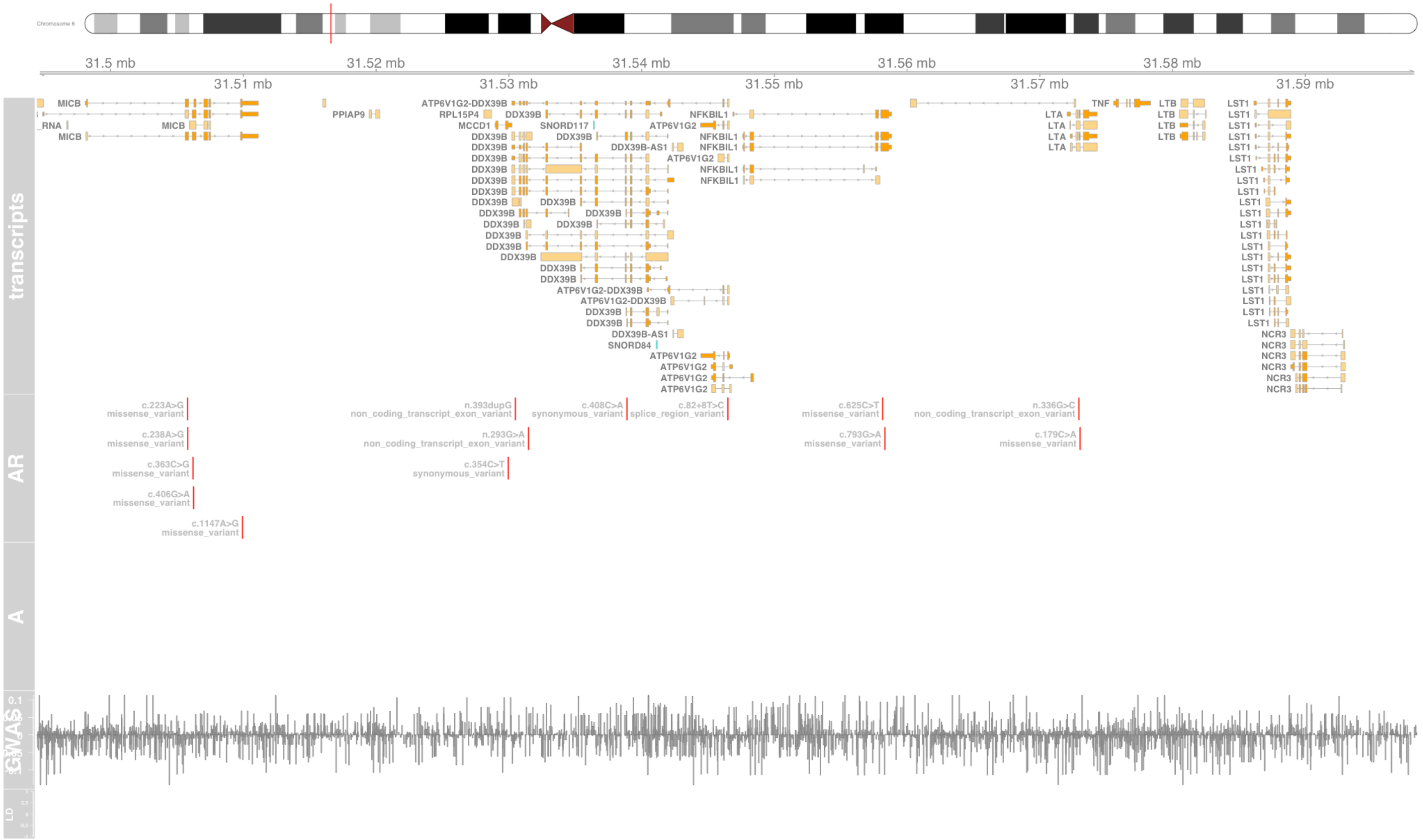

#### Fig S22 BRD2

bromodomain containing 2 [Source:HGNC Symbol;Acc:HGNC:1103]

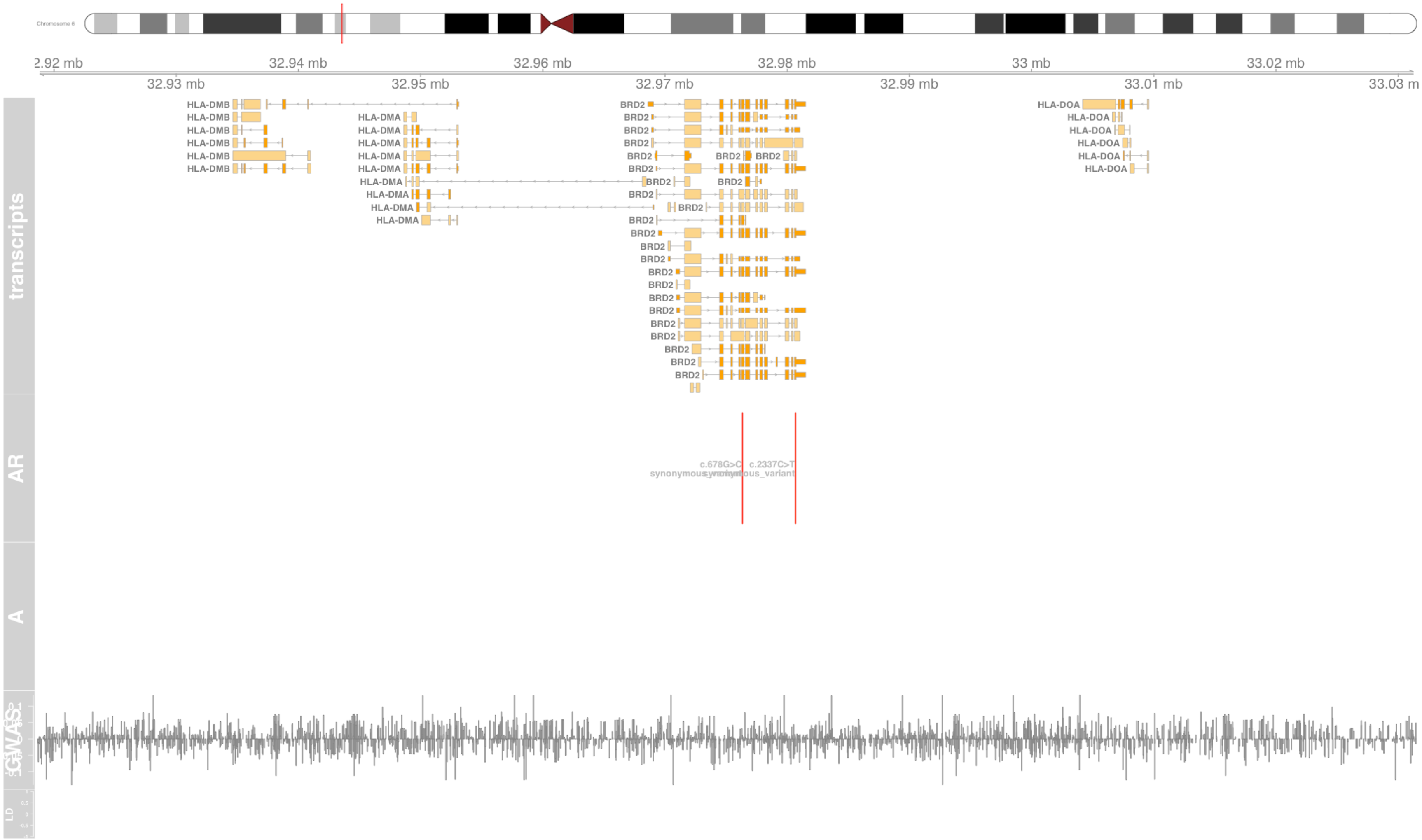

#### Fig S23 BTN2A1

butyrophilin subfamily 2 member A1 [Source:HGNC Symbol;Acc:HGNC:1136]

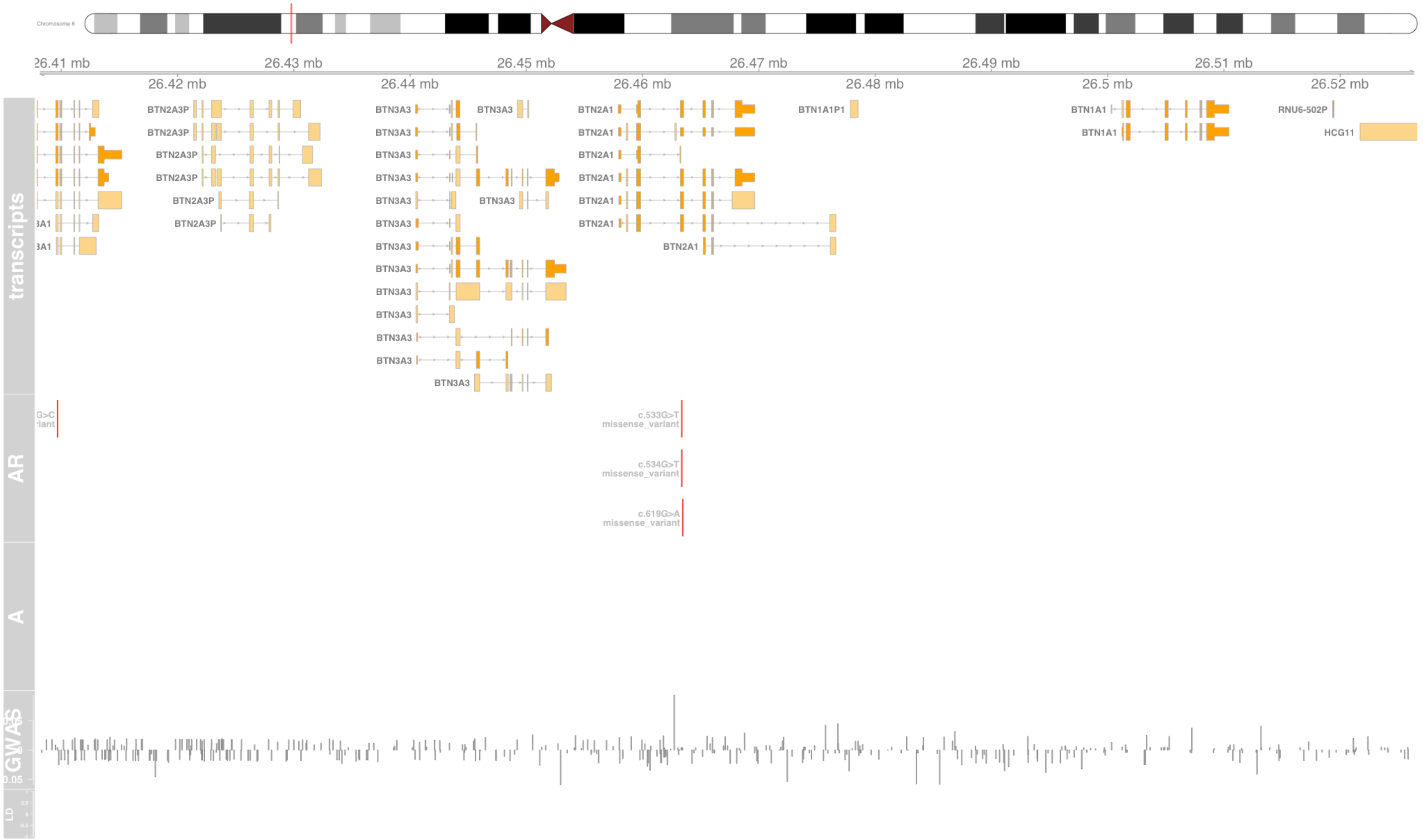

#### Fig S24 BTN3A1

butyrophilin subfamily 3 member A1 [Source:HGNC Symbol;Acc:HGNC:1138]

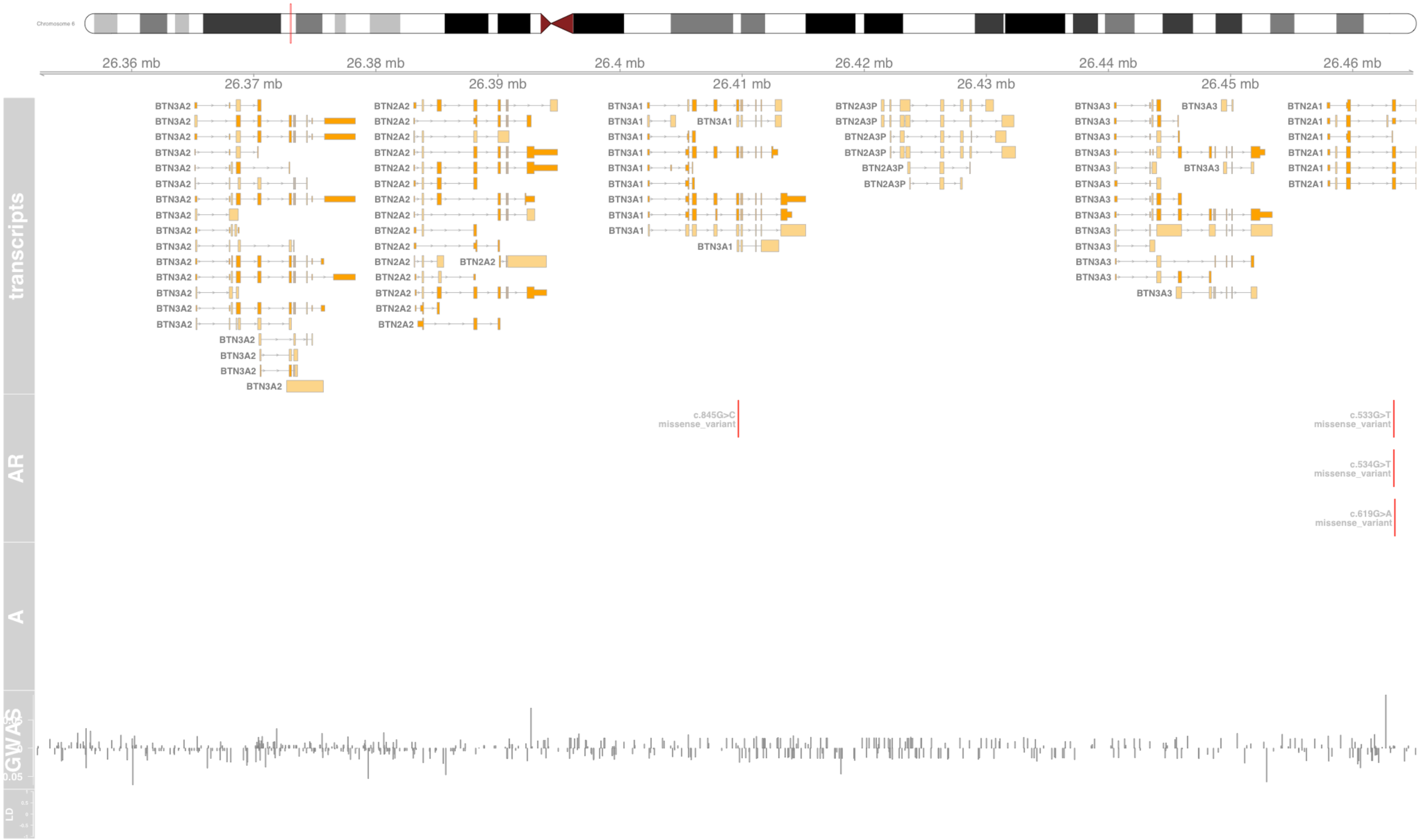

### Fig S25 BTNL2

butyrophilin like 2 [Source:HGNC Symbol;Acc:HGNC:1142]

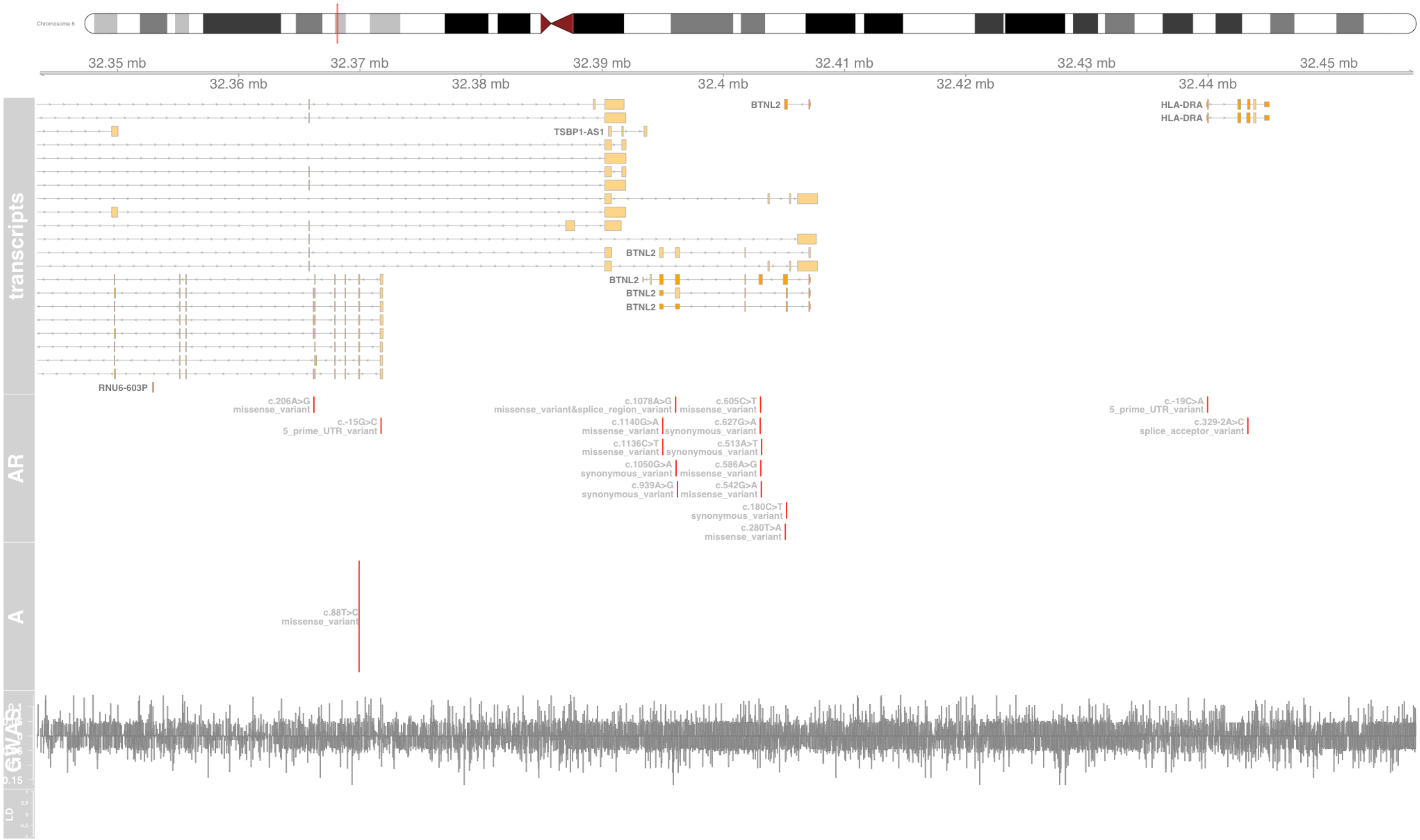

#### Fig S26 C6orf15

chromosome 6 open reading frame 15 [Source:HGNC Symbol;Acc:HGNC:13927]

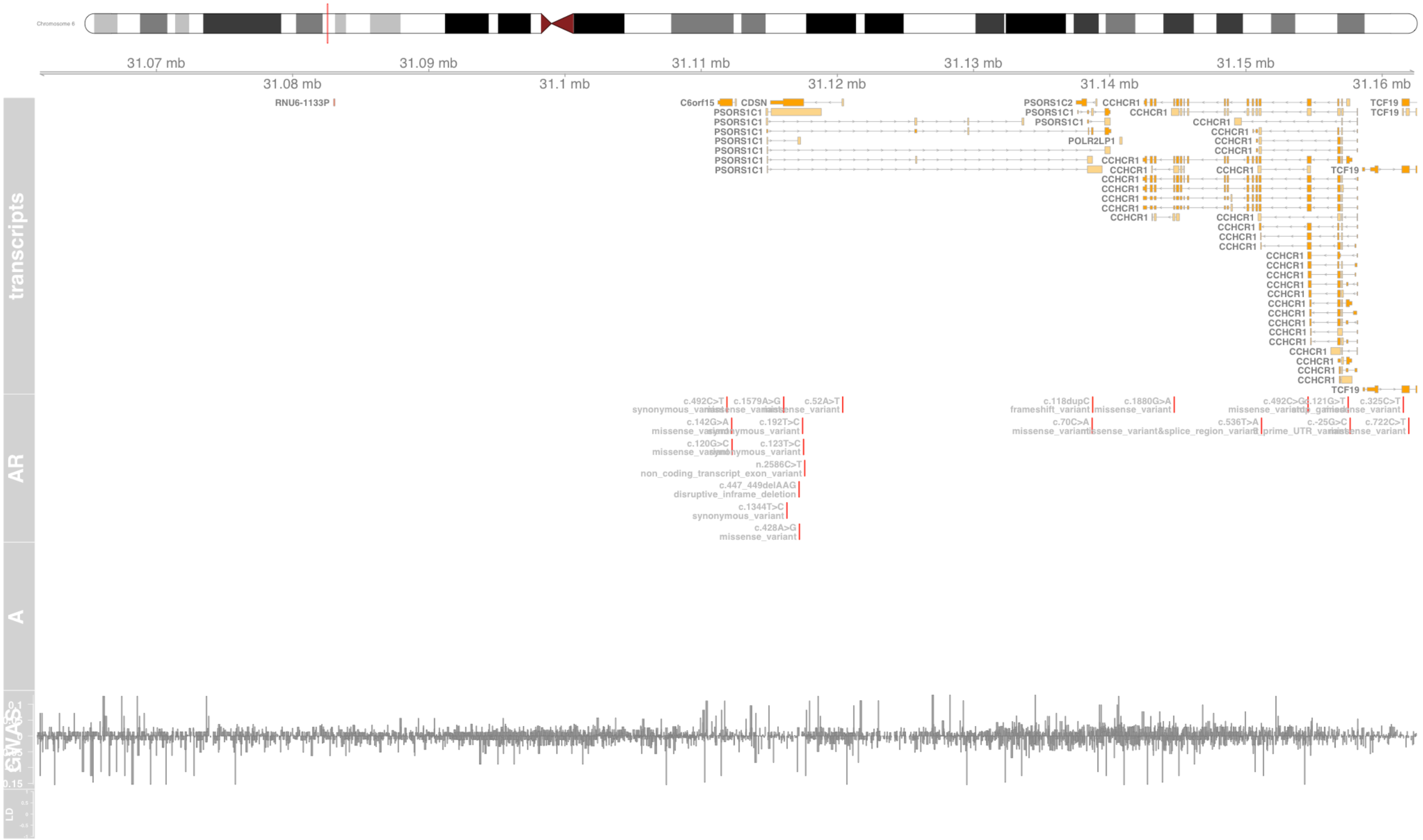

#### Fig S27 CCHCR1

coiled-coil alpha-helical rod protein 1 [Source:HGNC Symbol;Acc:HGNC:13930]

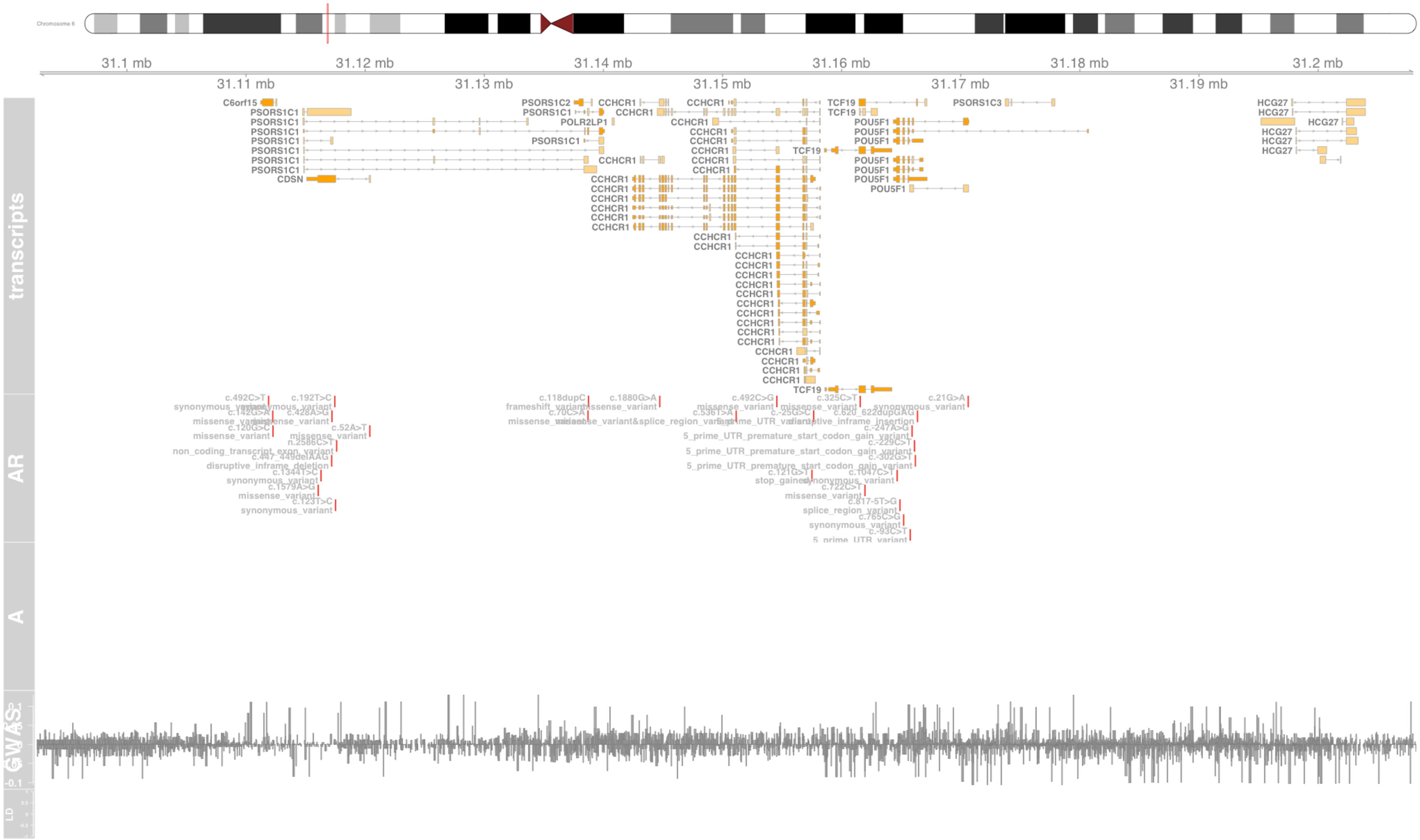

#### Fig S28 CD247

CD247 molecule [Source:HGNC Symbol;Acc:HGNC:1677]

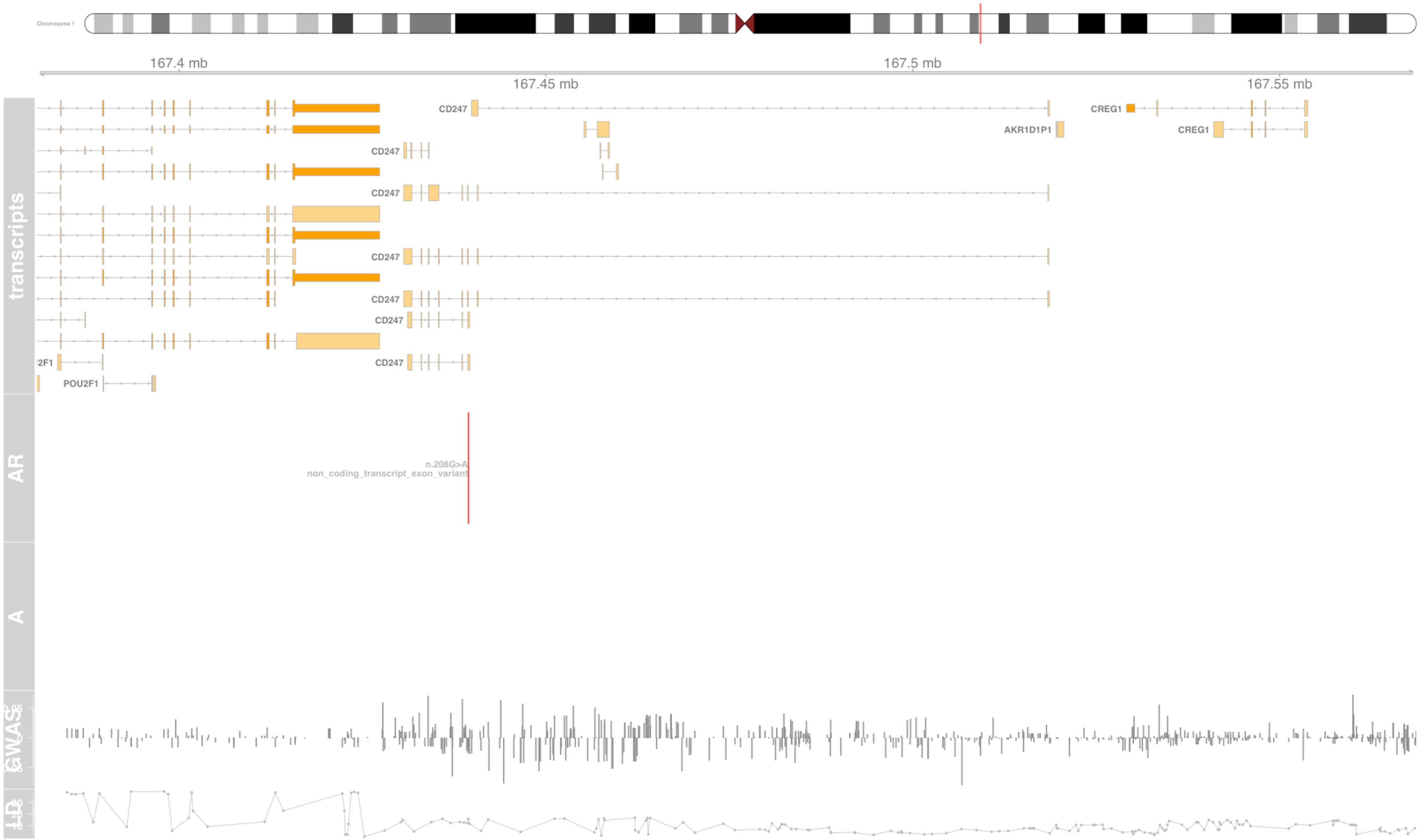

### Fig S29 CDSN

corneodesmosin [Source:HGNC Symbol;Acc:HGNC:1802]

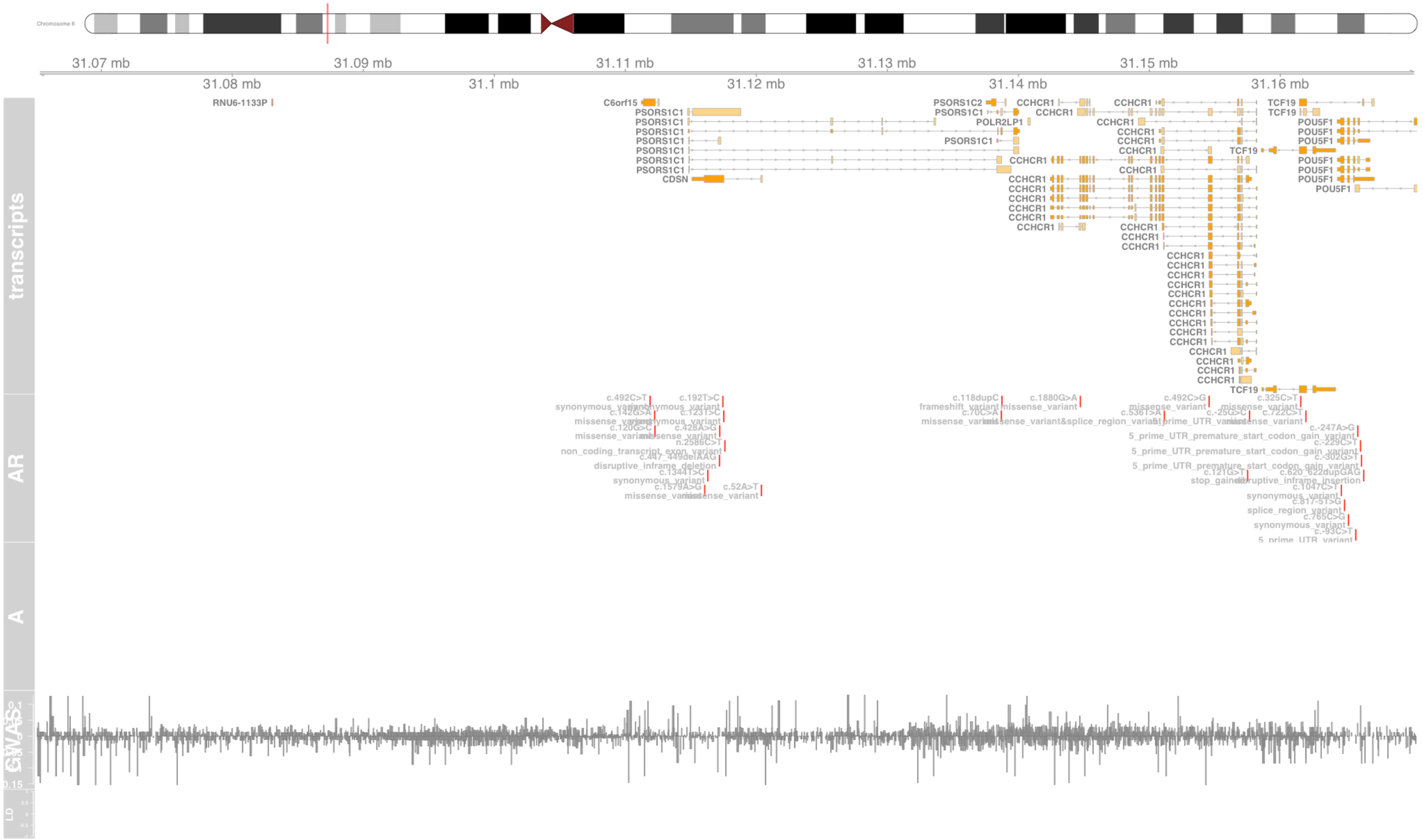

#### Fig S30 CFB

complement factor B [Source:HGNC Symbol;Acc:HGNC:1037]

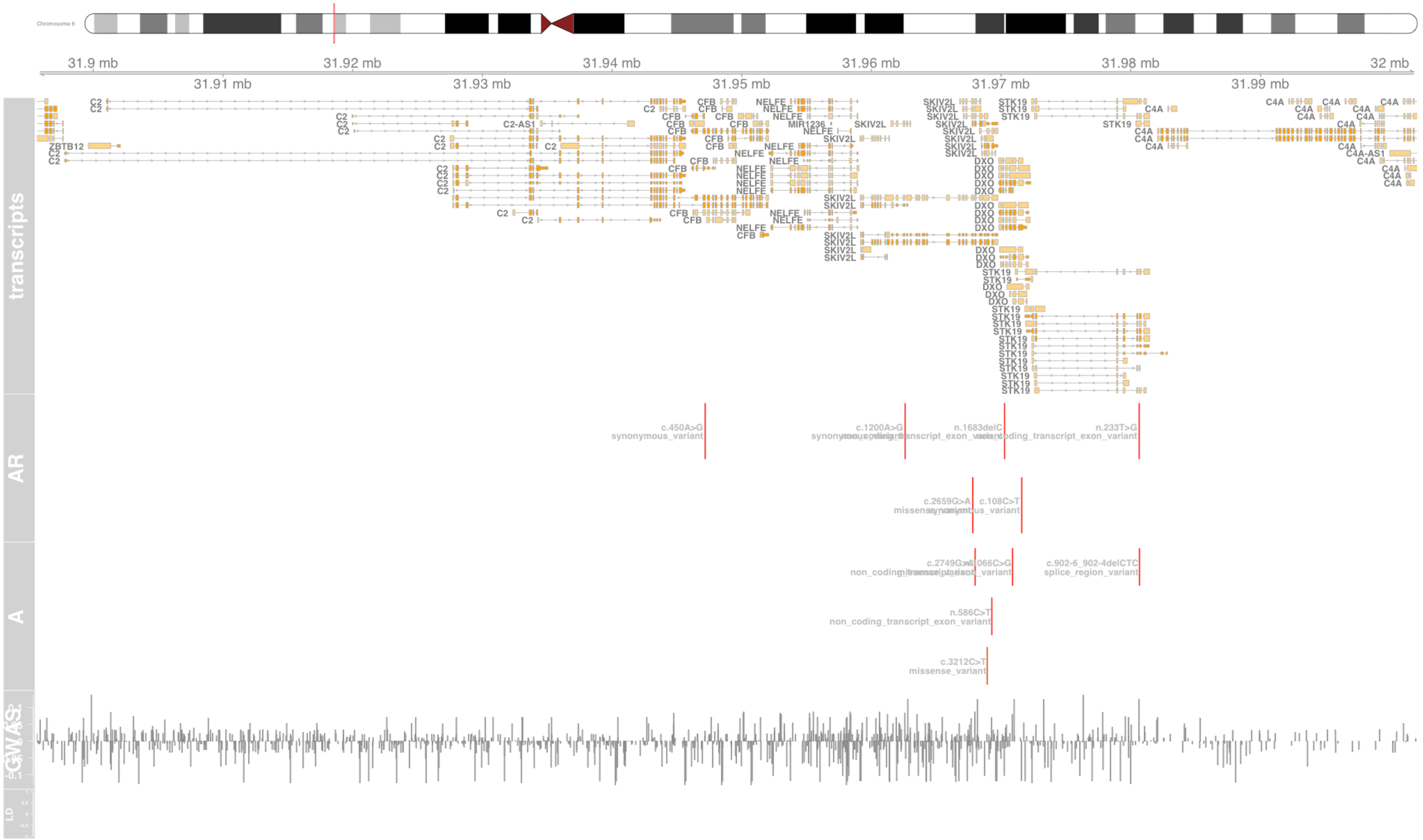

#### Fig S31 CSF3

colony stimulating factor 3 [Source:HGNC Symbol;Acc:HGNC:2438]

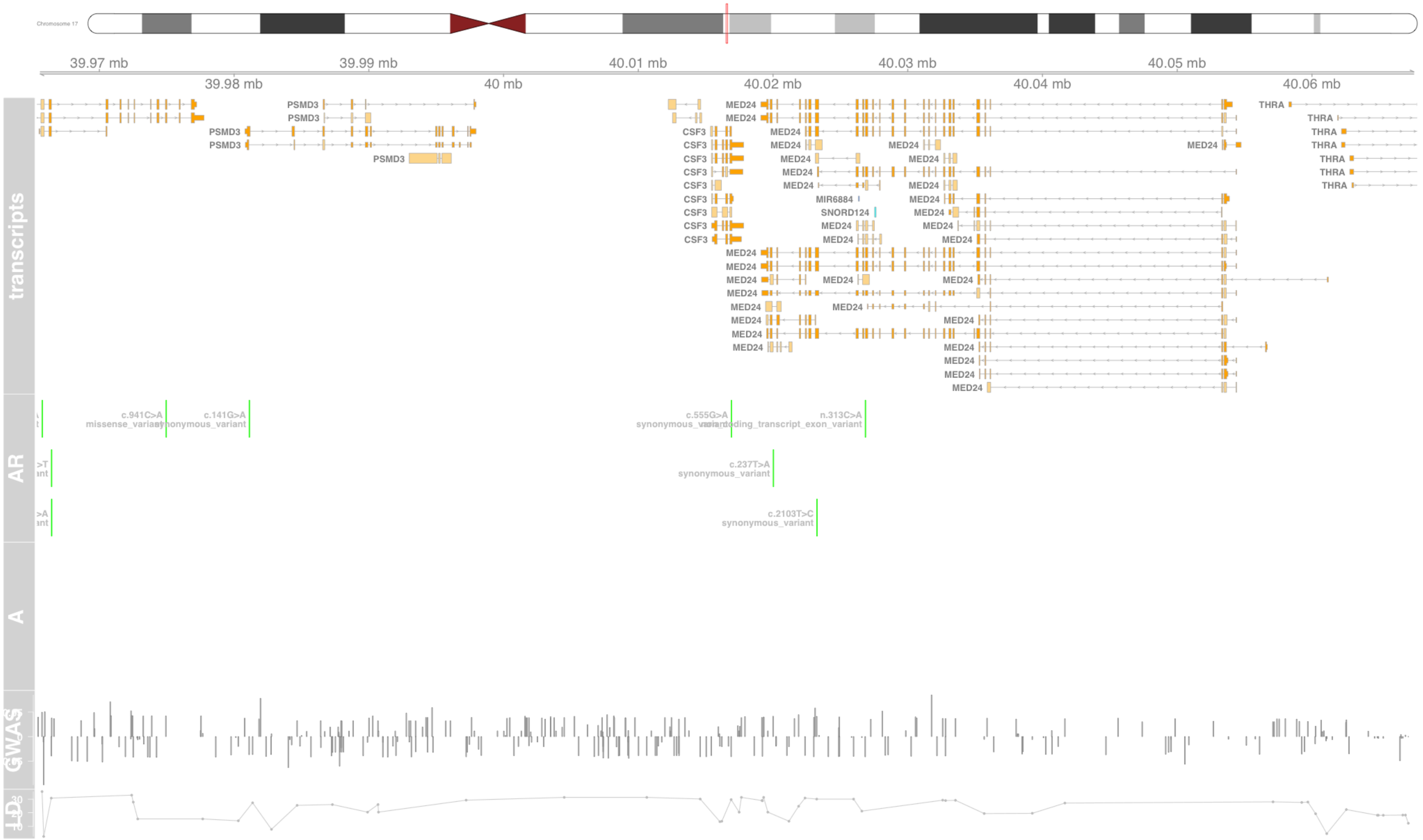

### Fig S32 CYP21A1P

cytochrome P450 family 21 subfamily A member 1, pseudogene [Source:HGNC Symbol;Acc:HGNC:2599]

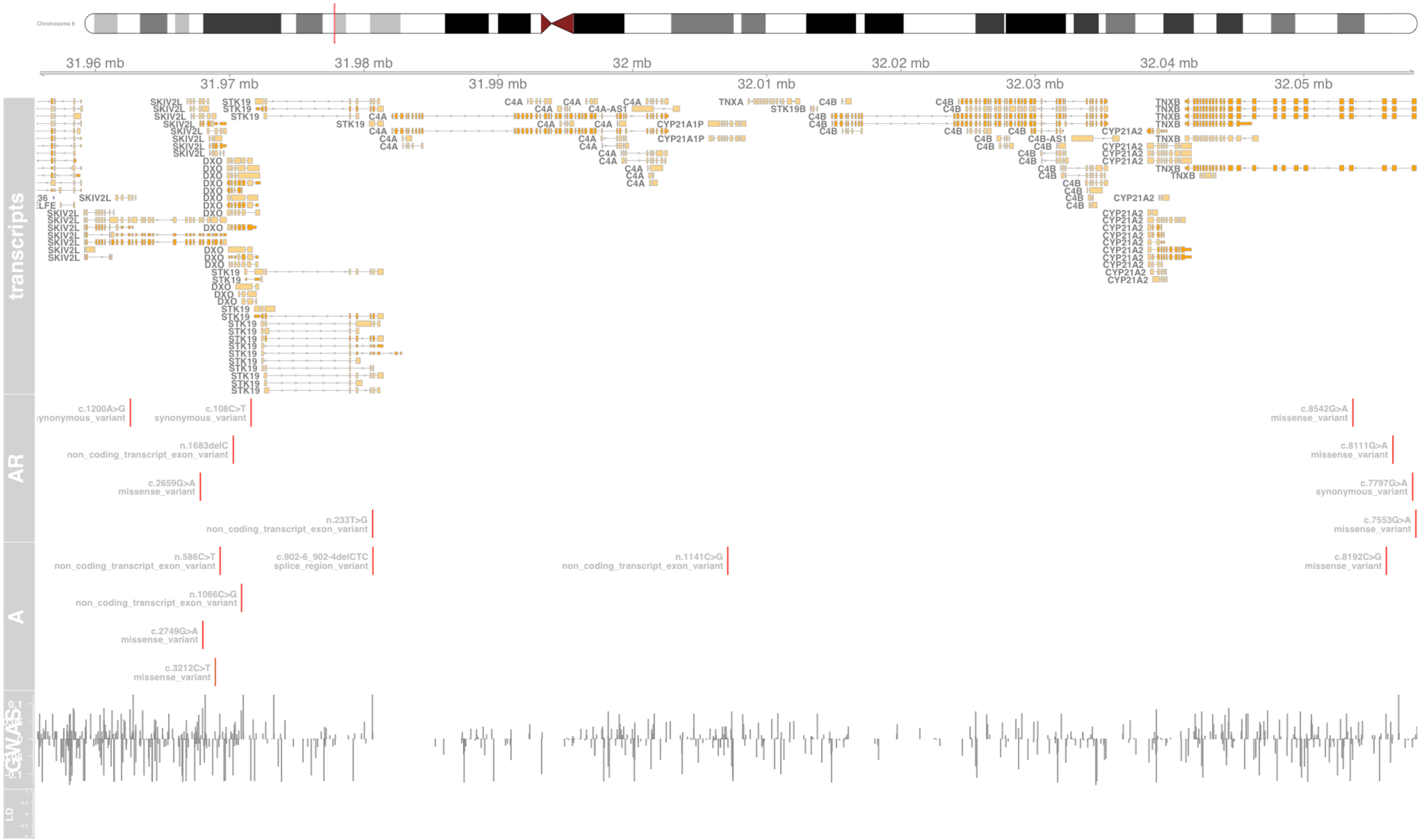

### Fig S33 D2HGDH

D-2-hydroxyglutarate dehydrogenase [Source:HGNC Symbol;Acc:HGNC:28358]

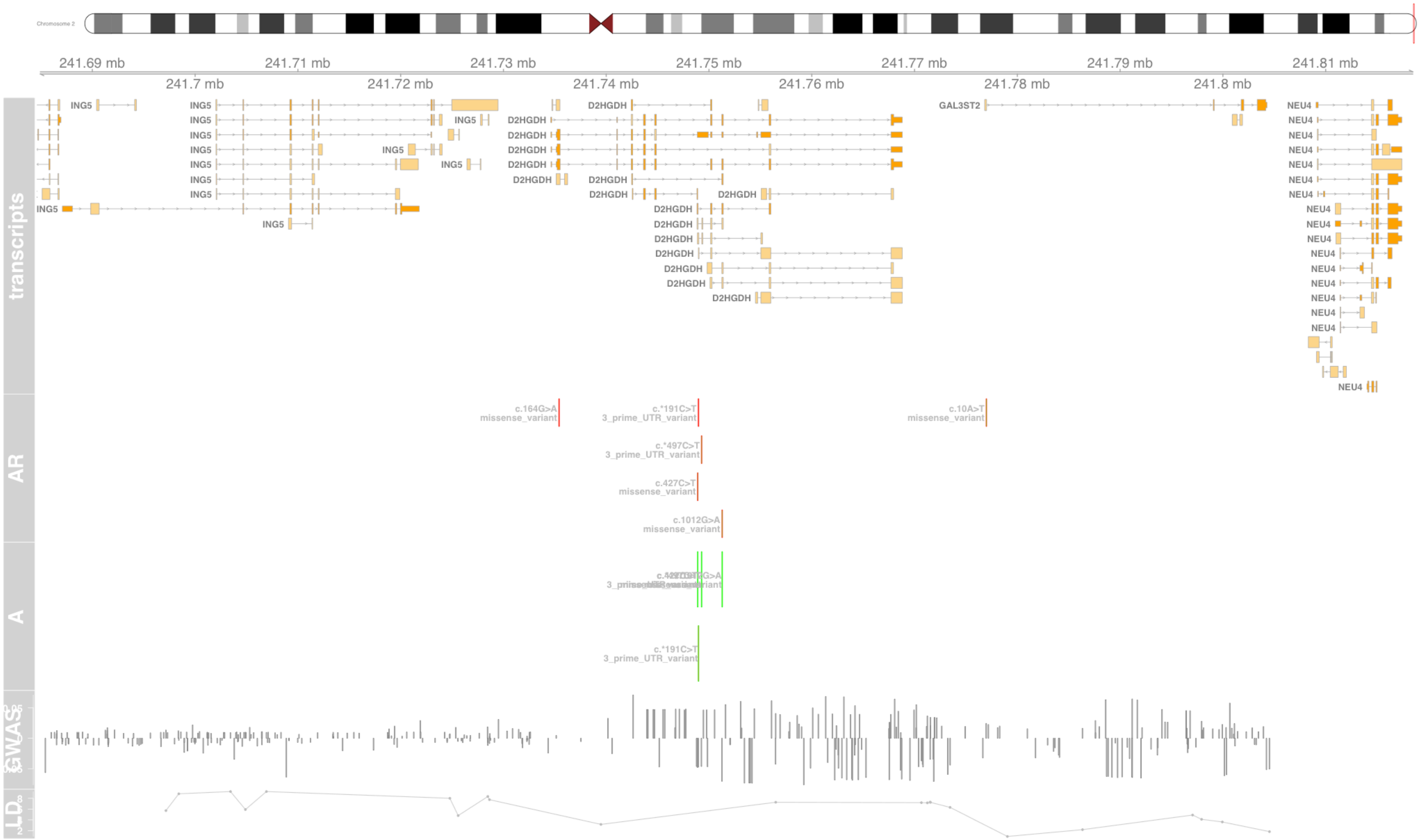

### Fig S34 DDX39B

DExD-box helicase 39B [Source:HGNC Symbol;Acc:HGNC:13917]

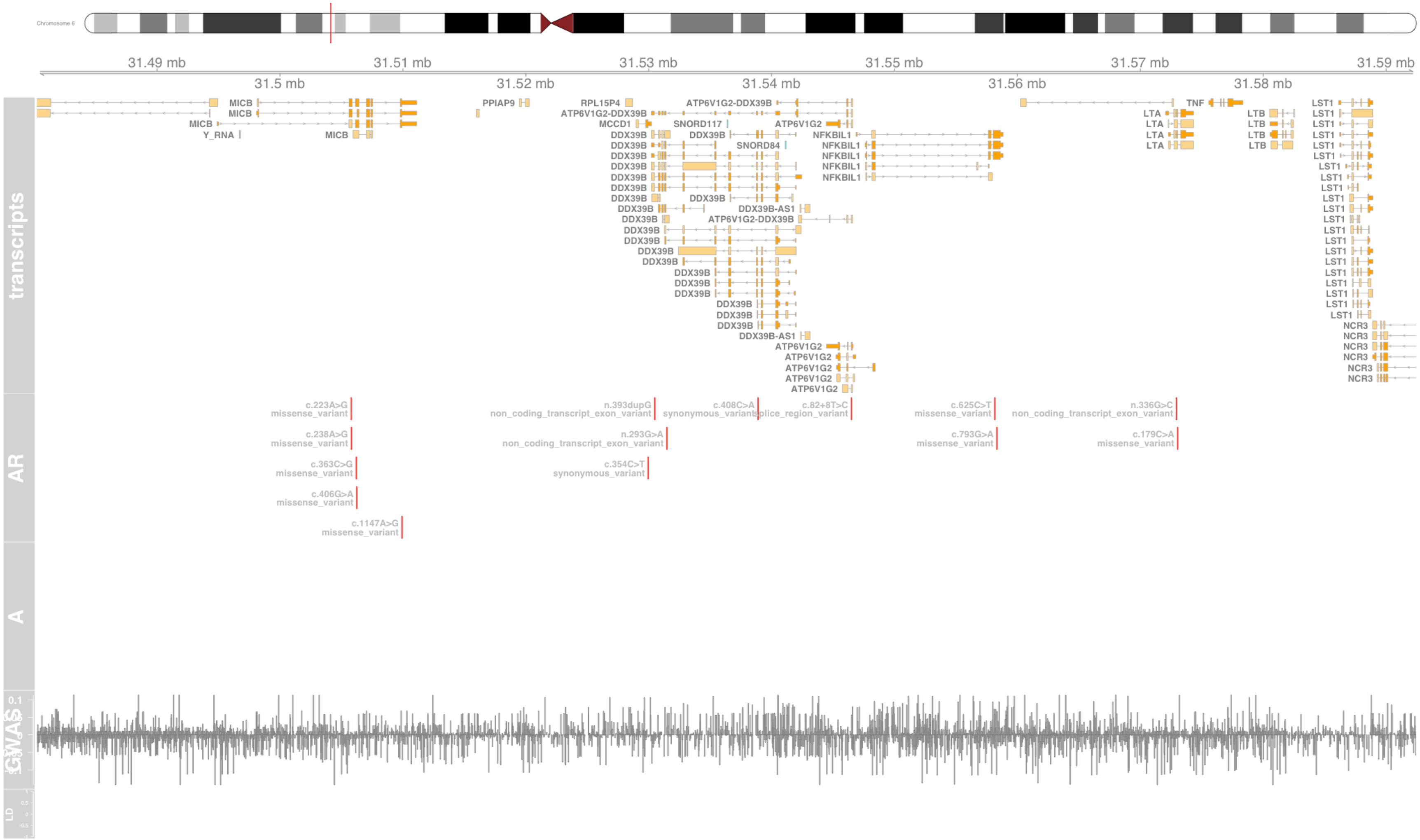

#### Fig S35 DXO

decapping exoribonuclease [Source:HGNC Symbol;Acc:HGNC:2992]

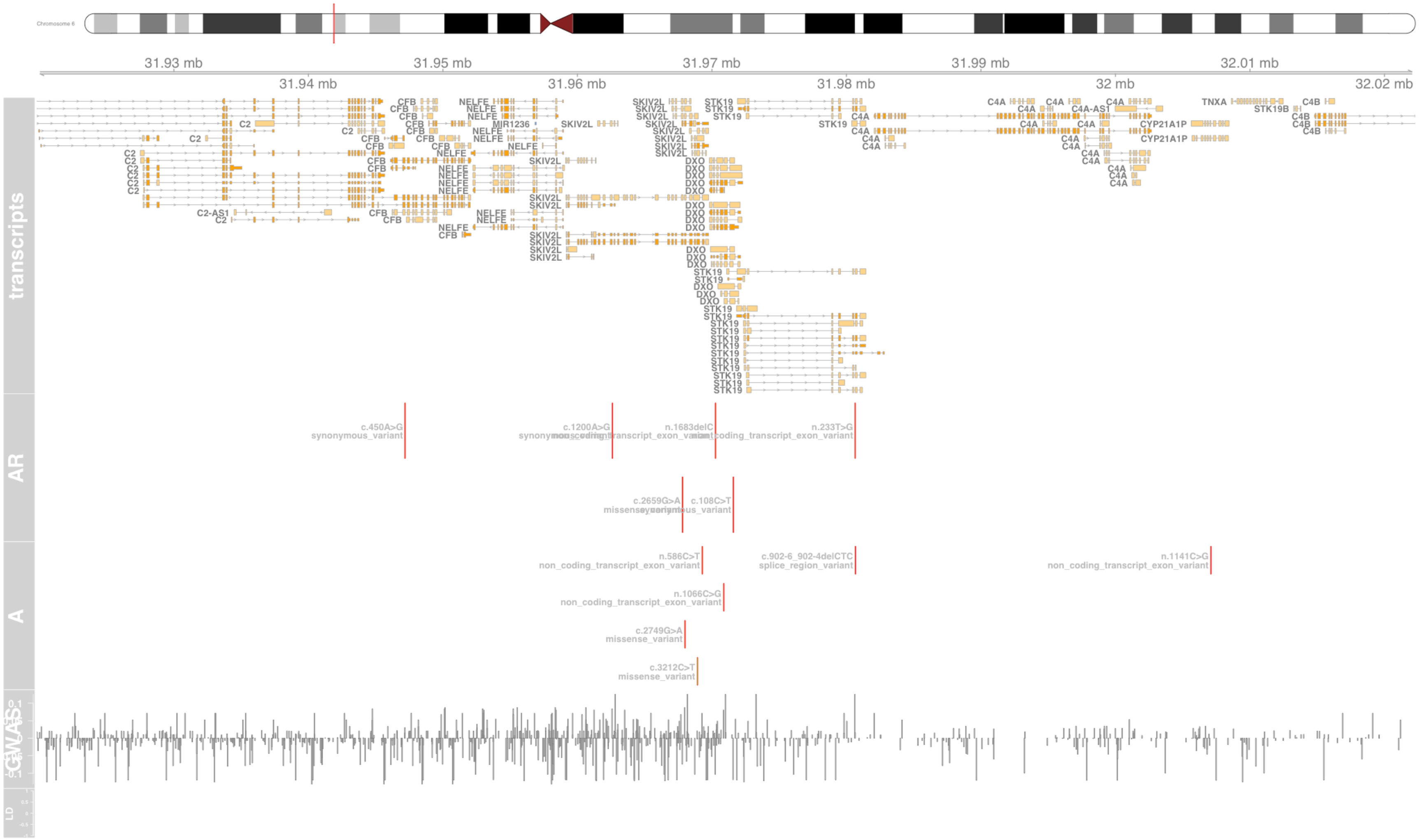

#### Fig S36 EGFL8

EGF like domain multiple 8 [Source:HGNC Symbol;Acc:HGNC:13944]

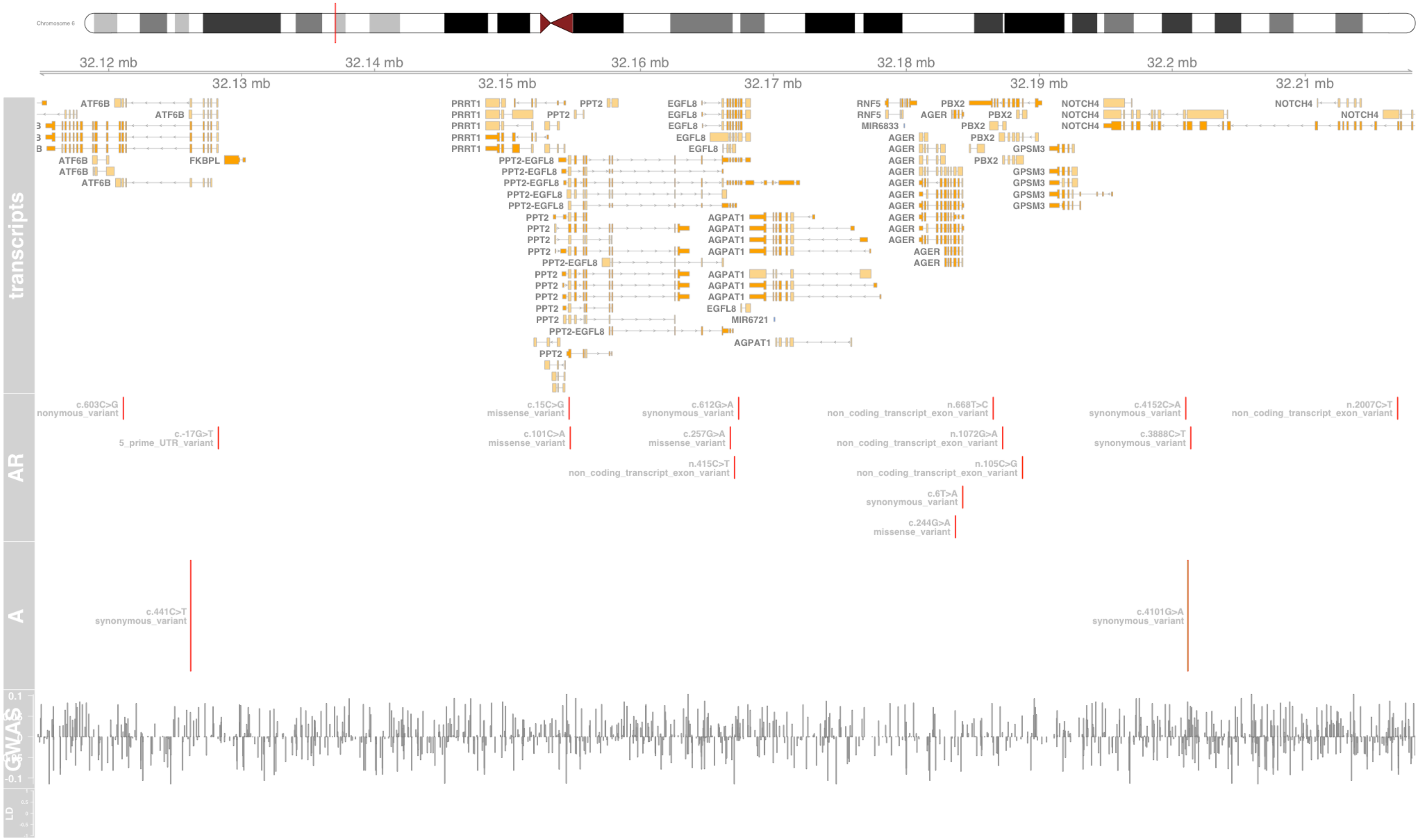

### Fig S37 ERBB2

erb-b2 receptor tyrosine kinase 2 [Source:HGNC Symbol;Acc:HGNC:3430]

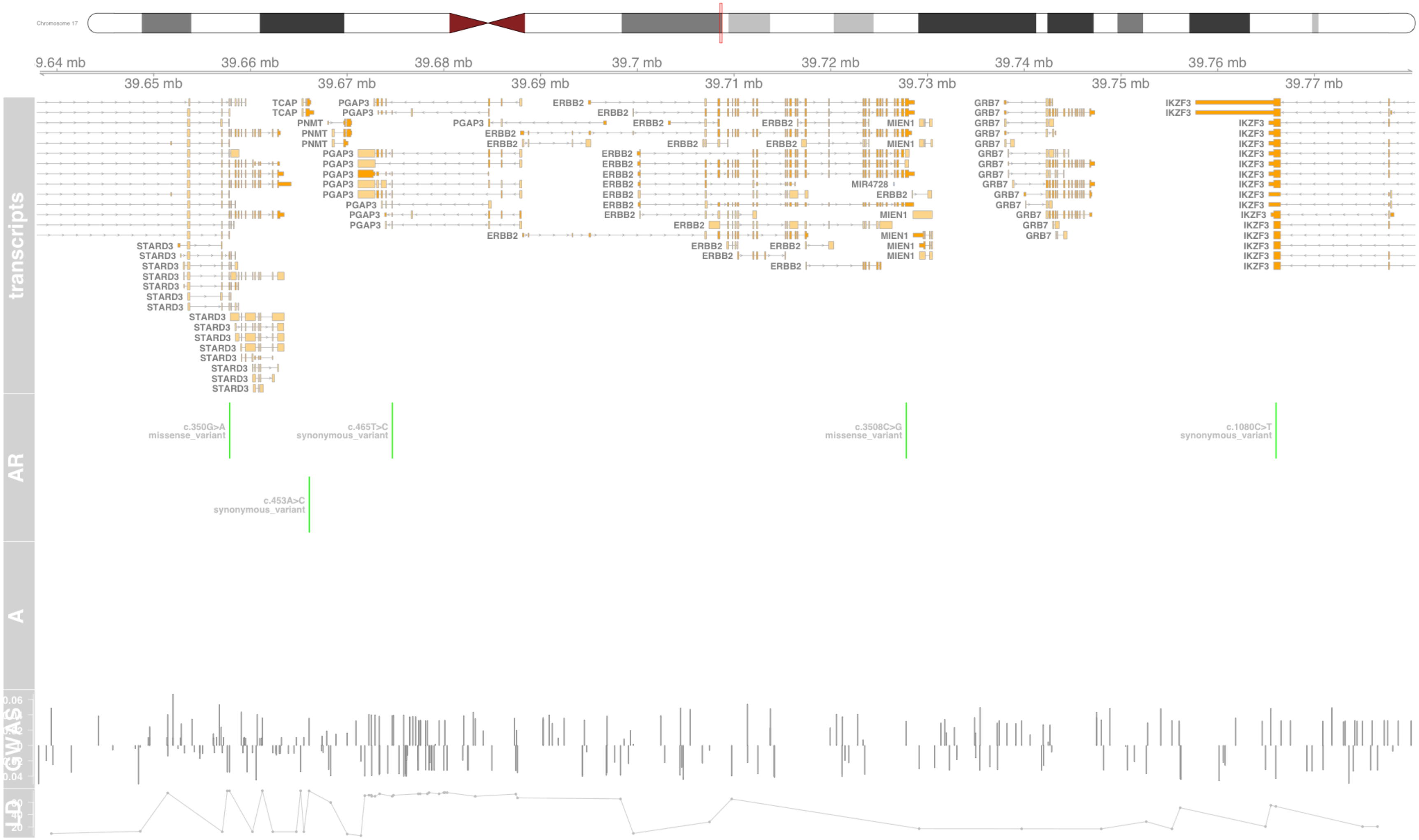

#### Fig S38 ERBB3

erb-b2 receptor tyrosine kinase 3 [Source:HGNC Symbol;Acc:HGNC:3431]

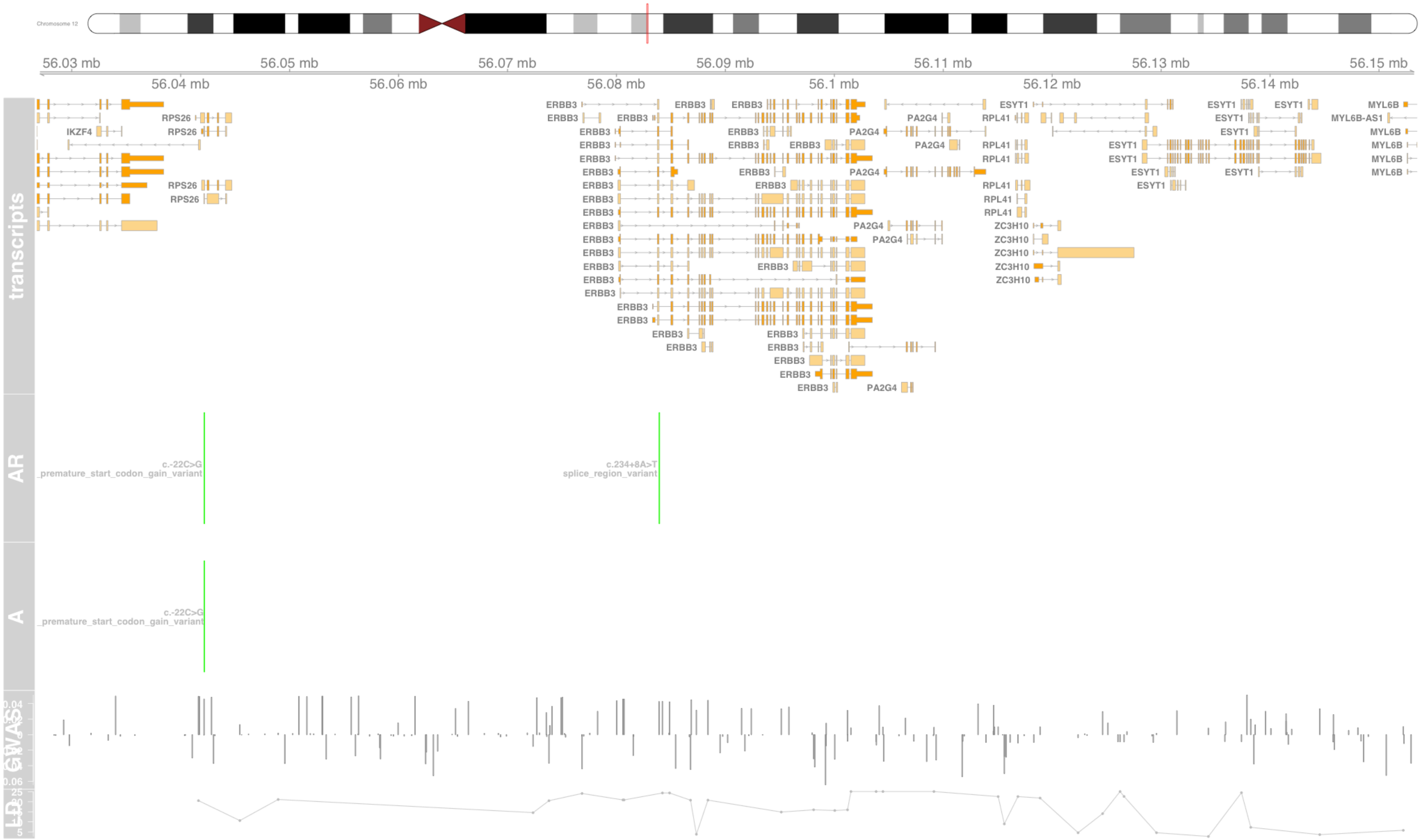

#### Fig S39 FLG

filaggrin [Source:HGNC Symbol;Acc:HGNC:3748]

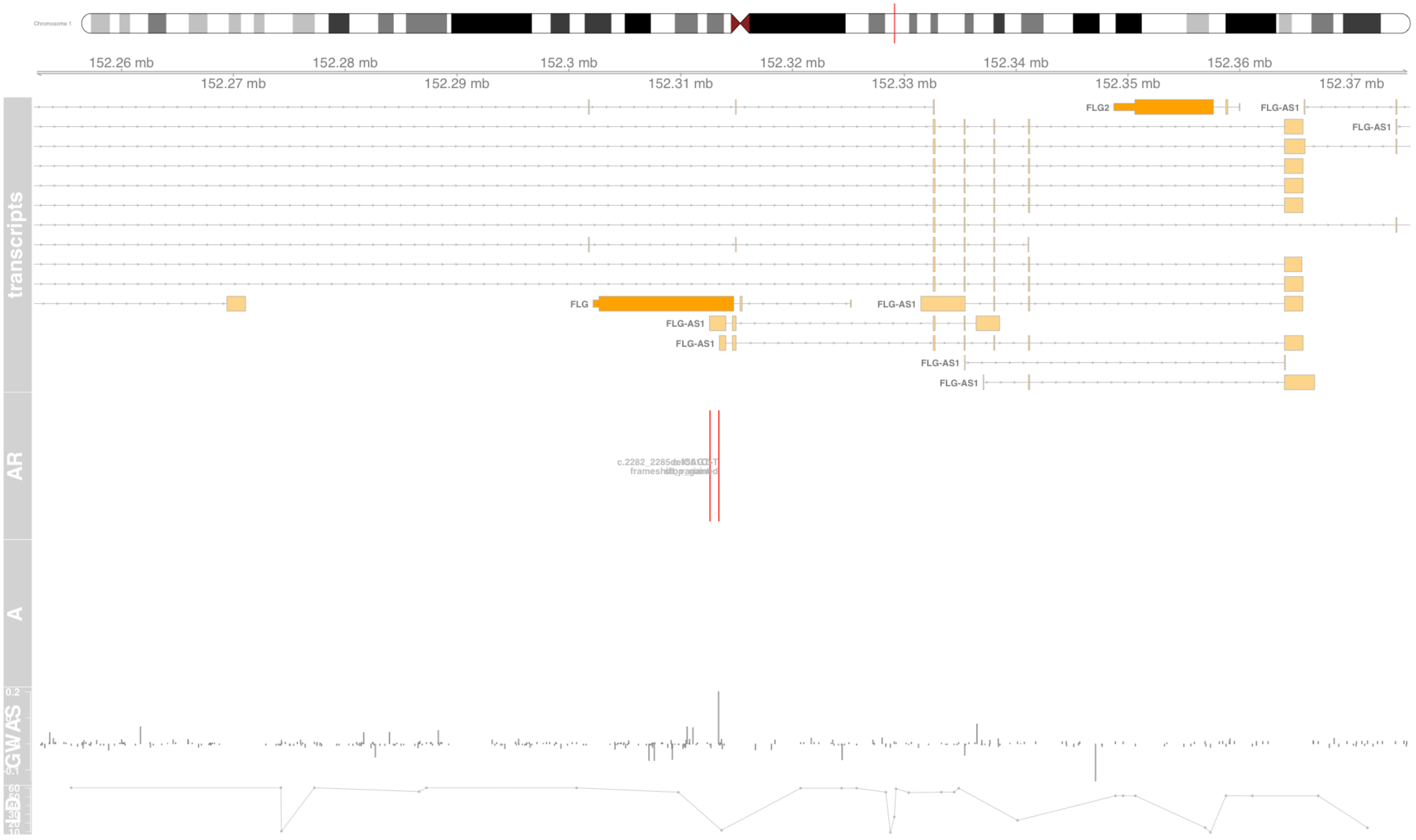

#### Fig S40 FLOT1

flotillin 1 [Source:HGNC Symbol;Acc:HGNC:3757]

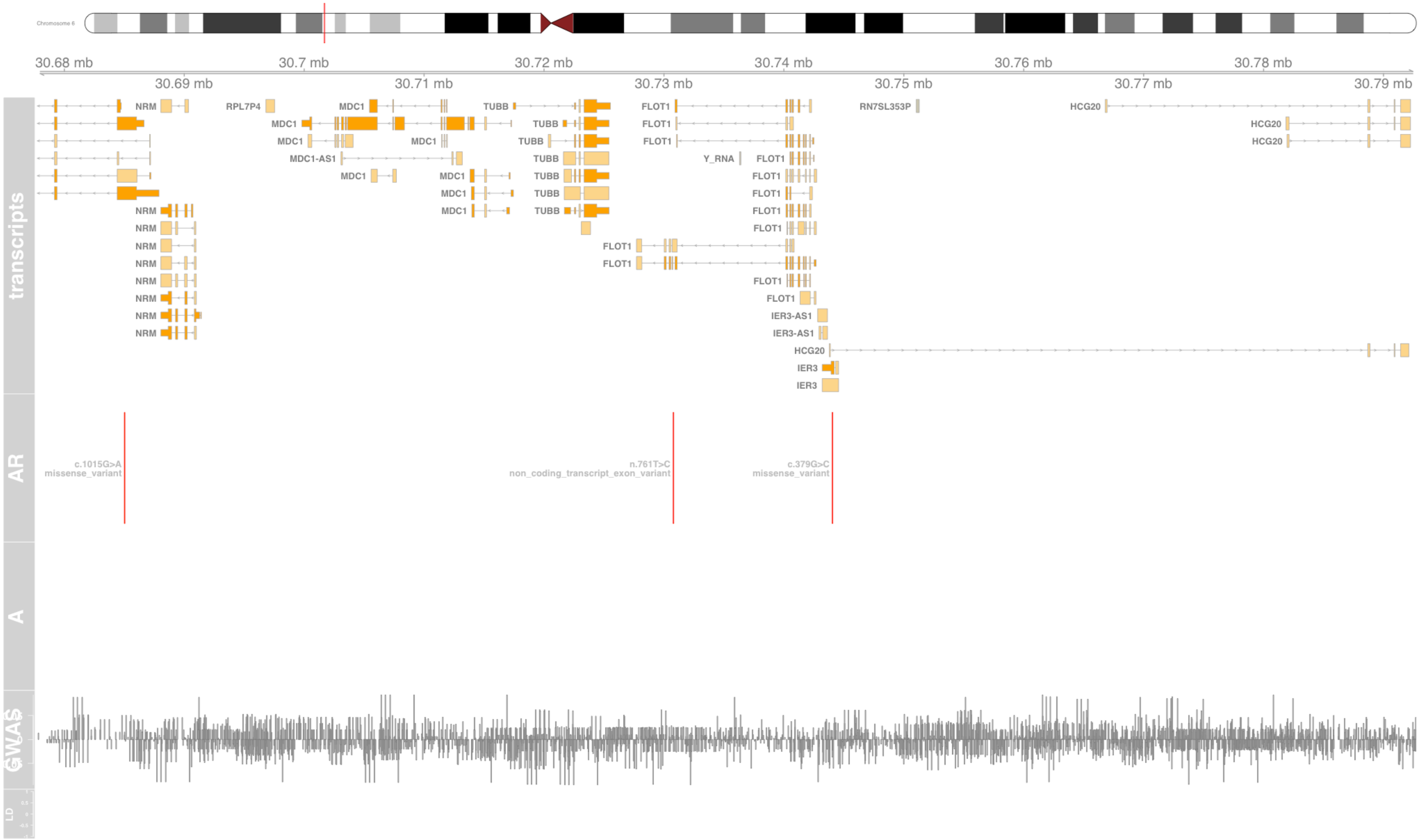

#### Fig S41 GAL3ST2

galactose-3-O-sulfotransferase 2 [Source:HGNC Symbol;Acc:HGNC:24869]

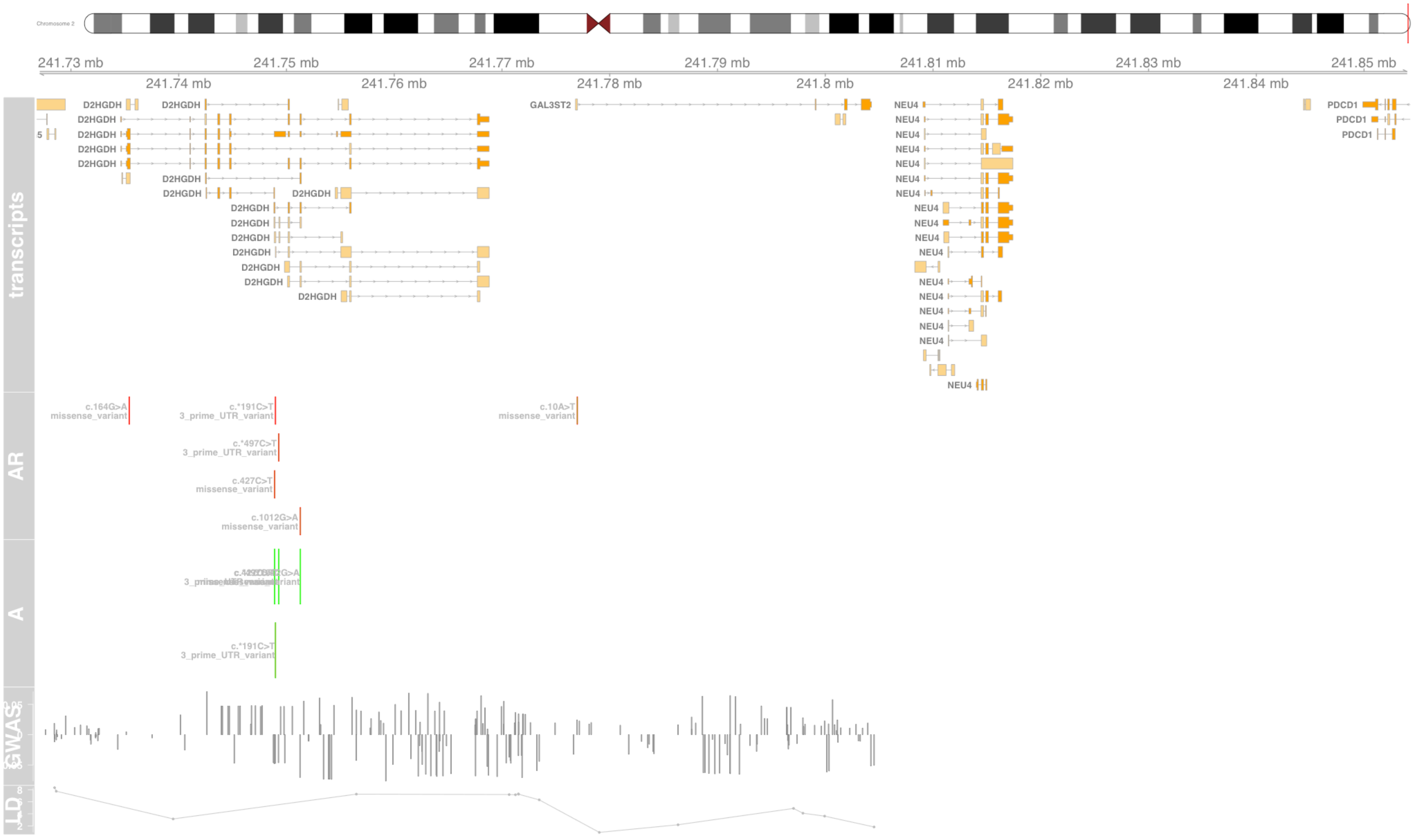

#### Fig S42 GNL1

G protein nucleolar 1 (putative) [Source:HGNC Symbol;Acc:HGNC:4413]

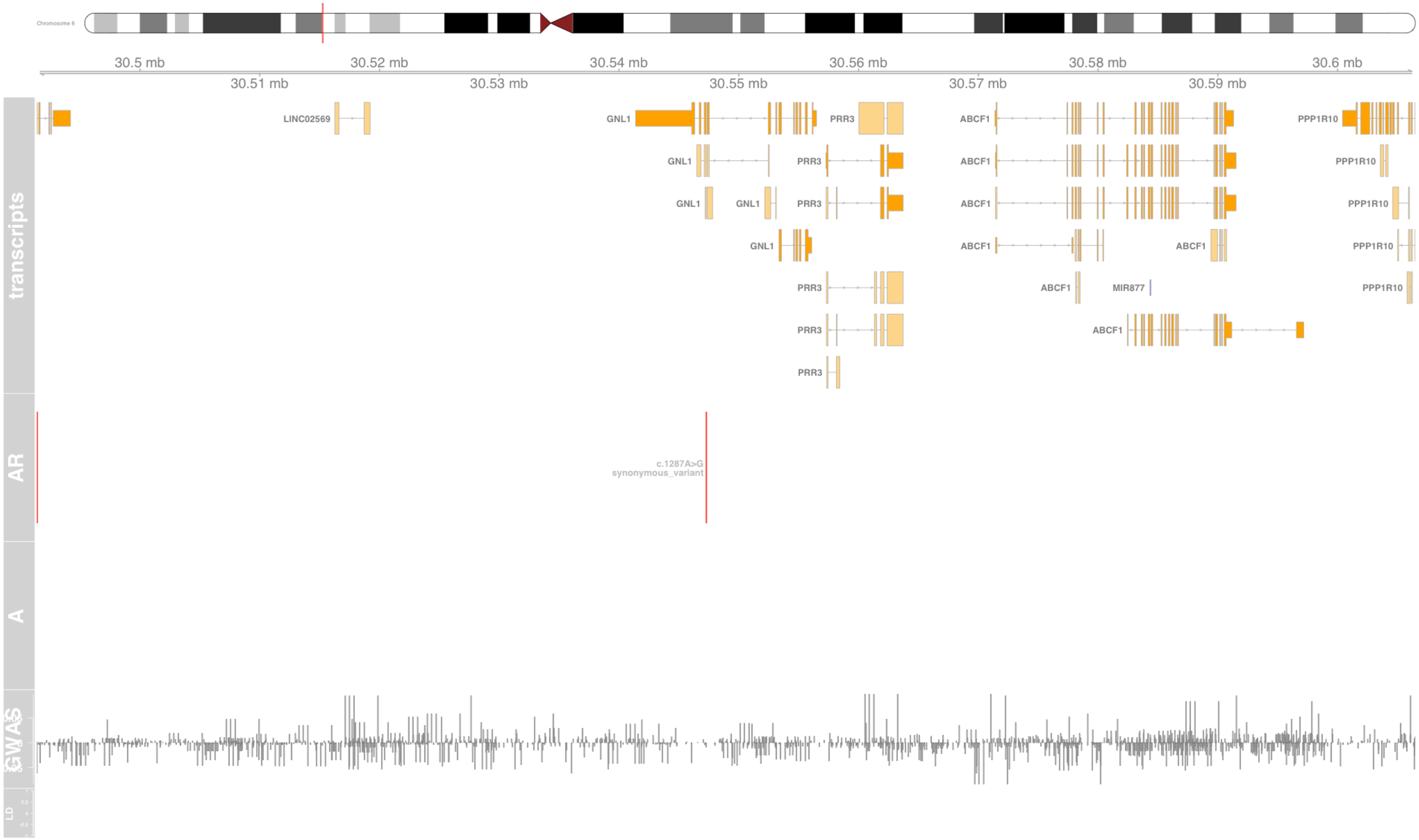

### Fig S43 GSDMA

gasdermin A [Source:HGNC Symbol;Acc:HGNC:13311]

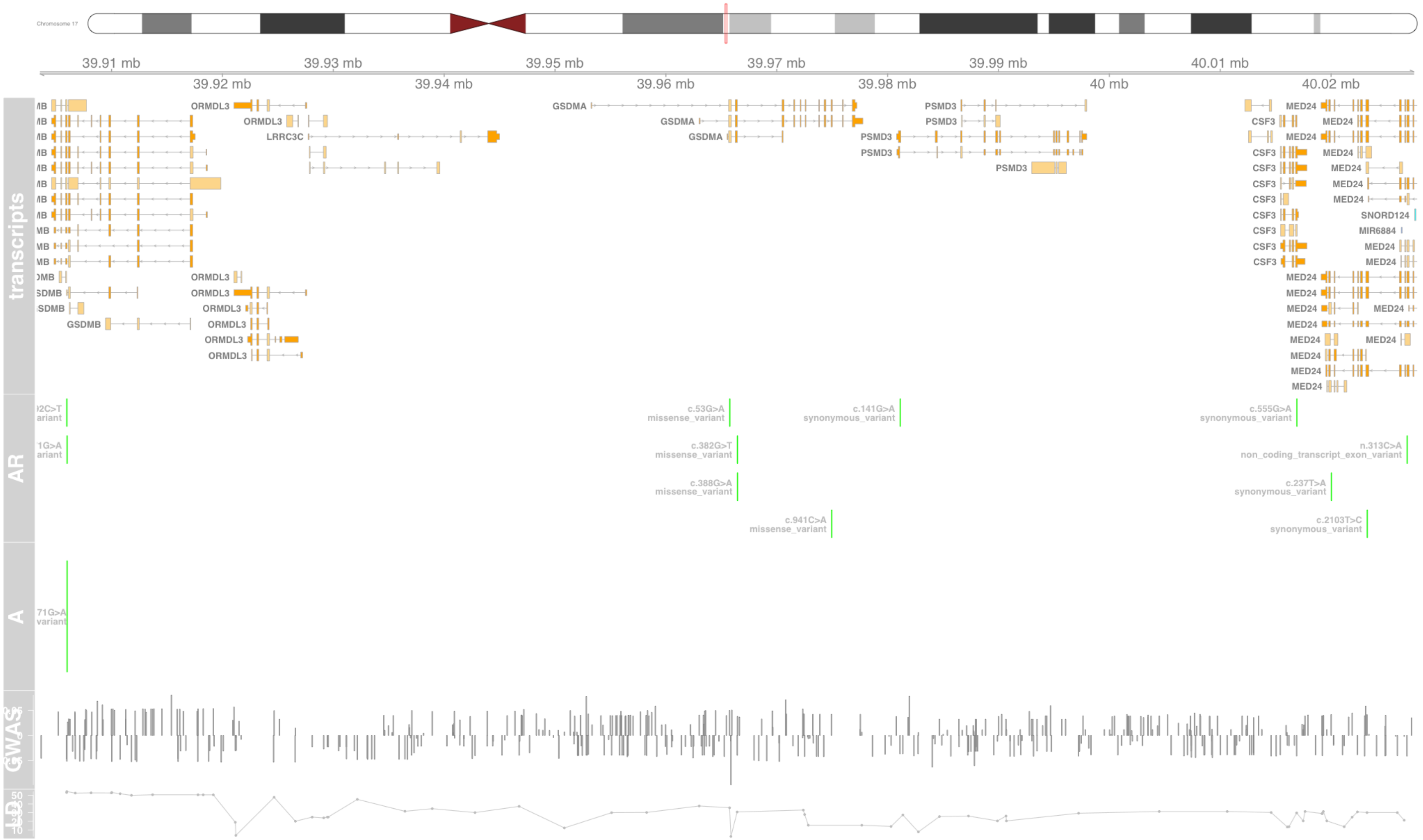

#### Fig S44 GSDMB

gasdermin B [Source:HGNC Symbol;Acc:HGNC:23690]

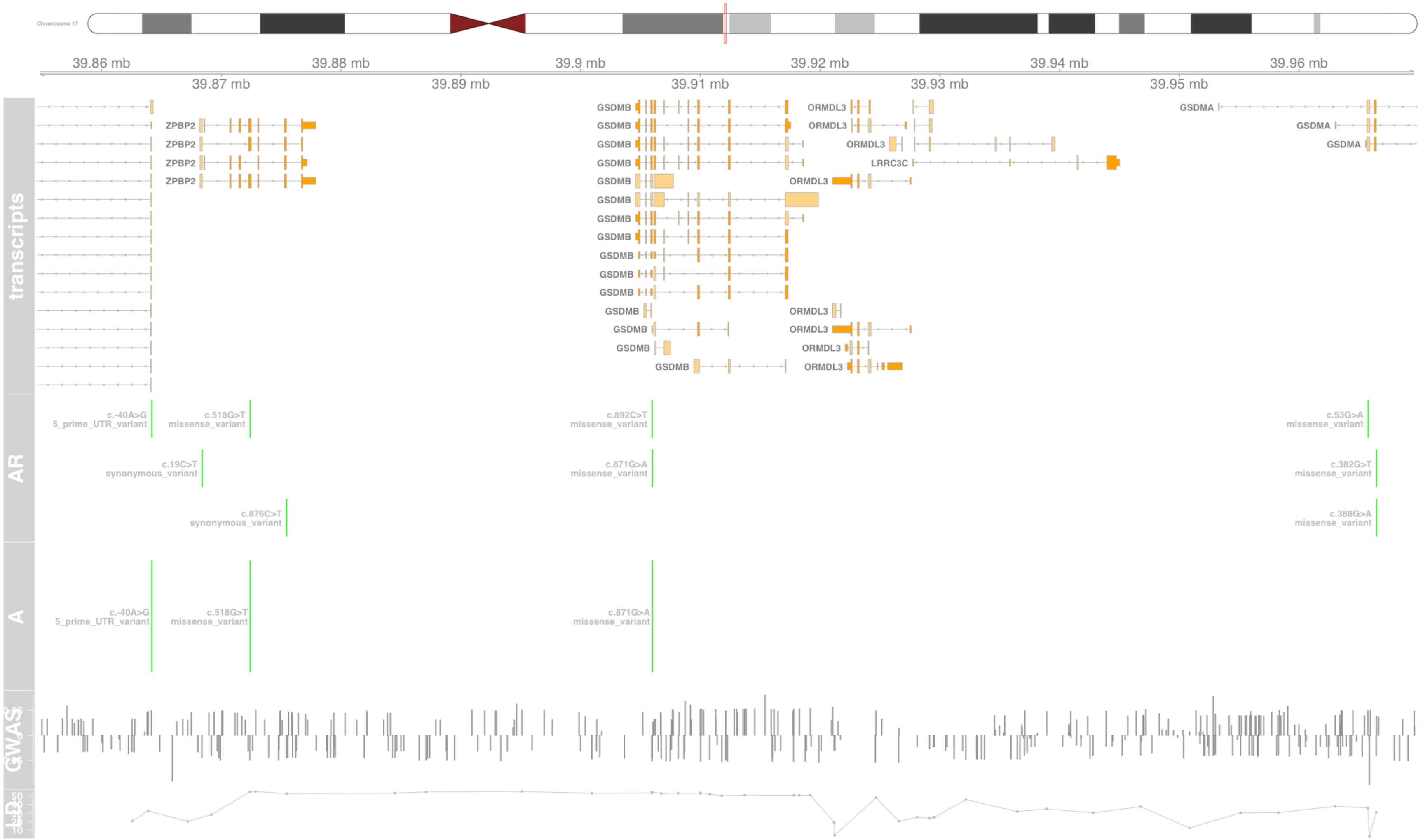

### Fig S45 HCG4

HLA complex group 4 [Source:HGNC Symbol;Acc:HGNC:21241]

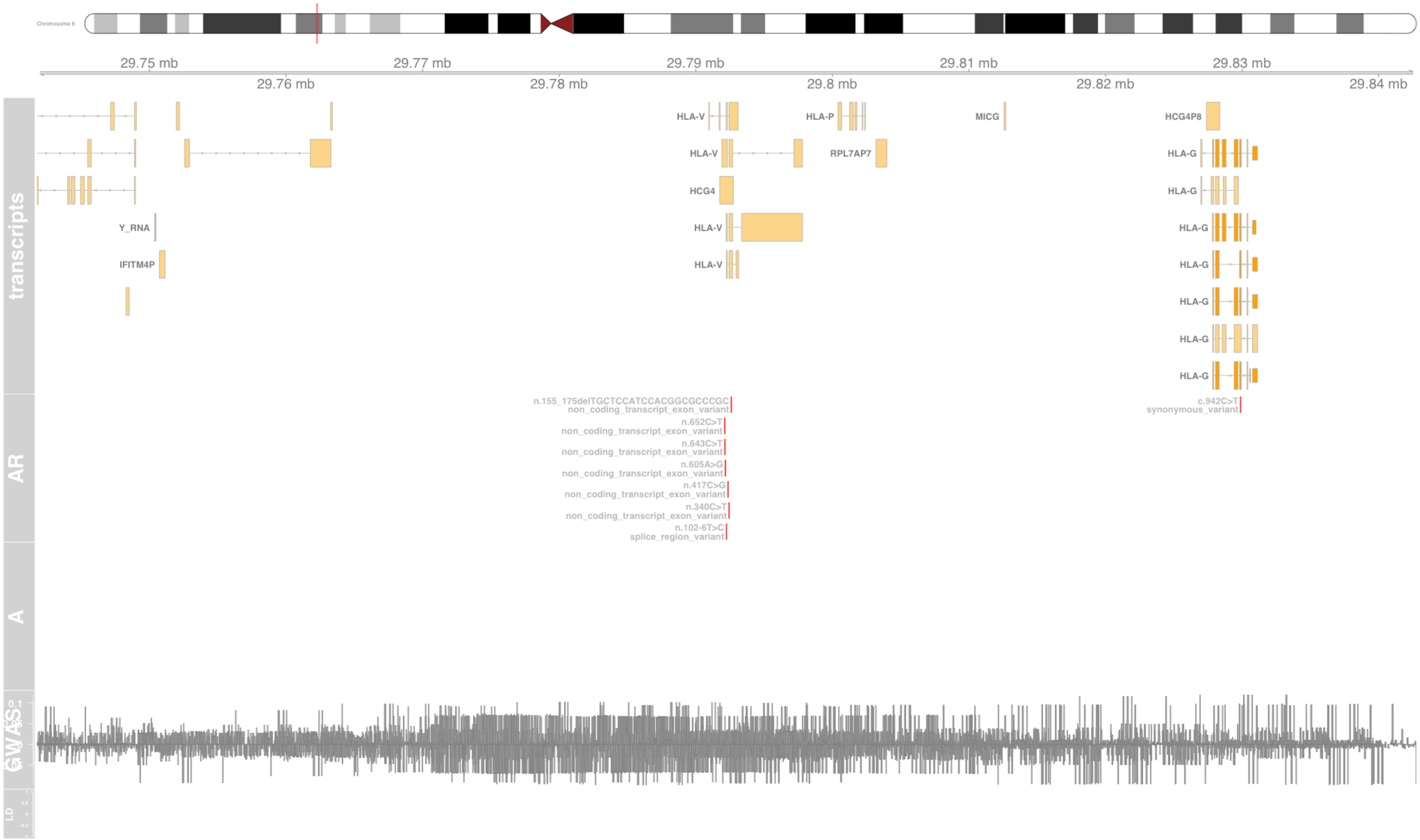

#### Fig S46 HLA-A

major histocompatibility complex, class I, A [Source:HGNC Symbol;Acc:HGNC:4931]

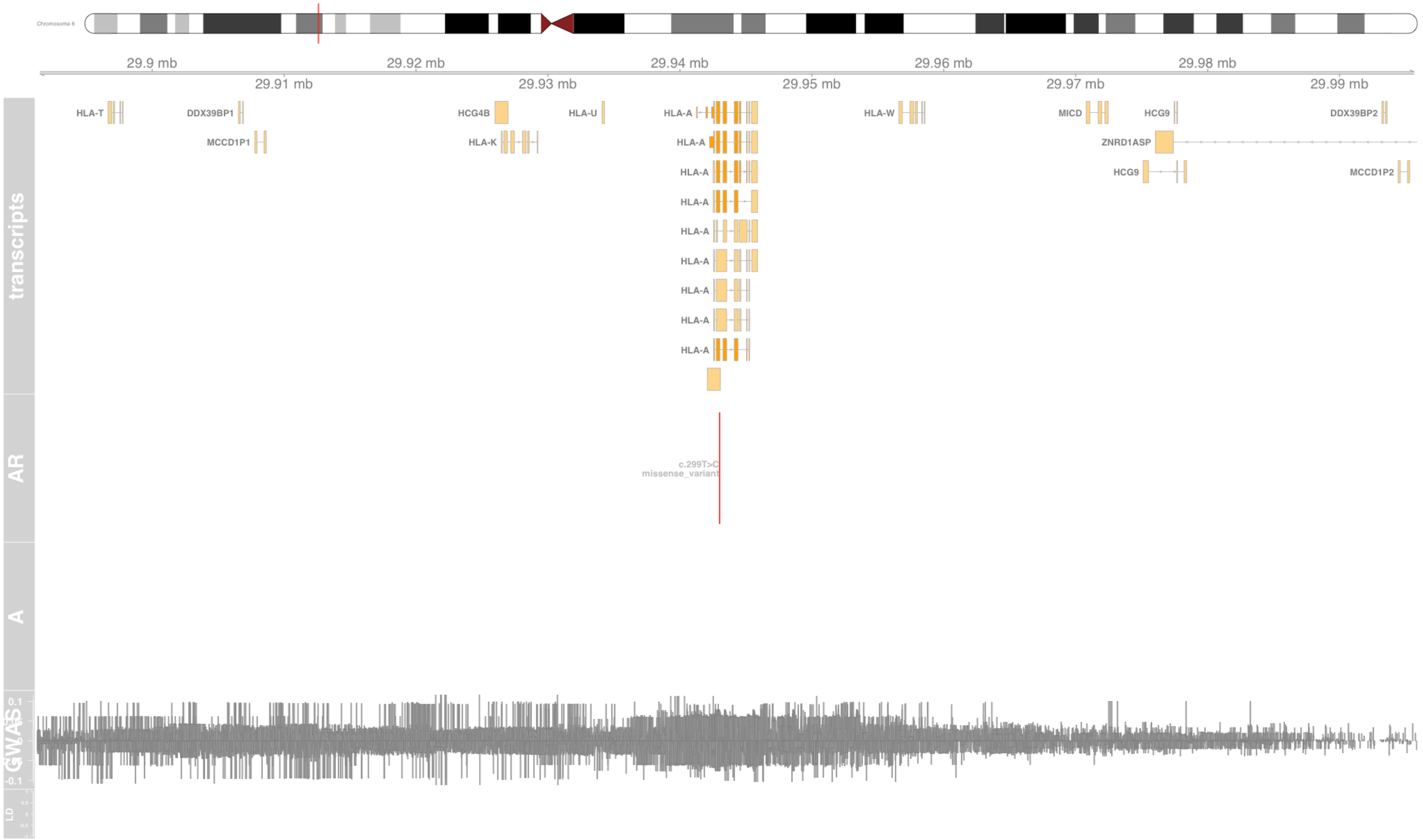

#### Fig S47 HLA-B

major histocompatibility complex, class I, B [Source:HGNC Symbol;Acc:HGNC:4932]

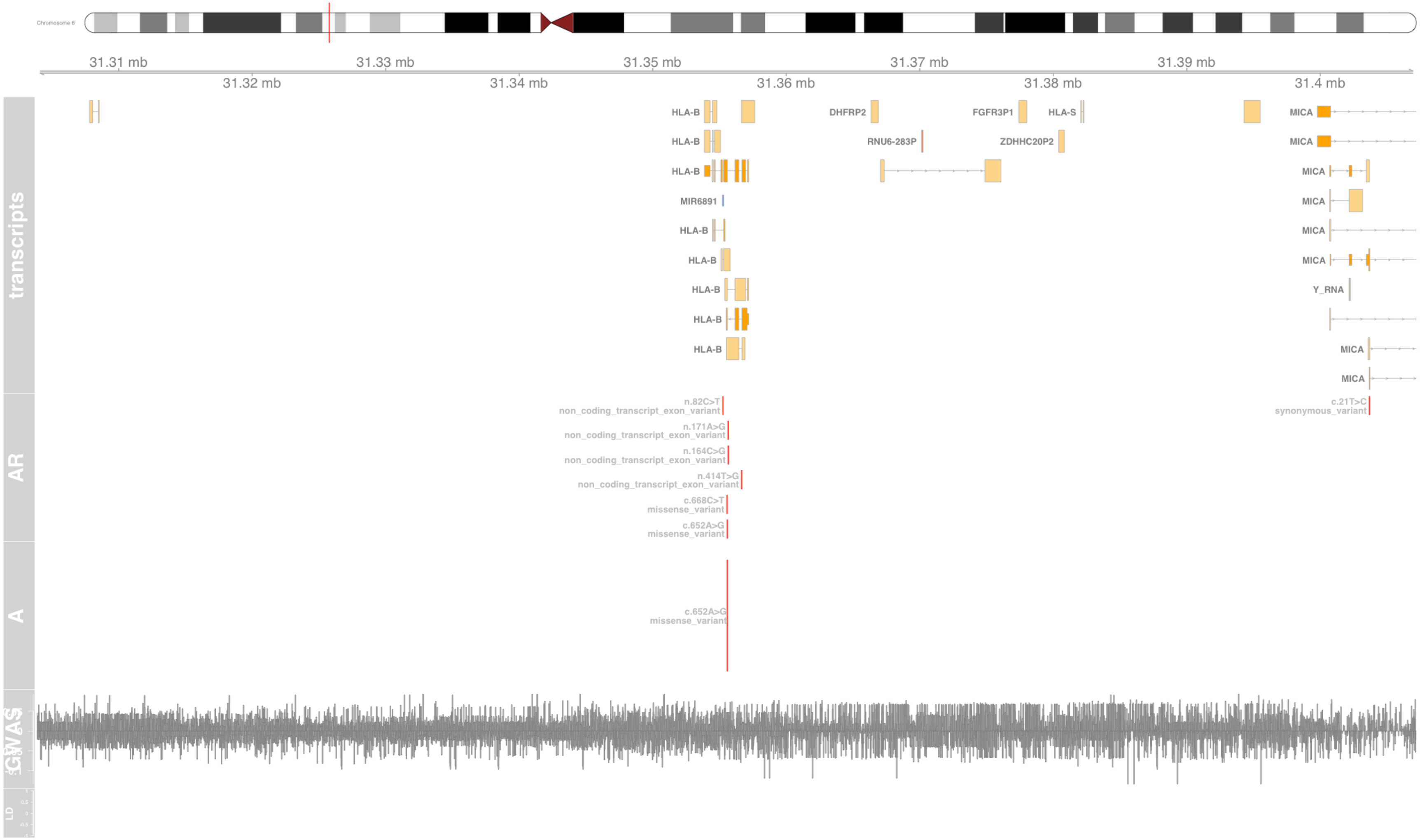

#### Fig S48 HLA-C

major histocompatibility complex, class I, C [Source:HGNC Symbol;Acc:HGNC:4933]

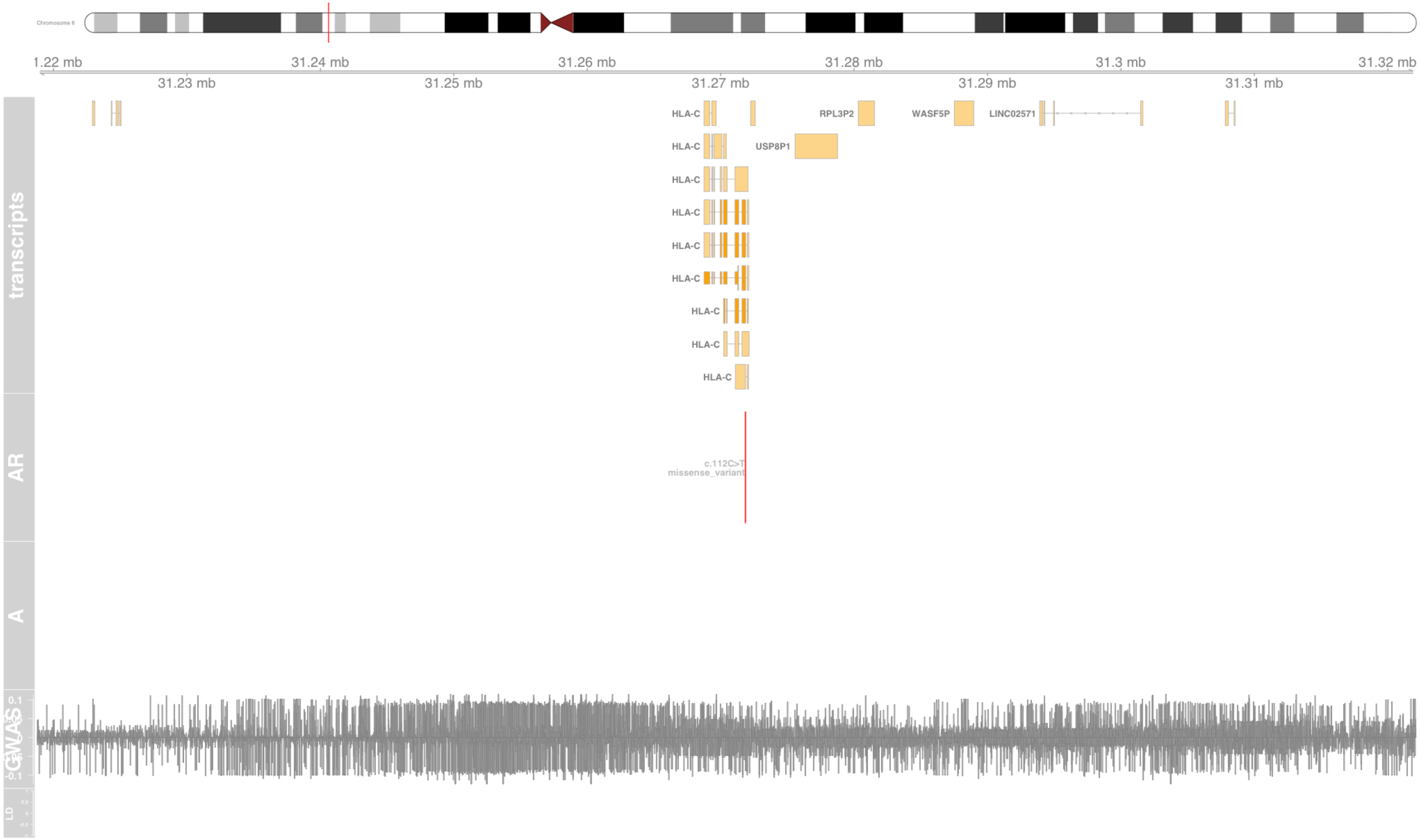

### Fig S49 HLA-DOB

major histocompatibility complex, class II, DO beta [Source:HGNC Symbol;Acc:HGNC:4937]

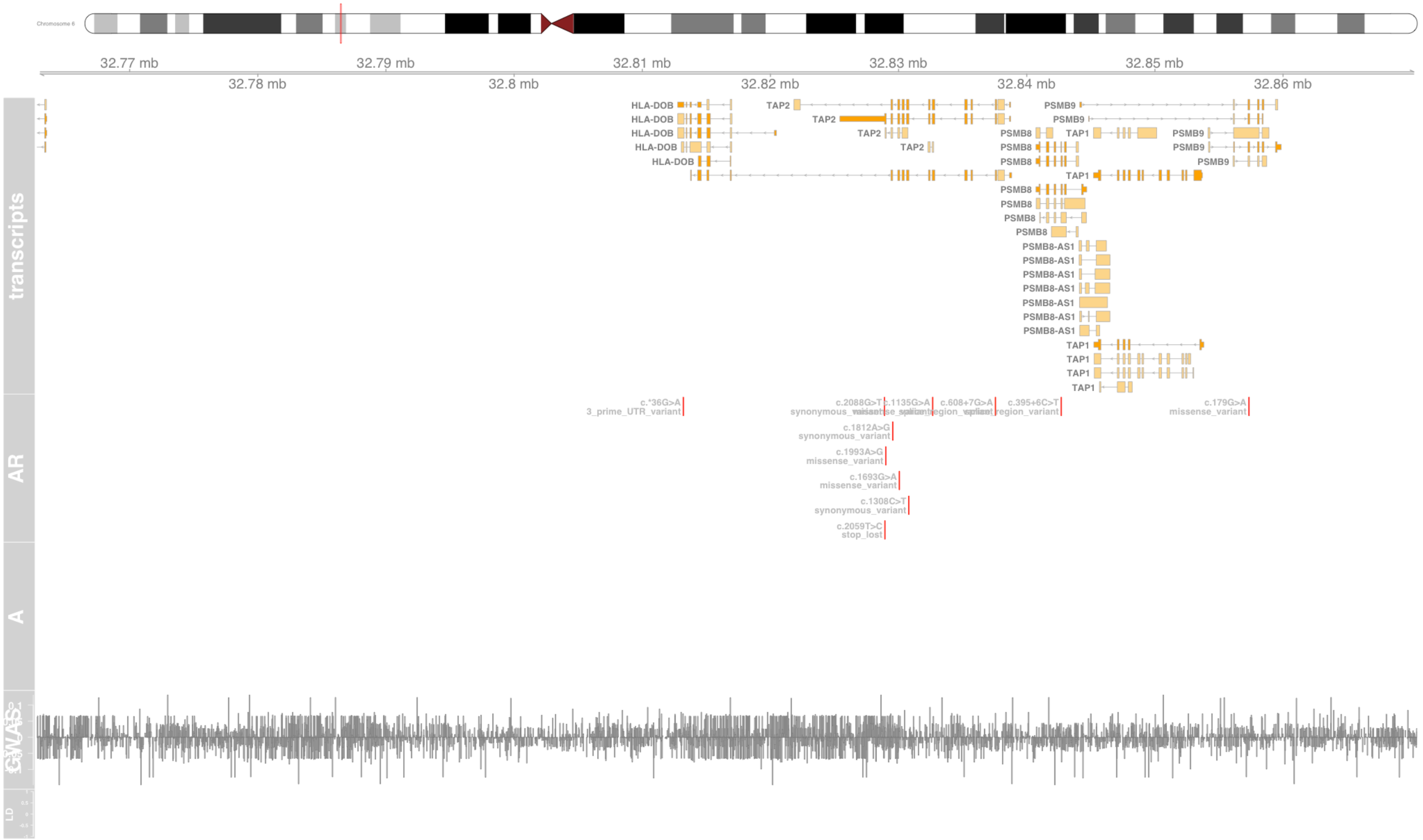

#### Fig S50 HLA-DQA1

major histocompatibility complex, class II, DQ alpha 1 [Source:HGNC Symbol;Acc:HGNC:4942]

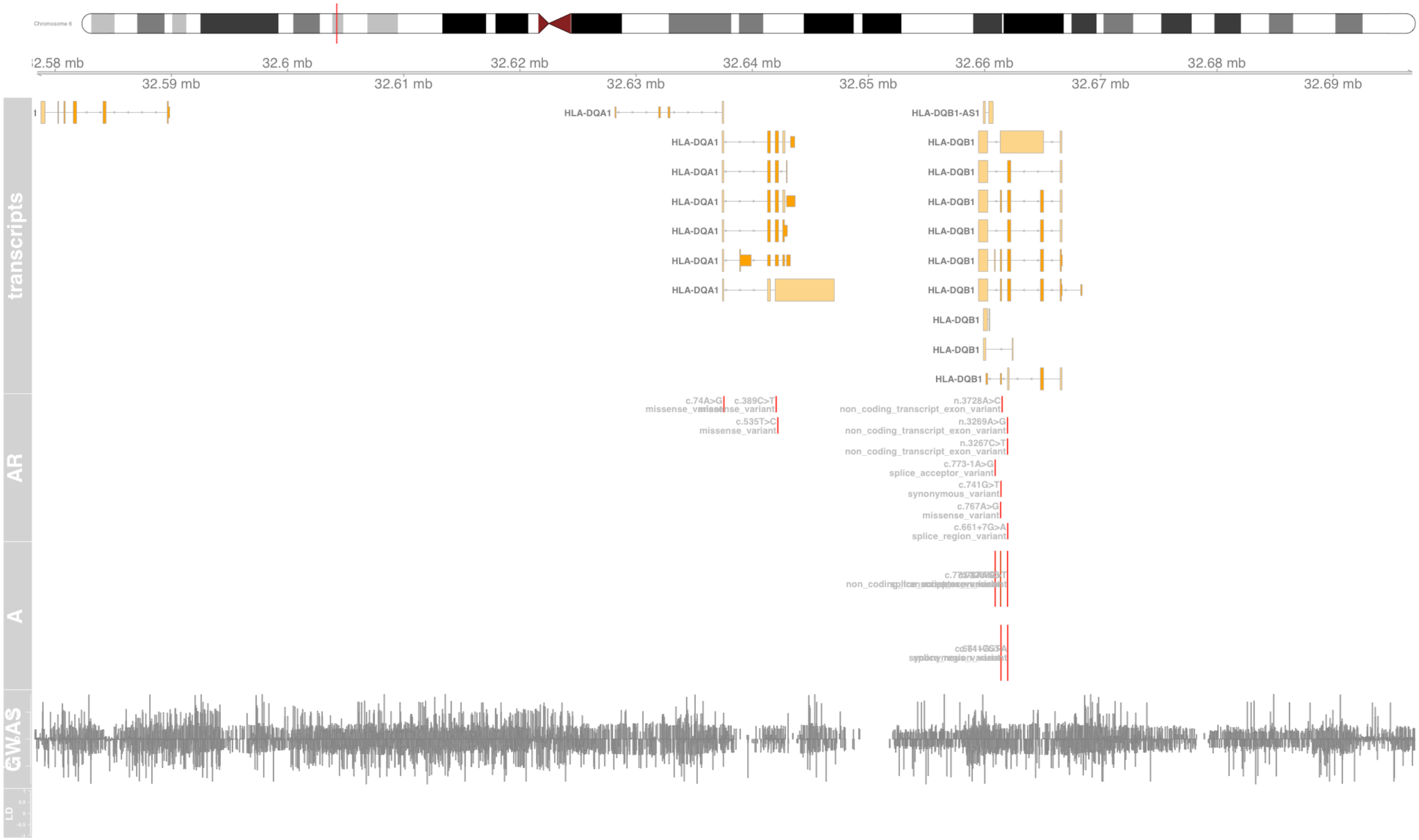

#### Fig S51 HLA-DQA2

major histocompatibility complex, class II, DQ alpha 2 [Source:HGNC Symbol;Acc:HGNC:4943]

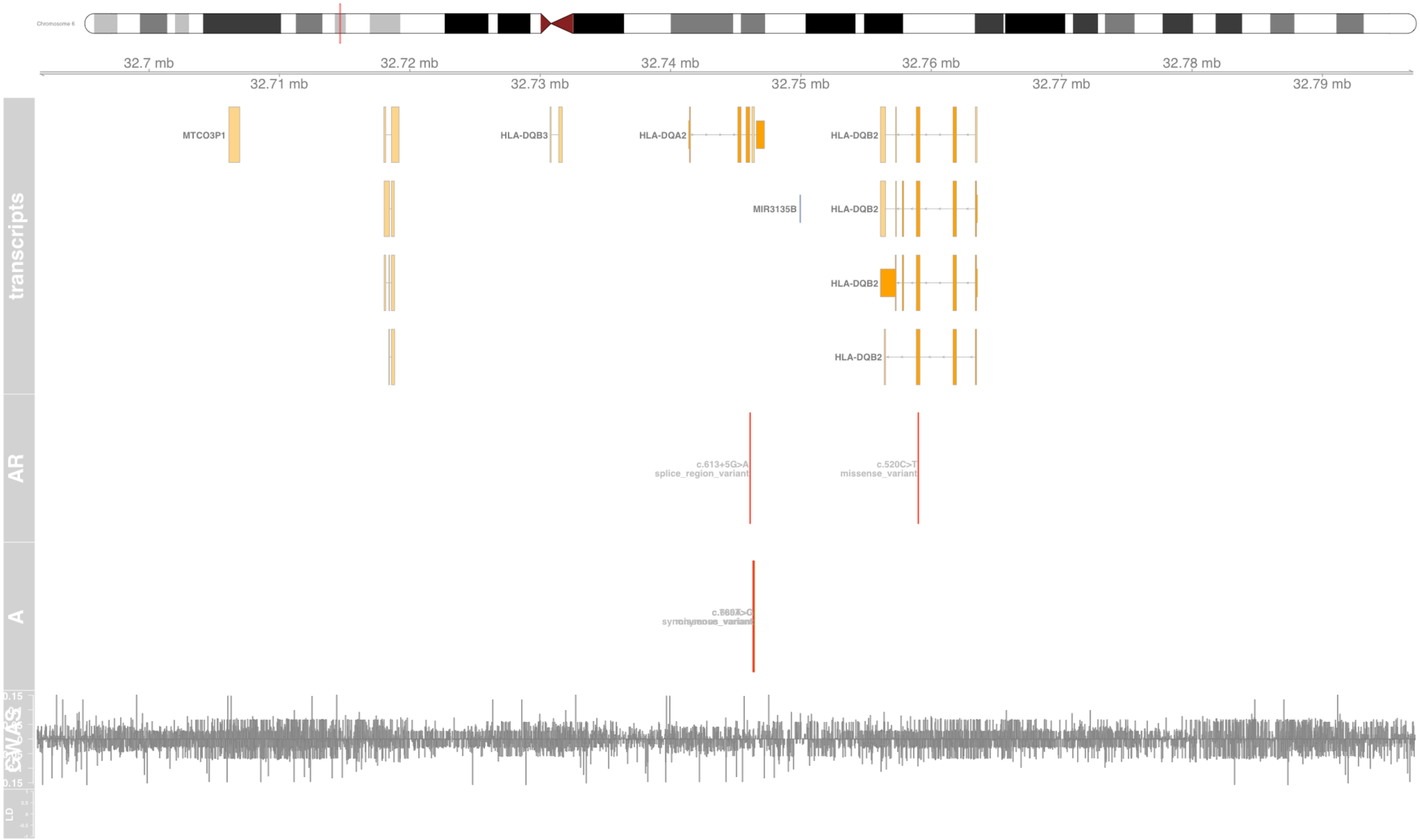

#### Fig S52 HLA-DQB1

major histocompatibility complex, class II, DQ beta 1 [Source:HGNC Symbol;Acc:HGNC:4944]

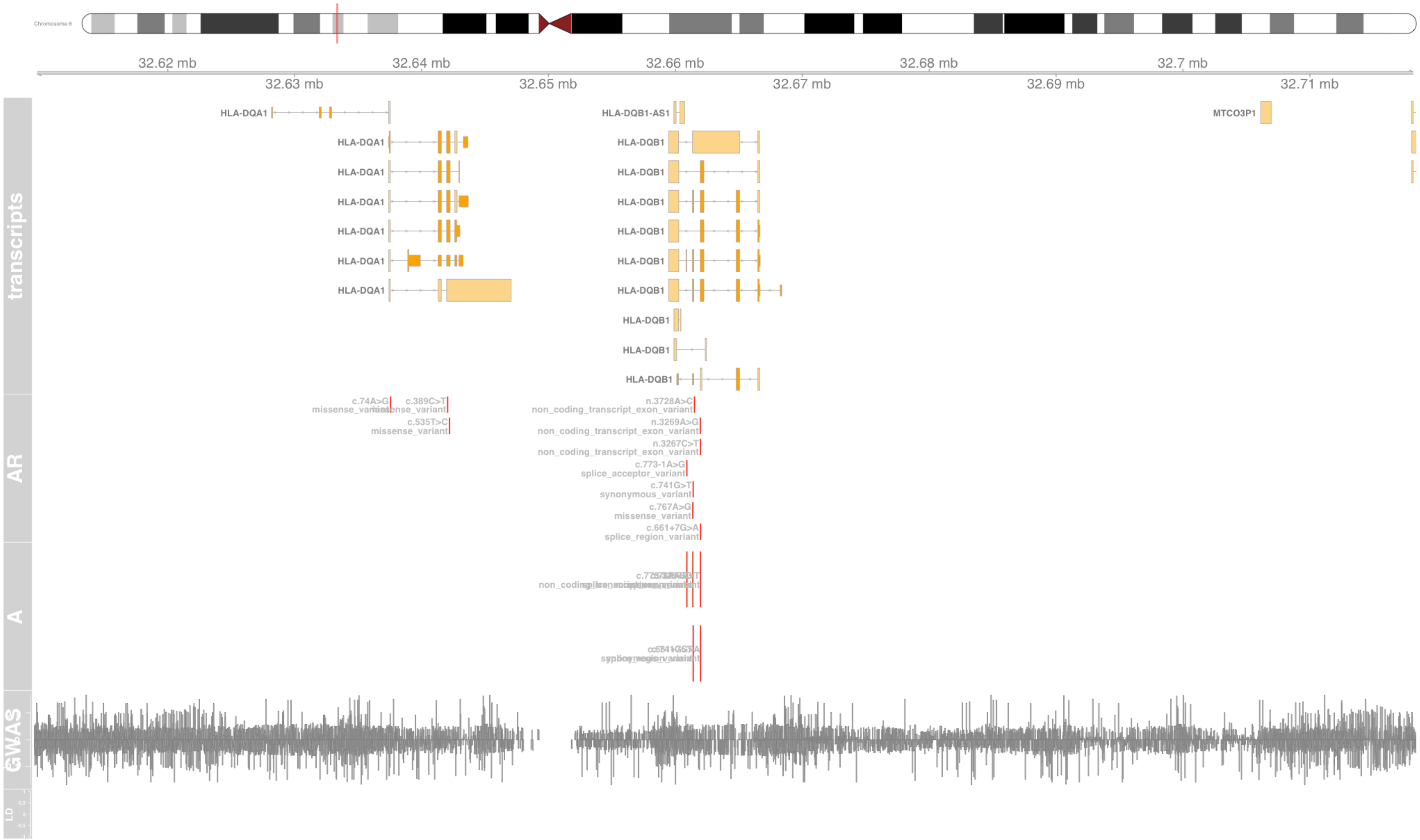

### Fig S53 HLA-DQB2

major histocompatibility complex, class II, DQ beta 2 [Source:HGNC Symbol;Acc:HGNC:4945]

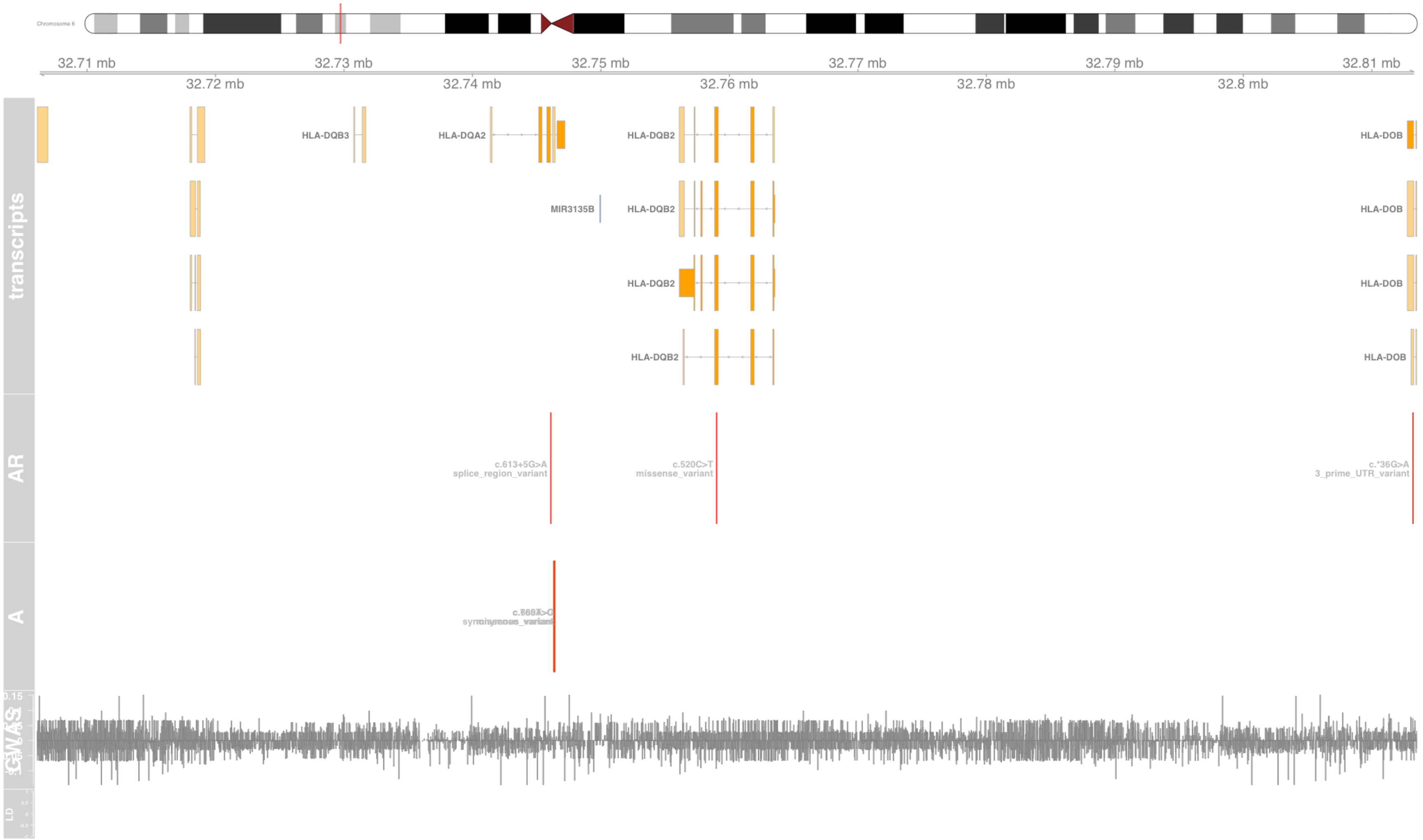

#### Fig S54 HLA-DRA

major histocompatibility complex, class II, DR alpha [Source:HGNC Symbol;Acc:HGNC:4947]

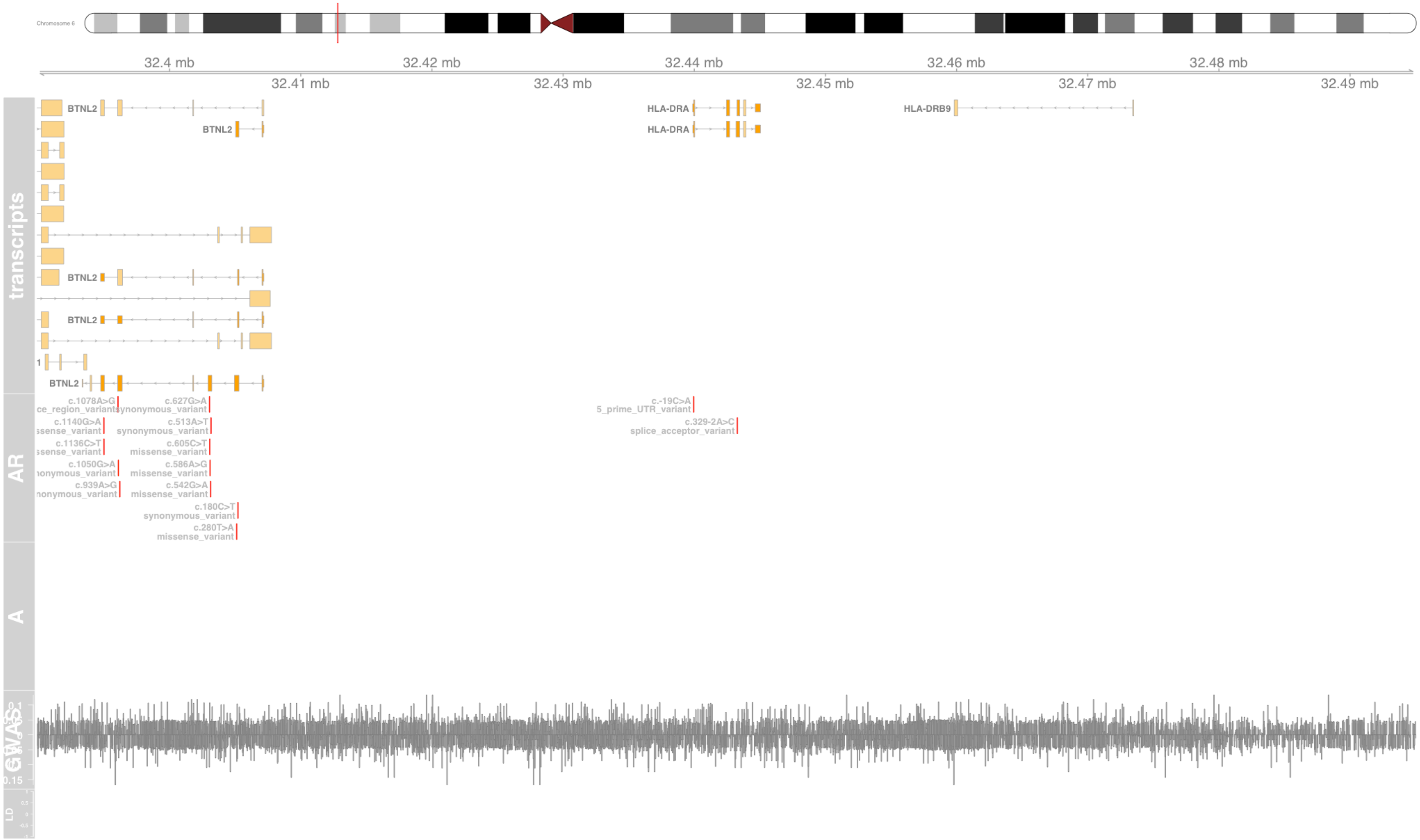

#### Fig S55 HLA-E

major histocompatibility complex, class I, E [Source:HGNC Symbol;Acc:HGNC:4962]

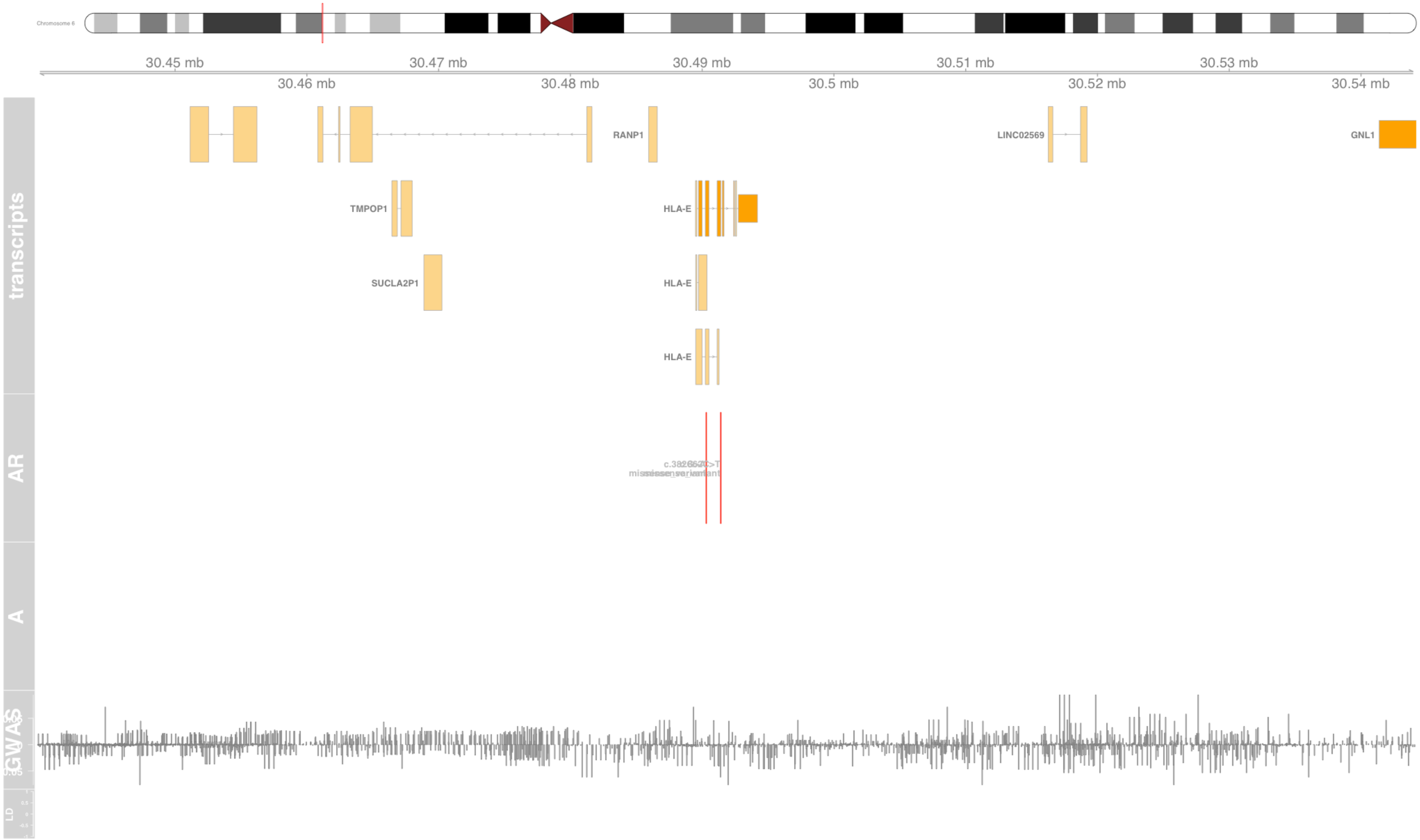

#### Fig S56 HLA-G

major histocompatibility complex, class I, G [Source:HGNC Symbol;Acc:HGNC:4964]

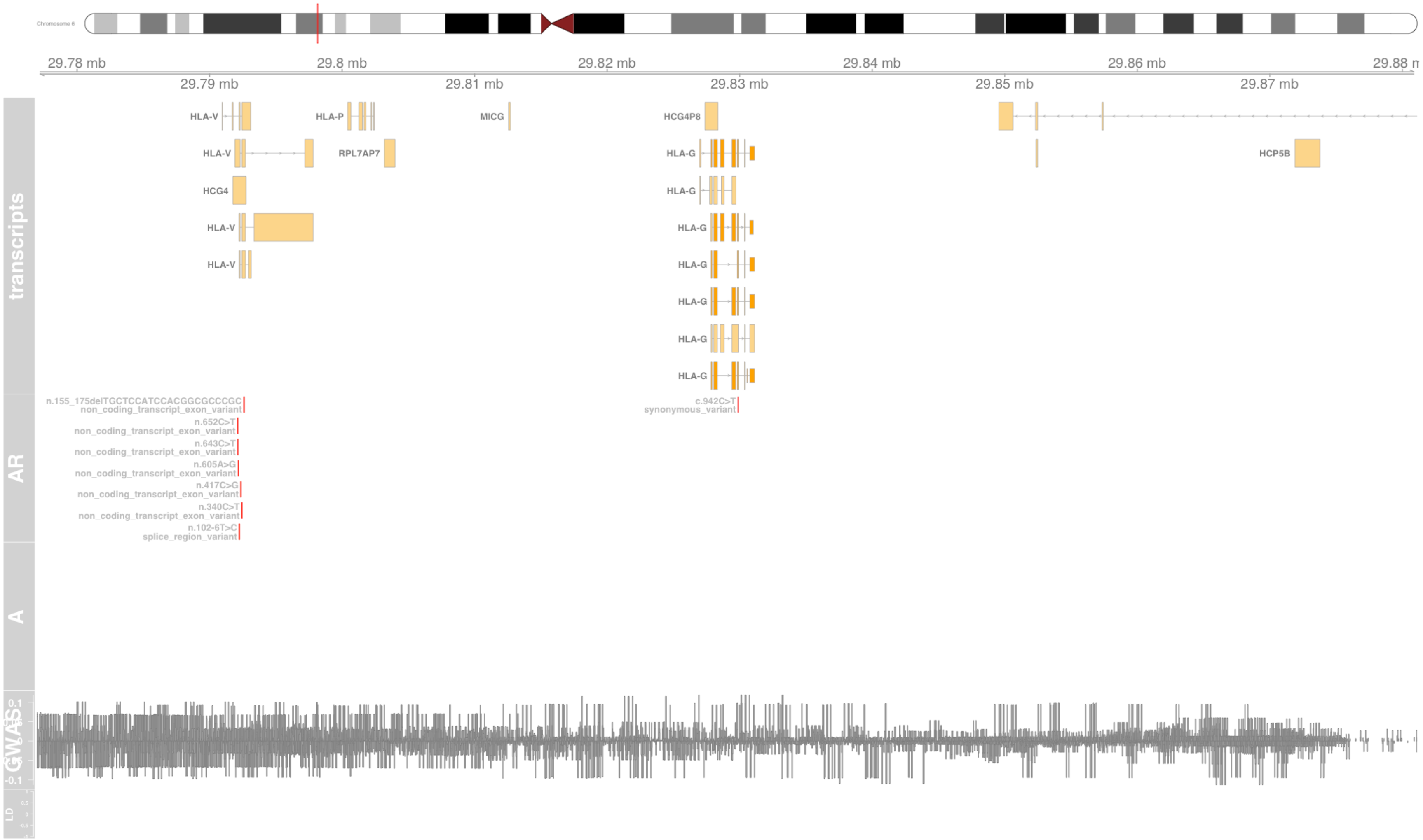

#### Fig S57 HLA-H

major histocompatibility complex, class I, H (pseudogene) [Source:HGNC Symbol;Acc:HGNC:4965]

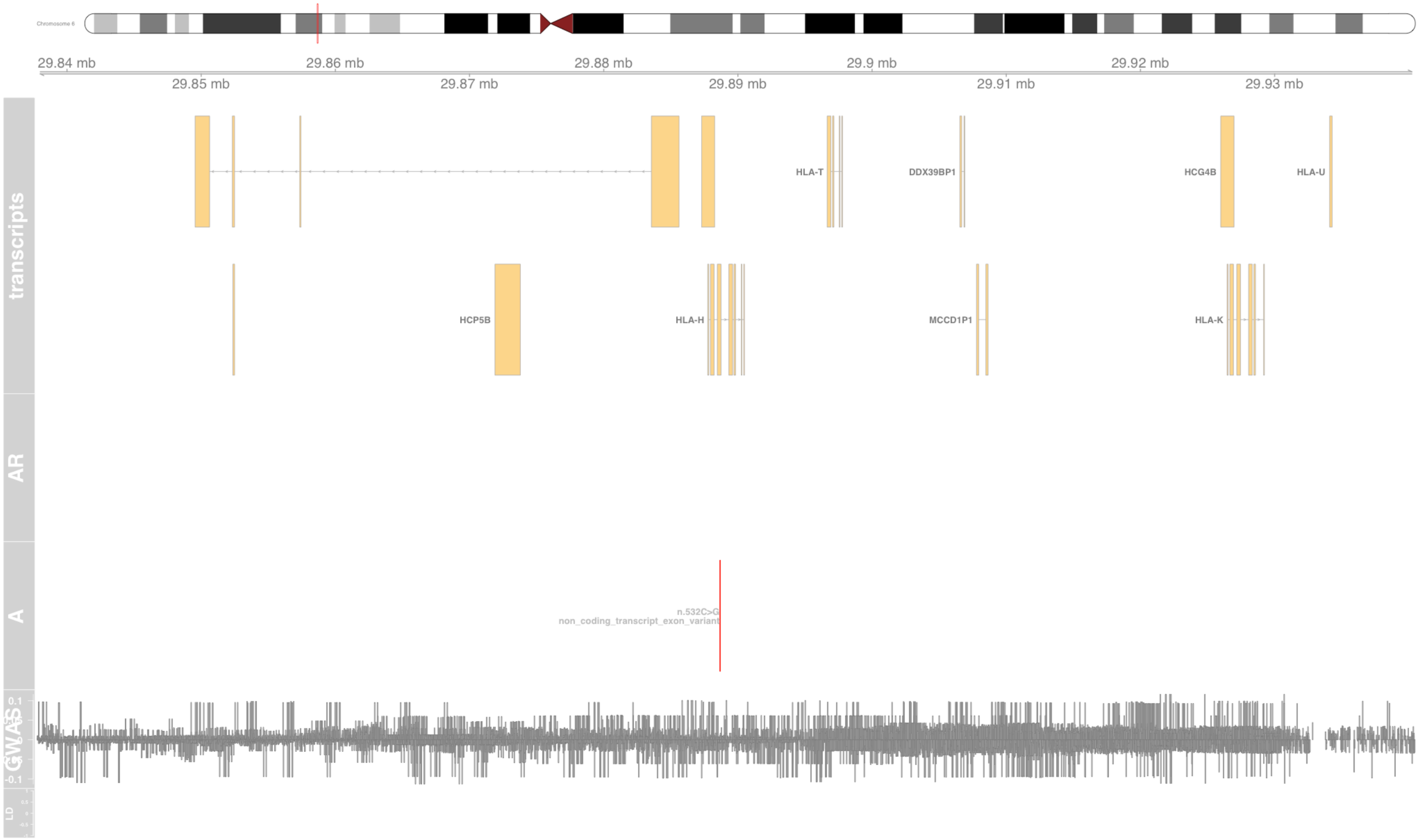

#### Fig S58 HLA-V

major histocompatibility complex, class I, V (pseudogene) [Source:HGNC Symbol;Acc:HGNC:23482]

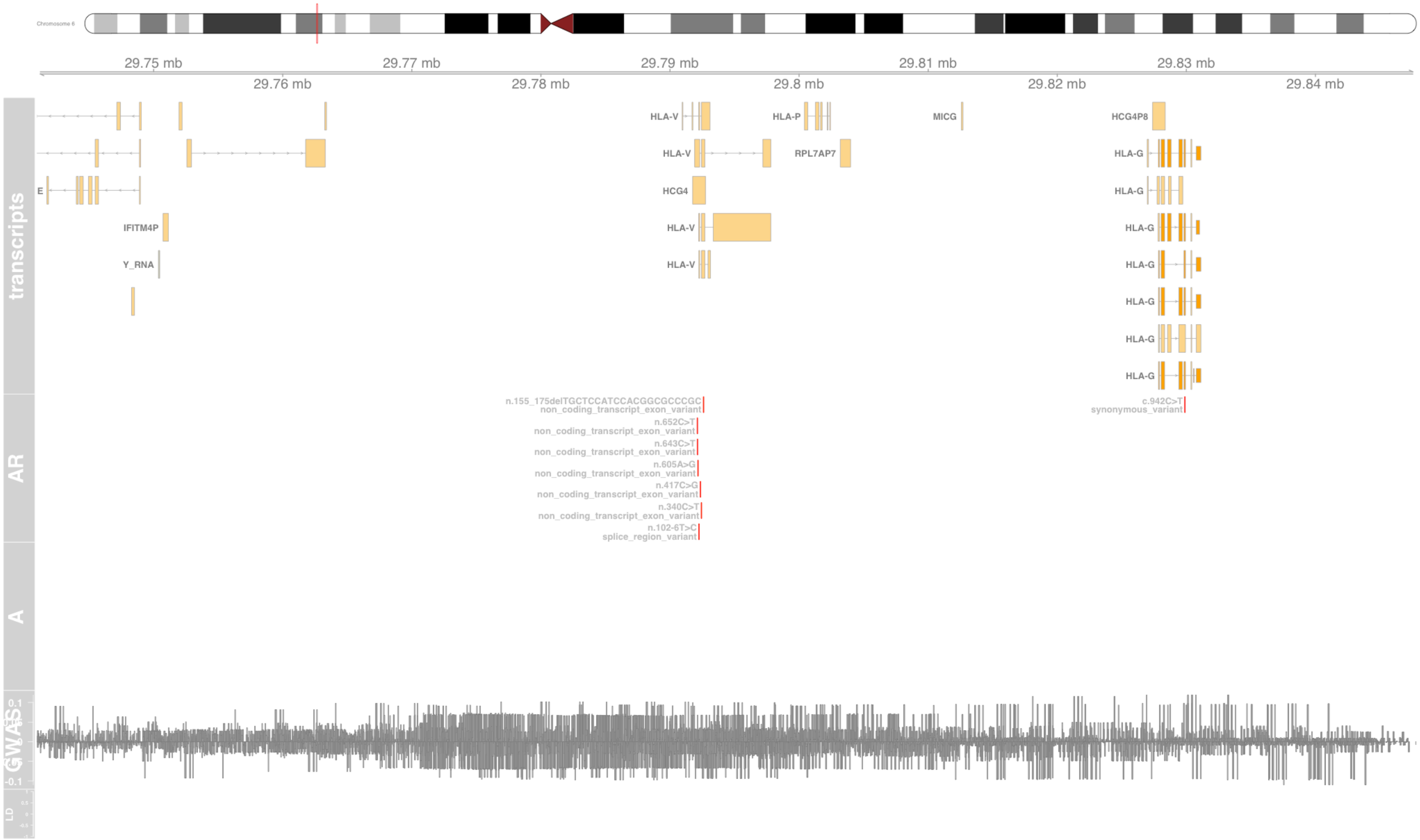

#### Fig S59 HSPA1L

heat shock protein family A (Hsp70) member 1 like [Source:HGNC Symbol;Acc:HGNC:5234]

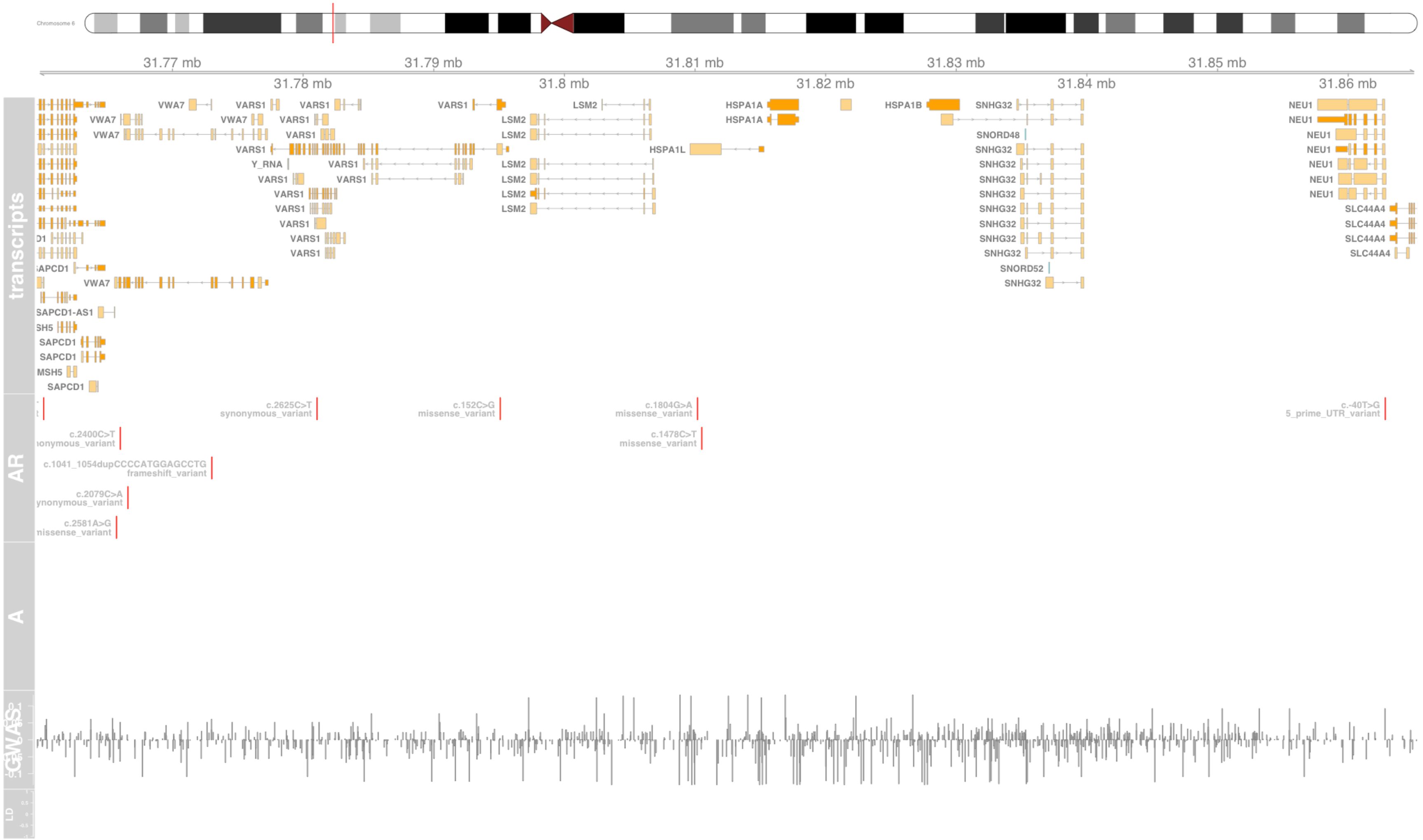

#### Fig S60 IER3

immediate early response 3 [Source:HGNC Symbol;Acc:HGNC:5392]

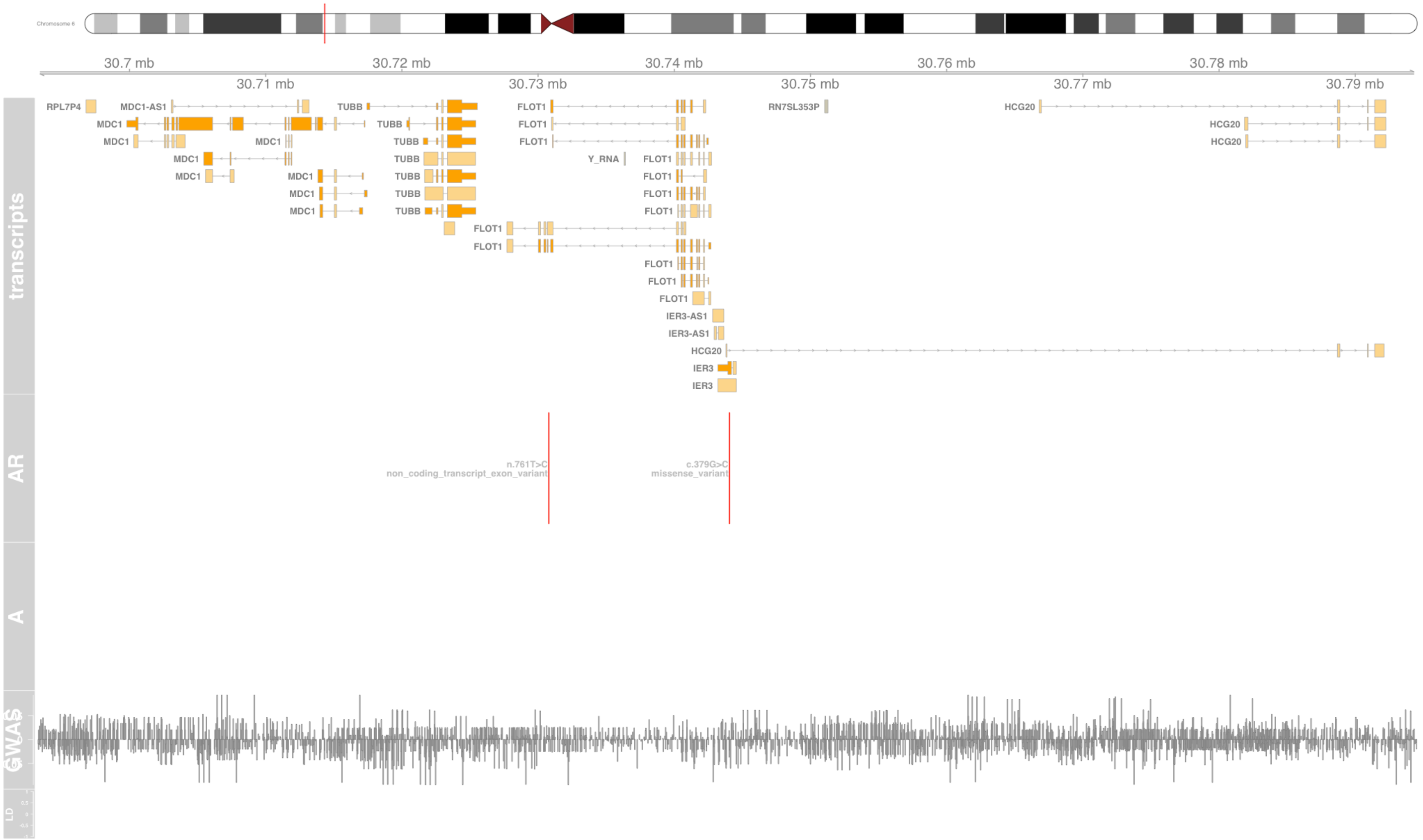

#### Fig S61 IKZF3

IKAROS family zinc finger 3 [Source:HGNC Symbol;Acc:HGNC:13178]

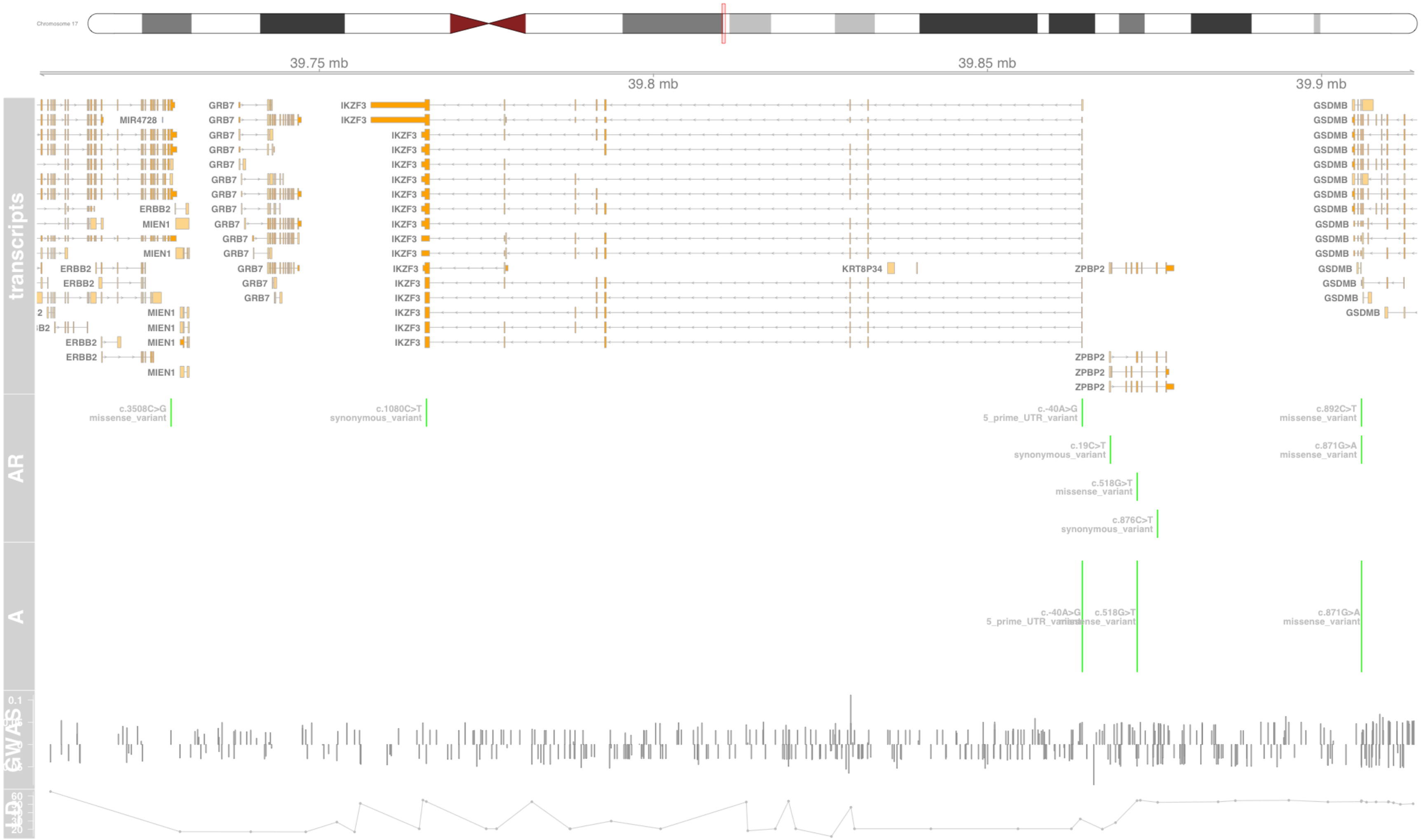

#### Fig S62 IL13

interleukin 13 [Source:HGNC Symbol;Acc:HGNC:5973]

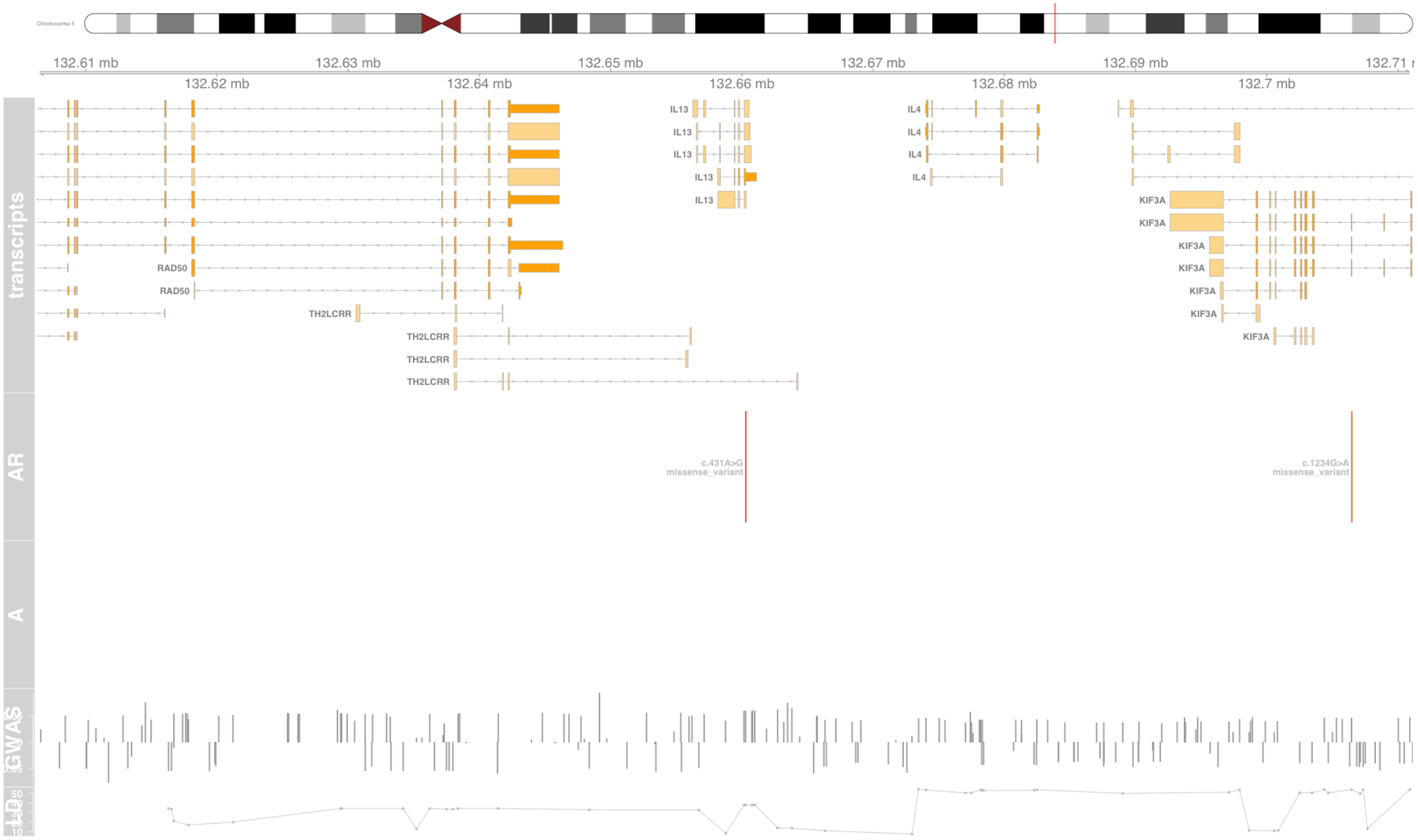

#### Fig S63 IL18R1

interleukin 18 receptor 1 [Source:HGNC Symbol;Acc:HGNC:5988]

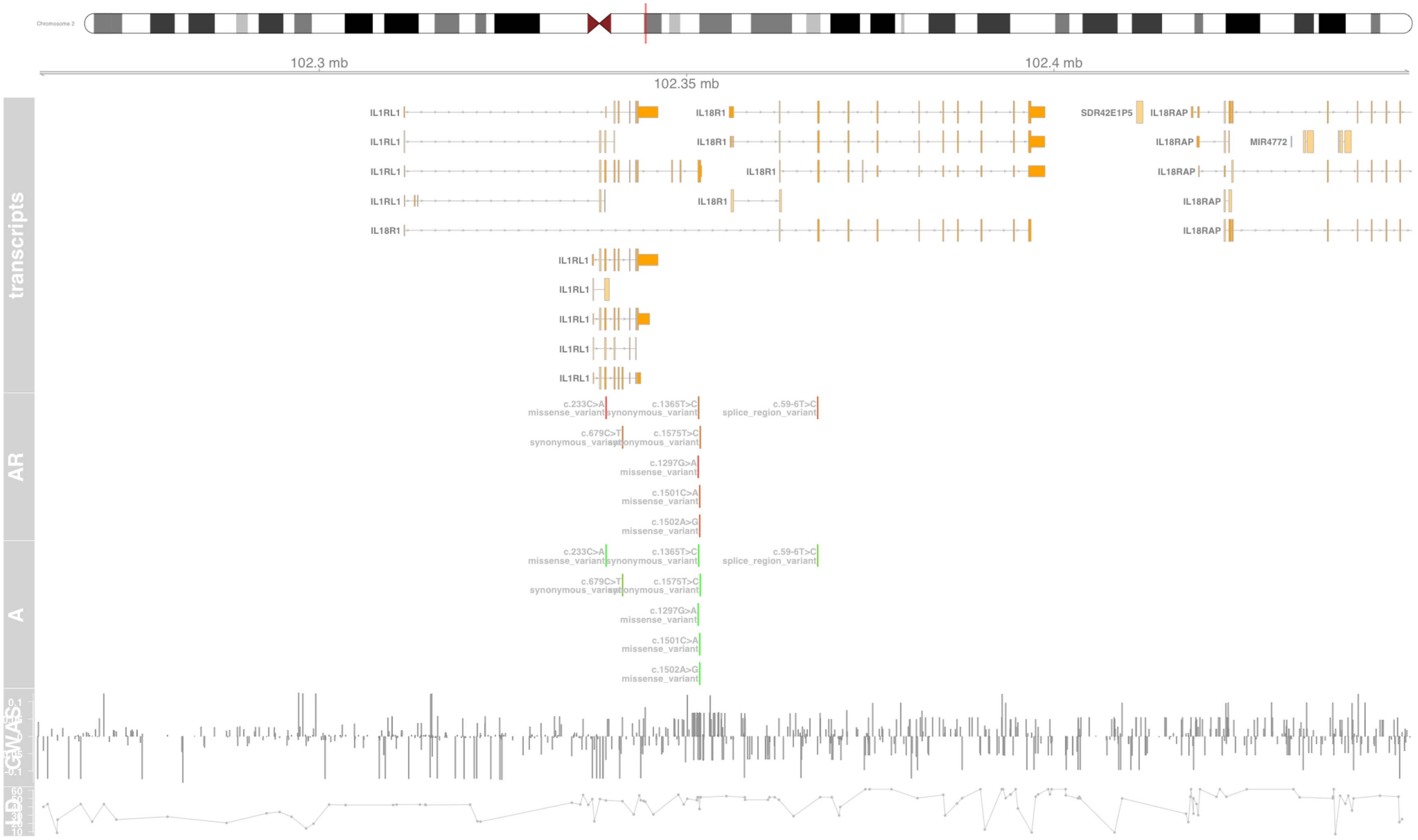

#### Fig S64 IL1RL1

interleukin 1 receptor like 1 [Source:HGNC Symbol;Acc:HGNC:5998]

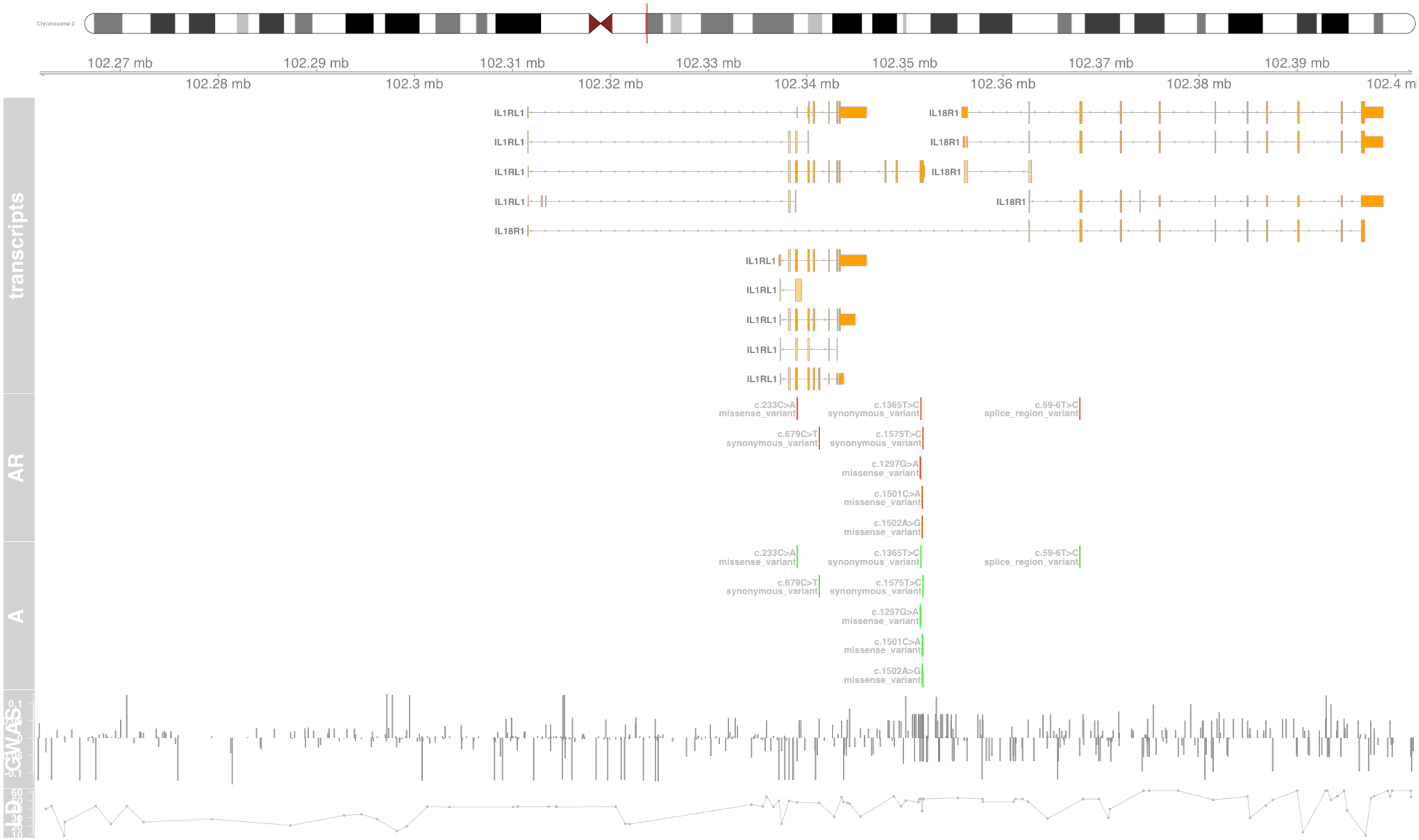

#### Fig S65 IL1RL2

interleukin 1 receptor like 2 [Source:HGNC Symbol;Acc:HGNC:5999]

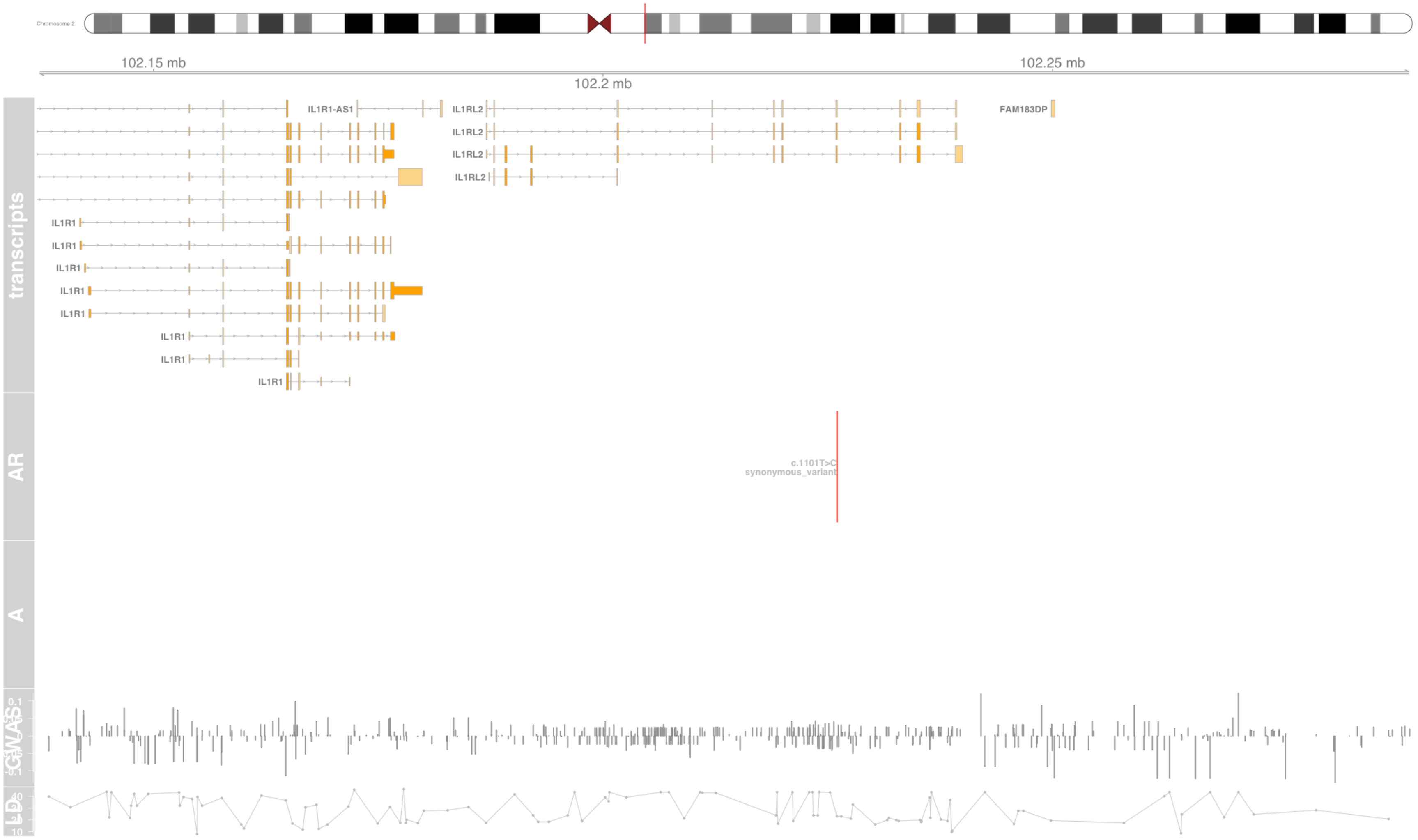

#### Fig S66 IL2

interleukin 2 [Source:HGNC Symbol;Acc:HGNC:6001]

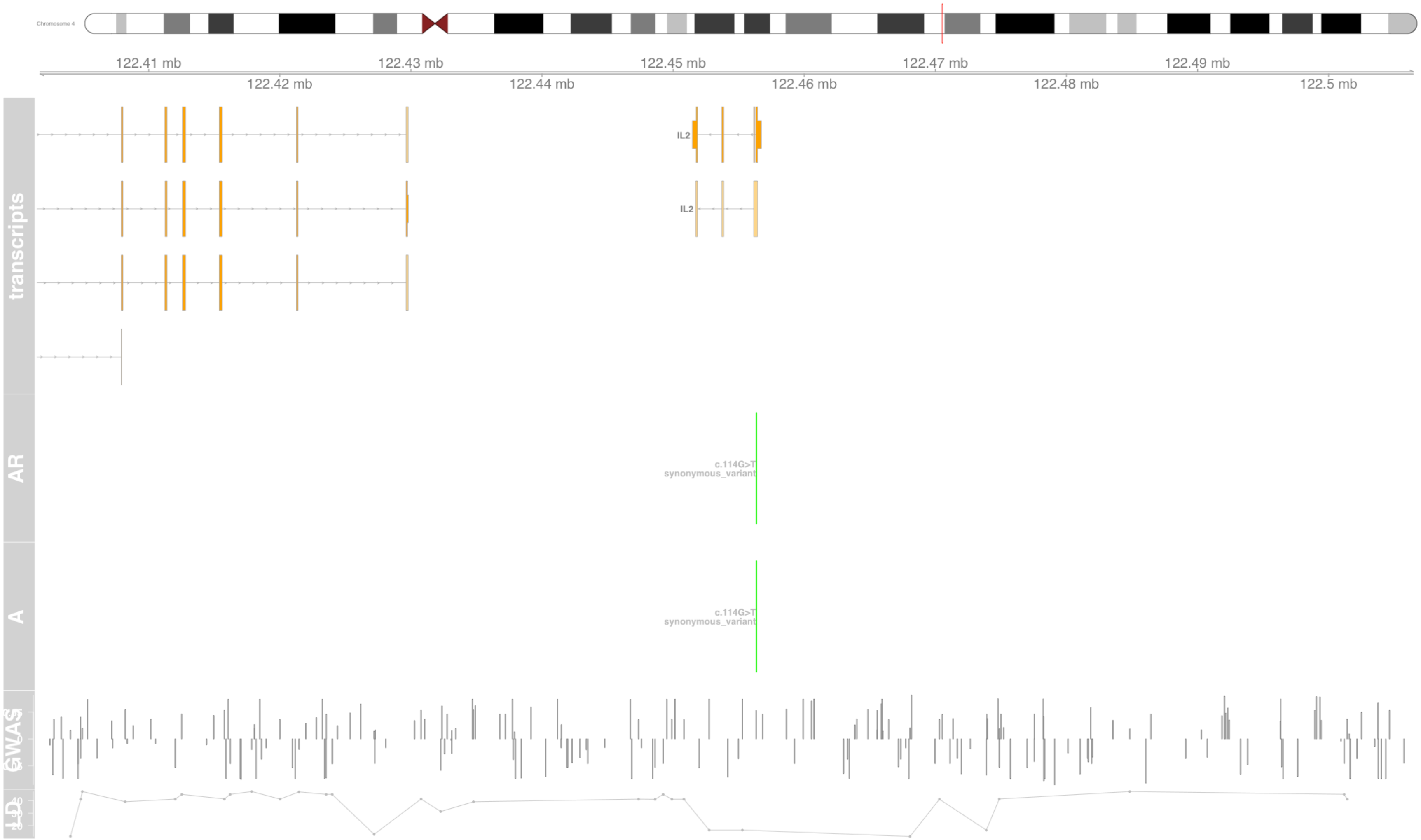

#### Fig S67 IL33

interleukin 33 [Source:HGNC Symbol;Acc:HGNC:16028]

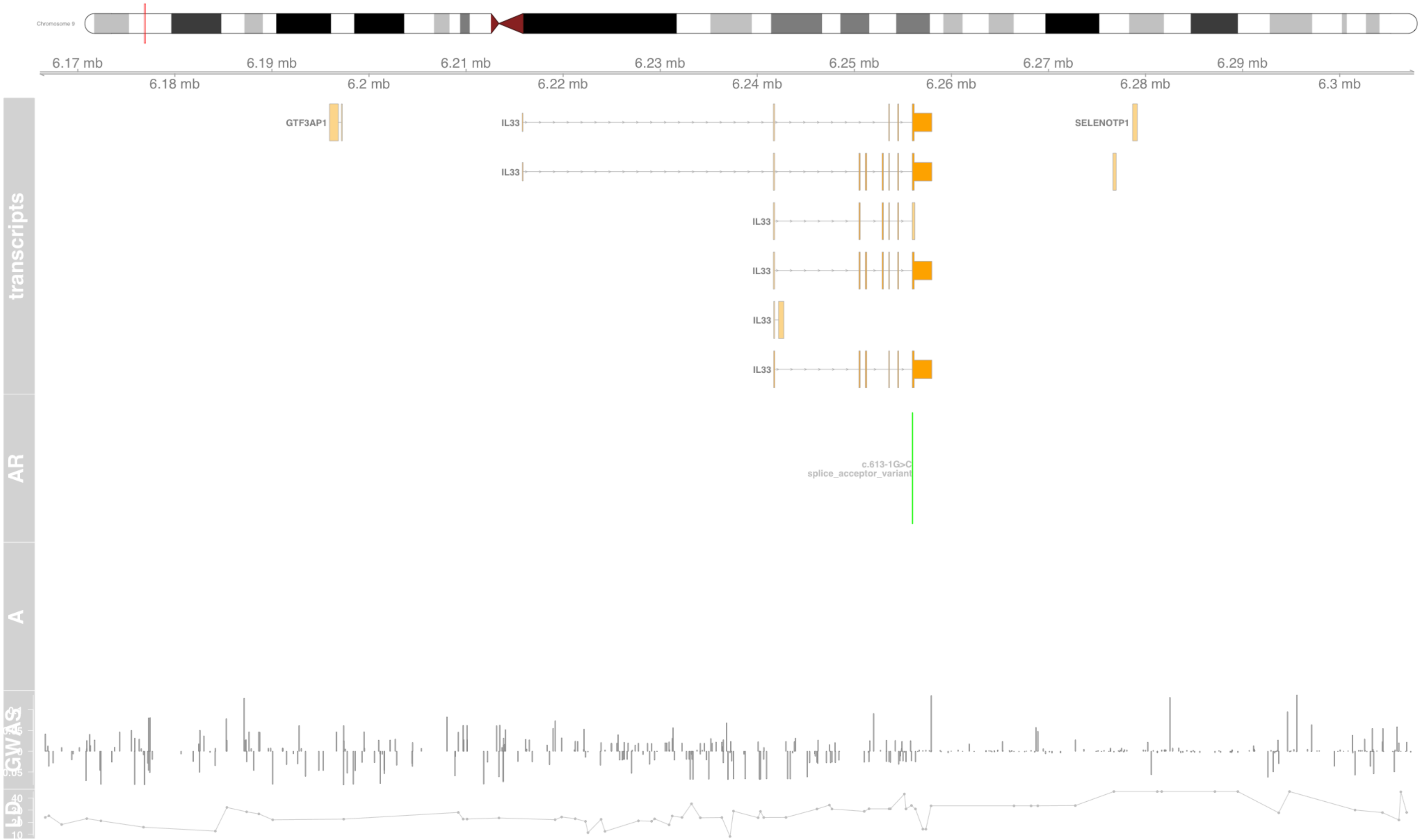

### Fig S68 IL4R

interleukin 4 receptor [Source:HGNC Symbol;Acc:HGNC:6015]

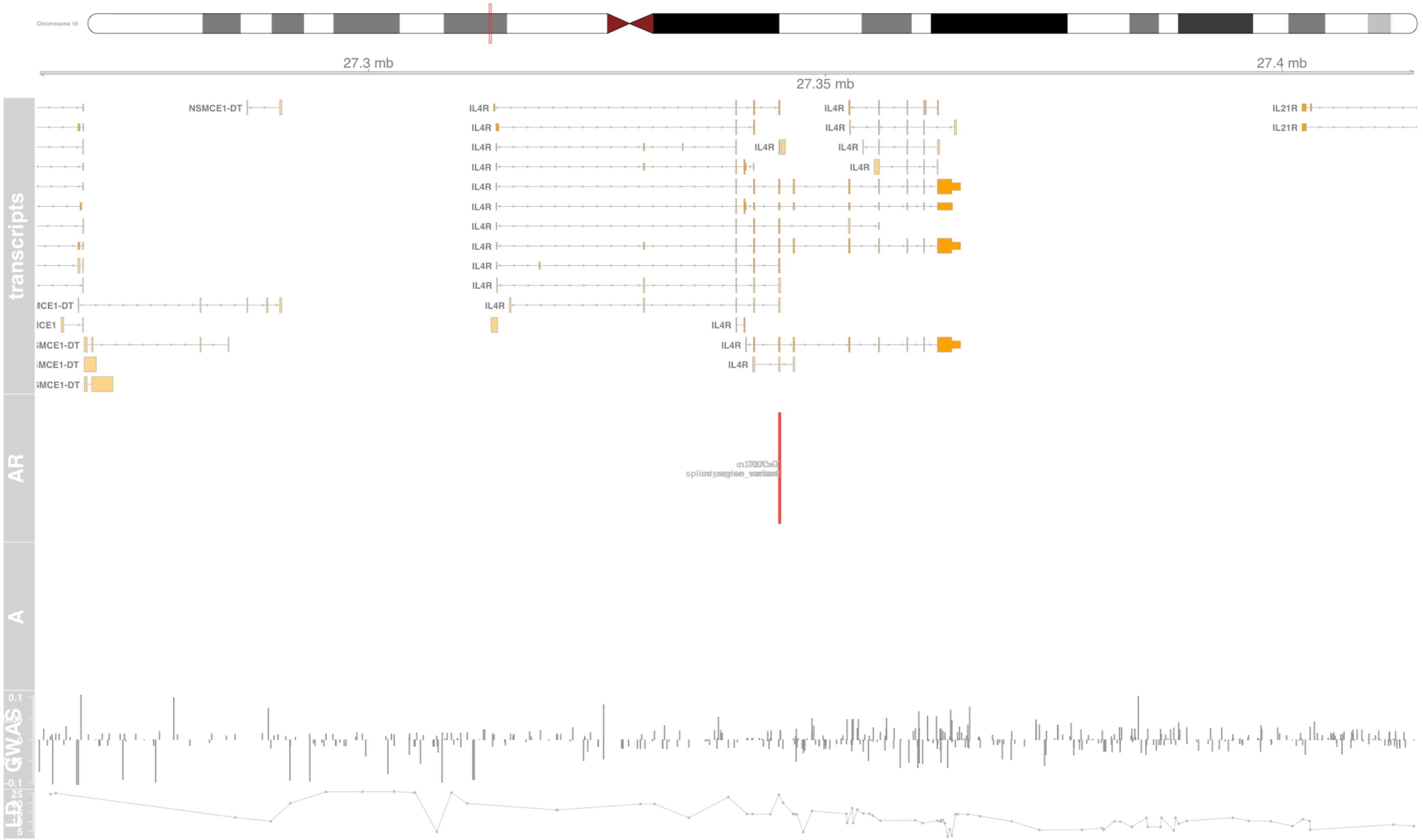

### Fig S69 IRF1

interferon regulatory factor 1 [Source:HGNC Symbol;Acc:HGNC:6116]

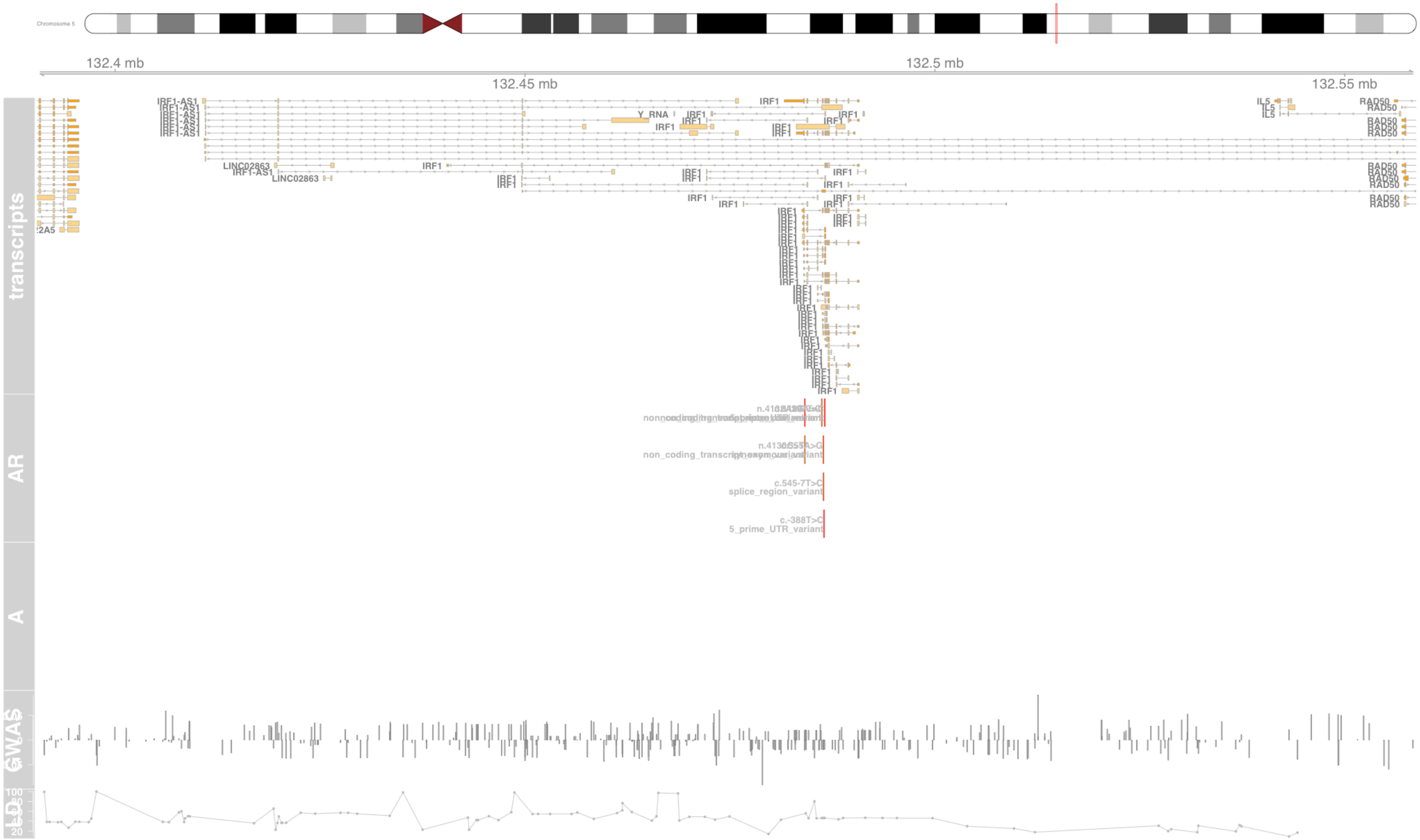

### Fig S70 KIAA1109

KIAA1109 [Source:HGNC Symbol;Acc:HGNC:26953]

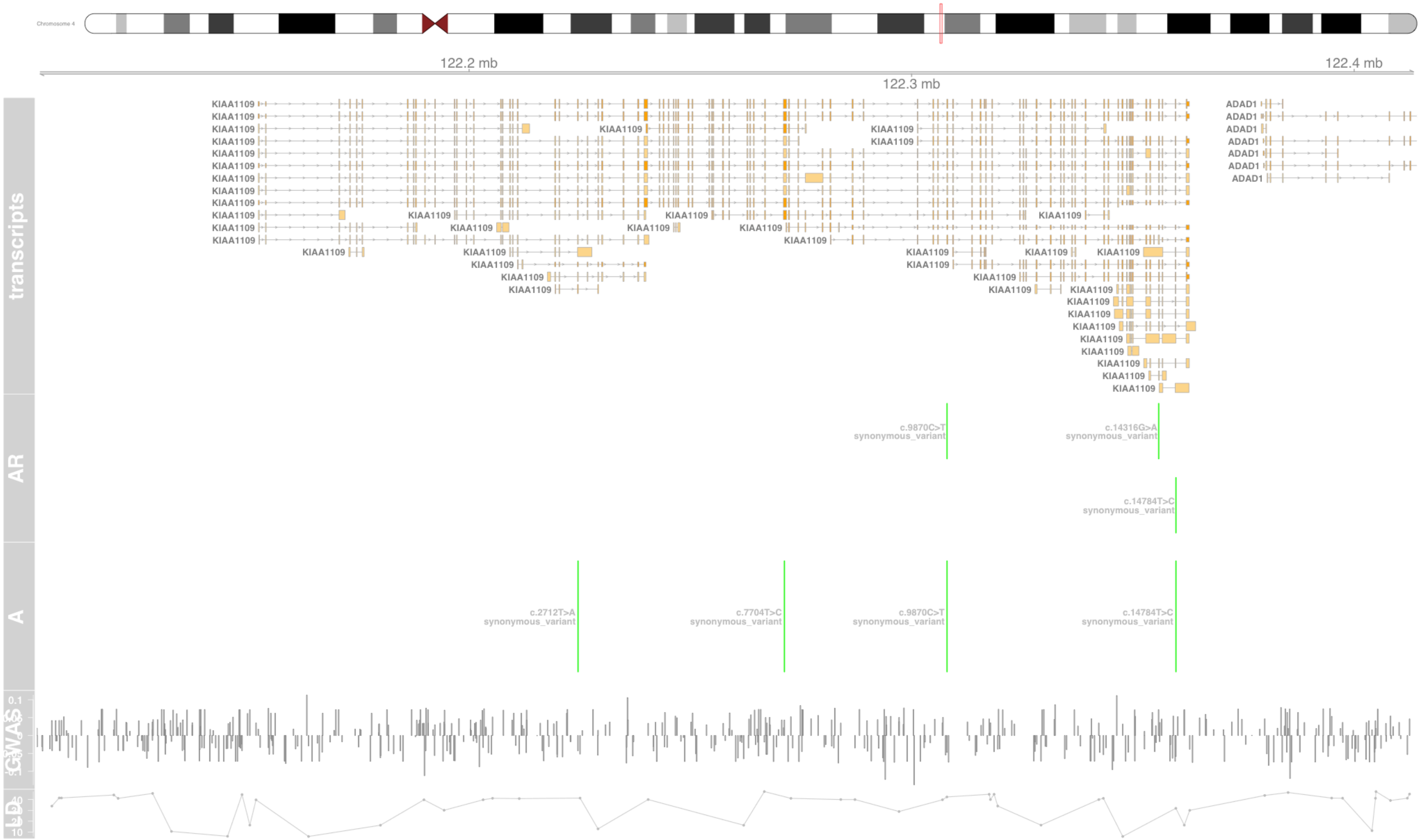

### Fig S71 KIF3A

kinesin family member 3A [Source:HGNC Symbol;Acc:HGNC:6319]

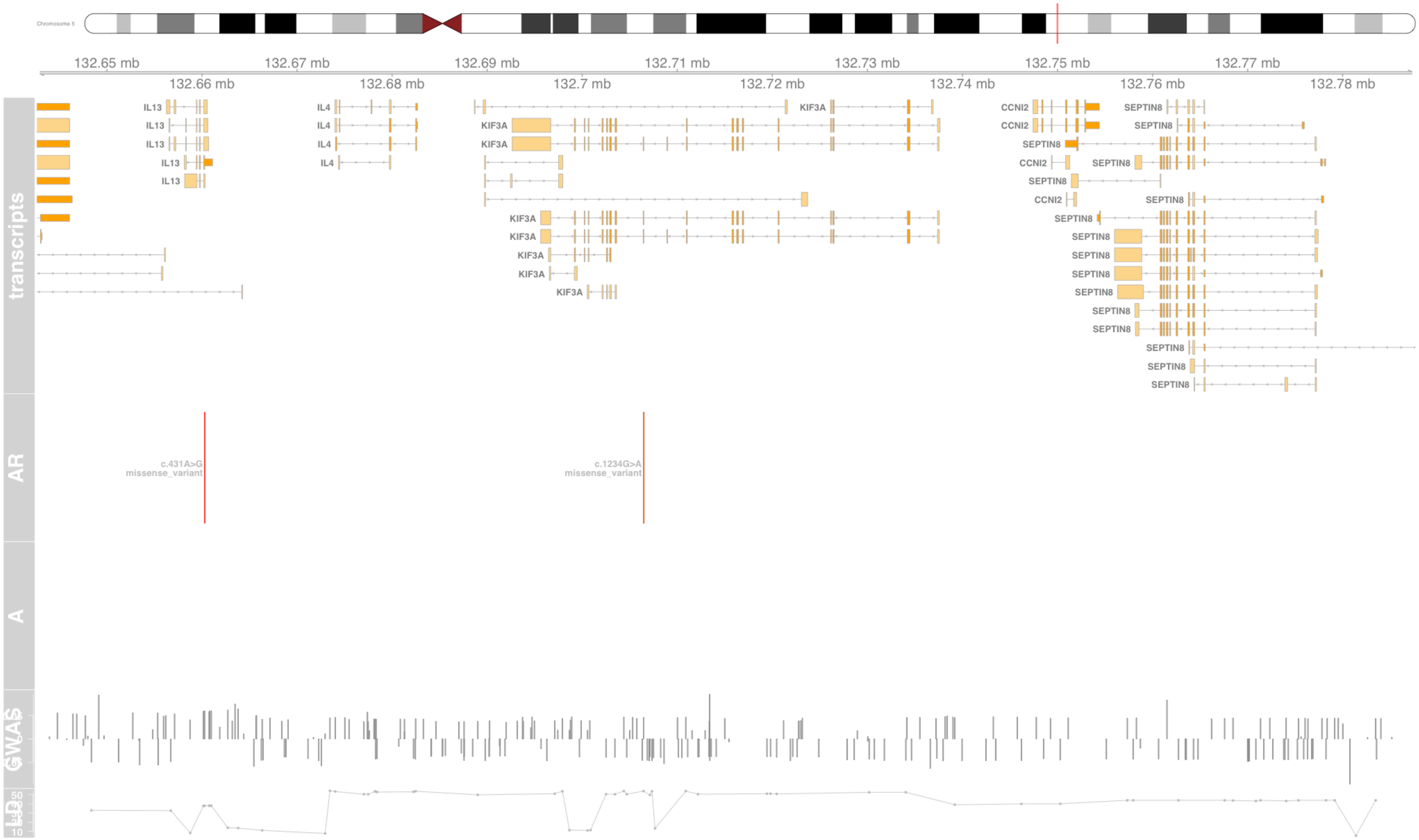

#### Fig S72 LRP1

LDL receptor related protein 1 [Source:HGNC Symbol;Acc:HGNC:6692]

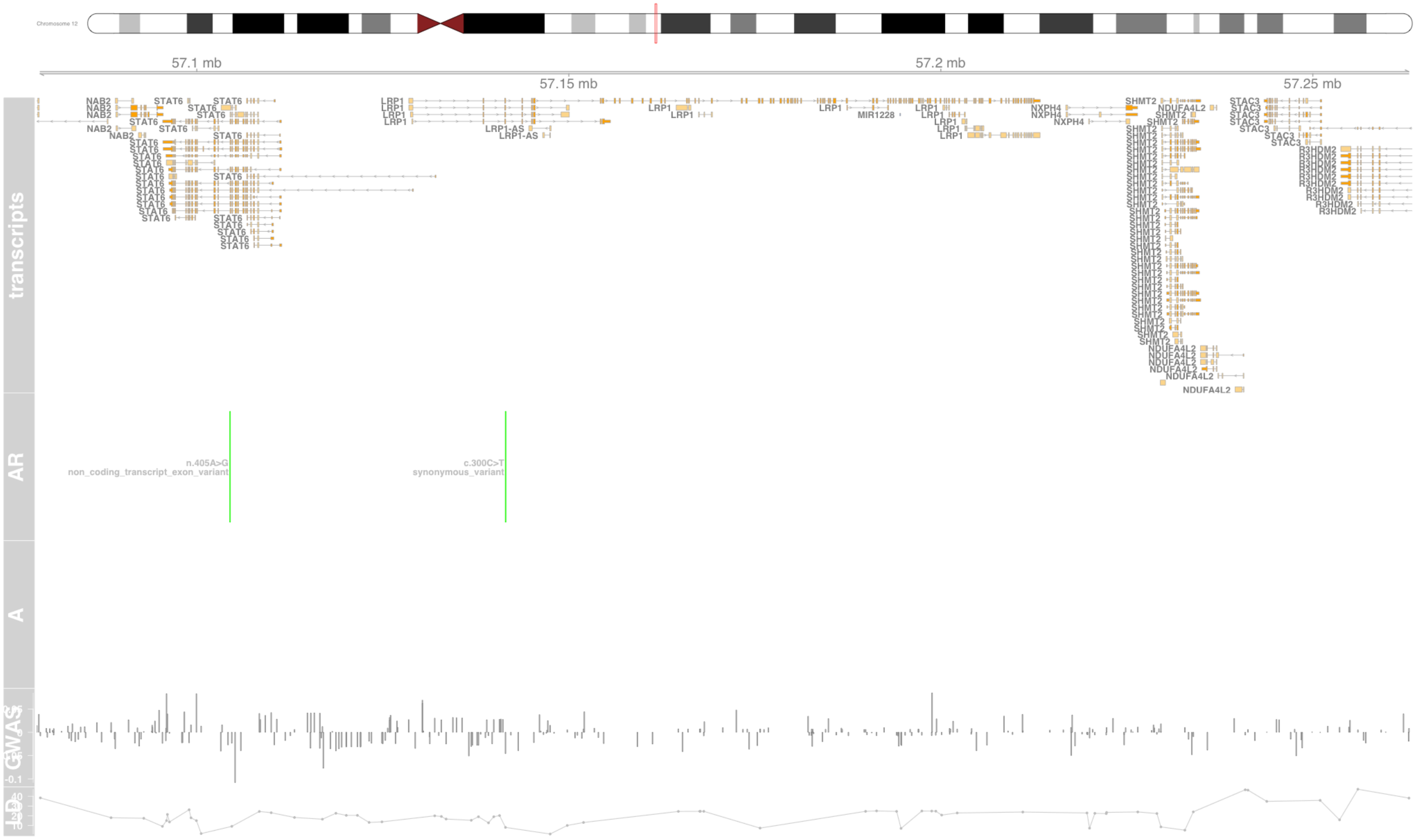

#### Fig S73 LTA

lymphotoxin alpha [Source:HGNC Symbol;Acc:HGNC:6709]

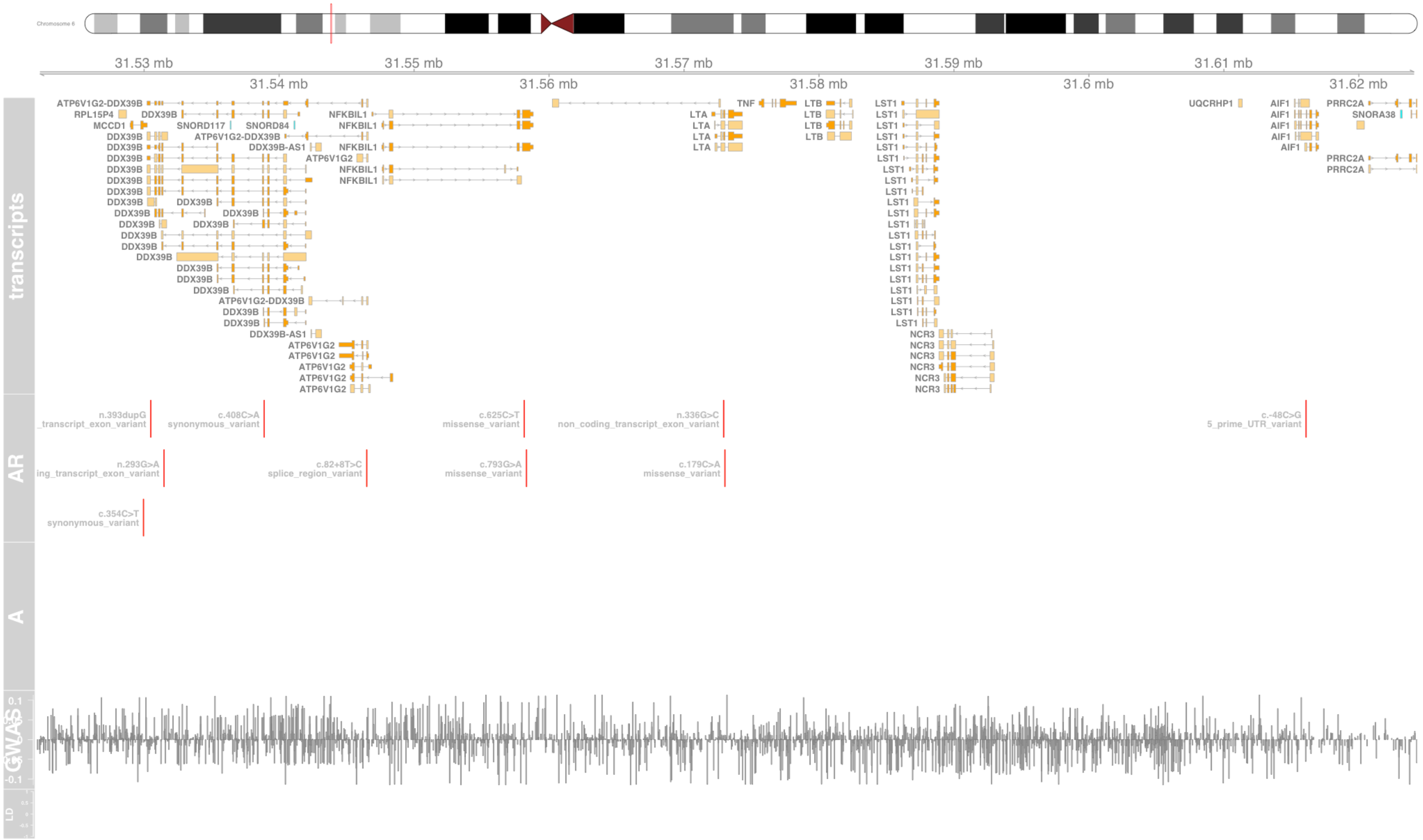

### Fig S74 LY6G6C

lymphocyte antigen 6 family member G6C [Source:HGNC Symbol;Acc:HGNC:13936]

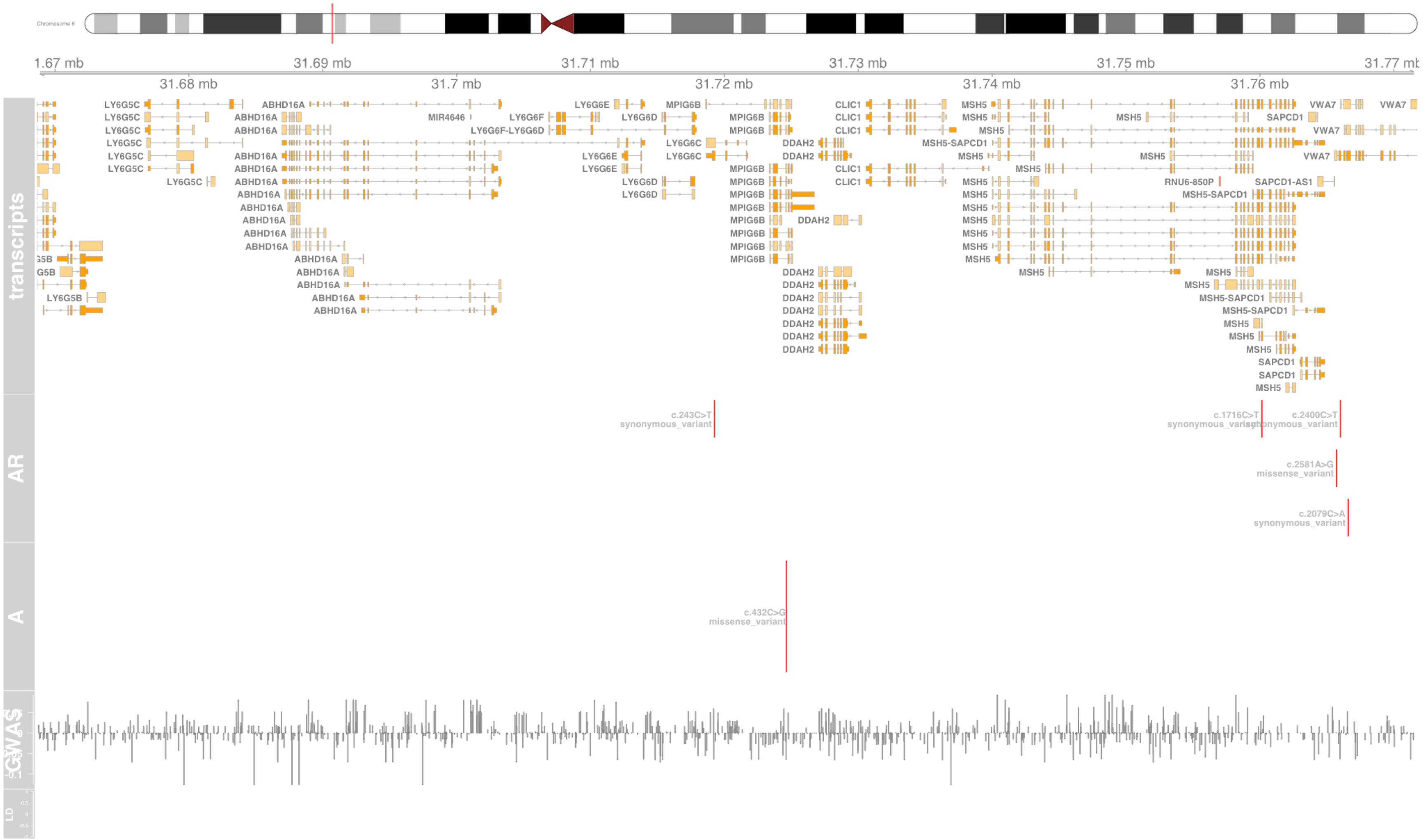

#### Fig S75 MCCD1

mitochondrial coiled-coil domain 1 [Source:HGNC Symbol;Acc:HGNC:20668]

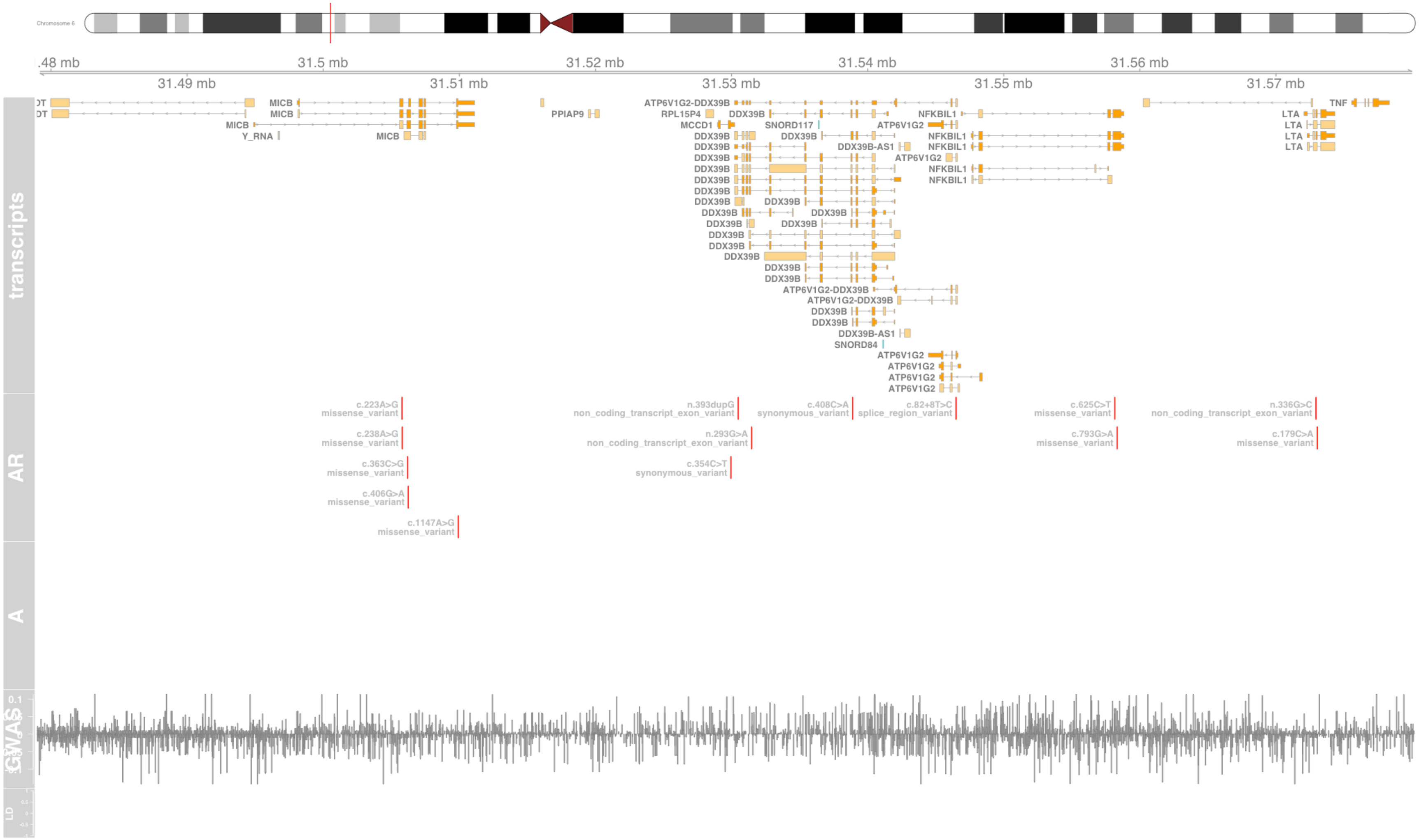

#### Fig S76 MED24

mediator complex subunit 24 [Source:HGNC Symbol;Acc:HGNC:22963]

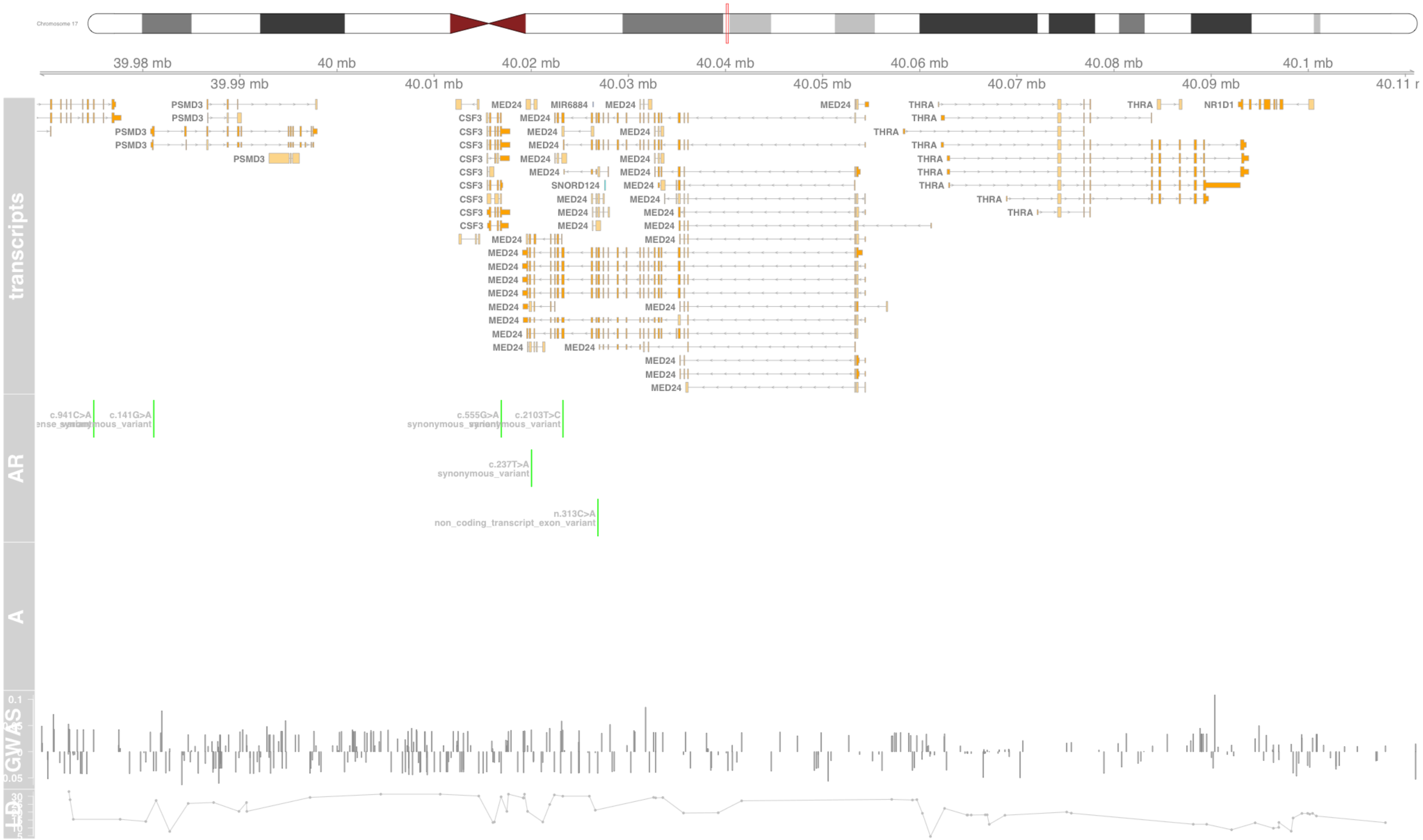

#### Fig S77 MEI1

meiotic double-stranded break formation protein 1 [Source:HGNC Symbol;Acc:HGNC:28613]

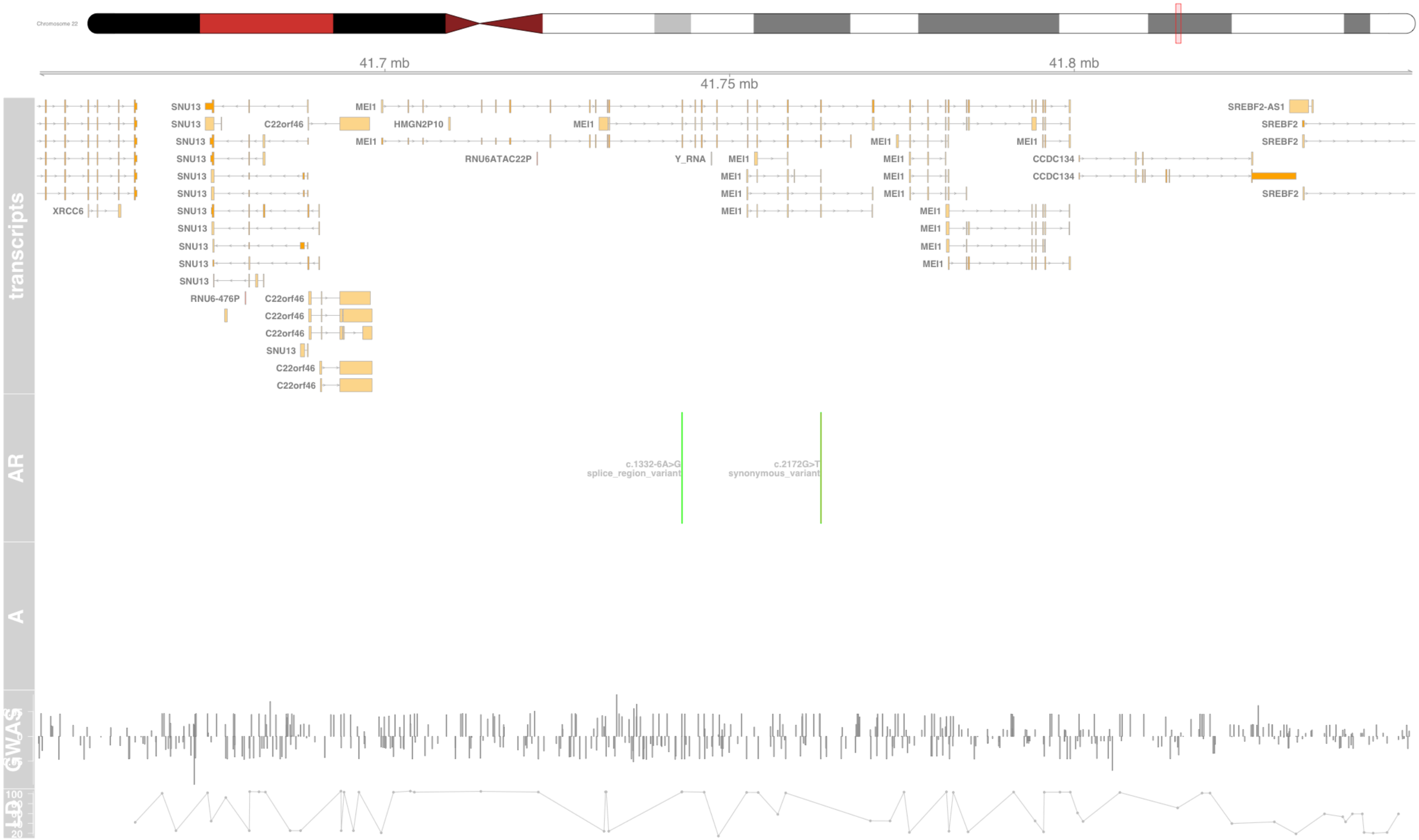

#### Fig S78 MICA

MHC class I polypeptide-related sequence A [Source:HGNC Symbol;Acc:HGNC:7090]

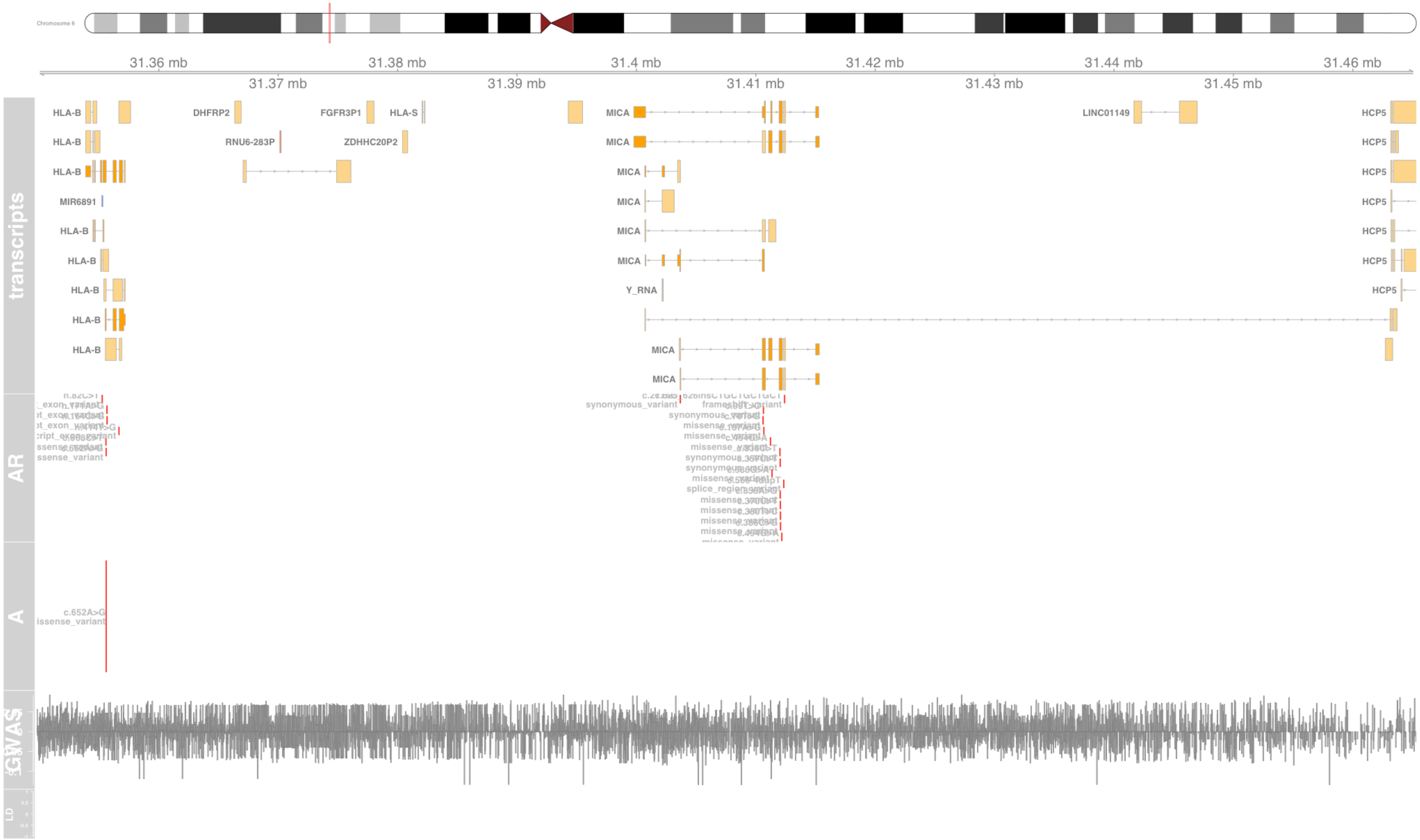

### Fig S79 MICB

MHC class I polypeptide-related sequence B [Source:HGNC Symbol;Acc:HGNC:7091]

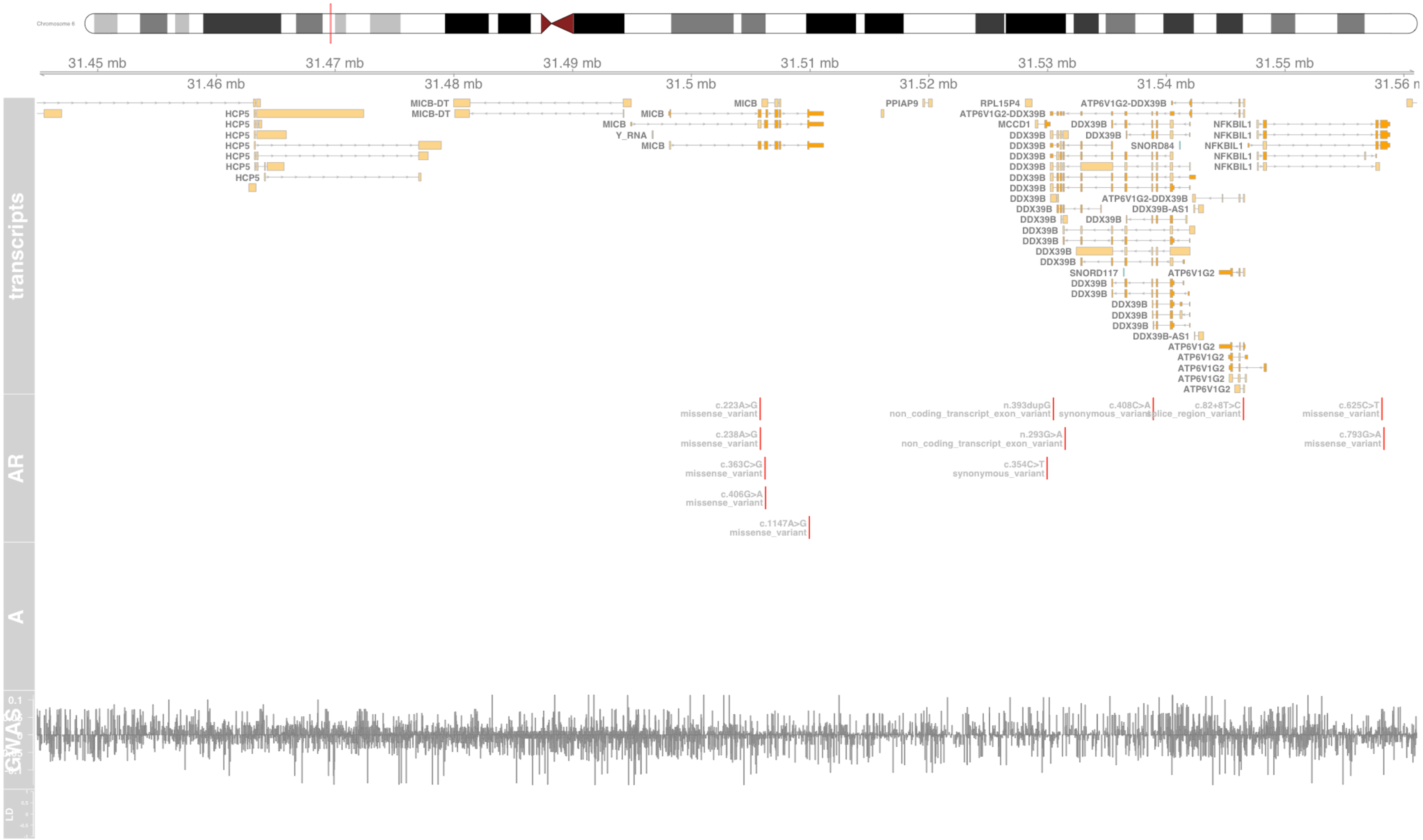

#### Fig S80 MIR6891

microRNA 6891 [Source:HGNC Symbol;Acc:HGNC:50243]

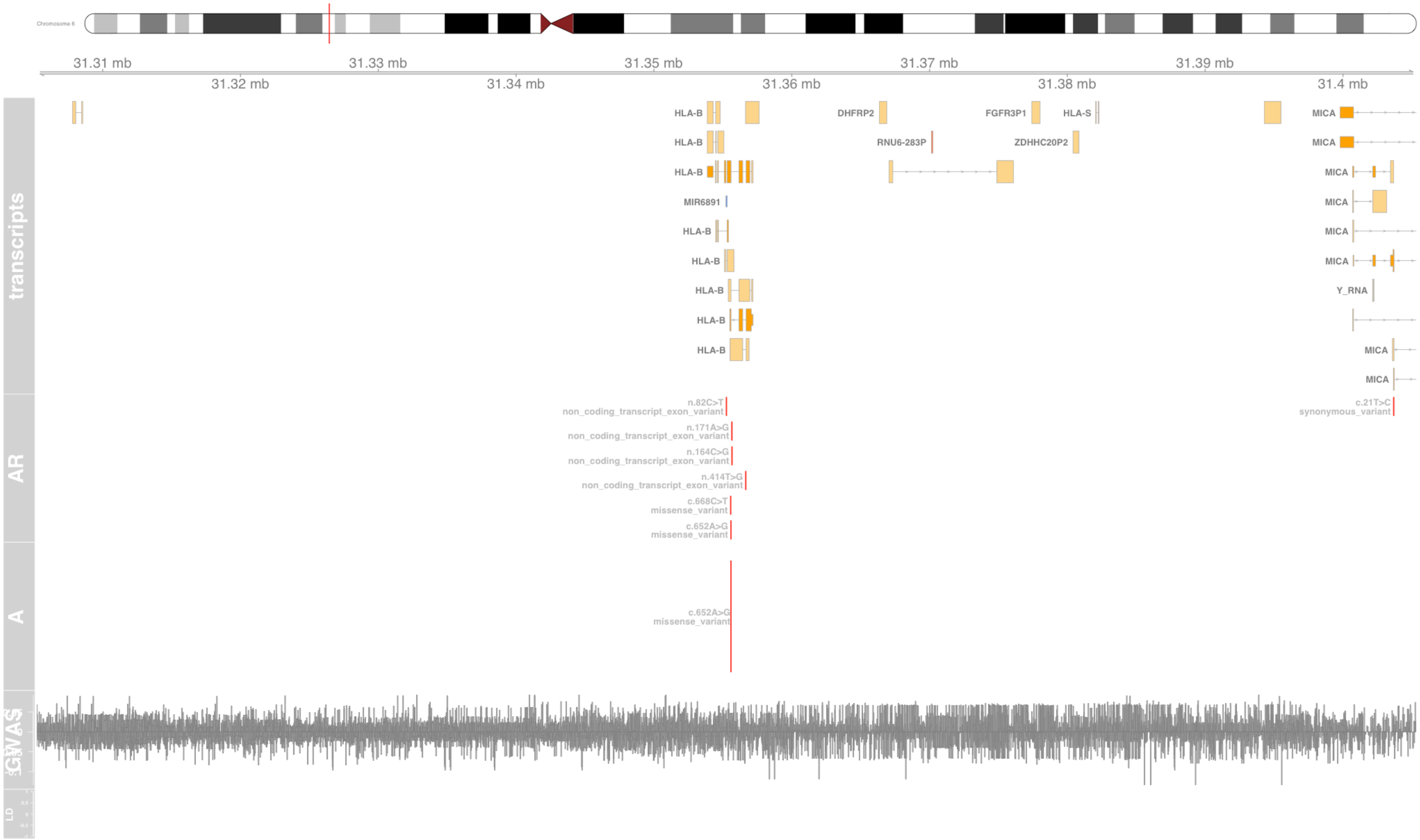

#### Fig S81 MPIG6B

megakaryocyte and platelet inhibitory receptor G6b [Source:HGNC Symbol;Acc:HGNC:13937]

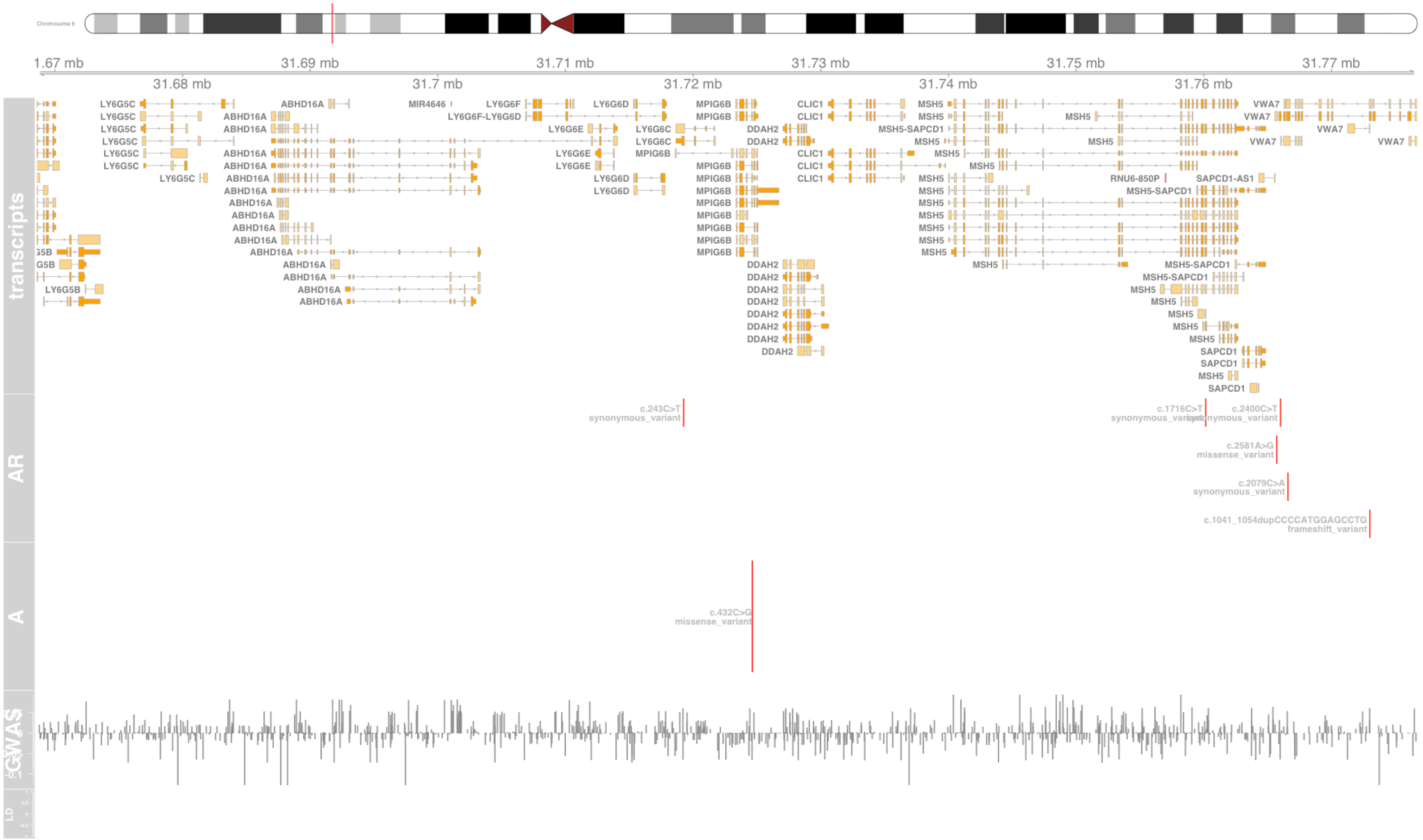

### Fig S82 MSH5

mutS homolog 5 [Source:HGNC Symbol;Acc:HGNC:7328]

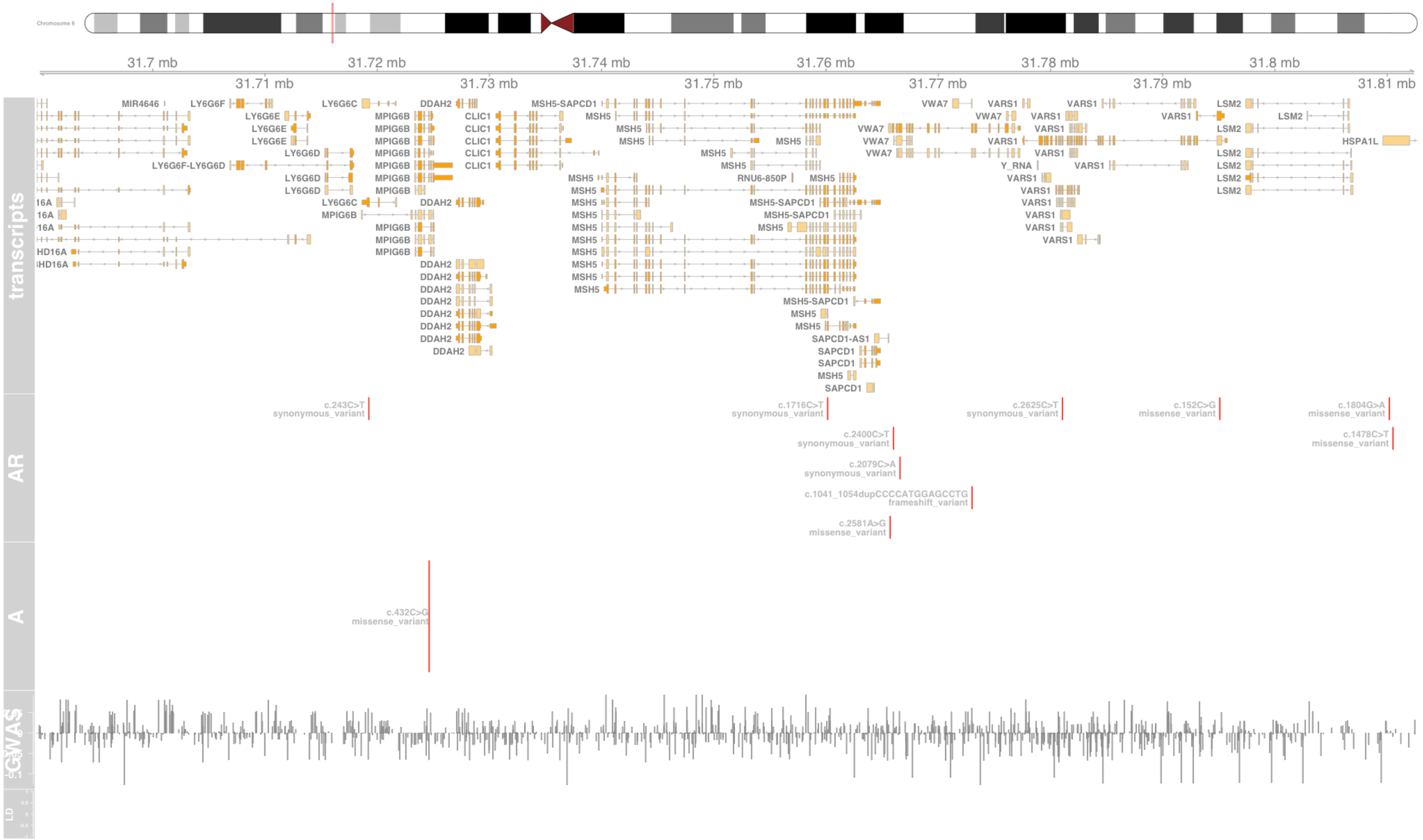

#### Fig S83 MUC22

mucin 22 [Source:HGNC Symbol;Acc:HGNC:39755]

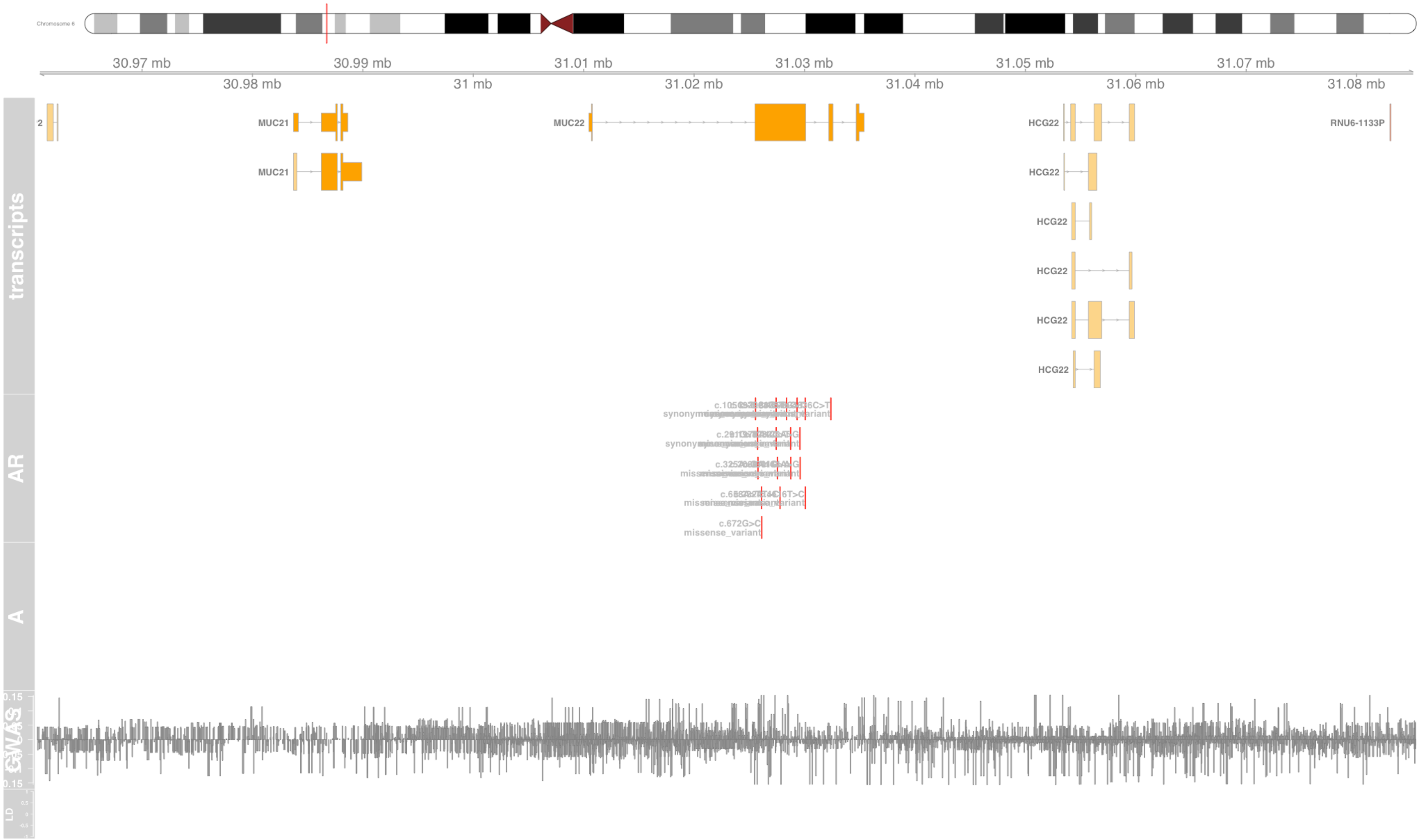

#### Fig S84 MYRF

myelin regulatory factor [Source:HGNC Symbol;Acc:HGNC:1181]

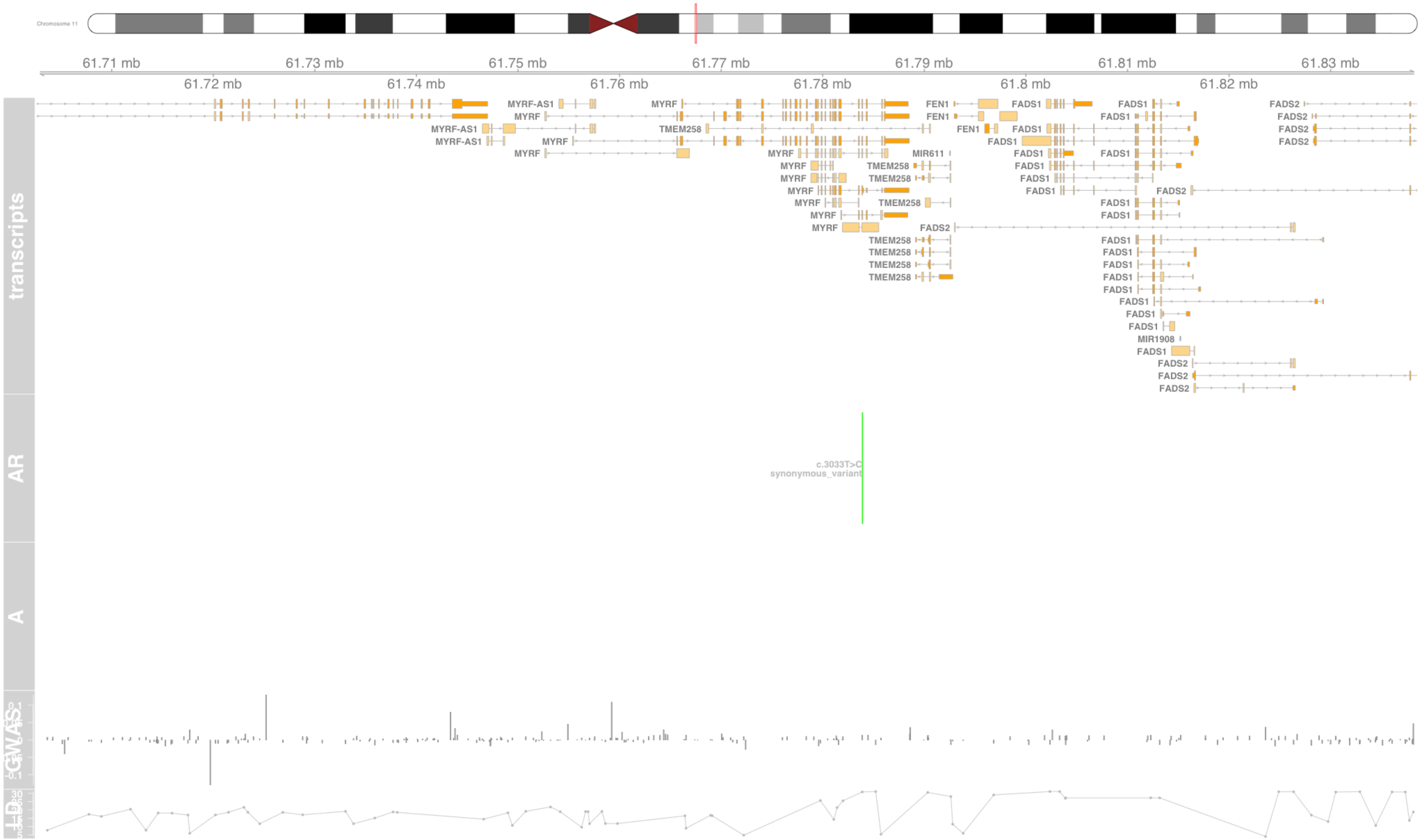

#### Fig S85 NEU1

neuraminidase 1 [Source:HGNC Symbol;Acc:HGNC:7758]

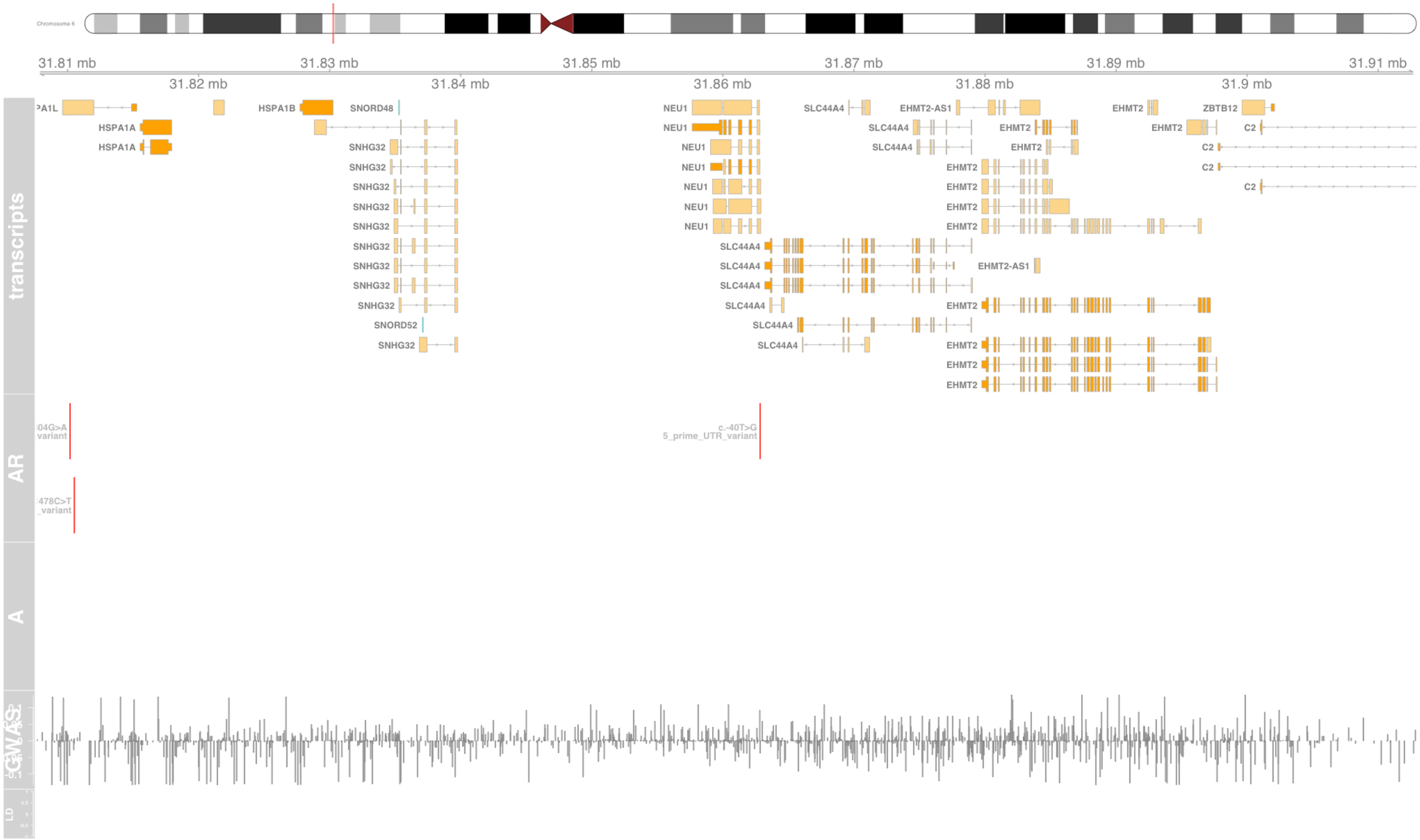

#### Fig S86 NFKBIL1

NFKB inhibitor like 1 [Source:HGNC Symbol;Acc:HGNC:7800]

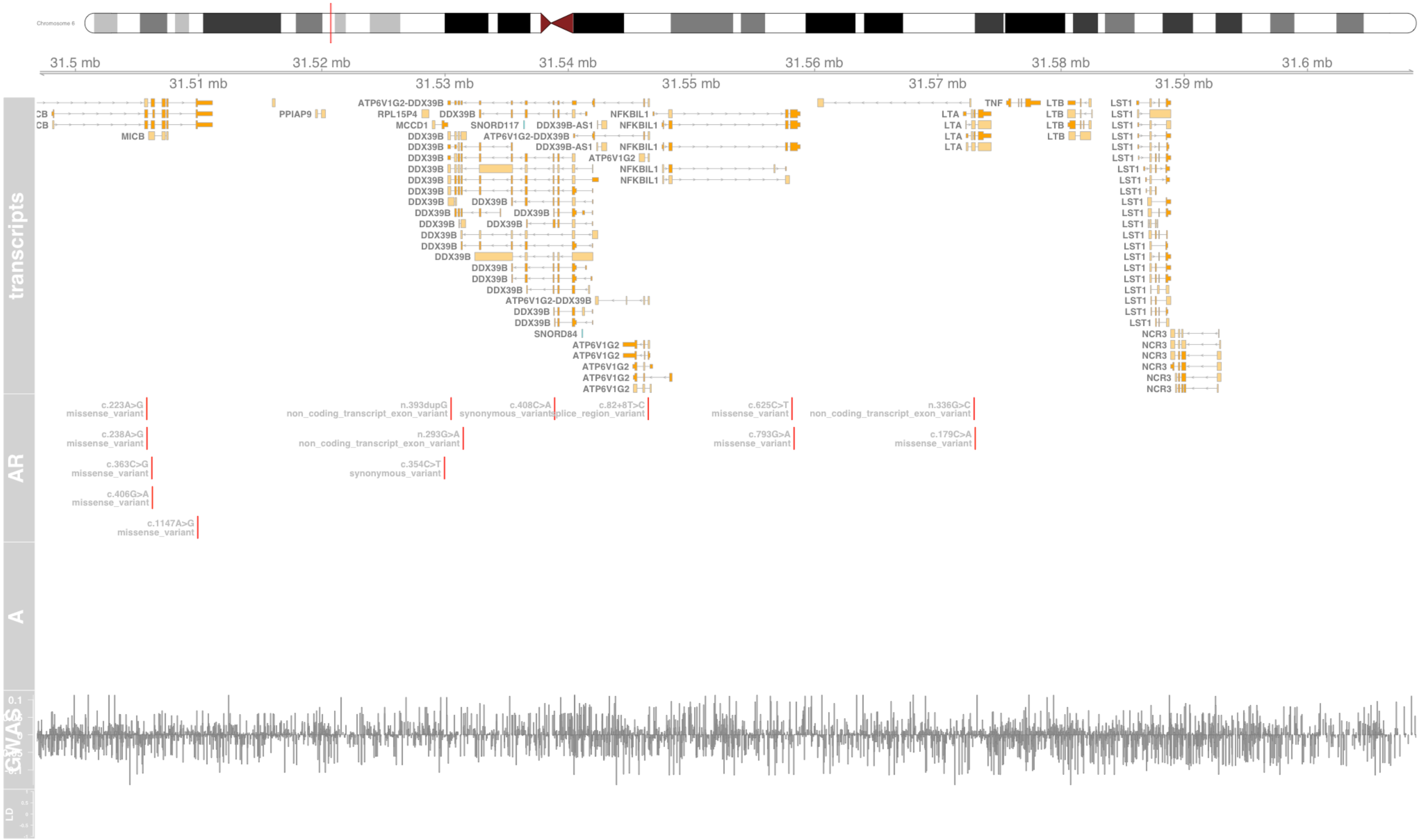

### Fig S87 NOTCH4

notch receptor 4 [Source:HGNC Symbol;Acc:HGNC:7884]

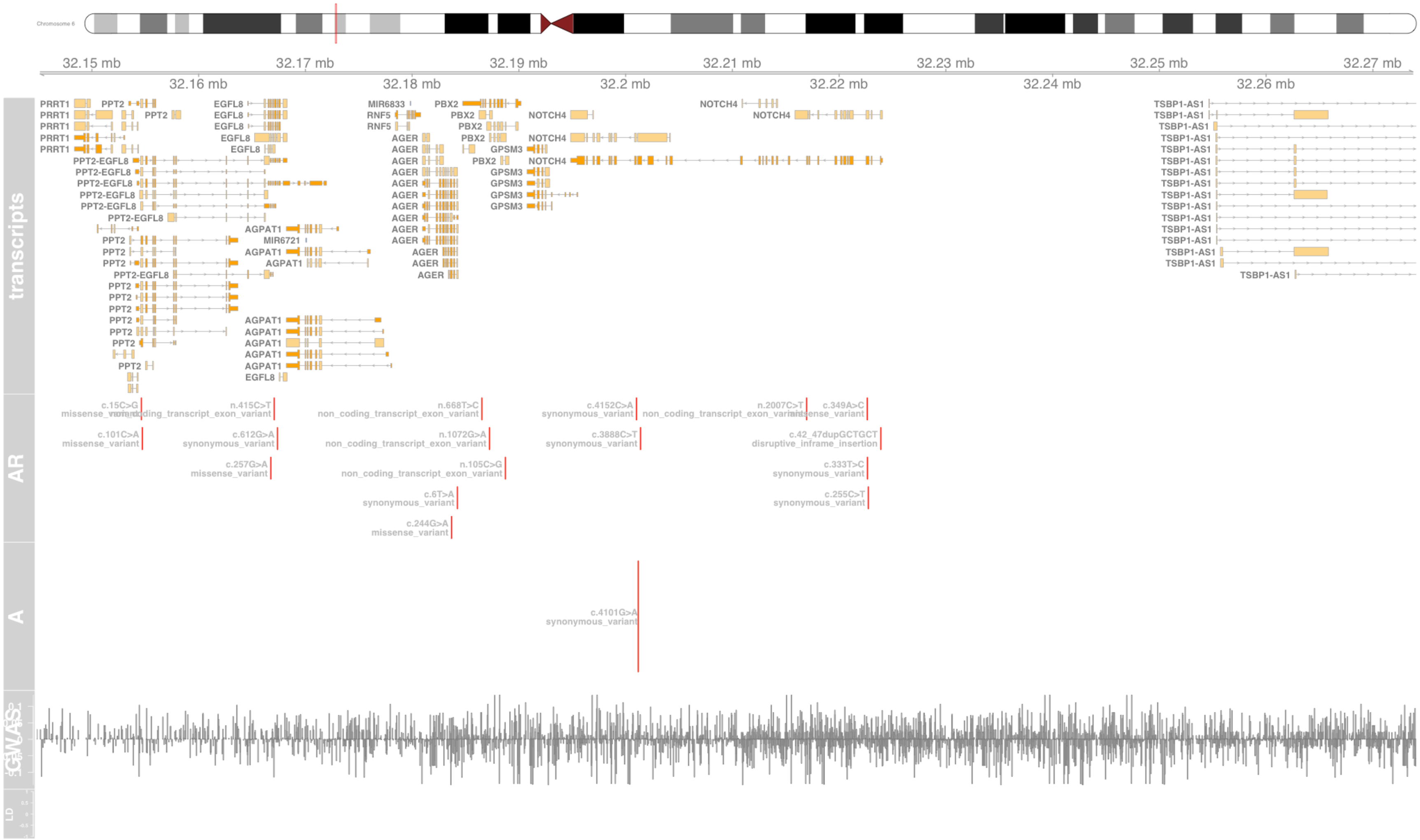

#### Fig S88 NSMCE1

NSE1 homolog, SMC5-SMC6 complex component [Source:HGNC Symbol;Acc:HGNC:29897]

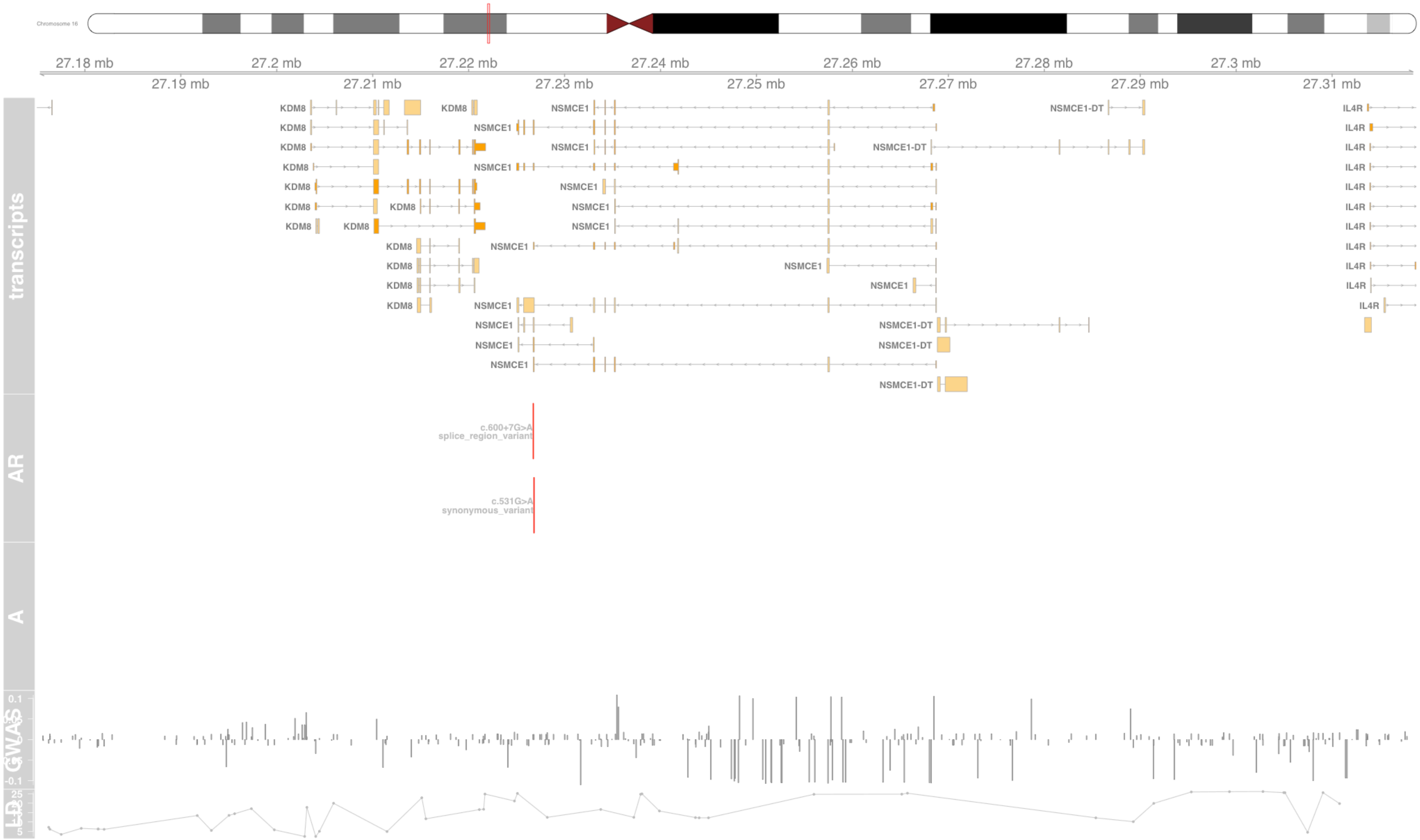

#### Fig S89 OR12D3

olfactory receptor family 12 subfamily D member 3 [Source:HGNC Symbol;Acc:HGNC:13963]

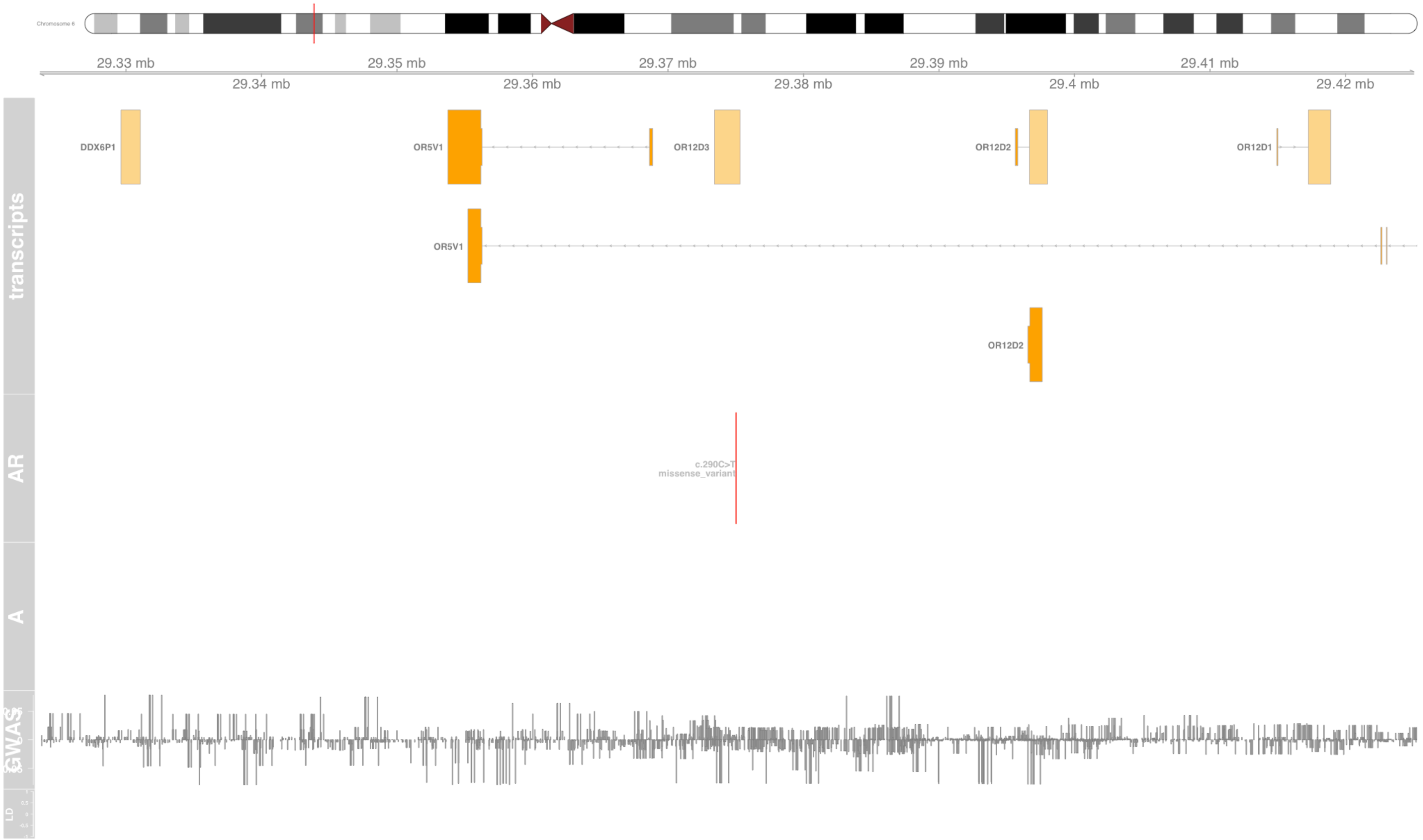

#### Fig S90 OR2B2

olfactory receptor family 2 subfamily B member 2 [Source:HGNC Symbol;Acc:HGNC:13966]

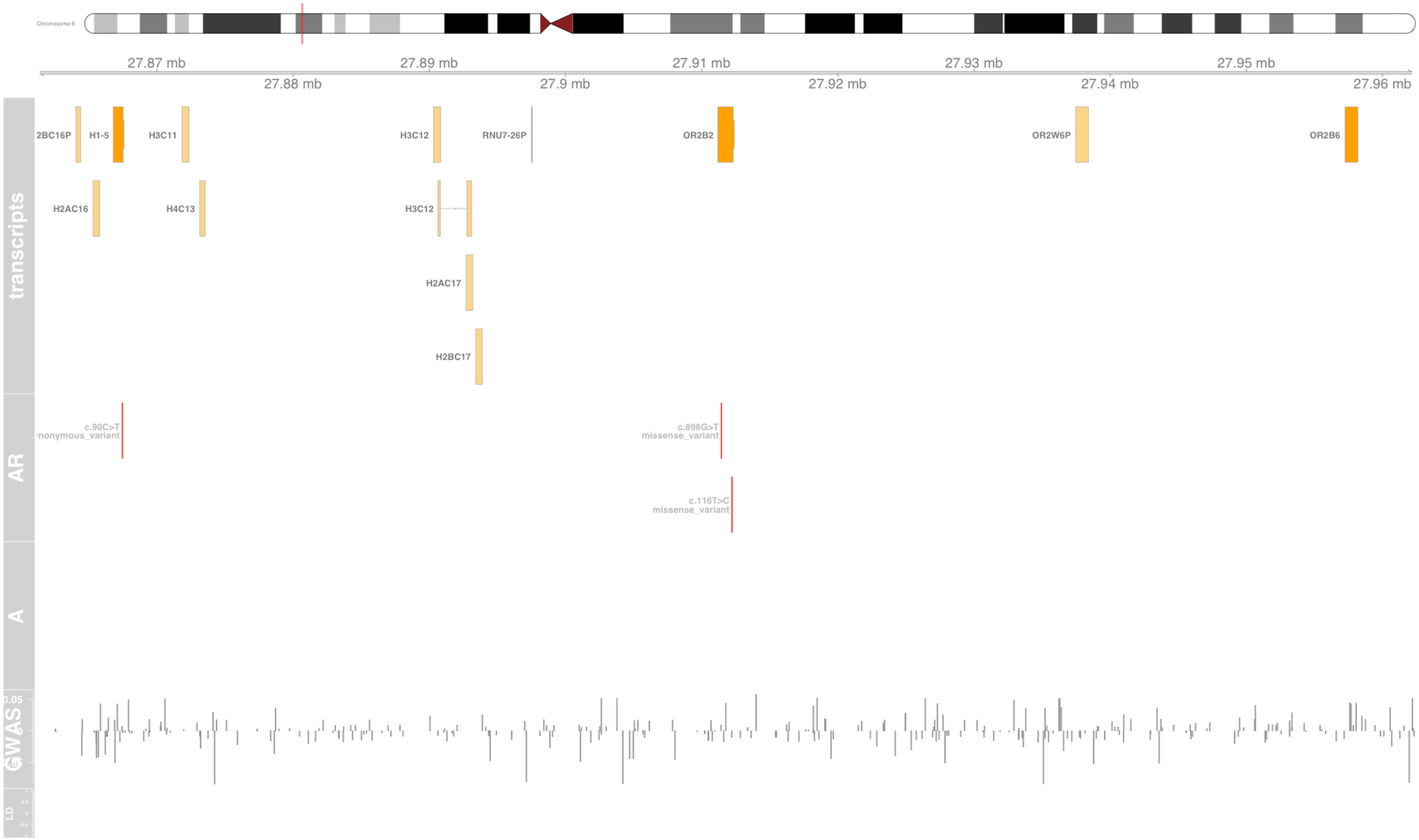

#### Fig S91 PBX2

PBX homeobox 2 [Source:HGNC Symbol;Acc:HGNC:8633]

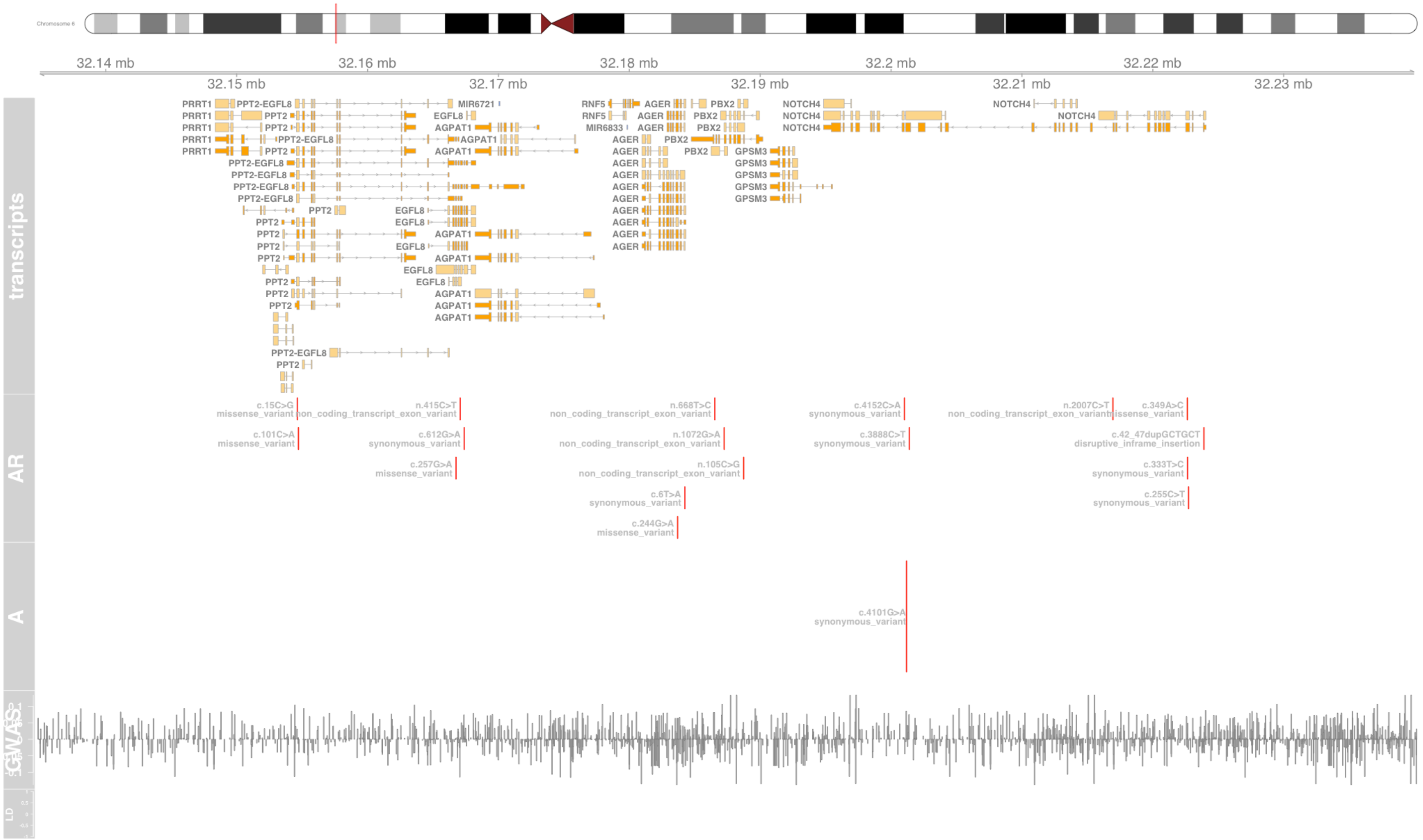

#### Fig S92 PDLIM4

PDZ and LIM domain 4 [Source:HGNC Symbol;Acc:HGNC:16501]

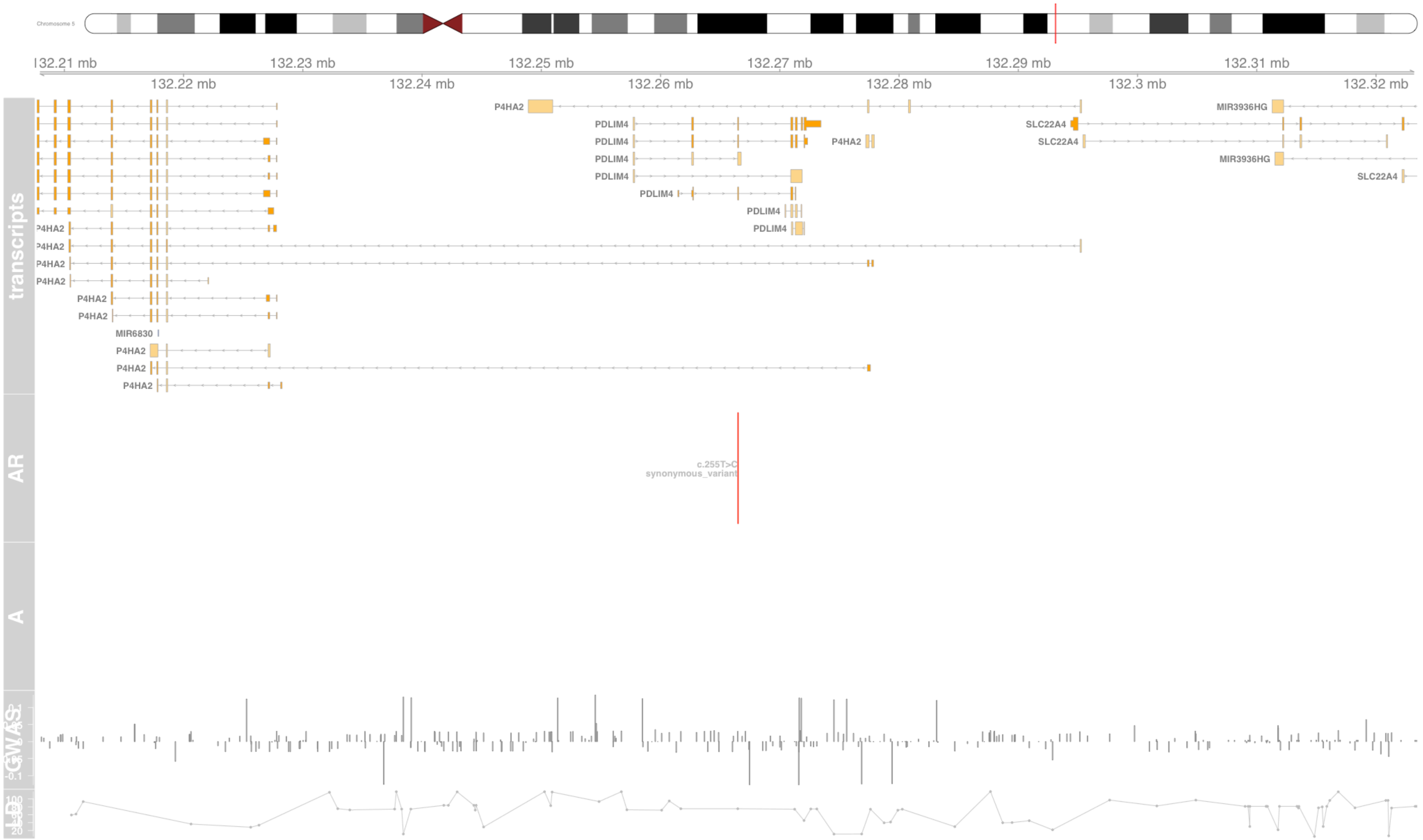

#### Fig S93 PGAP3

post-GPI attachment to proteins phospholipase 3 [Source:HGNC Symbol;Acc:HGNC:23719]

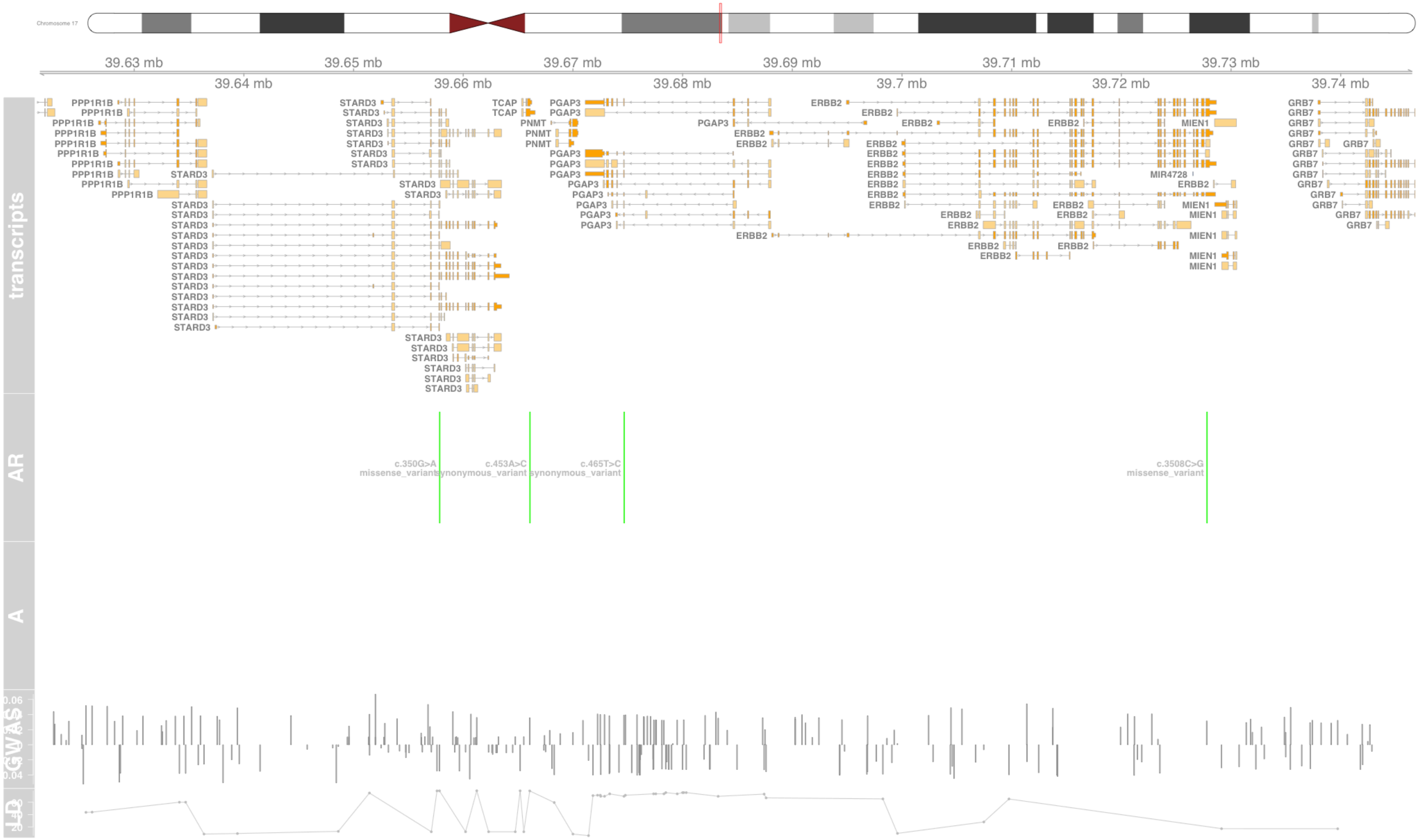

#### Fig S94 PGBD1

piggyBac transposable element derived 1 [Source:HGNC Symbol;Acc:HGNC:19398]

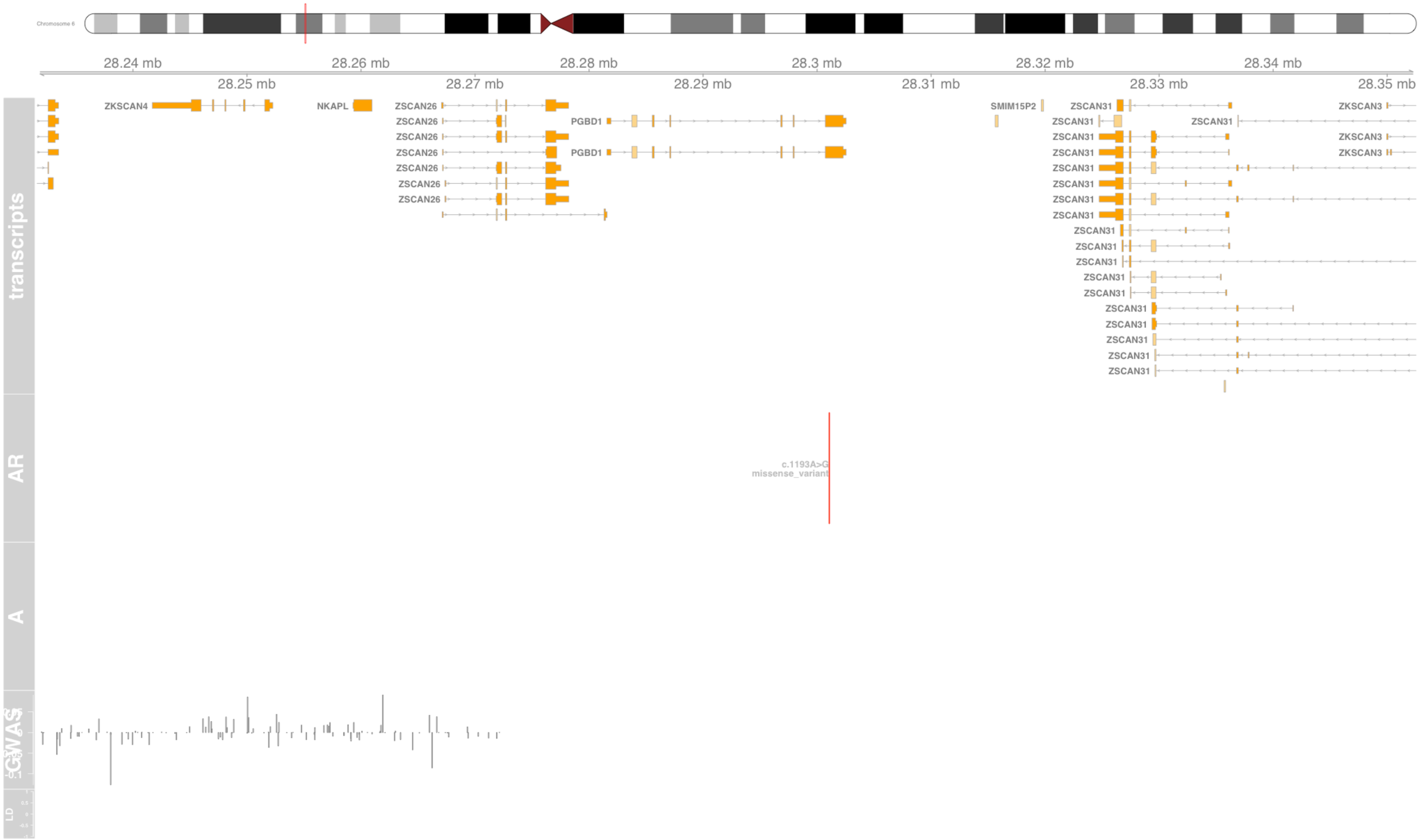

#### Fig S95 PHB

prohibitin [Source:HGNC Symbol;Acc:HGNC:8912]

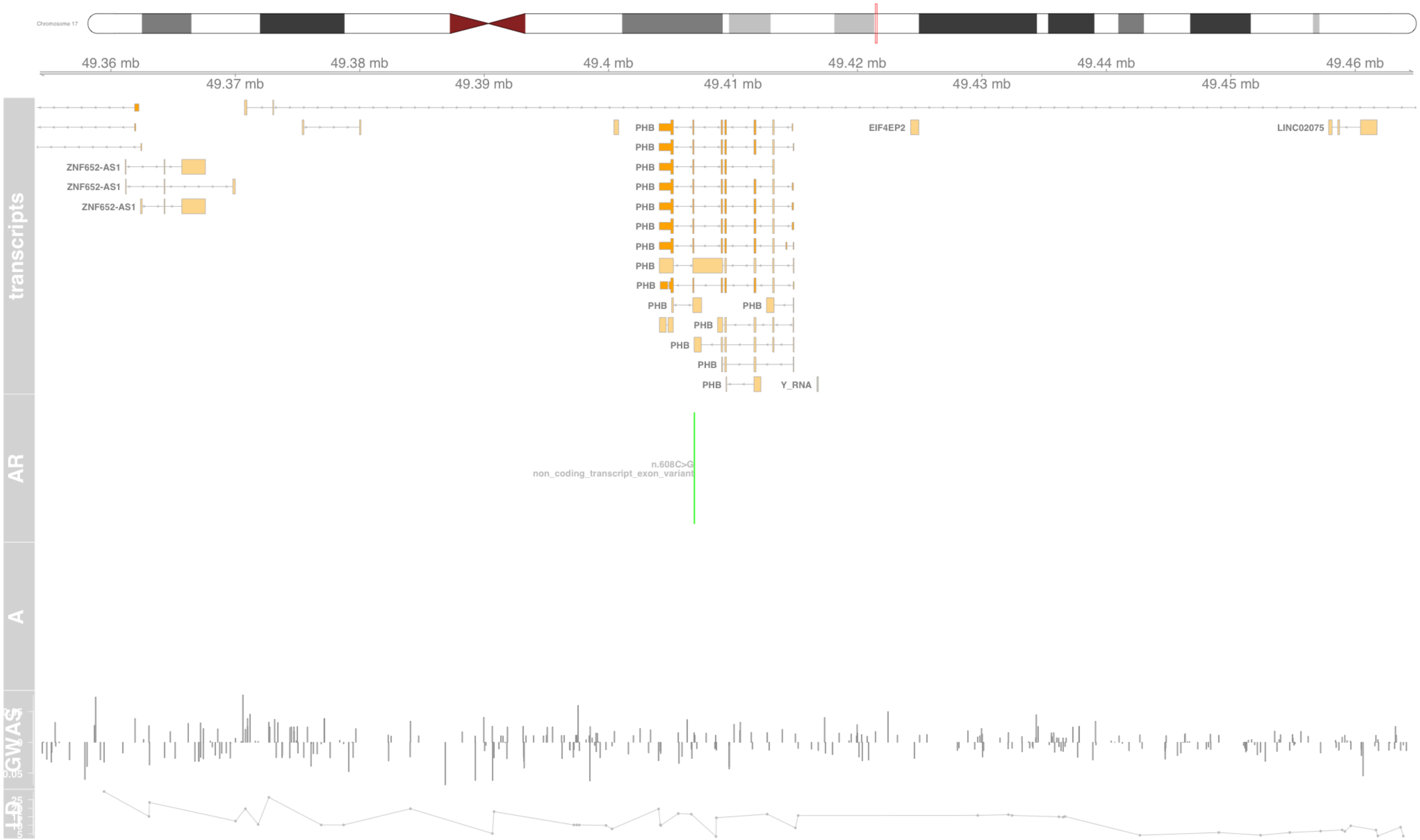

### Fig S96 POU5F1

POU class 5 homeobox 1 [Source:HGNC Symbol;Acc:HGNC:9221]

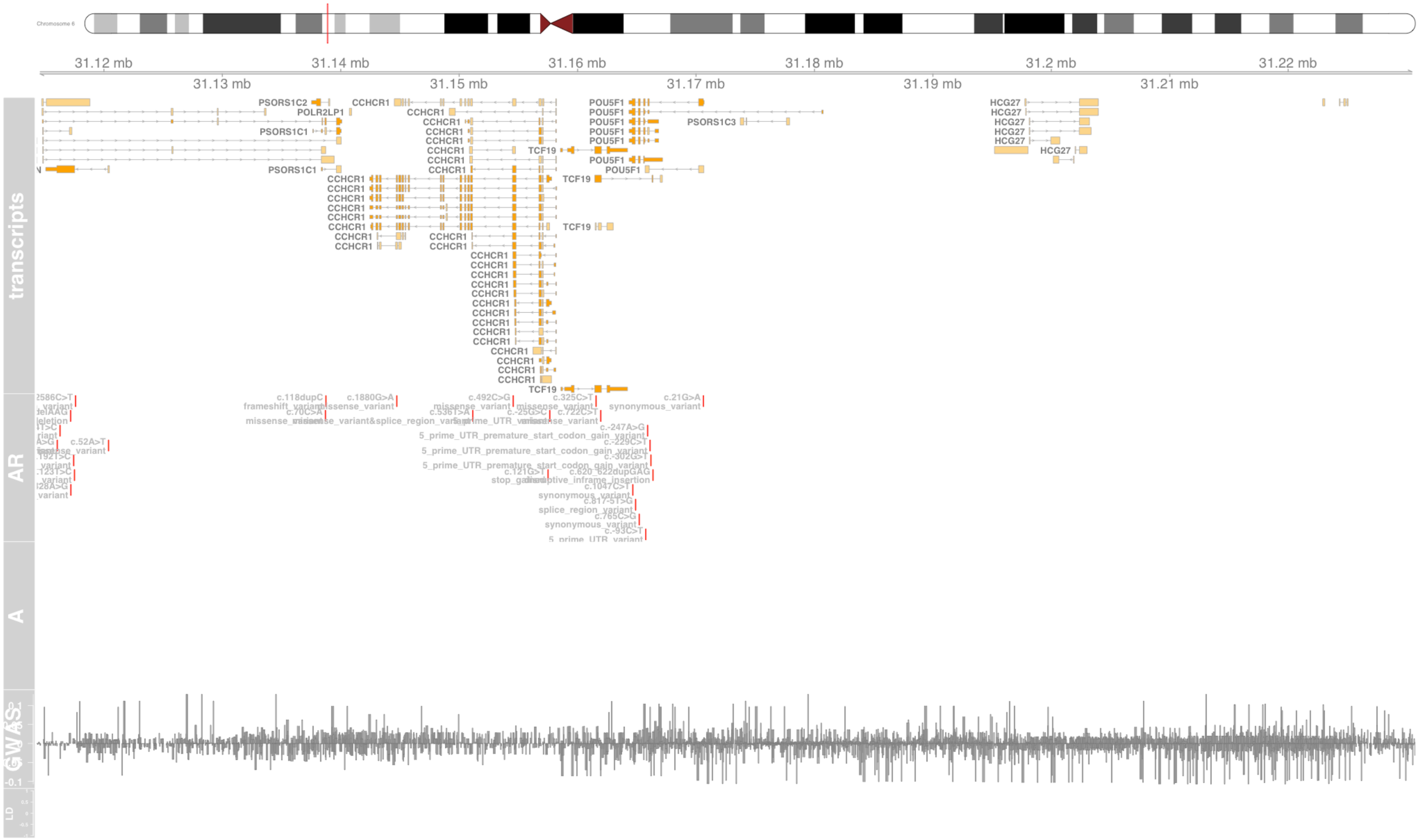

#### Fig S97 PPP1R18

protein phosphatase 1 regulatory subunit 18 [Source:HGNC Symbol;Acc:HGNC:29413]

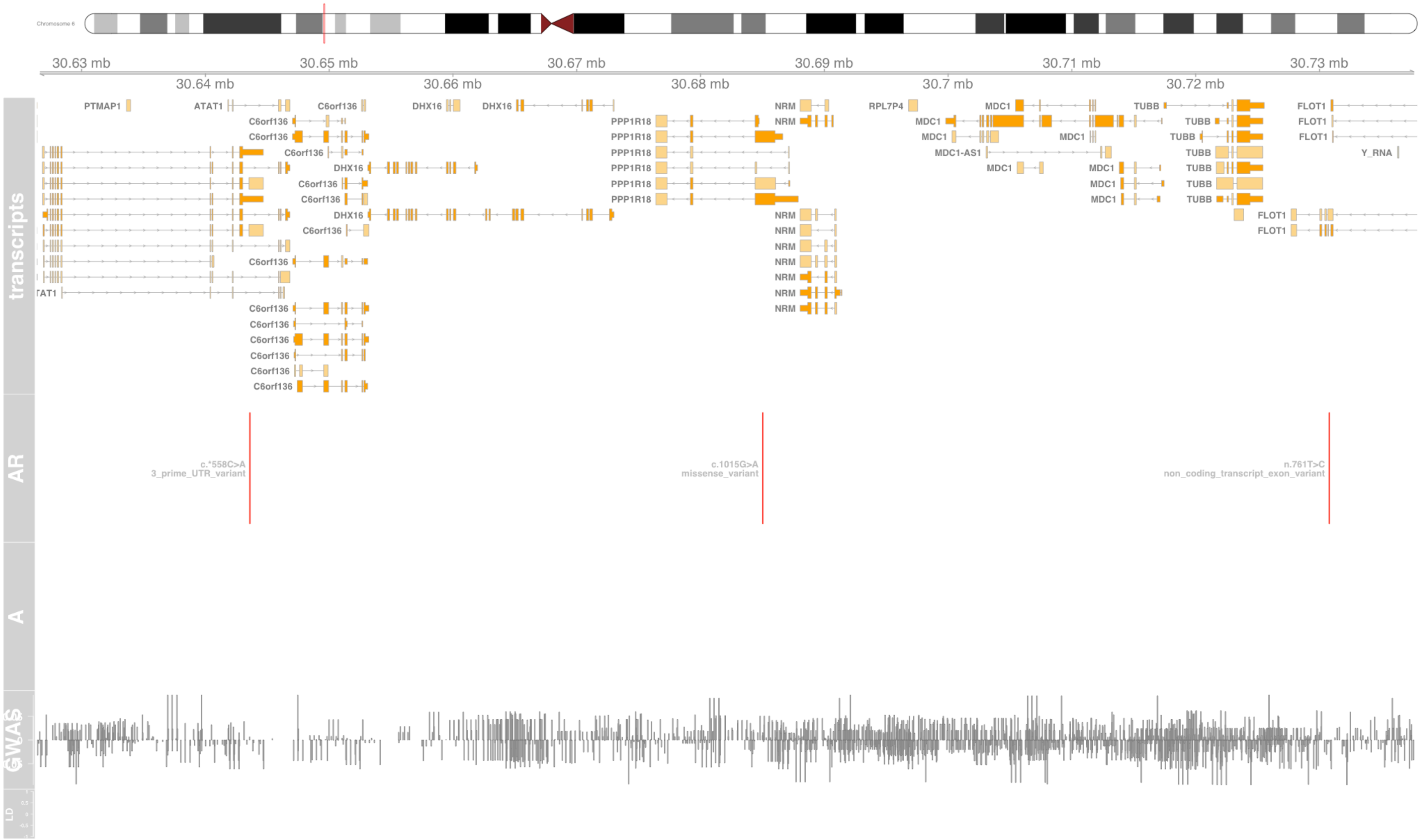

#### Fig S98 PPT2

palmitoyl-protein thioesterase 2 [Source:HGNC Symbol;Acc:HGNC:9326]

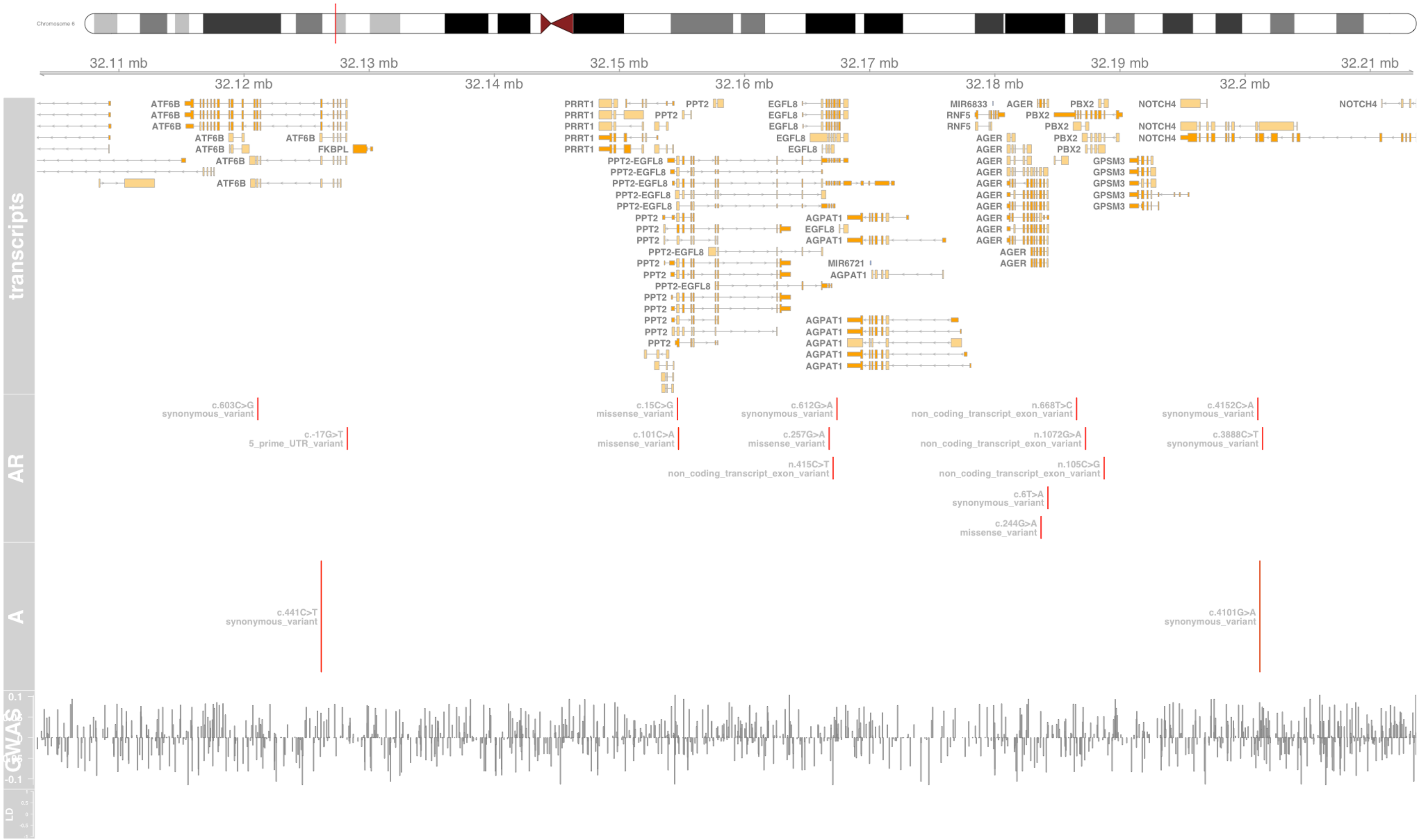

#### Fig S99 PRRC2A

proline rich coiled-coil 2A [Source:HGNC Symbol;Acc:HGNC:13918]

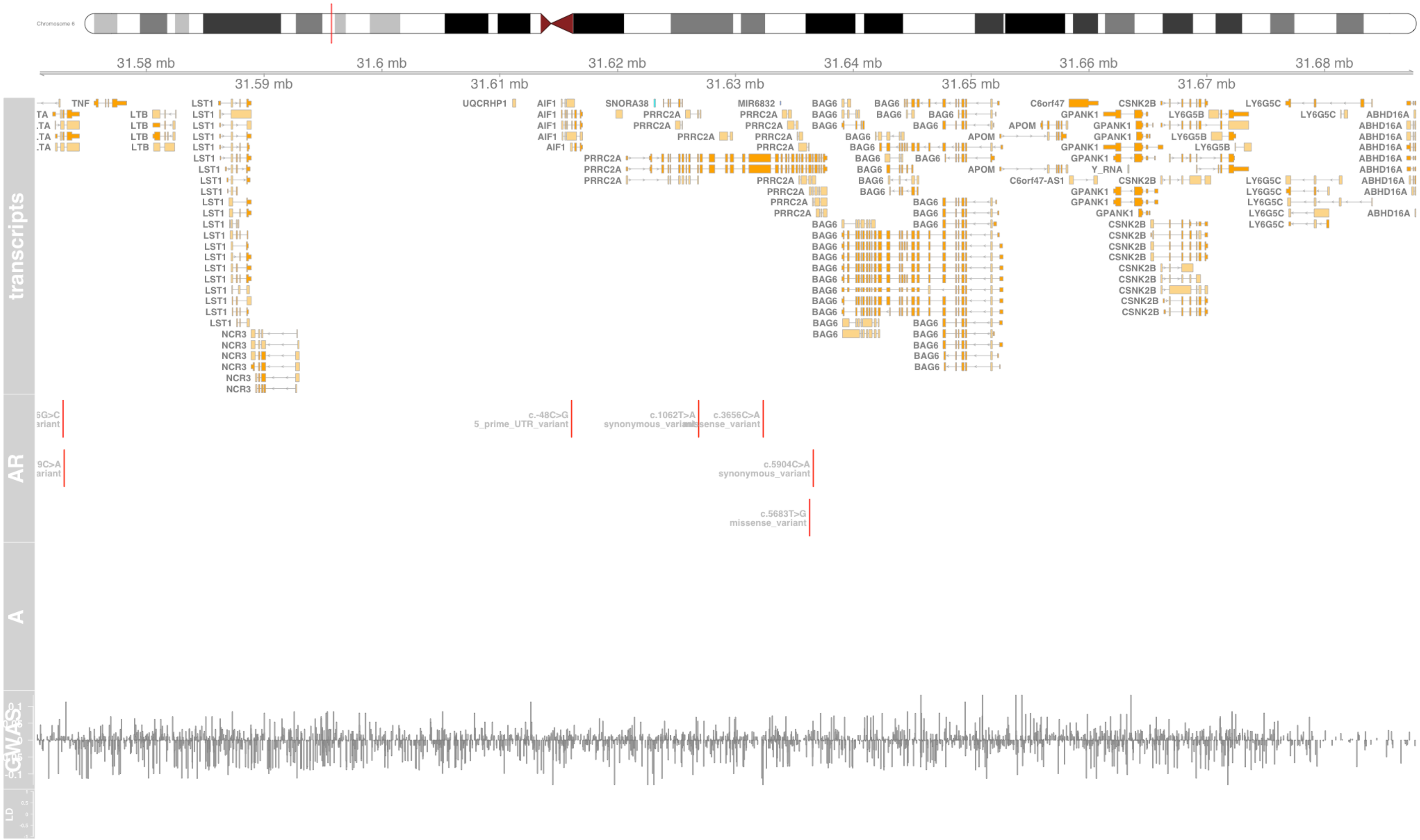

#### Fig S100 PSMB8

proteasome 20S subunit beta 8 [Source:HGNC Symbol;Acc:HGNC:9545]

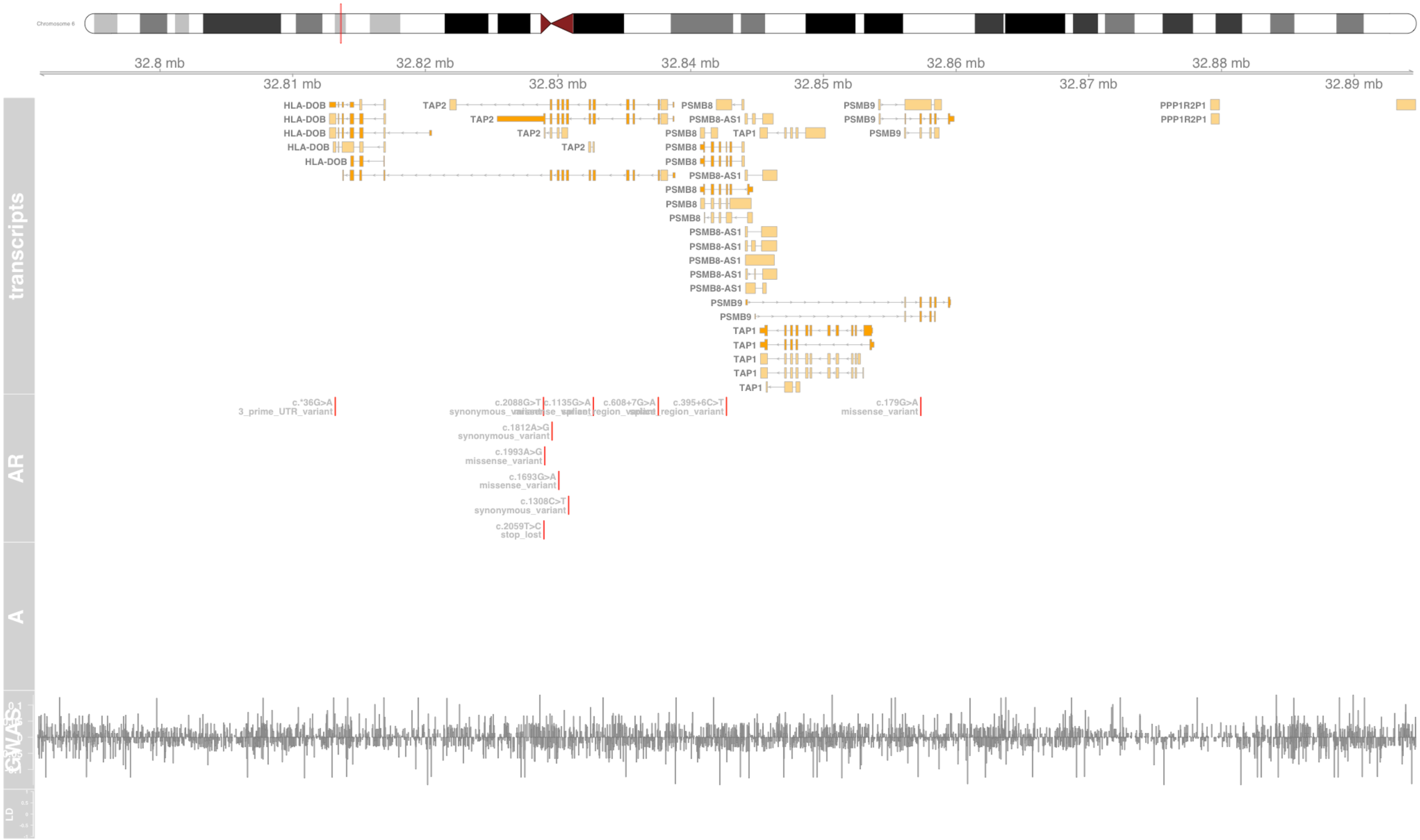

#### Fig S101 PSMB9

proteasome 20S subunit beta 9 [Source:HGNC Symbol;Acc:HGNC:9546]

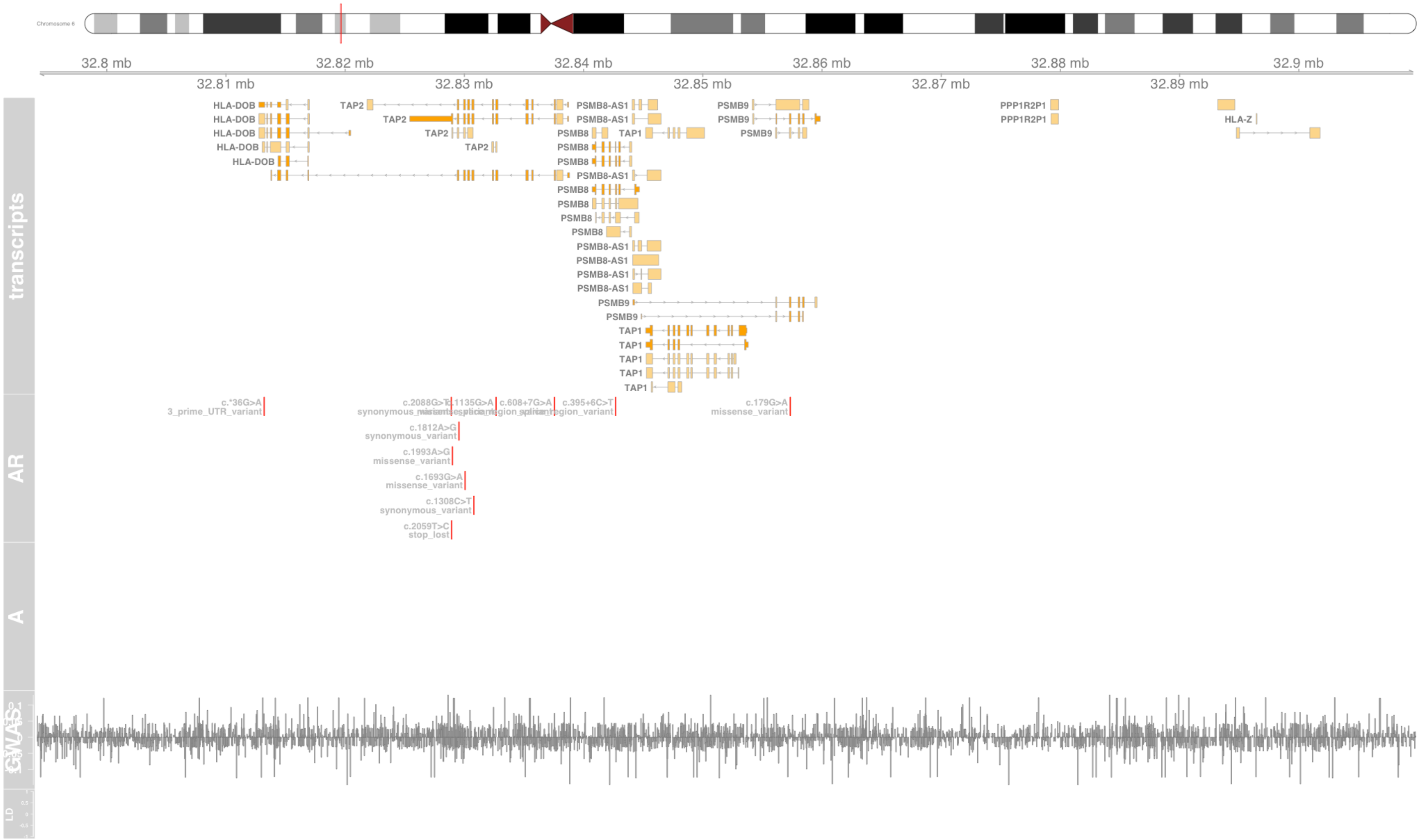

#### Fig S102 PSMD3

proteasome 26S subunit, non-ATPase 3 [Source:HGNC Symbol;Acc:HGNC:9560]

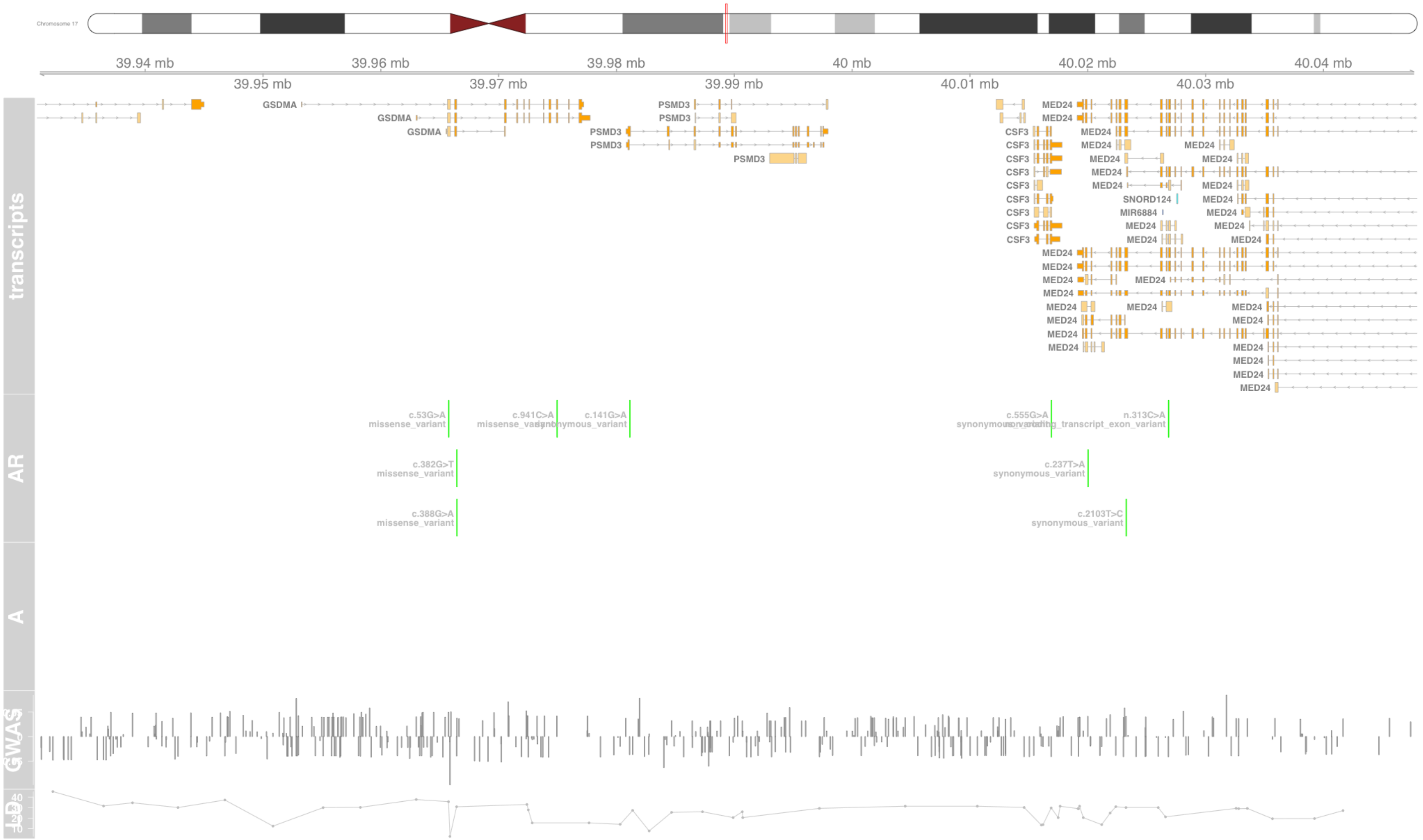

#### Fig S103 PSORS1C1

psoriasis susceptibility 1 candidate 1 [Source:HGNC Symbol;Acc:HGNC:17202]

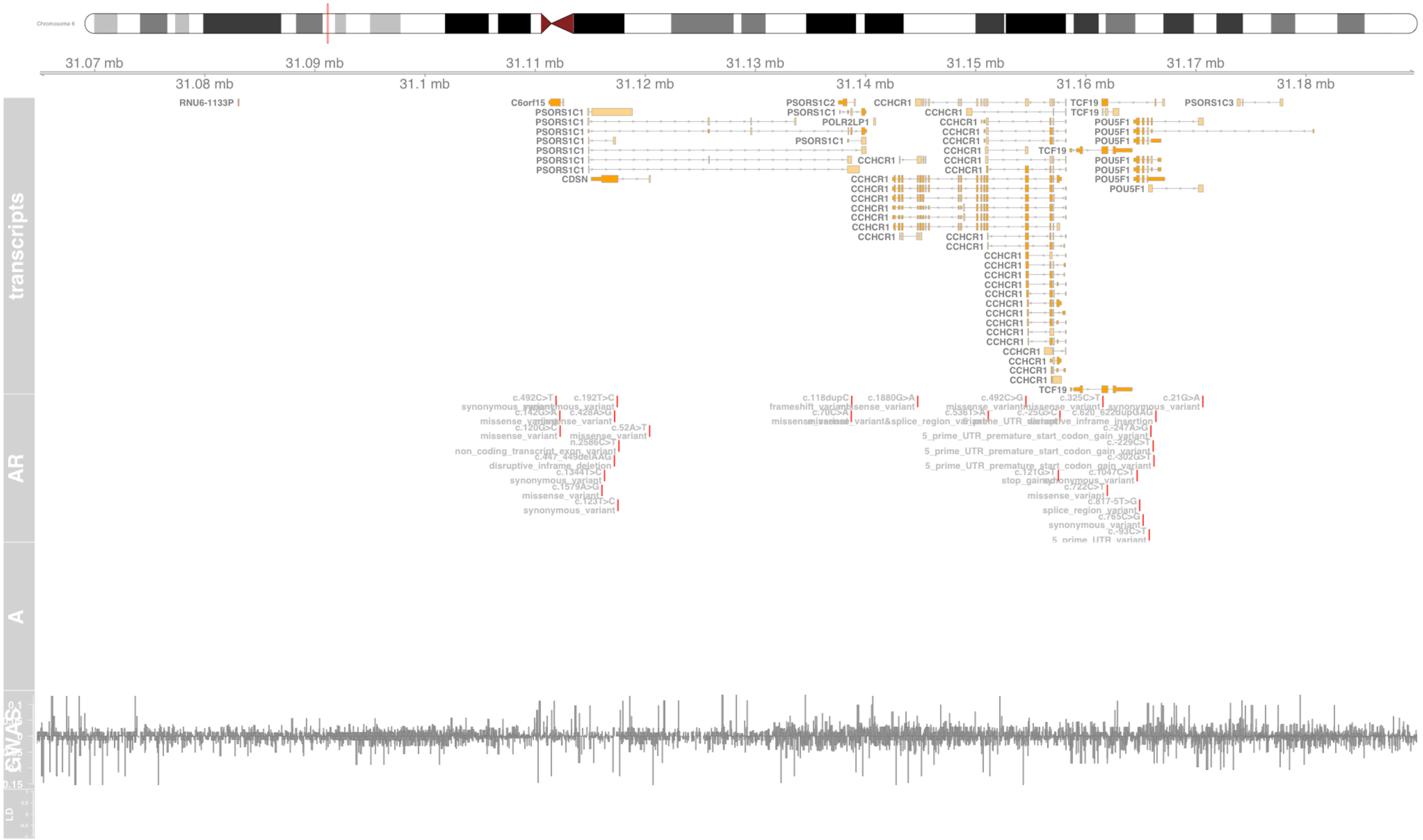

### Fig S104 RPS26

ribosomal protein S26 [Source:HGNC Symbol;Acc:HGNC:10414]

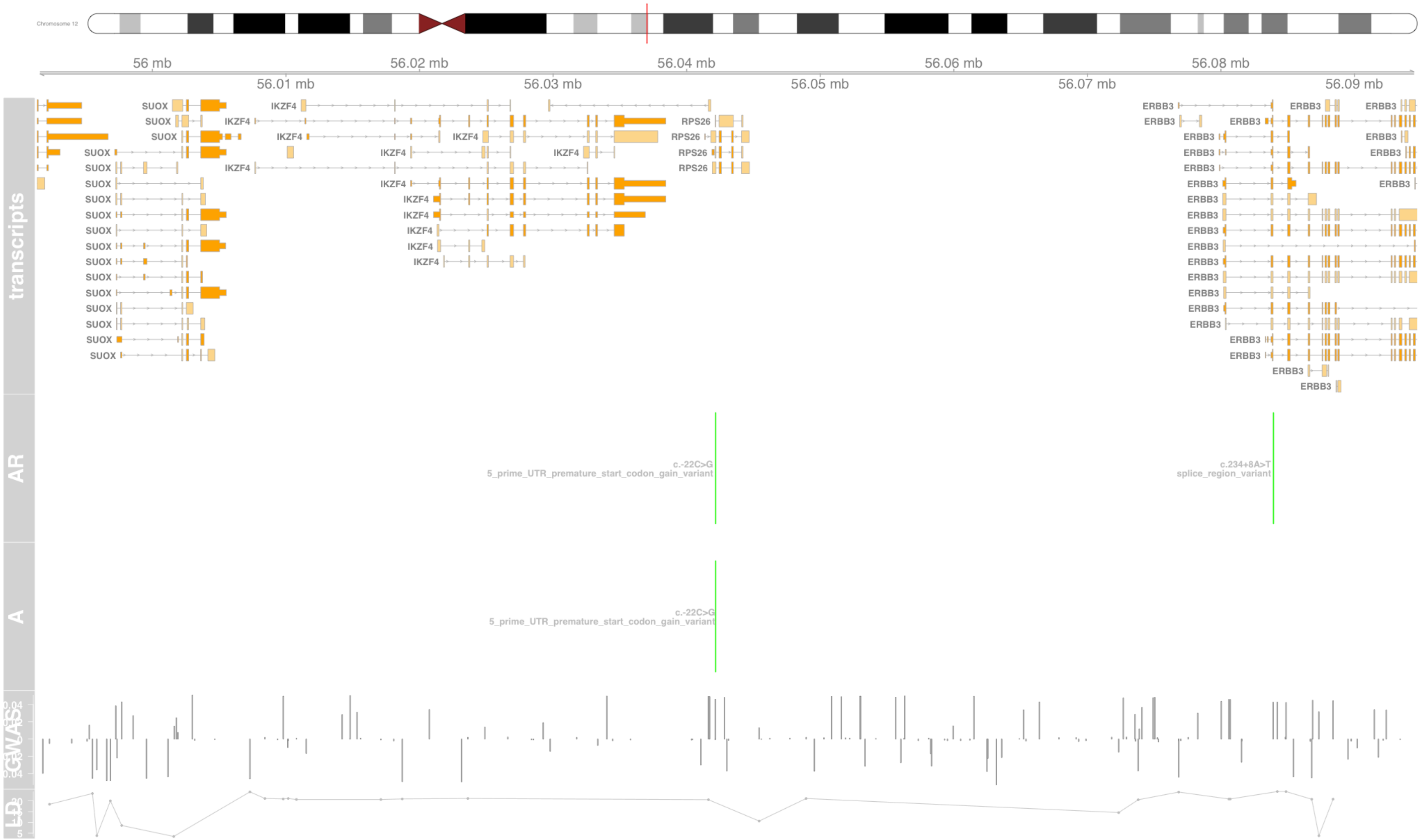

#### Fig S105 SKIV2L

Ski2 like RNA helicase [Source:HGNC Symbol;Acc:HGNC:10898]

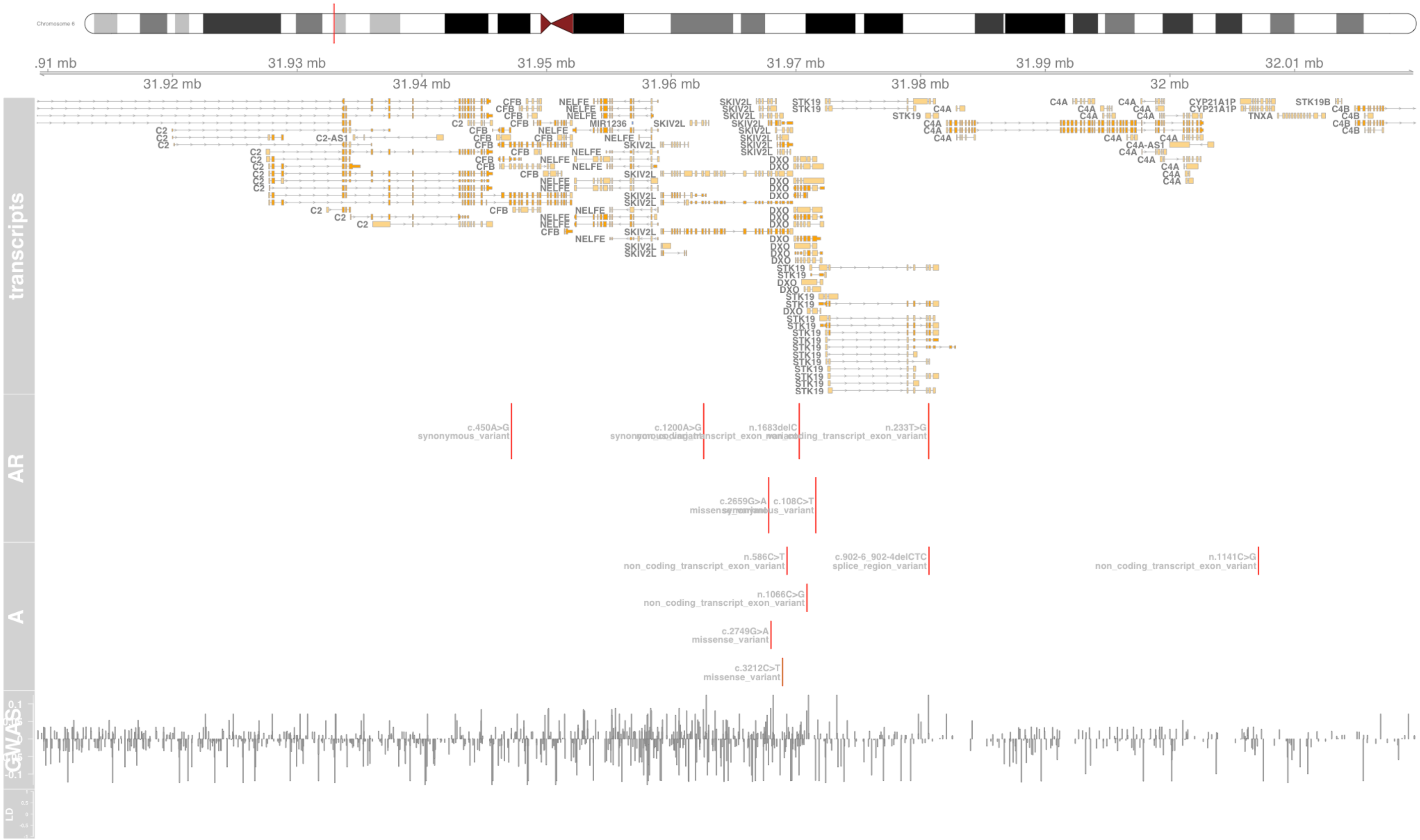

### Fig S106 SLC22A4

solute carrier family 22 member 4 [Source:HGNC Symbol;Acc:HGNC:10968]

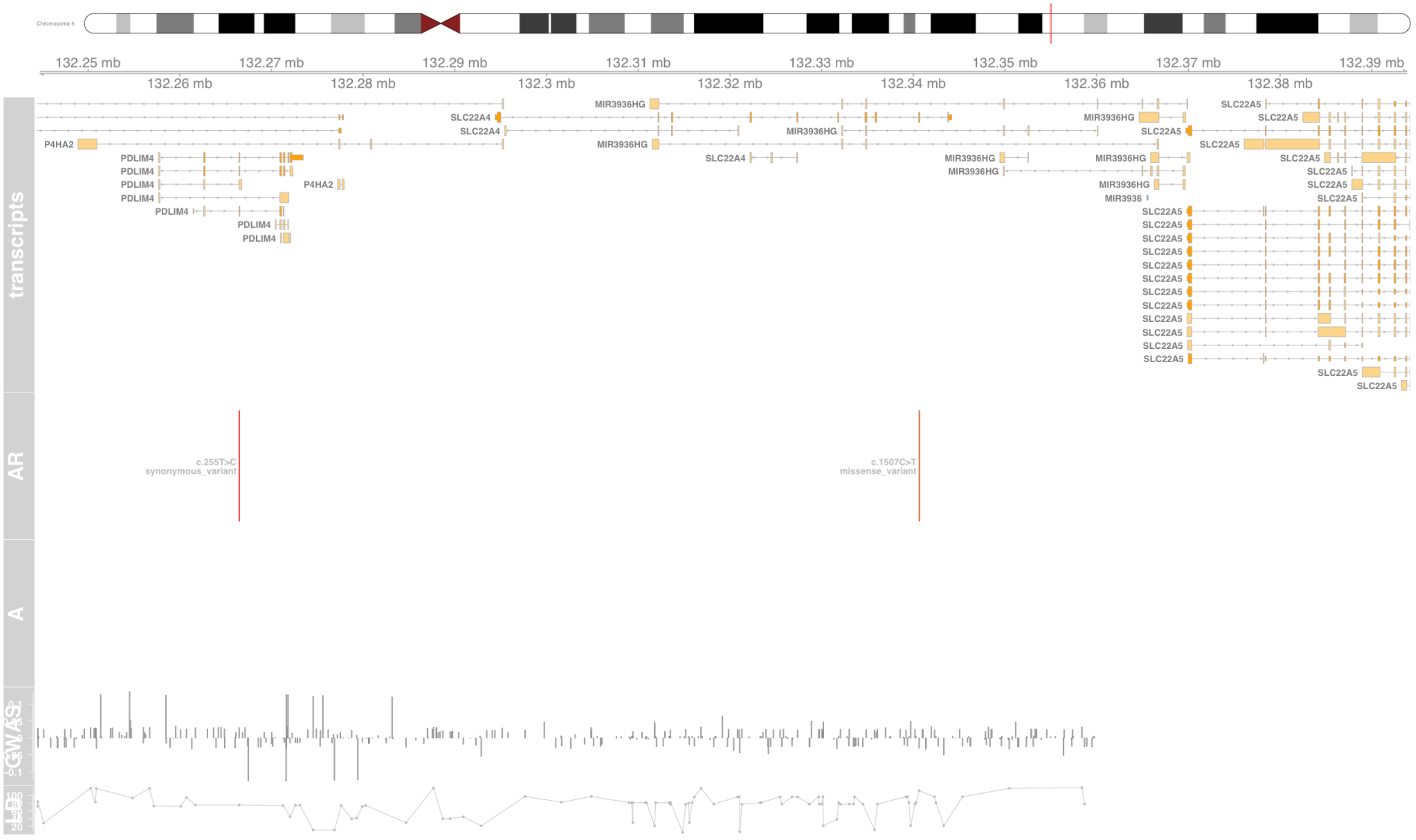

#### Fig S107 STARD3

StAR related lipid transfer domain containing 3 [Source:HGNC Symbol;Acc:HGNC:17579]

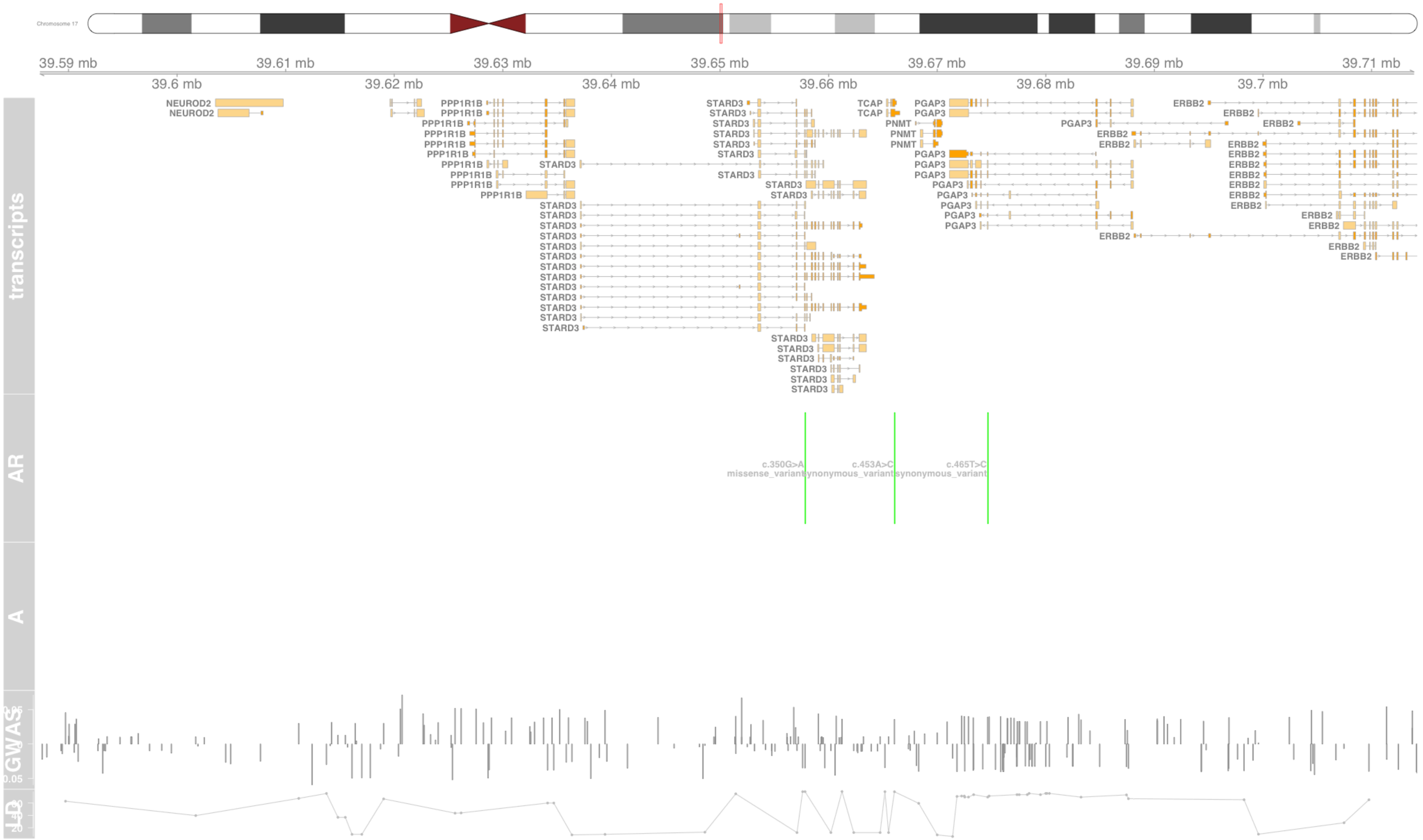

#### Fig S108 STAT6

signal transducer and activator of transcription 6 [Source:HGNC Symbol;Acc:HGNC:11368]

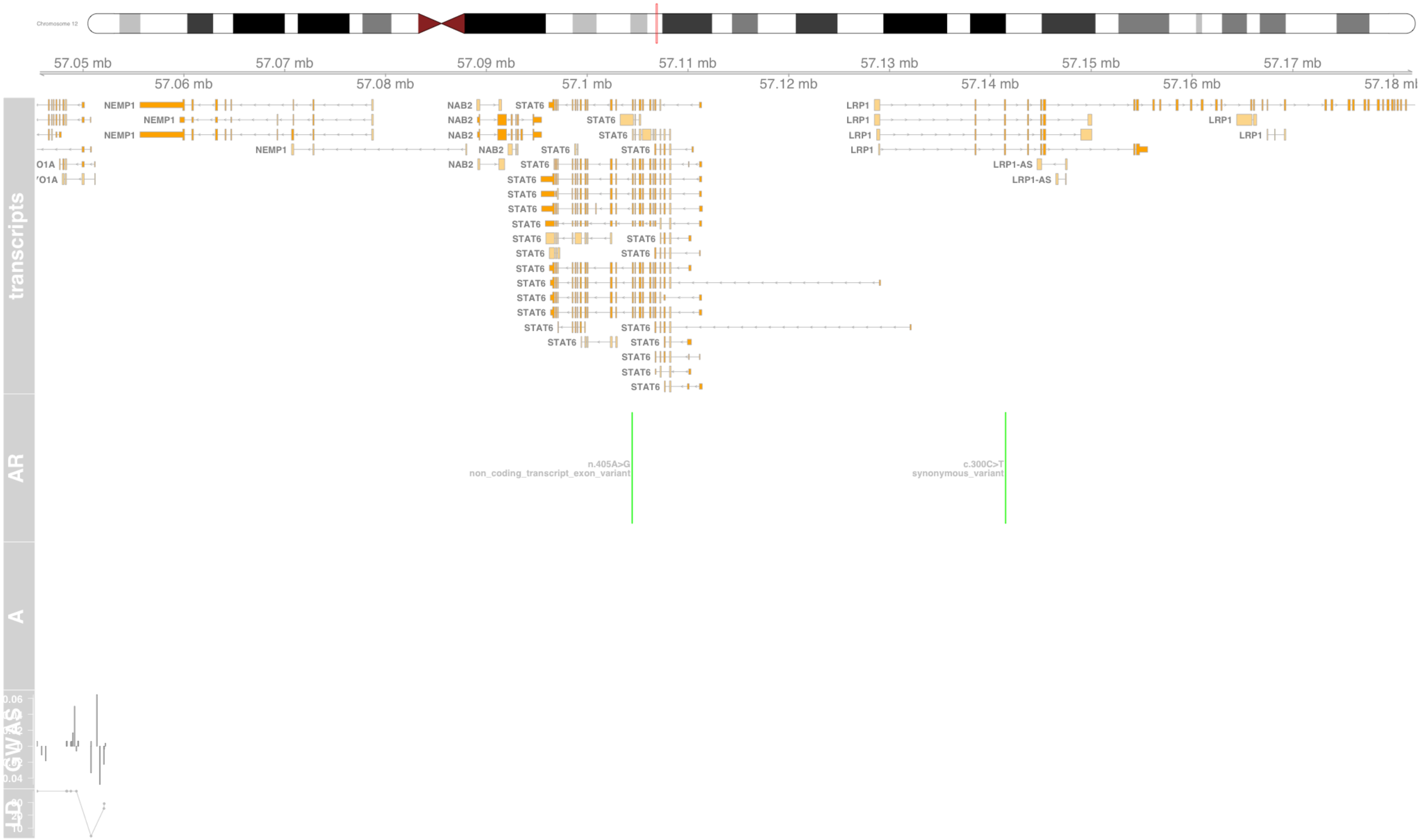

### Fig S109 STK19

serine/threonine kinase 19 [Source:HGNC Symbol;Acc:HGNC:11398]

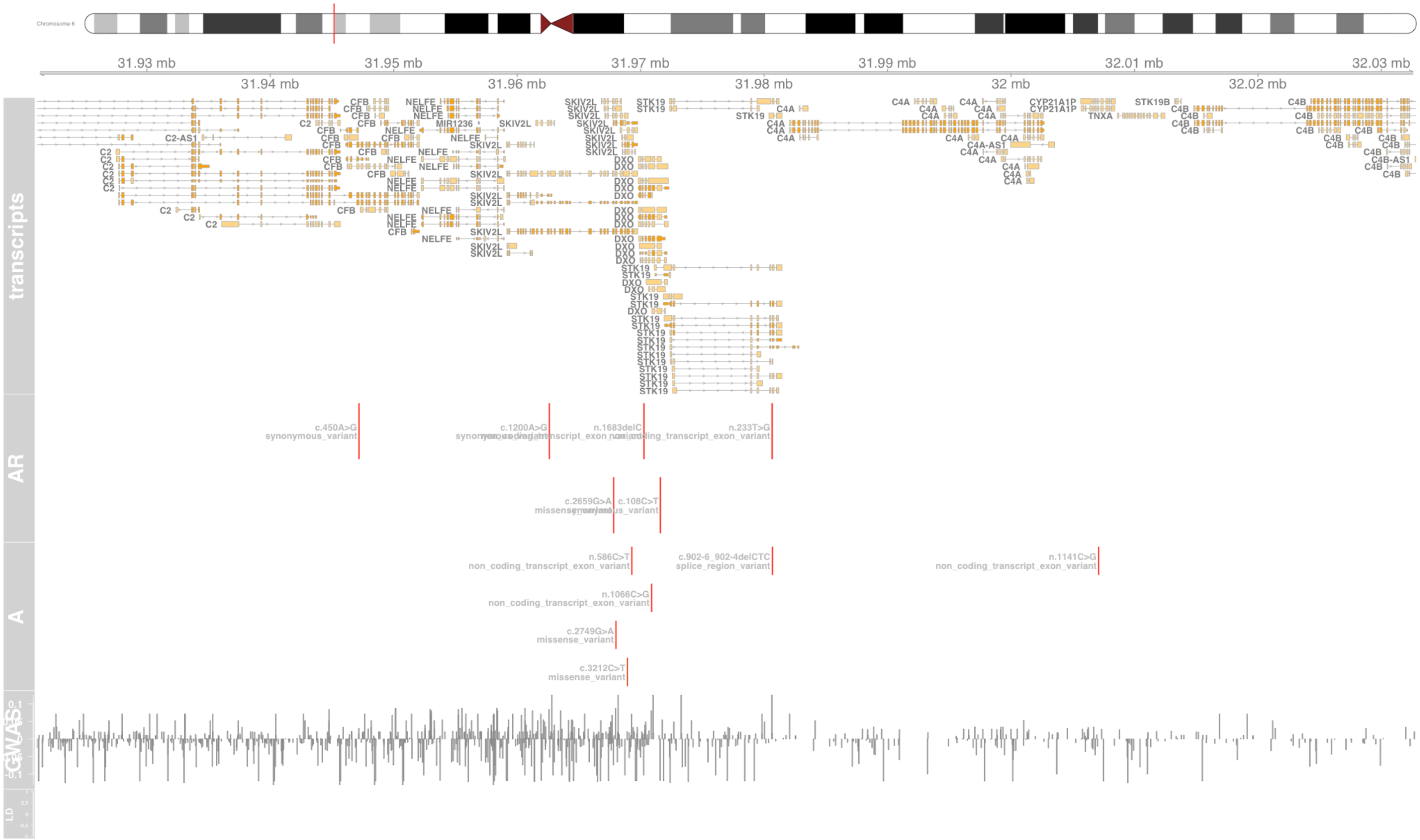

#### Fig S110 TAP2

transporter 2, ATP binding cassette subfamily B member [Source:HGNC Symbol;Acc:HGNC:44]

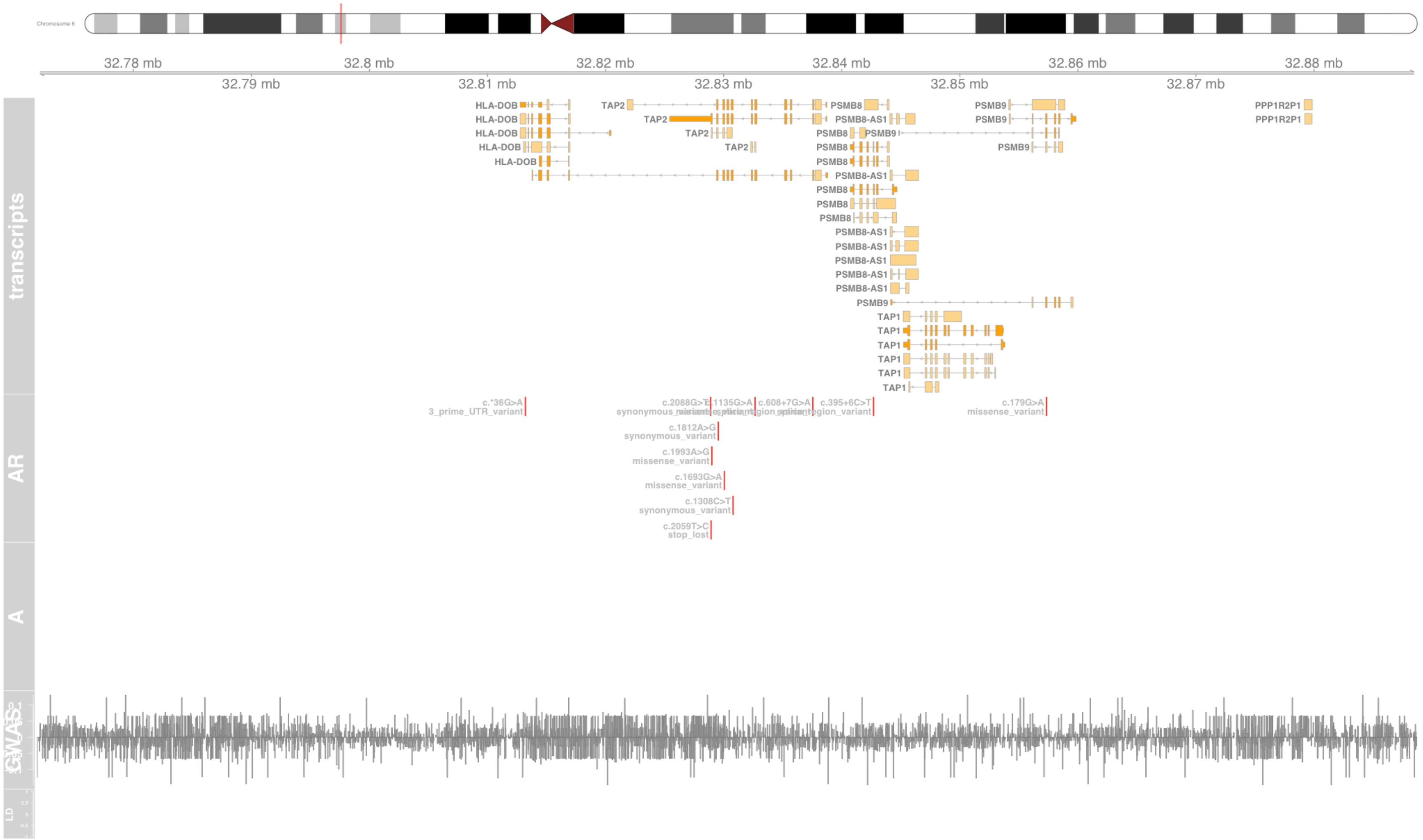

#### Fig S111 TCAP

titin-cap [Source:HGNC Symbol;Acc:HGNC:11610]

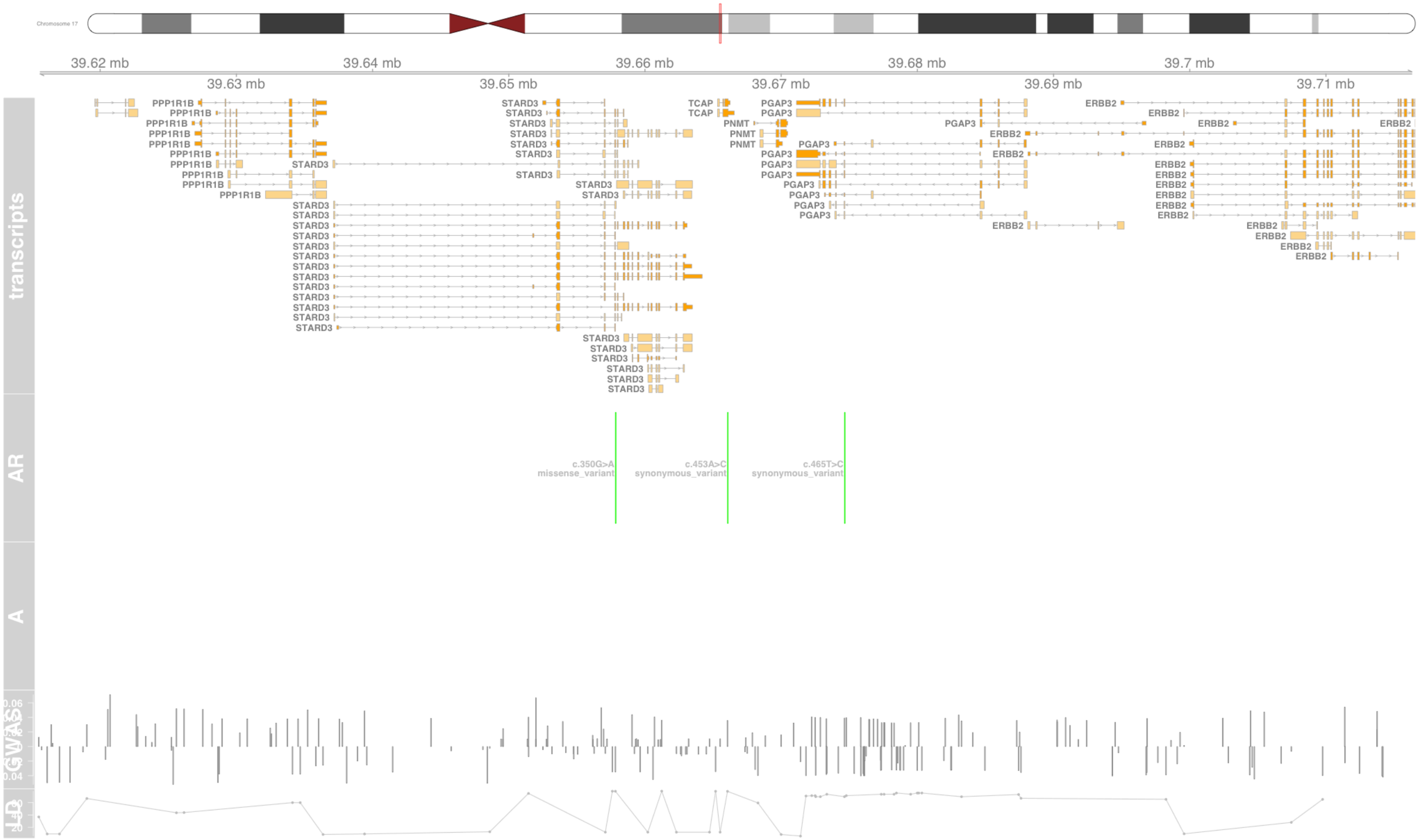

### Fig S112 TCF19

transcription factor 19 [Source:HGNC Symbol;Acc:HGNC:11629]

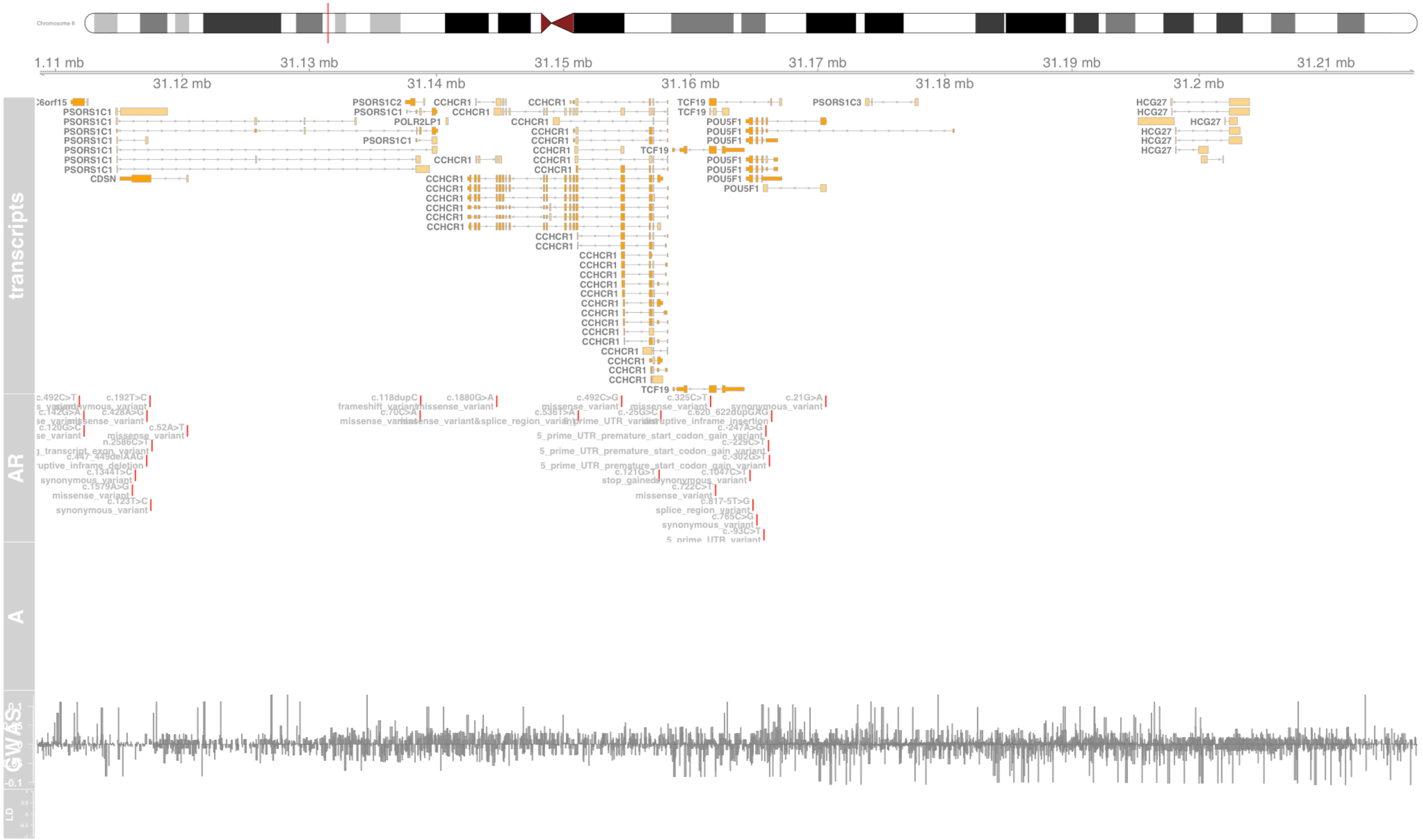

### Fig S113 TLR1

toll like receptor 1 [Source:HGNC Symbol;Acc:HGNC:11847]

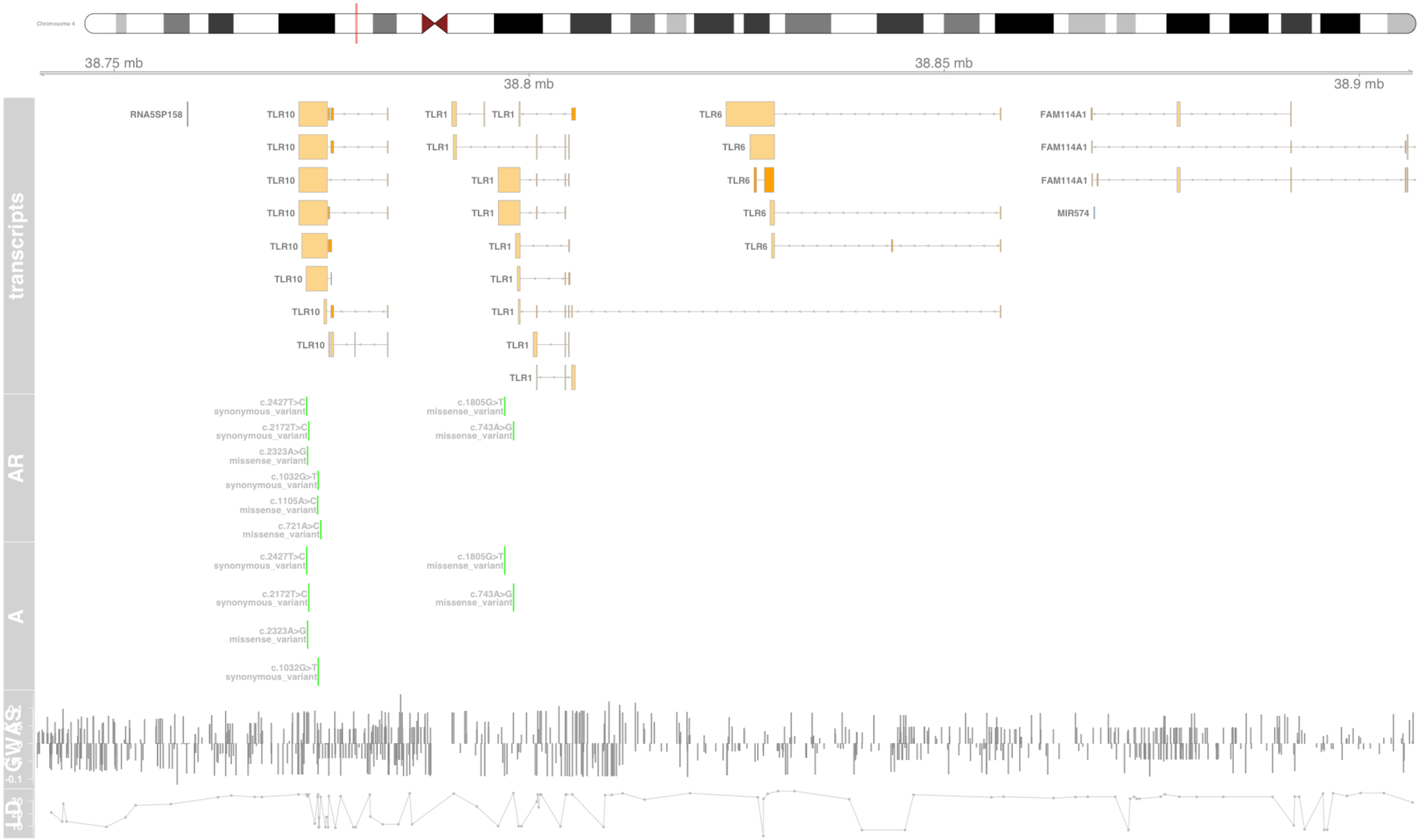

#### Fig S114 TLR10

toll like receptor 10 [Source:HGNC Symbol;Acc:HGNC:15634]

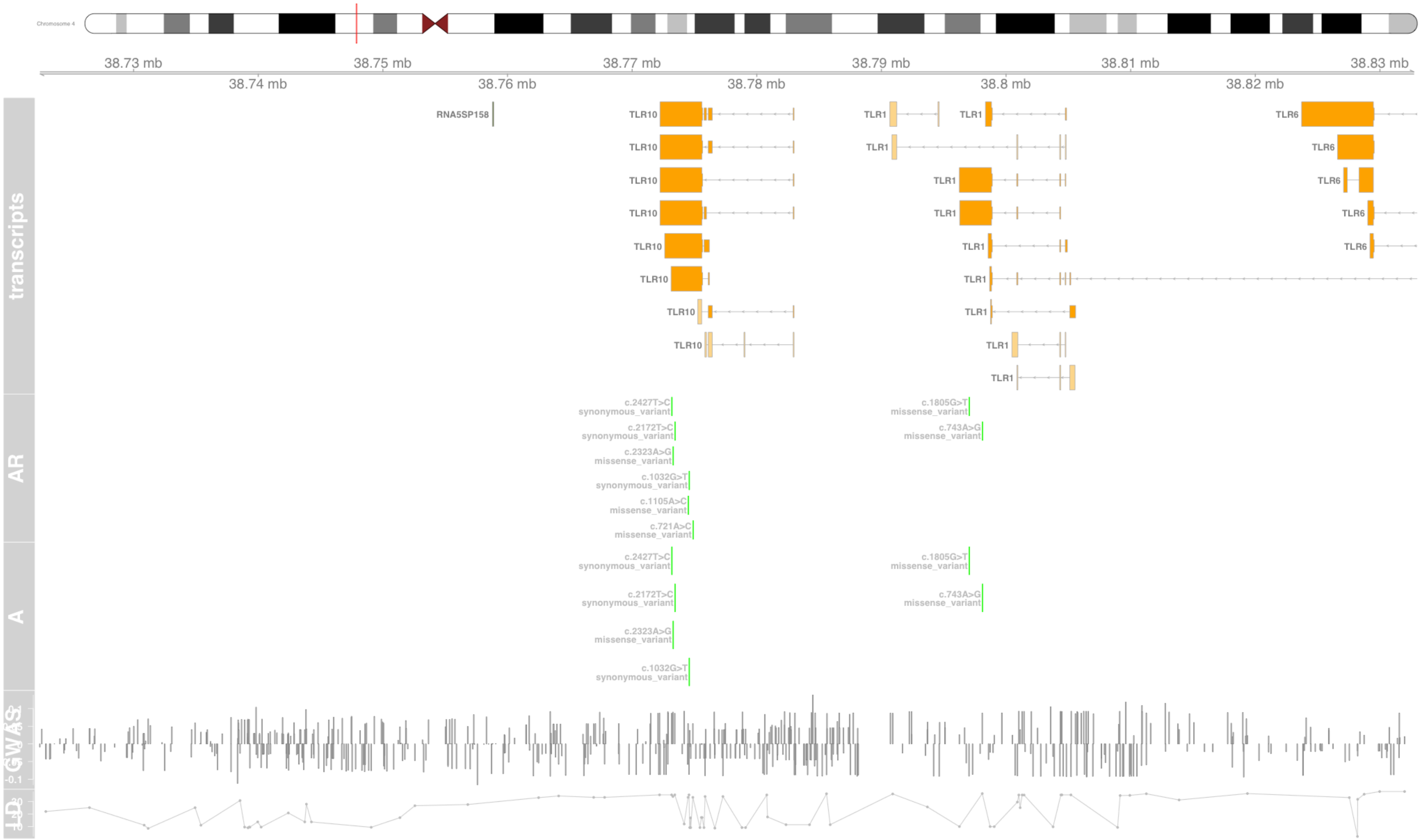

#### Fig S115 TNXB

tenascin XB [Source:HGNC Symbol;Acc:HGNC:11976]

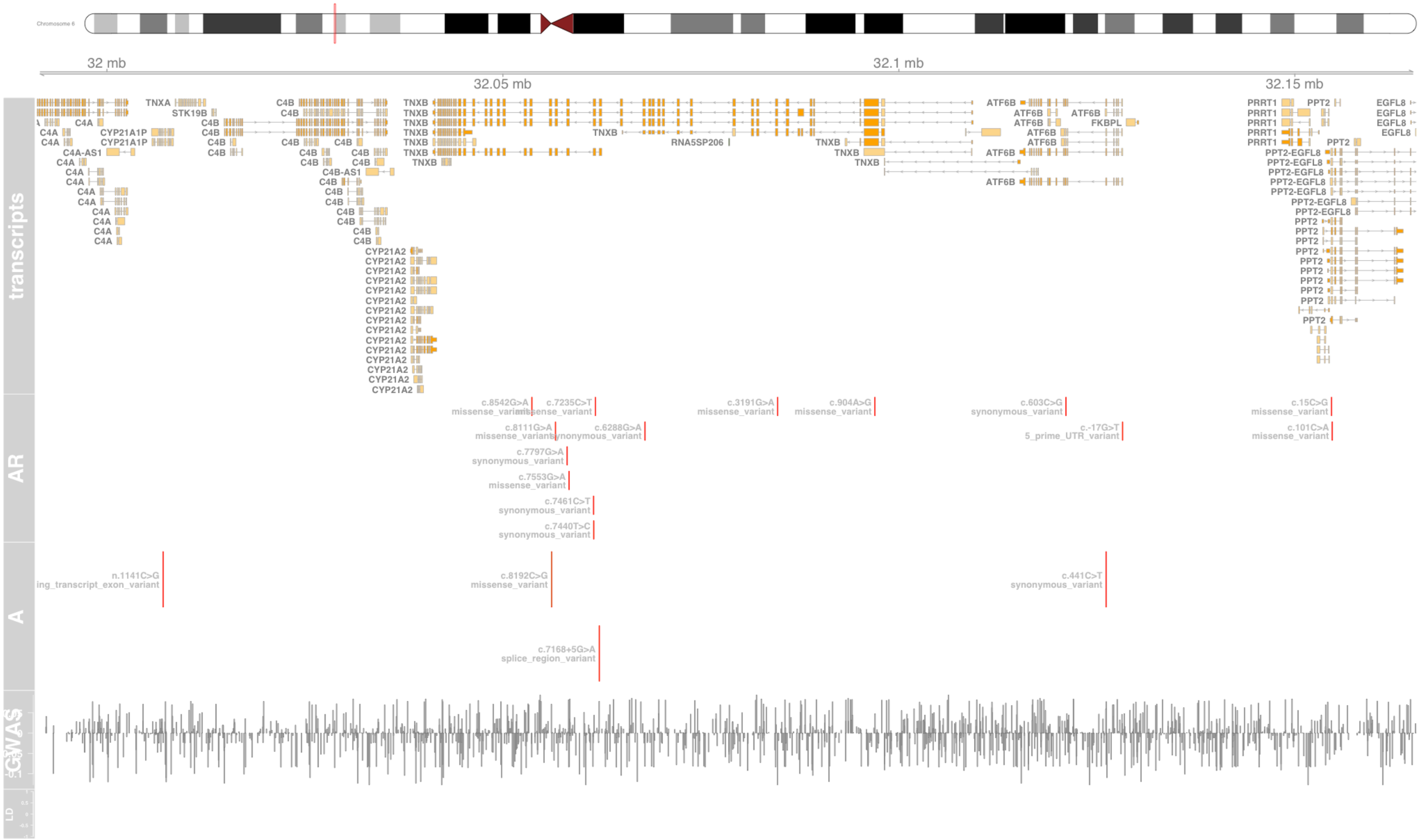

#### Fig S116 TOB2

transducer of ERBB2, 2 [Source:HGNC Symbol;Acc:HGNC:11980]

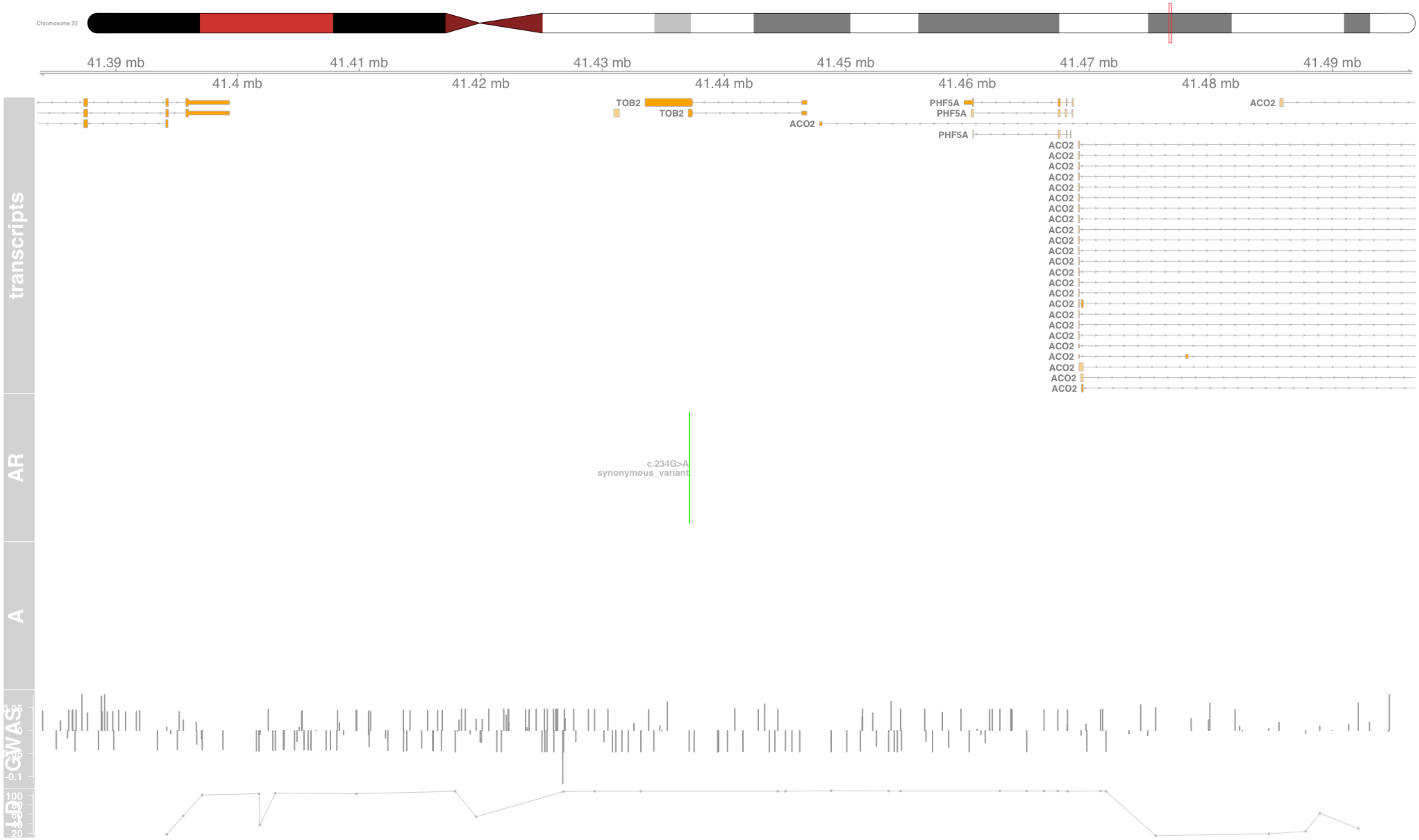

### Fig S117 TRIM26

tripartite motif containing 26 [Source:HGNC Symbol;Acc:HGNC:12962]

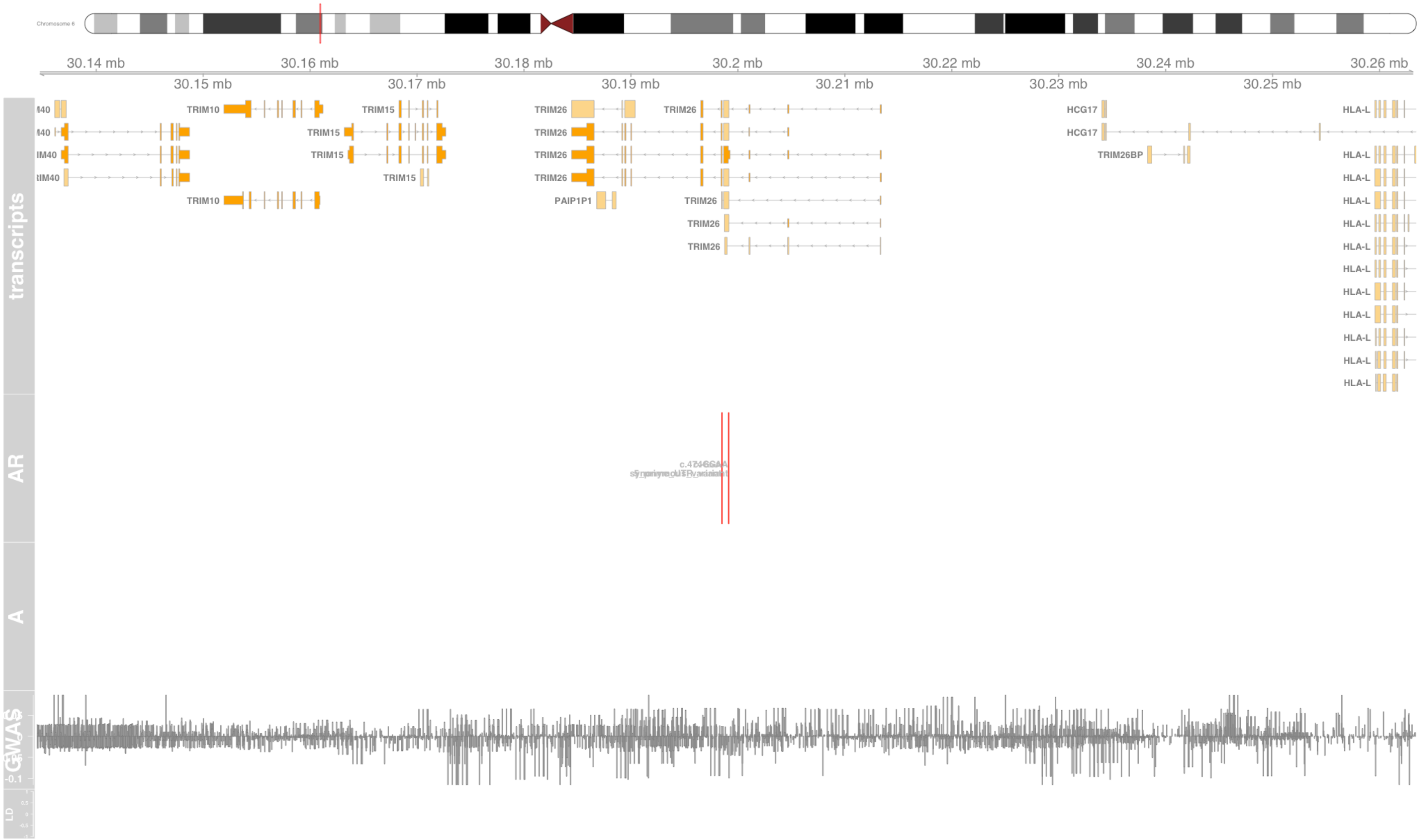

#### Fig S118 TRIM27

tripartite motif containing 27 [Source:HGNC Symbol;Acc:HGNC:9975]

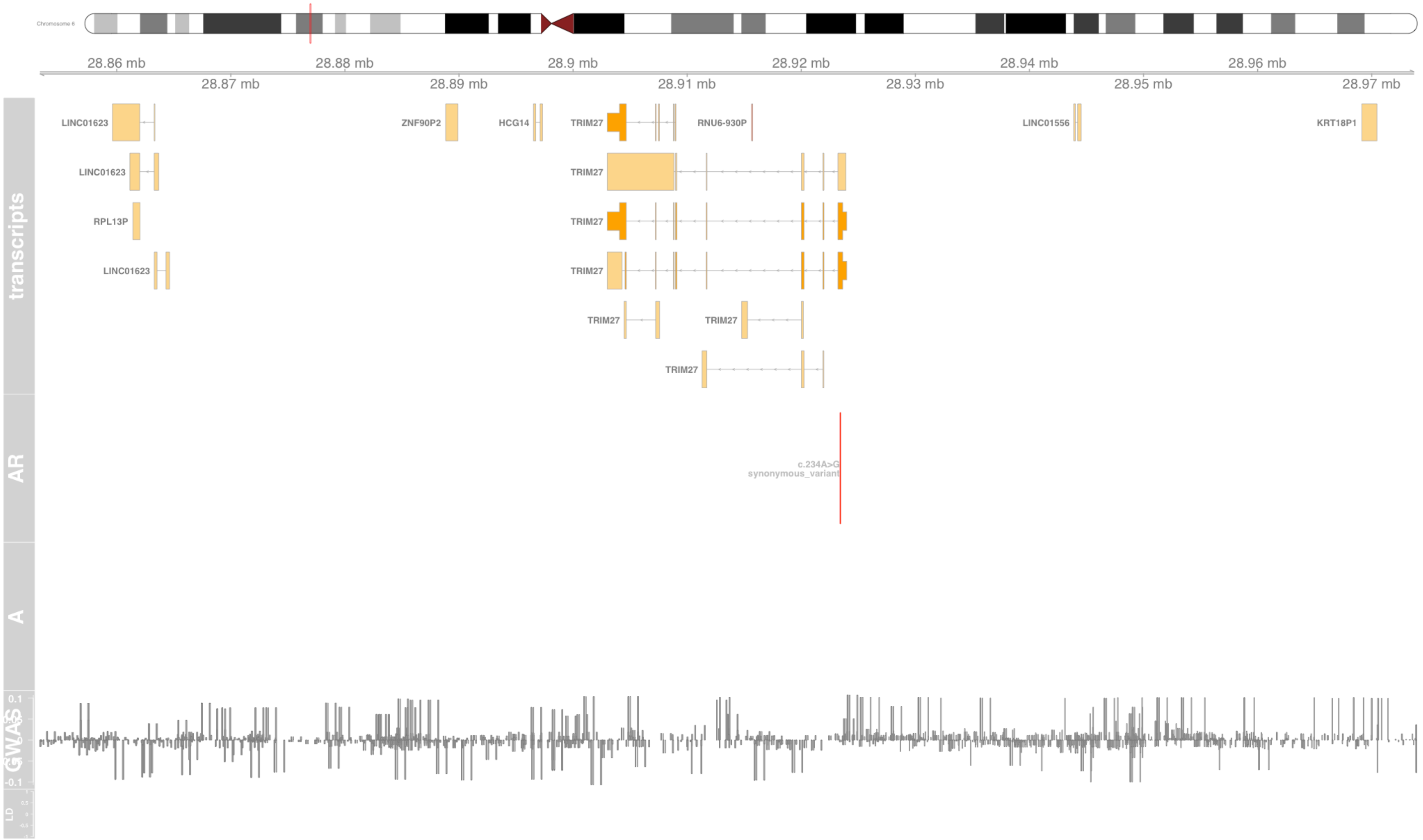

#### Fig S119 TRIM31

tripartite motif containing 31 [Source:HGNC Symbol;Acc:HGNC:16289]

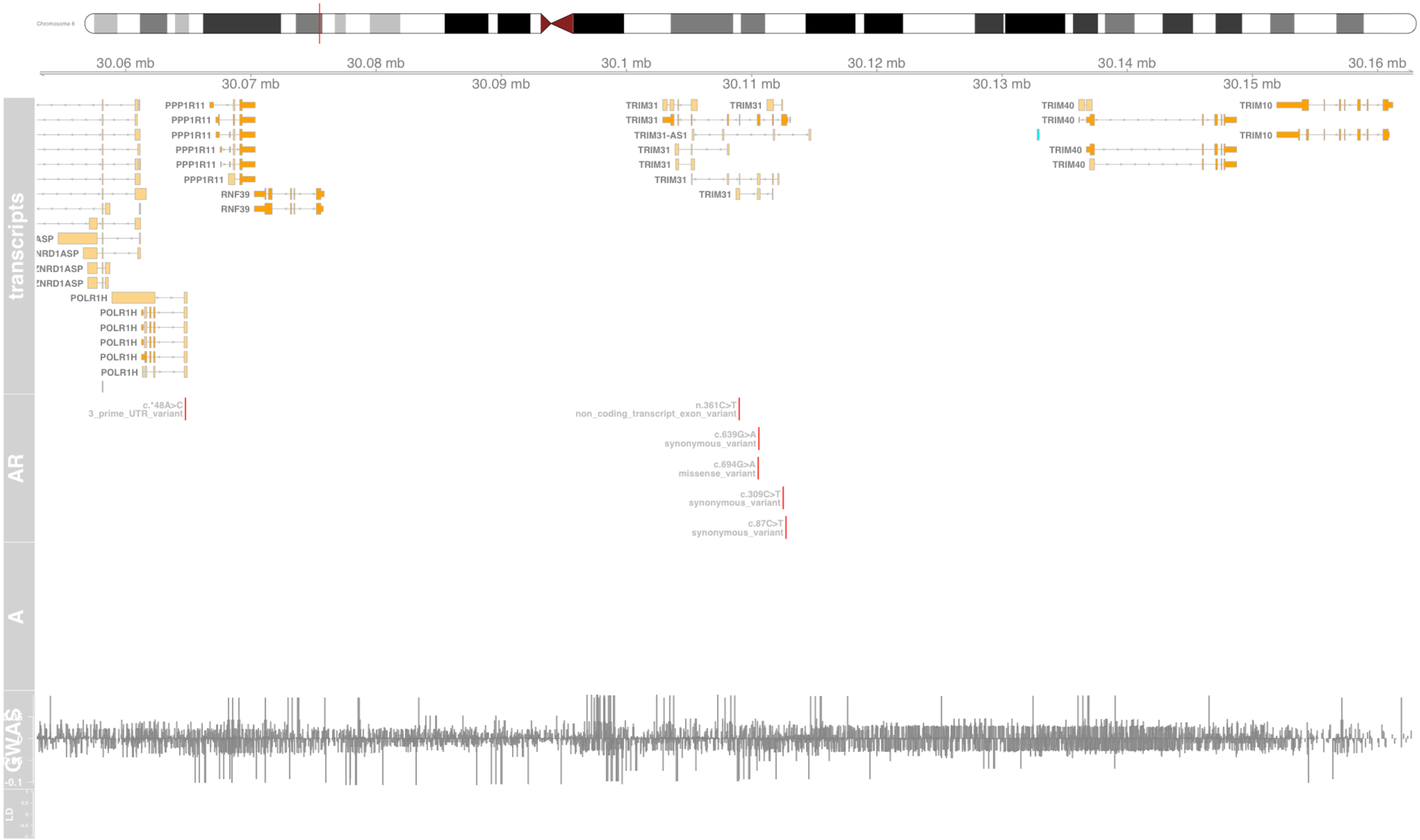

#### Fig S120 TSPAN8

tetraspanin 8 [Source:HGNC Symbol;Acc:HGNC:11855]

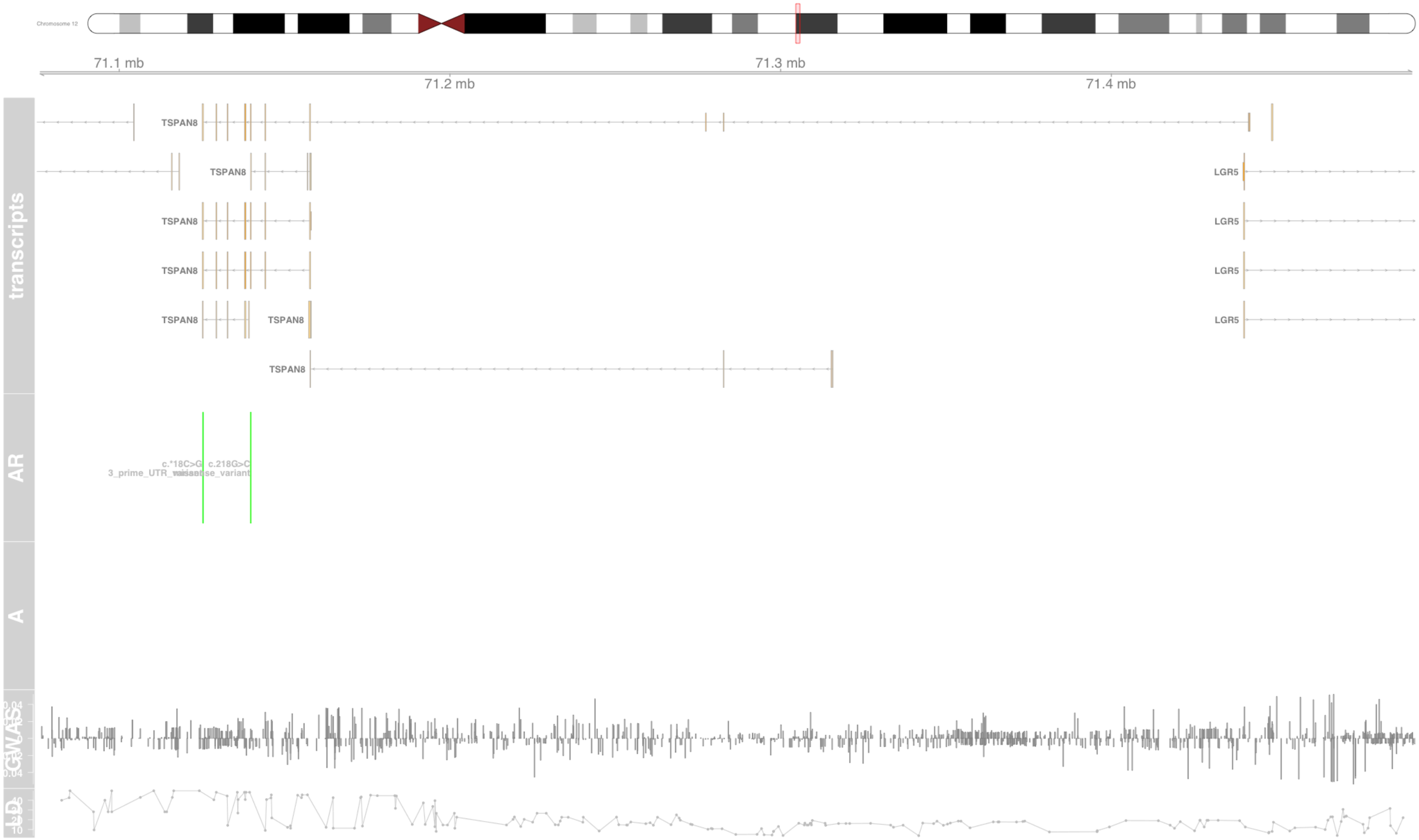

#### Fig S121 UBD

ubiquitin D [Source:HGNC Symbol;Acc:HGNC:18795]

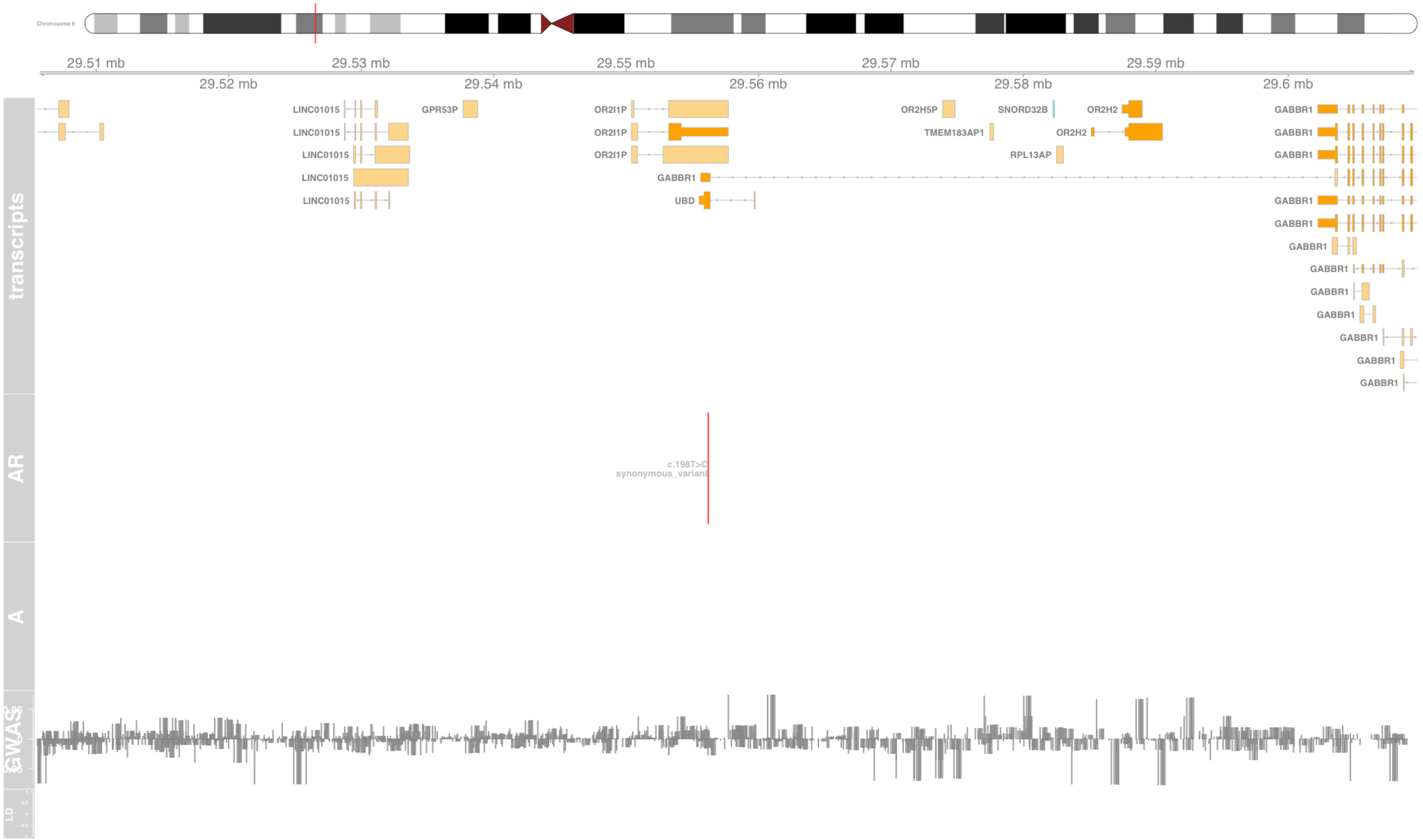

#### Fig S122 UGT3A1

UDP glycosyltransferase family 3 member A1 [Source:HGNC Symbol;Acc:HGNC:26625]

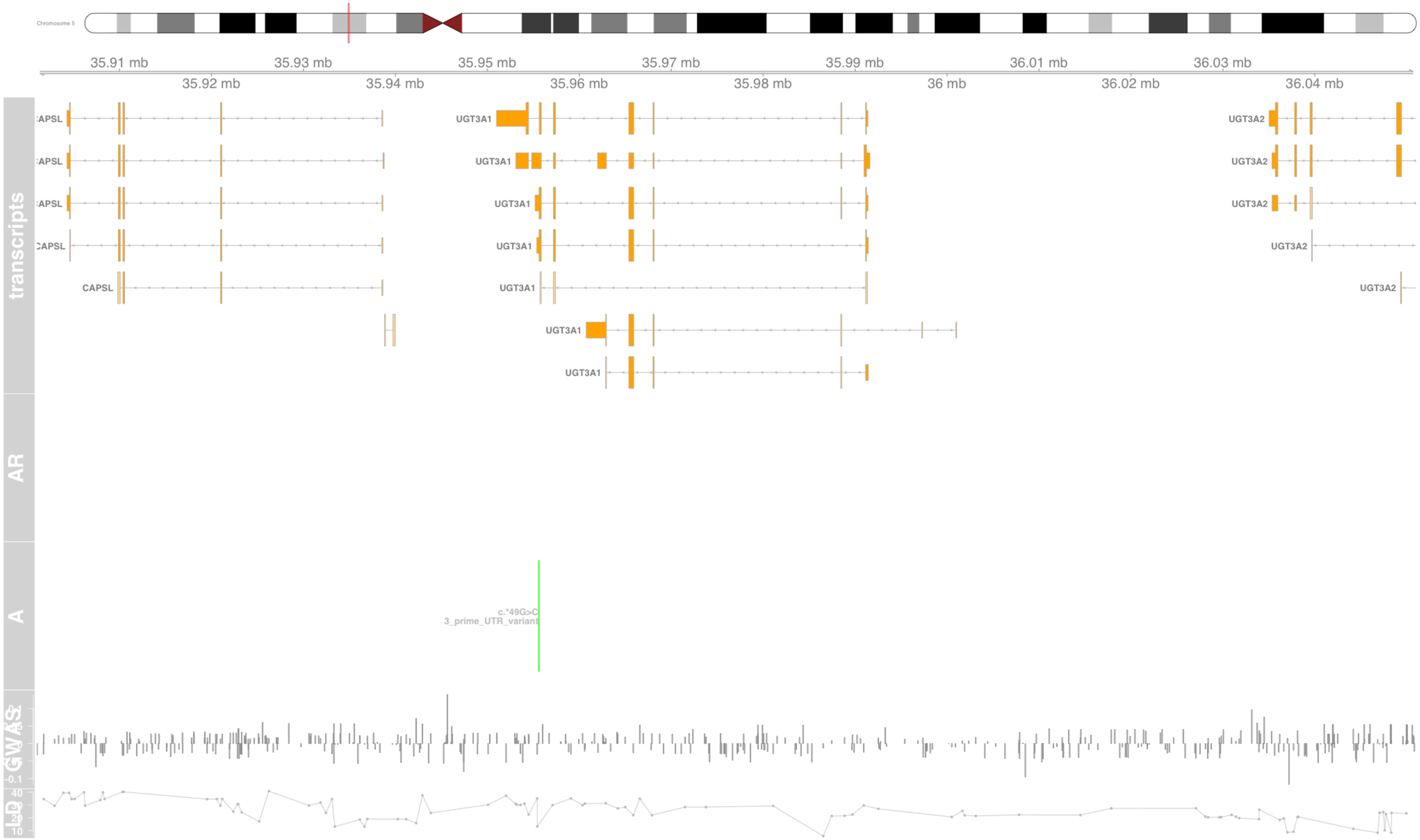

#### Fig S123 VWA7

von Willebrand factor A domain containing 7 [Source:HGNC Symbol;Acc:HGNC:13939]

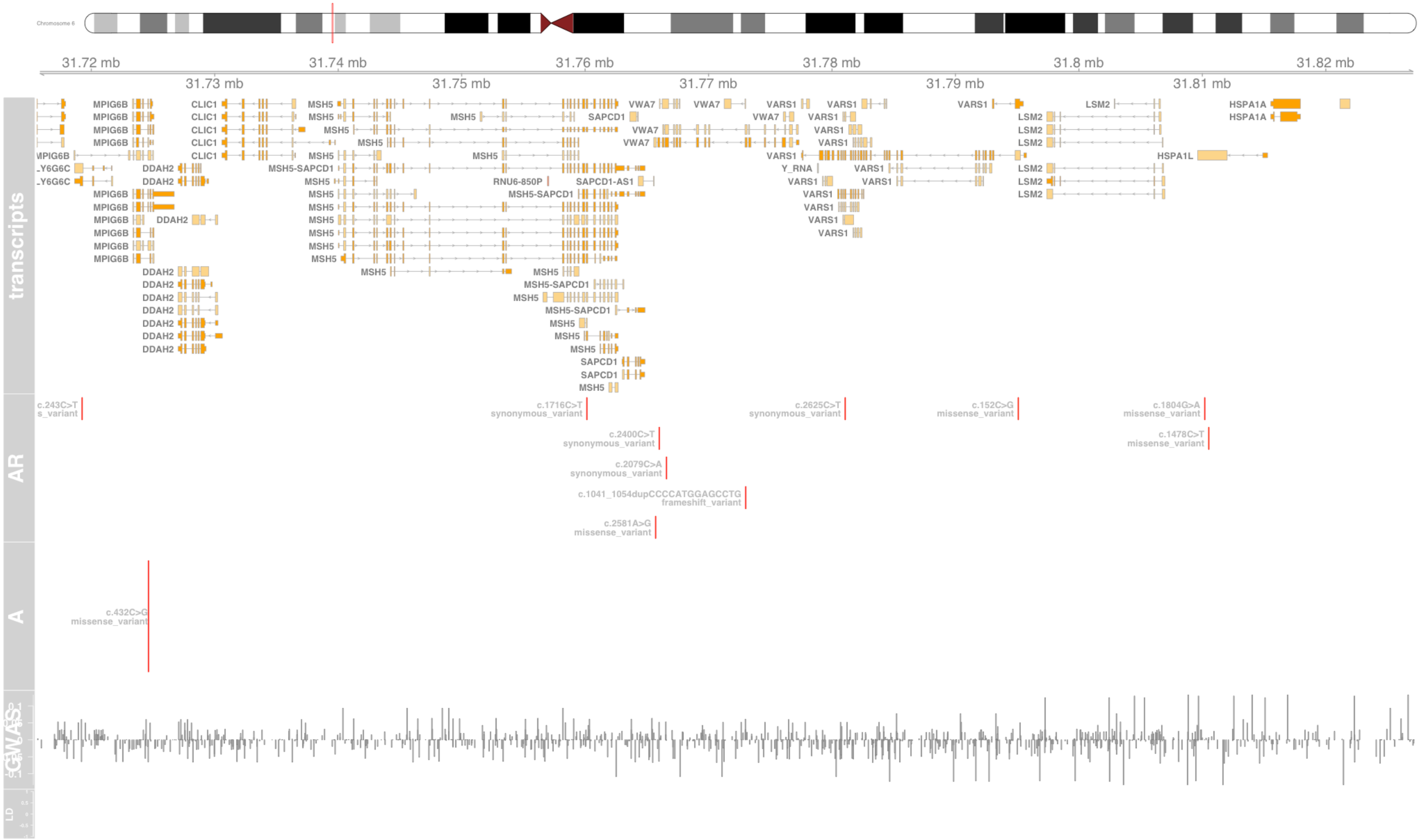

#### Fig S124 WDR36

WD repeat domain 36 [Source:HGNC Symbol;Acc:HGNC:30696]

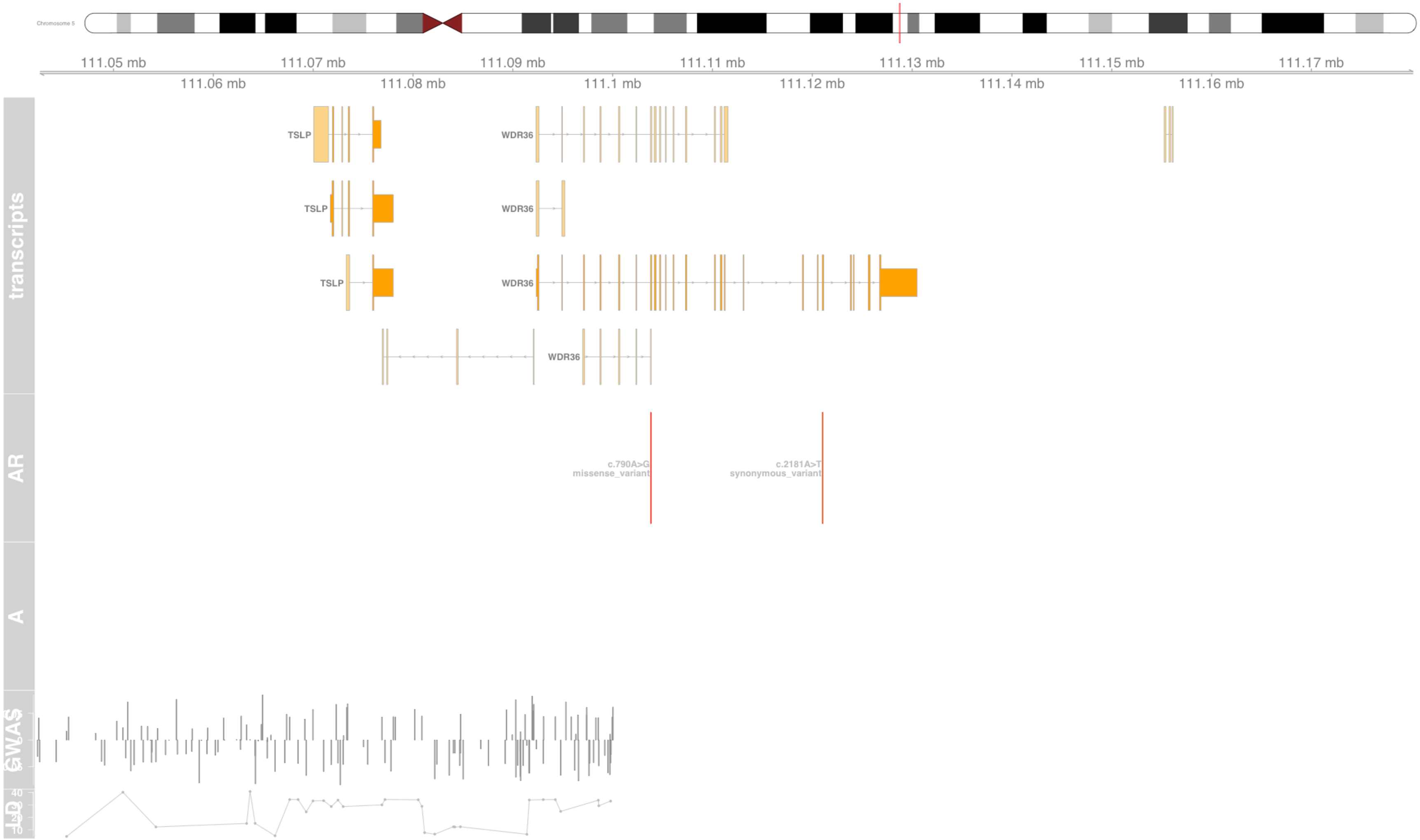

#### Fig S125 ZBED9

zinc finger BED-type containing 9 [Source:HGNC Symbol;Acc:HGNC:13851]

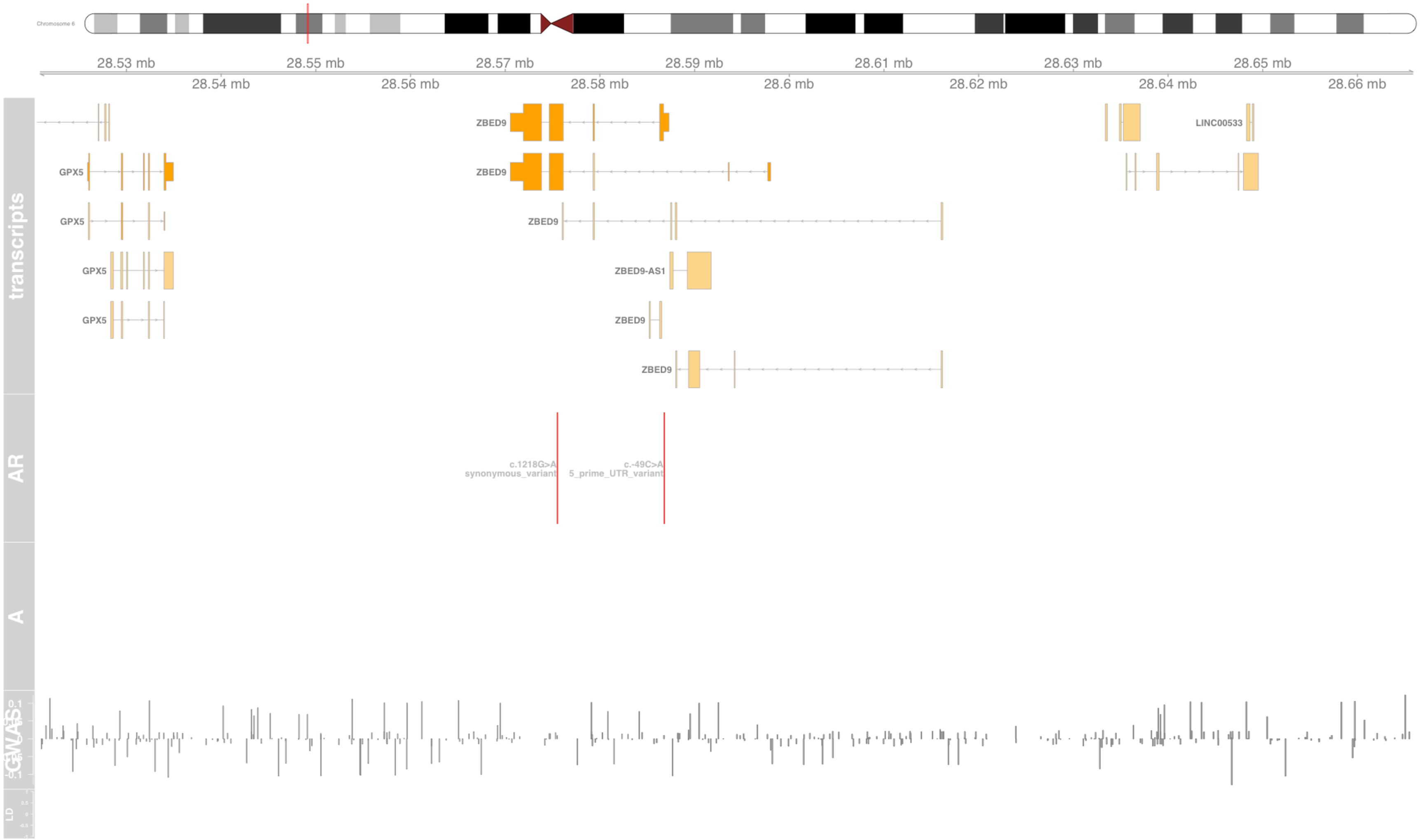

#### Fig S126 ZPBP2

zona pellucida binding protein 2 [Source:HGNC Symbol;Acc:HGNC:20678]

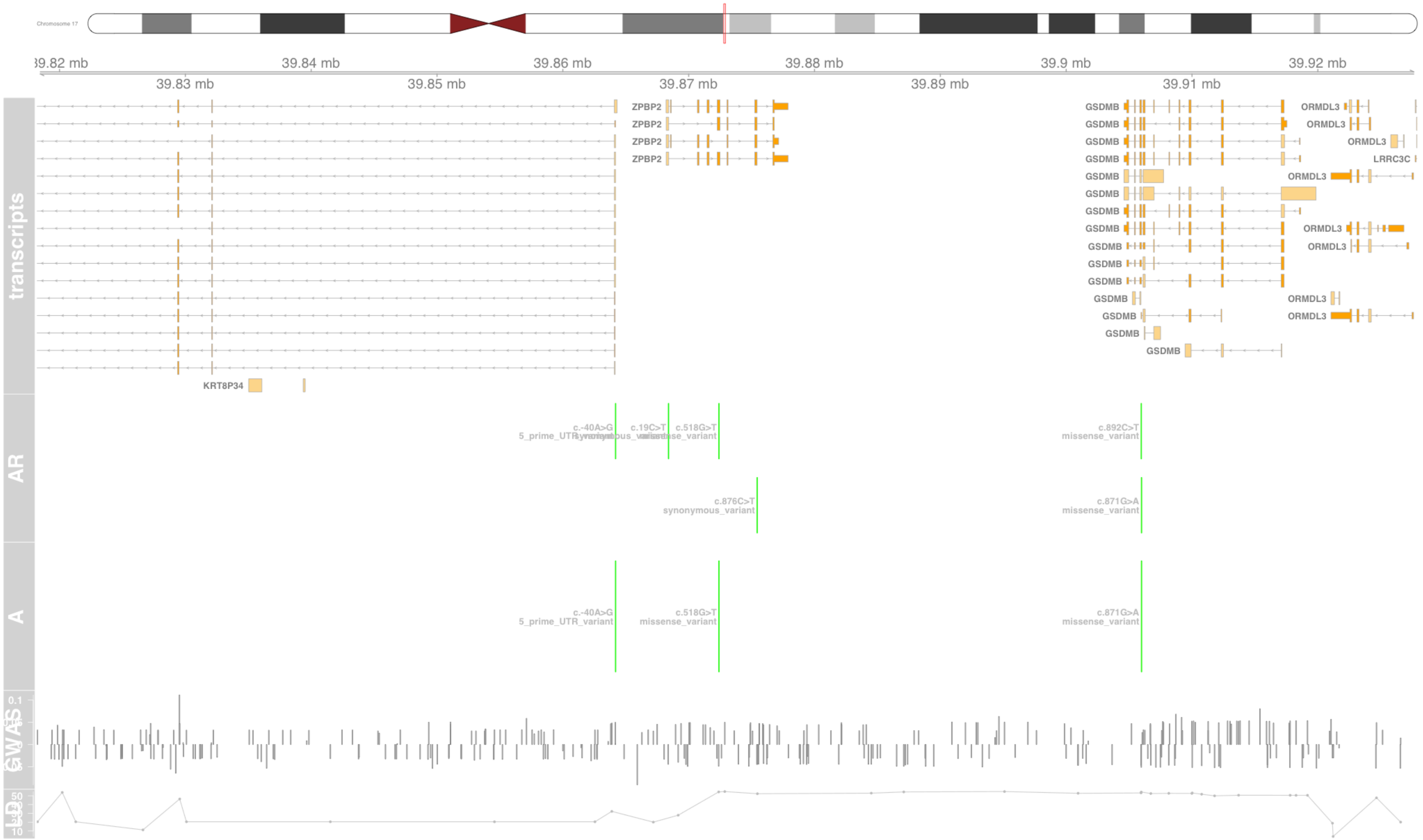

#### Fig S127 ZSCAN12

zinc finger and SCAN domain containing 12 [Source:HGNC Symbol;Acc:HGNC:13172]

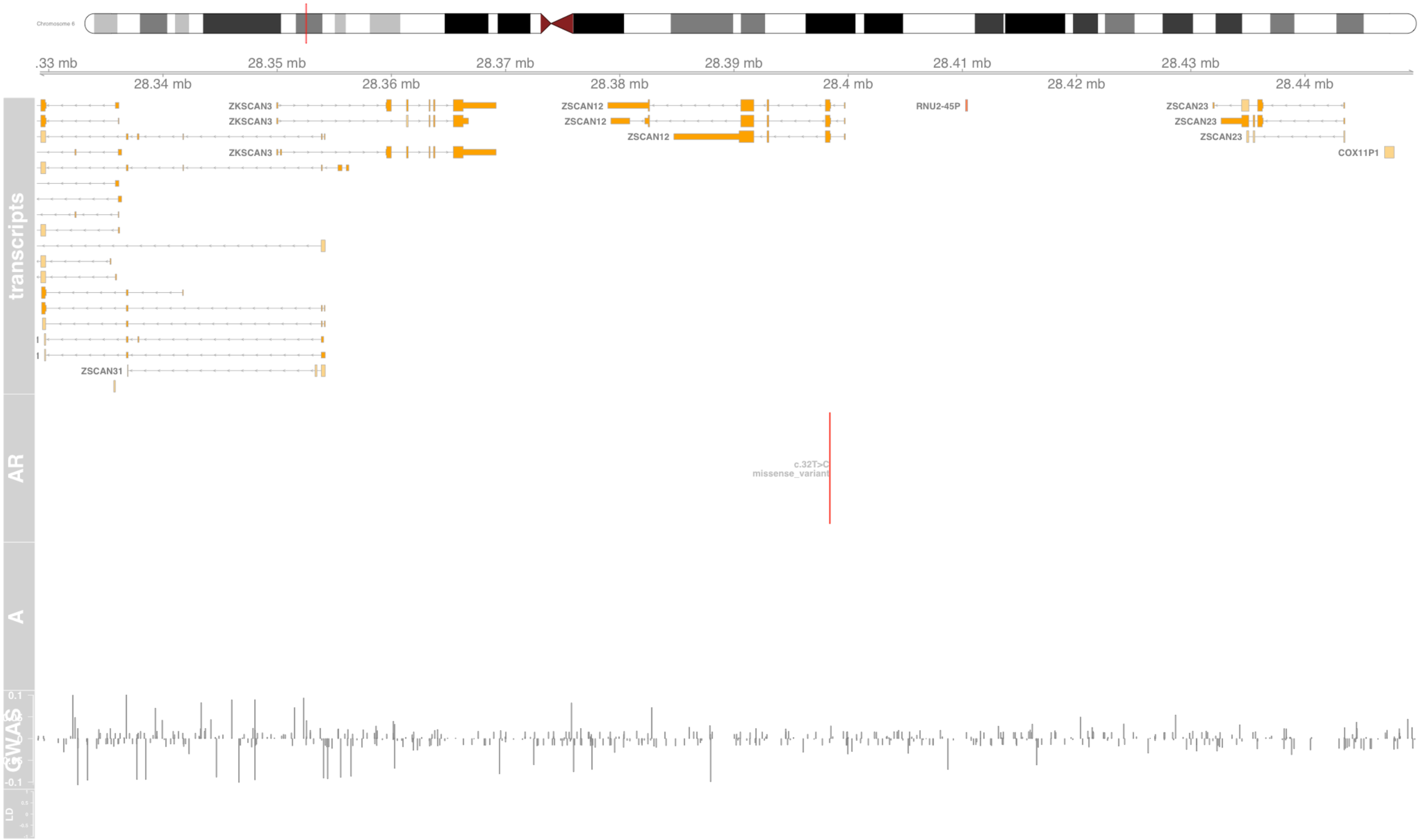
